# Insights into the limited global spread of the immune evasive SARS-CoV-2 variant Mu

**DOI:** 10.1101/2022.03.28.22273077

**Authors:** Mary E. Petrone, Carolina Lucas, Bridget Menasche, Mallery I. Breban, Inci Yildirim, Melissa Campbell, Saad B. Omer, Albert I. Ko, Nathan D. Grubaugh, Akiko Iwasak, Craig B. Wilen, Chantal B.F. Vogels, Joseph R. Fauver

**Affiliations:** Department of Epidemiology of Microbial Diseases, Yale School of Public Health, New Haven, CT, USA; School of Life and Environmental Sciences, University of Sydney, Sydney, NSW, Australia; Department of Immunobiology, Yale University School of Medicine, New Haven, CT, USA; Department of Laboratory Medicine, Yale University School of Medicine, New Haven, CT, USA; Department of Pediatric, Section of Infectious Diseases and Global Health, Yale University School of Medicine, New Haven, CT, USA; Yale Institute for Global Health, Yale University, New Haven, CT, USA; Department of Medicine, Section of Infectious Diseases, Yale University School of Medicine, New Haven, CT, USA; Department of Ecology and Evolutionary Biology, Yale University, New Haven, CT, USA; Howard Hughes Medical Institute, Chevy Chase, MD, USA; College of Public Health, University of Nebraska Medical Center, Omaha, NE, USA

**Author notes:** These authors contributed equally (Mary E. Petrone, Carolina Lucas, Bridget Menasche). These authors contributed equally (Chantal B.F. Vogels, Joseph R. Fauver).

## Abstract

SARS-CoV-2 ‘Variants of Concern’ (VOCs) continue to reshape the trajectory of the COVID-19 pandemic. However, why some VOCs, like Omicron, become globally dominant while the spread of others is limited is not fully understood. To address this question, we investigated the VOC Mu, which was first identified in Colombia in late 2020. Our study demonstrates that, although Mu is less sensitive to neutralization compared to variants that preceded it, it did not spread significantly outside of South and Central America. Additionally, we find evidence that the response to Mu was impeded by reporting delays and gaps in the global genomic surveillance system. Our findings suggest that immune evasion alone was not sufficient to outcompete highly transmissible variants that were circulating concurrently with Mu. Insights into the complex relationship between genomic and epidemiological characteristics of previous variants should inform our response to variants that are likely to emerge in the future.

## INTRODUCTION

The emergence of new SARS-CoV-2 variants sheds light on the complex interactions of virus genetic mutations, immune evasion, and transmissibility. By the end of 2021, 13 variants of public health significance were identified by the World Health Organization (WHO)^1^. Of these, the highly transmissible Alpha and Delta variants sequentially became dominant in many parts of the world^2–9^, while variants characterized by immune evasive mutations resulting in amino acid substitutions in the spike protein gene were arguably less impactful^10^. The emergence of Omicron, which is both highly transmissible and capable of evading vaccine-induced immunity^11–14^, and the exponential rise in Omicron infections that ensued, suggested that the combination of these factors conferred a higher level of viral fitness than was previously observed in the COVID-19 pandemic. Because new variants will inevitably emerge, there is an urgent need to better understand why certain constellations of concerning mutations result in international spread while others do not.

The variant Mu (pango lineage B.1.621) presents an apt case study for addressing this question. The lineage B.1.621 was first detected in Colombia in late 2020, and its emergence coincided with a rise in reported COVID-19 cases^15^. B.1.621 was designated as a Variant of Concern (VOC, Mu) by the WHO and the United States Centers for Disease Control and Prevention (CDC) in 2021^16,17^. Three sublineages of B.1.621 have emerged to comprise the Mu clade: B.1.621.1, B.1.621.2, and BB.2 (cov-lineages.org). As of January 1, 2022, Mu clade genome sequences had been identified through sequencing in 52 countries (gisaid.org). Mu initially elicited concern due to a number of mutations associated with immune evasion. Specifically, substitutions in the spike gene receptor binding domain (RBD) (E484K, N501Y, and R346K) are found in other VOCs including Beta, Gamma, and Omicron that have been shown to be less sensitive to neutralization by vaccine-induced antibodies^18^. Despite these characteristics, Mu did not become dominant in countries outside of South and Central America, and minimal local transmission has been reported in other regions^19,20^.

In this study, we adopt a multidisciplinary approach to investigate the emergence and limited spread of Mu. Previous studies have separately documented the reduced sensitivity of Mu to neutralization^18,21,22^ and characterized local outbreaks^15,19,23^, but none have comprehensively analyzed these correlates of variant fitness in conjunction. We begin by evaluating the biological characteristics of Mu using neutralization and replication assays and find that Mu is more resistant to neutralization compared to the VOCs that preceded it. In contrast, we do not observe elevated replication rates among any of the variants tested. We subsequently perform a phylogenetic analysis and show that there was a substantial lag between when Mu likely emerged and when it was first detected in South America. Subsequent investigation of the Mu genome reveals that a frameshift mutation causing a premature stop codon in ORF3a may have delayed the publication of Mu clade genome sequences on the public repository GISAID (gisaid.org). Finally, we conduct discrete phylogeographic inference, which reveals that the Mu clade did not circulate widely outside of South America, contrary to our expectations given its immune evasive properties and long period of undetected transmission. The multidisciplinary nature of this study, in which we use known circulating SARS-CoV-2 genomes to generate hypotheses for biomedical and epidemiological assessment, provides valuable insight into the genomic correlates of variant fitness.

## RESULTS

### The SARS-CoV-2 lineage B.1.621 (Mu) is less susceptible to neutralization compared to previous lineages

The SARS-CoV-2 lineage B.1.621 (Mu) was designated as a Variant of Concern (VOC) in August 2021 by the World Health Organization (WHO) due to its clade-defining mutations known to impact immune evasion^16^. Mu, which was first detected in Colombia at the end of 2020, is defined by 23 non-synonymous mutations, the plurality of which (n = 9) encoded amino acids in the spike protein (**Fig. 1a**). A number of these mutations are shared by other VOCs and are associated with immune evasion, including R346K24, E484K6,25–28, N501Y6,28, and P681H6,29–31. Notably, Mu shares the spike substitutions T95I, N501Y, P681H, and R346K with Omicron lineages (CoVariants.org). Mu also contains two notable insertions/deletions (indels). First, a 3 nucleotide insertion in the N-terminal domain results in amino acid changes Y144S and Y145N. Second, a 4 nucleotide deletion causes a frameshift mutation in ORF 3a (Δ256/257) that truncates the reading frame by encoding a premature stop codon (P258*).

**Figure 1:**
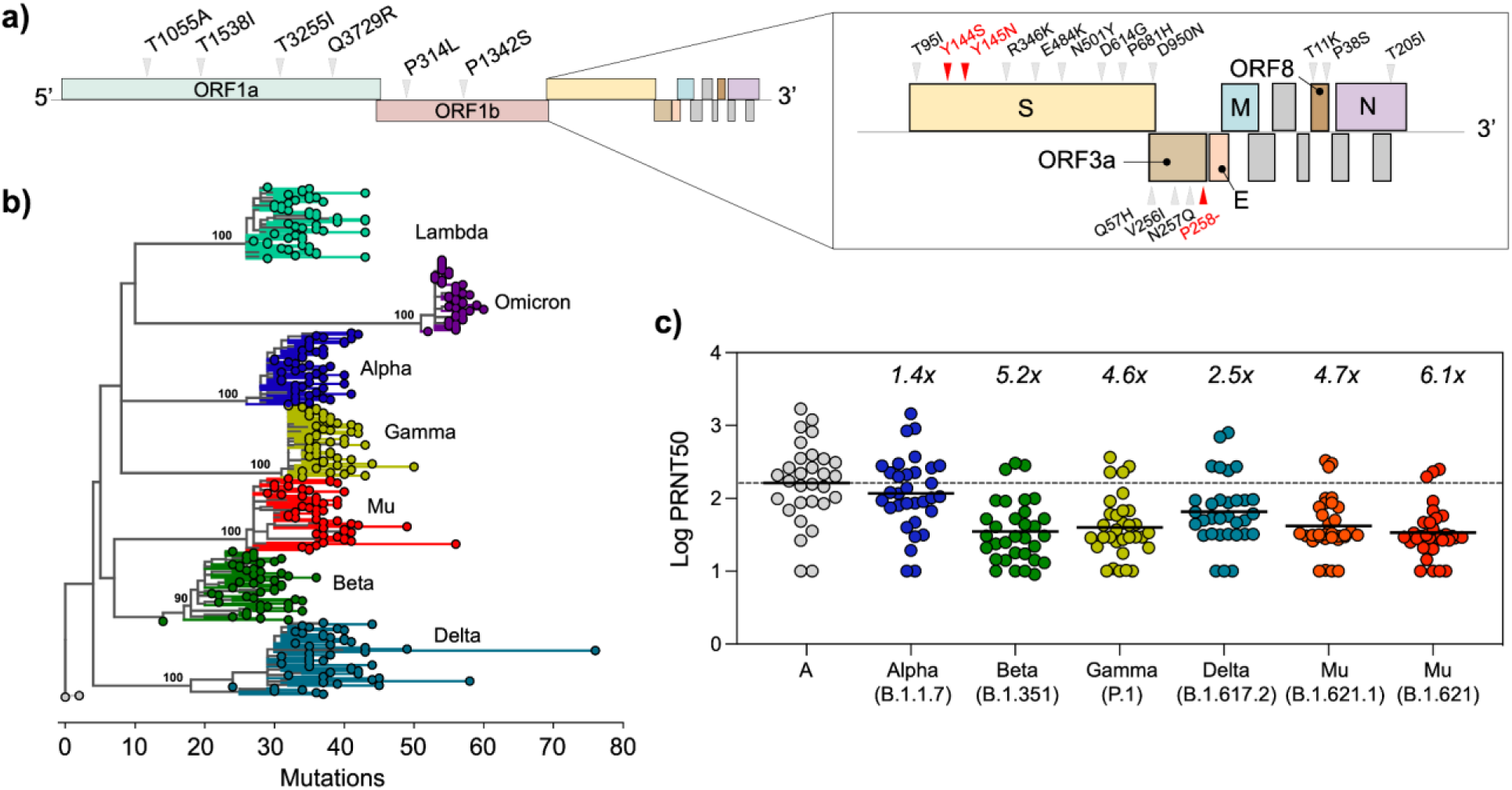
Mu is characterized by mutations that enhance immune evasion. **(a)** Schematic of SARS-CoV-2 genome with Mu clade defining mutations marked. The substitutions Y114S and Y145N (red) are the result of a 3bp insertion in the N-terminal domain. P258*, the premature stop codon resulting from a frameshift in ORF3a, is also shown in red. **(b)** Maximum likelihood phylogenetic tree of WHO Variants of Concern (n = 50 genome sequences per variant). We constructed this tree in IQTree using a GTR+G substitution model and 1000 bootstraps. Bootstrap values are indicated for internal nodes. **(c)** Plasma neutralization titers of key SARS-CoV-2 lineages 28 days post second vaccination dose.

Three sublineages of B.1.621 have been identified. The emergence of the sublineage B.1.621.1 followed shortly after that of Mu (**Fig. 1b**). B.1.621.1, which also contains the substitutions E484K, N501Y, and R346K, was designated as a ‘Variant Being Monitored’ by the CDC in conjunction with B.1.621 in September 202117. This lineage was not given a designation by the WHO at that time. Two additional sublineages of Mu, B.1.621.2 and BB.2, were later characterized (cov-lineages.org). Of note, the former contains two additional amino acid changes in the spike protein RBD, Y449N and E583D.

The utility of genomic surveillance based on mutation profiles as described above is well established and has been further reinforced during this pandemic. It is crucial that VOCs are further investigated using experimental systems to measure effects on transmissibility and immune evasion. Previously, we found that the added effects of the spike amino acid changes E484K and N501Y on vaccine-induced neutralization resulted in the greatest reduction in neutralization^6^. Given that Mu has these two amino acid changes in common with Beta and Gamma, we hypothesized that serum neutralization for Mu would be significantly decreased.

We tested this hypothesis by comparing the neutralization capacity of plasma samples from vaccinated individuals against a panel of authentic SARS-CoV-2 isolates representing the ancestral lineage A virus, as well as the B.1.1.7 (Alpha), B.1.351 (Beta), B.1.617.2 (Delta), P.1 (Gamma), B.1.621 (Mu), and B.1.621.1 (Mu) variants. Forty healthcare worker (HCW) volunteers from the Yale-New Haven Hospital (YNHH) received two doses of the mRNA vaccine (Moderna or Pfizer). Plasma samples were collected at baseline, prior to vaccination, and 28 days post second vaccination dose. Supporting our hypothesis, we found the lowest neutralization titers for Mu, representing a 4.7-fold and 6.1-fold decrease relative to lineage A (**Fig. 1c**). Our results were consistent with previous findings^18^. To further investigate the biological impact of Mu’s mutation profile, we measured the intrinsic transmissibility of Mu relative to other VOCs by establishing growth curves of our panel of SARS-CoV-2 isolates in human lung cancer (Calu3) cells. In our *in vitro* system, we found no differences in replication between lineage A and the variants we tested (**Supp. Fig. 1**). This suggests that there were no changes in *in vitro* replication phenotypes within our panel of SARS-CoV-2 isolates when tested on Calu3 cells. Together, our experimental data show that B.1.621 and B.1.621.1 have a decreased mRNA vaccine-induced neutralization, but they do not exhibit changes in intrinsic transmissibility in Calu3 cells.

### The public health response to B.1.621 and its sublineages was delayed by gaps in the global genomic surveillance system

Our phylogenetic analysis revealed that B.1.621 likely emerged in South America in mid-2020, well before it was detected. The first sequenced B.1.621 case was from a sample collected on December 18, 2020, in Bogota, Colombia (**Table 1**), and evidence for international spread was documented on January 21, 2021 with the a B.1.621 genome sequence identified in Venezuela. Interestingly, the collection of the first genome sequence of the sublineage BB.2 predates that of B.1.621 by two months (October 2020, in Antioquia Colombia), and one genome sequence assigned to the lineage B.1.621.1 was collected in December 2020, in Florida, USA. Due to the limited volume of genomic data for these lineages, the reasons for which are described in more detail below, it is not possible to resolve the likely location of emergence beyond South America. Regardless, the timing of collection of B.1.621.1 and BB.2 suggests that B.1.621 and its sublineages were circulating for an extended period prior to detection. To test this hypothesis, we constructed a time-resolved phylogenetic tree using 2478 sequences from B.1.621 and related lineages (**Supp. Table 1**) collected between October 14, 2020 and December 31, 2021 (**Fig. 2a**). We estimated that B.1.621 likely emerged by September 2020, but as early as April of the same year (time to the most recent common ancestor (tMRCA) range = 2020-04-15 to 2020-09-18; **Fig. 2a, bar**). The sublineages B.1.621.1, B.1.621.2, and BB.2 emerged shortly thereafter. These lineages persisted through the end of our study period, with the most recent genome sequence collected (lineage B.1.621.1) on December 24, 2021 in New Jersey, USA (gisaid.org). Despite this evidence that B.1.621 emerged in mid-2020, neither the WHO nor the CDC assigned an official designation to this variant until August and September of 2021, respectively^16,17^, by which time documented outbreaks of B.1.621 and sublineages had already peaked globally (**Fig. 2b**).

**Table 1:** Mu and B.1.621 sublineages likely emerged in the Americas in mid-2020. ‘Submission Date’ indicates the date on which the genome became publicly available on GISAID.

| Lineage | Collection Date | Submission Date | Location Collected |
| --- | --- | --- | --- |
| B.1.621 | Dec. 15, 2020 | Dec. 18, 2021 | Bogota, Colombia |
| B.1.621.1 | Dec. 2020* | July 19, 2021 | Florida, USA |
| B.1.621.2 | May 2, 2021 | Aug. 13, 2021 | Cesar, Colombia |
| BB.2 | Oct. 14, 2020 | Nov. 19, 2021 | Antioquia, Colombia |

\*Exact date not provided in GISAID submission

**Figure 2:**
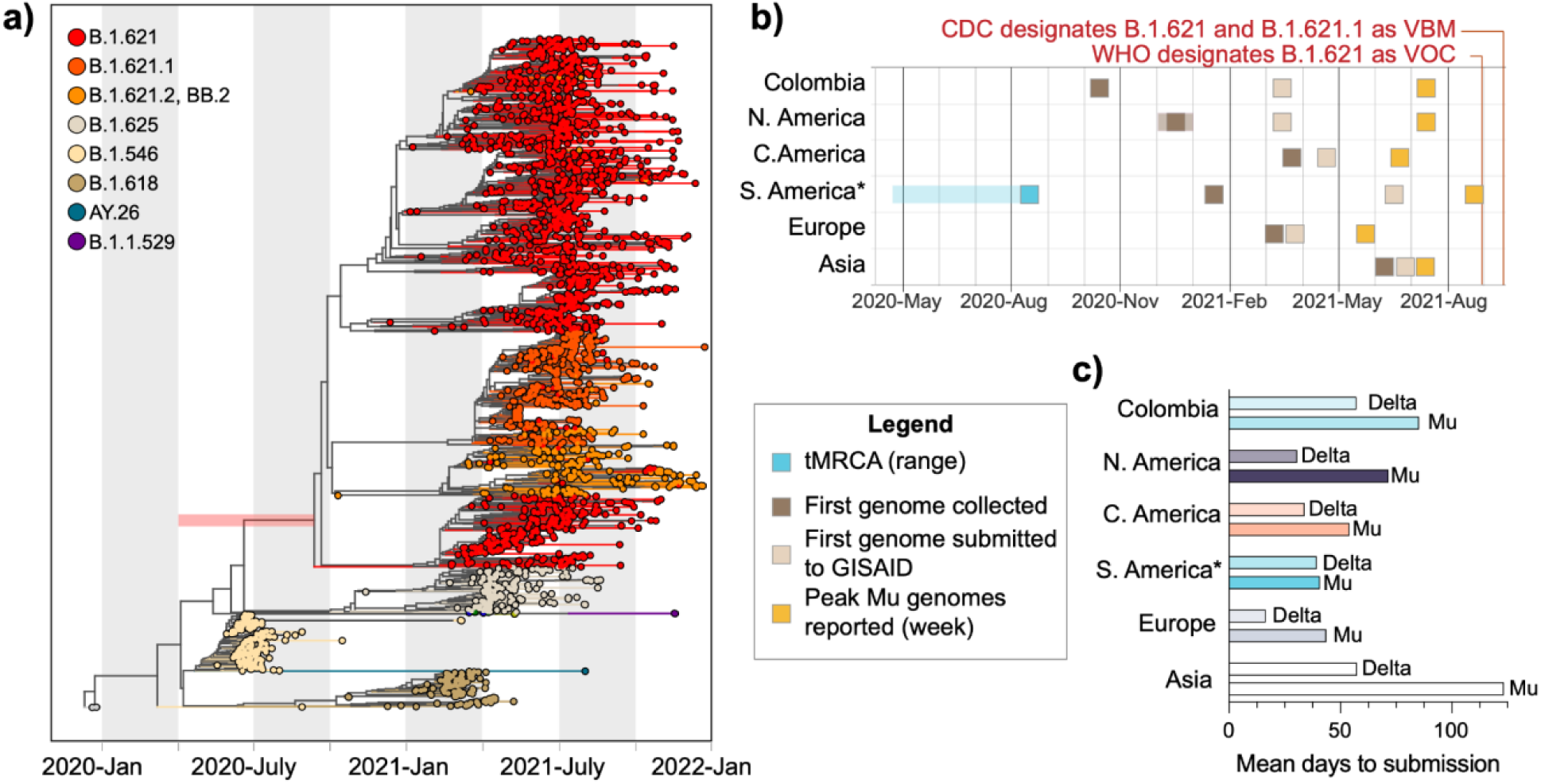
Surveillance gaps contributed to delayed reporting of Mu outbreaks. **(a)** Time-resolved phylogenetic tree generated using TreeTime. The estimated tMRCA range for Mu is indicated by the red bar. **(b)** *Excludes Colombia; transparent boxes indicate the range of possible dates. Collection and submission dates were retrieved from gisaid.org. Submission date refers to the date on which genomes became publicly available. **(c)** Exact values can be found in **Supp. Table 3**. The mean time to submission in days was calculated in R v.4.0.2.

One reason for this delay may be the initial challenges associated with publishing B.1.621 genome sequences on the public repository GISAID (gisaid.org). As described above (**Fig. 1a**), B.1.621 contains a 4 nucleotide deletion in ORF3a, which causes a frameshift mutation resulting in a premature stop codon and a truncated protein (**Supp. Fig. 2**). As a public repository, GISAID has necessarily stringent quality control measures to identify consensus genome sequences that may contain sequencing errors. As real frameshift mutations are uncommon, they are flagged as sequencing errors, the submitters are notified, and the virus genome sequences in question are not made publicly available. It is therefore the responsibility of the submitter to review rejected sequences for possible errors before resubmitting their data (see **Methods**). In overwhelmed or limited resource research settings, rejected genome sequences may not be reviewed for extended periods of time, thereby delaying the identification of new lineages with bona fide frameshift mutations. This issue arose with B.1.621, and GISAID has subsequently streamlined the quality control process by adding multiple confirmation options allowing submitters to verify that consensus sequence genomes with frameshift mutations are genuine prior to submission.

Though we cannot definitely pinpoint the precise reasons, we found that there were considerable lags between when samples were collected and when the corresponding genomes were published on GISAID. It was not until March 2021 that any B.1.621 genome sequences were made publicly available on GISAID (listed as ‘submission date’ in the repository, **Fig 2b**), and the earliest genome sequences were not made available for more than a year after their collection (**Table 1**). To quantify these delays, we measured the mean time between collection and submission for Colombia and every continent that reported cases of Mu (that is, all continents except Oceania and Antarctica). We elected to analyze countries in Central America separately. In doing so, we found that submission followed collection by more than a month on average, with South America (excluding Colombia) experiencing the shortest turn-around times (mean = 40.8 days) and Asia, represented mainly by Mongolia, experiencing the longest (mean = 123.4 days) (**Fig. 2c**). For comparison, the mean lag of Delta submissions was consistently shorter across all regions analyzed (shortest mean = 16.53 days in Europe, longest mean = 57.49 days in Asia) (**Fig. 2c**).

These extended turn-around times do not fully account for the timing of Mu’s VOC designation, but they importantly identify key gaps in the global genomic surveillance system, which in turn correspond to vulnerabilities in our public health responses. The uneven global distribution of laboratories performing genomic surveillance relative to population (**Supp. Table 2**)^23^, and therefore potential emergence of undetected SARS-CoV-2 variants, may partly explain these gaps. That is, delays in detection are more likely when variants emerge in regions with less robust surveillance infrastructure.

### The spread of the Mu clade was primarily limited to South and Central America

To further investigate this hypothesis, we next evaluated the global distribution and spread of Mu. Central and South America reported the highest proportion of sequenced Mu clade genomes, while North America and Europe experienced unevenly distributed and small waves (**Fig. 3a,b**). Four percent of genomes sequenced in South America and published on GISAID between October 14, 2020 and December 31, 2021 were attributed to the Mu clade (**Fig. 3a**). In contrast, the Mu clade constituted only 0.25% and 0.033% of SARS-CoV-2 genome sequences in North America and Europe during this time period, respectively (**Fig. 3a**). These small percentages reflect the large number of total genomes sequenced (**Supp. Table 2**), the truncated rise in frequency in these locations, and the uneven subcontinental distribution of the Mu clade.

**Figure 3:**
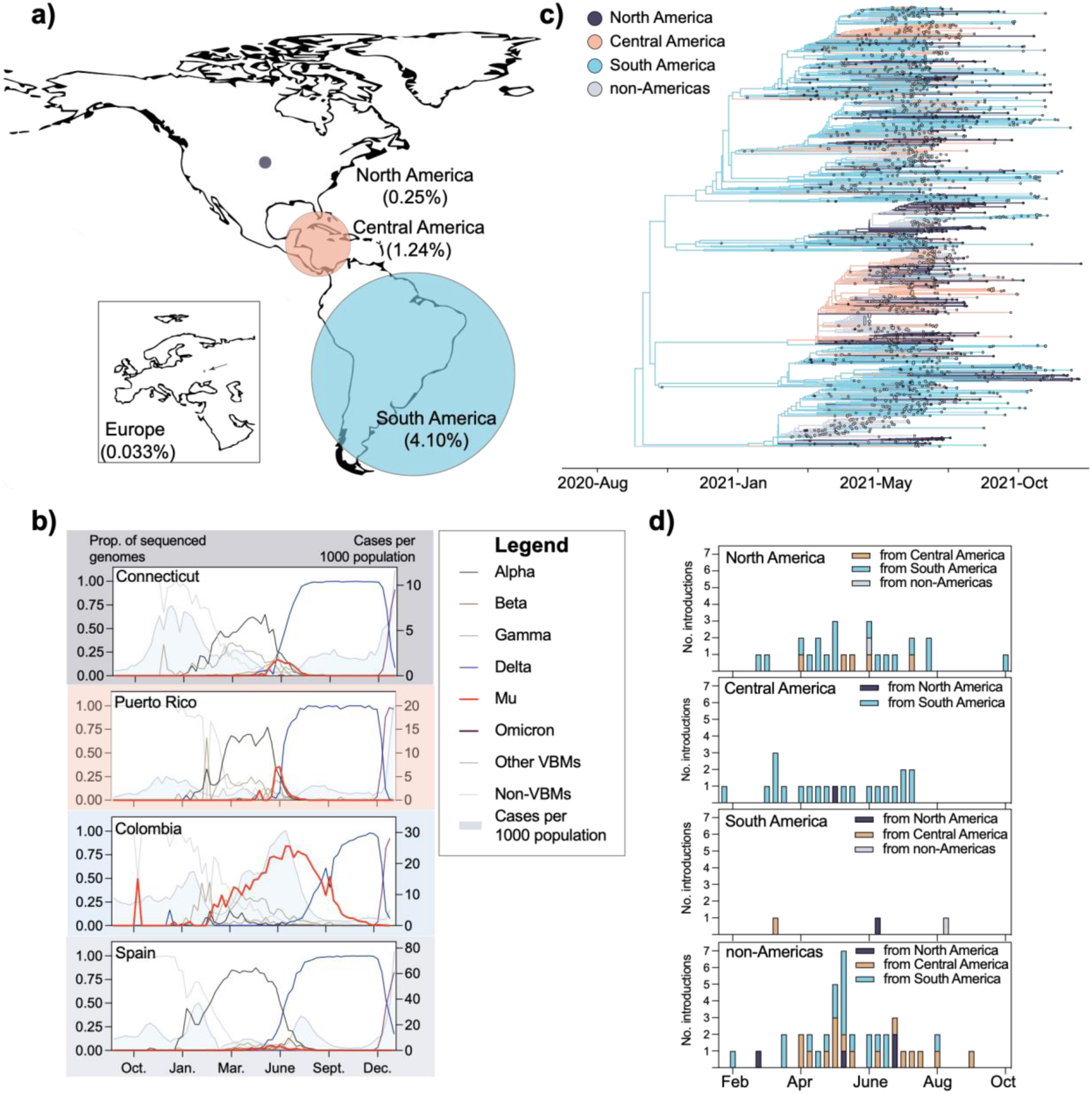
The spread of Mu was likely curtailed by the emergence of Delta in the Americas. **(a)** Percentage of Mu genomes submitted to GISAID as of December 31, 2021. **(b)** Genomic frequencies of variants in sub-continental regions. Case data were retrieved from the Johns Hopkins COVID-19 Data Repository (https://github.com/CSSEGISandData/COVID-19) **(c)** Geographically-resolved Mu clade (shown in **Fig. 2a**). The geographic location for internal nodes was inferred using BEAST v.1.10.5. **(d)** Temporal distribution of sustained introductions into continental regions. We defined sustained introductions as transitions between geographic states that resulted in clades containing at least 3 tips.

In fact, some subcontinental regions experienced a perceptible rise in the frequency of Mu clade genome sequences identified between the Alpha and Delta waves. To detect local patterns in Mu clade frequencies that deviated from those on the continental level, we tabulated the number of Mu clade genome sequences identified per week in the countries and US states that reported the highest burden of samples identified as Mu clade. We found that Connecticut and Puerto Rico experienced observable, albeit short increases in Mu clade frequencies between the Alpha and Delta waves. Colombia experienced a more substantial rise in samples identified as Mu, which reached ∼75% frequency at its peak. Though Spain reported the largest number of samples identified as Mu clade in Europe, the rise in frequency was almost imperceptible (maximum frequency = 0.05) (**Fig. 3b**).

One explanation for these inconsistent patterns is that the Mu clade did not circulate continuously through the Americas, instead spreading unidirectionally from South and Central America to North America and Europe. We initially hypothesized that the delay in detection would have enabled the Mu clade to disperse internationally and establish an intercontinental circulation pattern undetected. To test this hypothesis, we extracted the Mu clade from our time-resolved phylogenetic tree shown in **Fig. 2a** and inferred geographic transitions using a Bayesian discrete phylogeographic approach implemented in BEAST^32,33^. For simplicity, we assigned genome sequences to one of four categories: North America (n = 376), Central America (n = 342), South America (n = 698), and non-Americas (n = 538) (**Fig. 3c**). The vast majority of the genome sequences classified as ‘non-Americas’ were collected in Europe, and the remaining 31 were collected in Asia. From the geographically-resolved tree (**Fig. 3c**), we extracted sustained introductions, which we defined as transitions between two geographic categories that resulted in a clade with at least three tips.

This analysis revealed that the Mu clade spread intercontinentally from South and Central America, but was rarely re-introduced into those regions. We detected only three sustained introductions into South America during our study period. Similarly, though we detected introductions into Central America, all but one were likely from South America. These patterns are in sharp contrast to the other two geographic categories in which we detected sustained introductions almost continually through August 2021, including as many as seven introductions per week into the non-Americas region (**Fig. 3d**). They also reflect the likely origins of the Mu clade sublineages B.1.621.1, B.1.621.2, and BB.2. We inferred that B.1.621.1 likely emerged in Central America (location probability = 0.96) and B.1.621.2 and BB.2 likely emerged in South America (location probability = 1 for both) (**Fig. 3c**).

The limited geographic distribution of the Mu clade may be due to the rapid rise of the highly transmissible Delta variant. South America did not experience a large Alpha wave (**Supp. Fig. 3**), which seems to have facilitated the establishment of the Mu clade. However, even in Colombia where the Mu clade comprised the majority of sequenced genomes for weeks, the emergence of Delta coincided with a rapid decline in Mu clade prevalence. At the same time, three sublineages of B.1.621 emerged and spread between continents over a short time period, indicating that the reporting delays described above may have had an epidemiological impact.

## DISCUSSION

In this study, we combine experimental and genomic data to evaluate the surprisingly limited spread of a highly immune evasive SARS-CoV-2 variant. Genomic data are an important starting point for hypothesis generation around the biological and epidemiological characteristics of an emerging variant. Our analysis focused on Mu because its mutation profile indicated that it would exhibit elevated resistance to antibody neutralization (**Fig. 1a**). We used a neutralization assay to evaluate the immune evasive properties of Mu relative to other variants (**Fig. 1c**), but additional assays such as tissue tropism may be incorporated depending on the mutation profile of the variant under investigation. We next sought to answer important epidemiological questions using phylogenetics. First, we estimated when Mu and the B.1.621 sublineages likely emerged compared to when they were initially detected (**Fig. 2a**). Substantial lags in time to detection and reporting (**Fig. 2c**) led us to investigate the underlying bioinformatic challenges associated with Mu consensus genome sequences (**Supp. Fig. 2**). We then adopted an epidemiological approach to evaluate the global distribution (**Fig. 3a**) and spread (**Fig. 3c,d**) of this variant given its long period of cryptic transmission. Our analysis incorporated the putative weekly frequencies of individual variants (**Fig. 3b**), but more computationally rigorous approaches could be applied such as effective reproduction number estimates^34,35^. We utilized a streamlined method for phylogeographic inference33 to measure the rate of inter-continental spread. Despite its immune evasive properties and delayed detection, Mu and its B.1.621 sublineages did not circulate globally, instead spreading unidirectionally from South America to North America, Europe, and Asia (**Fig. 3c,d**). This may not be the case for future variants, at which point a more thorough assessment of circulation patterns will be needed. Together, our findings present a comprehensive (though not exhaustive) assessment of the previously designated VOC Mu.

Our findings suggest that immune evasion alone was not sufficient to outcompete other globally circulating VOCs during the second year of the COVID-19 pandemic. Due to its documented resistance to neutralization, Mu should have been able to spread even in highly vaccinated populations, but it comprised less than 10% of sequenced viruses in all but a few countries. Visual inspection of variant frequencies indicate that Mu was out-competed by Delta (**Fig. 3b**), though frequency data alone are not sufficient to assess direct competition. Interestingly, both Alpha and Delta exhibited the least reduction (1.4 and 2.5 fold, respectively) in sensitivity to neutralization in our study (**Fig. 1c**) but rose to global dominance in 2021^2–9^. This suggests that transmissibility yielded a greater advantage than immune evasion in the second year of the pandemic. The subsequent emergence of Omicron, which is both highly transmissible and immune evasive, indicates that these properties may act synergistically to drive the unprecedented global incidence rates of Omicron infections. Further analysis of this observation is needed to disentangle epidemiological and biological factors associated with variant fitness and spread.

In our study, we also highlight key gaps in the global genomic surveillance system that require immediate attention. The timeliness and accuracy of genomic data made available on public repositories is essential for driving an effective response. The genetic composition of SARS-CoV-2 lineages and rapid changes in their frequency are early indicators of an emerging VOC. As much as possible, genomic data should be made publicly available to facilitate the rapid detection of new variants; however, we recognize that there are many valid reasons why some researchers are hesitant to share their data prior to publication^36,37^. Similarly, errors in sequencing data could obfuscate genomic warning signs in addition to introducing inaccuracies in phylogenetic analyses. Therefore, it is of the utmost importance that individuals generating genomic data do so with high levels of accuracy. The WHO has provided extensive quality control standards and implementation guidelines to ensure high-quality SARS-CoV-2 genomic data^38^.

Our study was not without limitations. The analysis presented above is retrospective and therefore not explicitly actionable. Rather, our intention was to examine genomic and clinical correlates of variant fitness that may inform the response to future variants. The data we have presented should be carefully interpreted. The experimental assays were performed *in vitro*, and therefore may not directly reflect biological dynamics in vivo. However, our objective was to compare specific characteristics in individual variants, which may be accomplished *in vitro*. Genomic frequencies should be interpreted with caution due to large fluctuations in the percentage of cases sequenced by week in each location. For this reason, long-term trends are more reliable than short-term trends. Lastly, the phylogeographic analyses presented comprised substantially downsampled data, potentially biasing our findings. In particular, characterizing patterns of sustained introductions should be interpreted with understanding. That is, the relative number of introductions across regions should be considered rather than the absolute number. This challenge is in part due to the retrospective nature of our study. Downsampling is less likely to be needed when a more recently emerged variant is the subject of analysis.

Here, we present a multidisciplinary analysis of Mu, an immune evasive VOC that exhibited limited global spread. As new SARS-CoV-2 lineages emerge, their mutation profiles provide preliminary data pertaining to their potential epidemiological impact, which should be further tested using experimental and phylogenetic analyses. Due to the reliance on publicly available genomic data for the initial screening step, quality control should be a priority during consensus sequence generation. Finally, though the global health threat posed by Mu was quickly overshadowed by the threat posed by Delta and Omicron, our case study yielded insights into genetic correlates of immune evasion and surveillance gaps. As the circulating SARS-CoV-2 population continues to evolve, examining variants that came before can help us prepare for variants that will emerge in the future.

## METHODS

### Ethics

This study was approved by Yale Human Research Protection Program Institutional Review Board (IRB Protocol ID 2000028924). Informed consent was obtained from all enrolled vaccinated HCWs.

### Sampling Variants of Concern (Fig. 1b)

We randomly sampled 50 genome sequences per Variant of Concern (Alpha, Beta, Gamma, Delta, Lambda, Mu, Omicron) based on GISAID accession numbers using a random sampler implemented in R v.4.0.2.

### Frequency plots

We downloaded all metadata that were submitted to GISAID by January 19, 2022. Entries for which only the collection year was provided were removed. Entries for which only the month were provided were assigned to the 15th. We further filtered our dataset to include genome sequences collected between October 1, 2020 and December 31, 2021. We manually re-assigned genome sequences collected in Central American countries to Central America. To measure weekly frequencies of variants, we created 8 variant categories: Alpha, Beta, Gamma, Delta, Mu, Omicron, other-VBMs, non-VBMs (**Supp. Table 4**). We tabulated the number of each variant category collected by week and reported frequencies as a proportion of the total number of genome sequences collected by week by location. All analyses were performed in R v.4.0.2. We visualized the plots using Prism v.9.3.1.

### Sampling Mu clade genome sequences

We first selected all complete Mu genome sequences available on GISAID collected through December 31, 2021 (n = 14,477) as of January 8, 2022. We did not downsample genome sequences from countries or territories with less than or equal to 50 genome sequences during our study period (n = 452). For the remaining locations, we began by tabulating the weekly incident cases for each using the Johns Hopkins dashboard (https://github.com/CSSEGISandData/COVID-19) between October 2020 and December 2021. We normalized weekly incident cases per 1 million persons in the population. Next, we calculated the proportion of cases each location contributed to the total per week. We then multiplied this proportion by the number of available genome sequences per week per location to obtain the total number of genome sequences to sample (n = 1505). To improve our phylogenetic inference, we included genome sequences from closely related lineages (B.1.546, B.1.616, and B.1.625) as well as Variants of Concern (Alpha, Beta, Gamma, Delta, Omicron) (**Supp. Table 1**). For the former, we selected all complete genome sequences with high coverage collected during our study period available on GISAID. For the latter, we randomly selected 1-2 complete genome sequences. We rooted the tree using a reference genome sequence collected in Wuhan in 2019 (Wuhan/WH01/2019).

### Phylogenetic tree construction

Once fastas were downloaded from GISAID (**Supp. Table 5**), we aligned our genome sequences with MAFFT using MN908947.3 as the reference. We removed gaps and masked known problematic sites per De Maio et al. ^39^. We built our maximum likelihood trees (**Fig. 1b, Fig. 2a**) in IQTree v1.6.3 using a GTR+G substitution model and 1000 bootstraps. To obtain temporal resolution for **Fig. 2a**, we used the maximum likelihood tree and corresponding alignment as input for augur. All trees were visualized using the Python library ‘baltic’.

### Phylogeographic inference

We extracted the Mu clade from our time-resolved tree in R v.4.0.2 (**Fig. 2a**). For discrete phylogeographic reconstruction, we assigned tips to one of four categories: ‘North America’, ‘Central America’, ‘South America’, ‘non-Americas’. Genome sequences assigned to ‘non-Americas’ were primarily collected in Europe (n = 507). The remaining 31 were collected in Asia. Tips for which a complete date was not provided with the GISAID submission were assigned to the 15th of the month. We performed phylogeographic reconstruction implemented in BEAST v1.10.5 using an asymmetric CTMC model for discrete state reconstructions and running for 1 million states 32. We used Tracer v.1.7.1 to assess convergence and determined that all ESS values reached at least 200.

To count the number of sustained introductions into each geographic category, we defined a sustained introduction as a transition from a source to a sink that resulted in at least three tips. We tabulated sustained introductions per regional sink in Python v.3.8.3.

### Bioinformatic assessment of insertions and deletions in the Mu genome

The majority of SARS-CoV-2 consensus genome sequences deposited on GISAID are generated using bioinformatic pipelines that take a reference based assembly approach in lieu of a de novo assembly approach. While practical, this approach makes assumptions about genome synteny and can struggle to correctly identify structural changes in the genome, such as insertions/deletions (indels). Mu offers a unique opportunity to examine indels identified in the consensus genome sequence using read level data because it has both a 3 b.p. insertion and 4 b.p. deletion. The b.p. deletion in ORF3a causes a frameshift mutation and results in a premature stop codon. Premature stop codons can be an indication of a poor quality consensus sequence genome and are used by many databases, including GISAID, as a means for quality control. However, when multiple genome sequences from the same lineage are rejected by databases on the grounds of a truncated coding frame, further investigation is warranted.

To evaluate this, we suggest looking at the read-level data to determine if the deletion is genuine or the artifact of either laboratory or bioinformatic errors. Reference based genome assembly pipelines, such as iVar^40^ used in this example, align reads to a reference genome using BWA*41* to call variants and determine the most common nucleotide at any given position. An intermediate .bam file is produced and can be visualized in common GUI software, such as Geneious. If the deletion is genuine, it will be observed in read-level data, i.e. individual reads will not contain the deleted sequence seen in the reference genome (Supp. Fig. 2). In this case, we can see that the reference sequence “GUUA” is not contained in either forward or reverse reads aligning to this region is ORF3A. As well, we see that a 3 b.p. segment “TAC” is not observed in the reference genome, but is seen in both forward and reverse reads aligning to the region in the Spike gene. In both cases, the read-level data supports the indels observed in the consensus sequence. While not practical to do for all samples containing indels, this check is important when sequencing new SARS-CoV-2 variants to determine if indels identified in the consensus sequence are supported by the read-level data.

### Phylogenetic tree visualization

All phylogenetic trees (**Figs. 1b, 2a, 3c**) were visualized using baltic implemented in Python v.3.8.3.

### Vaccinated volunteers

Forty health care workers volunteers from the YNHH were enrolled and included in this study. The volunteers received the mRNA vaccine (Moderna or Pfizer) between November 2020 and January 2021. Plasma from vaccinated donors were collected at baseline, prior to vaccination, and 28-days post second vaccination dose. Plasma samples were collected after centrifugation of whole blood at 600 g for 20 min at room temperature (RT) without brake. The undiluted plasma was transferred to 15-ml polypropylene conical tubes, and aliquoted and stored at -80 °C for subsequent analysis. Demographic information was aggregated through a systematic review and the clinical data were collected using REDCap. None of the participants experienced serious adverse effects after vaccination.

### SARS-CoV-2 variants isolation

TMPRSS2-VeroE6 kidney epithelial cells were cultured in Dulbecco’s Modified Eagle Medium (DMEM) supplemented with 1% sodium pyruvate (NEAA) and 10% fetal bovine serum (FBS) at 37°C and 5% CO2. The cell line has been tested negative for contamination with mycoplasma. SARS-CoV-2 lineage A (USA-WA1/2020), was obtained from BEI Resources (#NR-52281). Alpha, Beta, Gamma, Delta and Mu variants were isolated from nasopharyngeal specimens. The isolates were cultured and resequenced as previously described^6,42^. In brief, samples were filtered through a 45μM filter and serially diluted from 1:50 to 1:19,531,250. The dilution was subsequently incubated with TMPRSS2-Vero E6 in a 96 well plate and adsorbed for 1 hour at 37°C. After adsorption, replacement medium was added, and cells were incubated at 37°C for up to 5 days. Supernatants from cell cultures with cytopathic effect (CPE) were collected, frozen, thawed and subjected to RT-qPCR. Fresh cultures were inoculated with the lysates as described above for viral expansion. Nucleic acid was extracted using the ThermoFisher MagMAX viral/pathogen nucleic acid isolation kit and libraries were prepared using the Illumina COVIDSeq Test RUO version. Pooled libraries were sequenced on the Illumina NovaSeq (paired-end 150) by the Yale Center for Genome Analysis. Data was analyzed and consensus genomes were generated using iVar (version 1.3.1). The resequenced genomes were submitted to NCBI (GenBank Accession numbers: ancestral lineage A = MZ468053, Alpha = MZ202178, Beta = MZ468007, Gamma = MZ202306, Delta = MZ468047, Mu = OK032350 (B.1.621) and OK032351 (B.1.621.1)). The pelleted virus was then resuspended in PBS and aliquoted for storage at -80°C. Viral titers were measured by standard plaque assay using TMPRSS2-VeroE6. Briefly, 300 µl of serial fold virus dilutions were used to infect Vero E6 cells in MEM supplemented NaHCO3, 4% FBS 0.6% Avicel RC-581. Plaques were resolved at 48h post-infection by fixing in 10% formaldehyde for 1h followed by 0.5% crystal violet in 20% ethanol staining. Plates were rinsed in water to plaques enumeration. All experiments were performed in a biosafety level 3 laboratory with approval from the Yale Environmental Health and Safety office.

### Neutralization assay

Sera from vaccinated individuals were heat treated for 30 min at 56°C. Sixfold serially diluted plasma, from 1:10 to 1:2430 were incubated with SARS-CoV-2 variants, for 1 h at 37 °C. The mixture was subsequently incubated with TMPRSS2-VeroE6 in a 12-well plate for 1h, for adsorption. Then, cells were overlayed with MEM supplemented NaHCO3, 4% FBS 0.6% Avicel mixture. Plaques were resolved at 40 h post infection by fixing in 10% formaldehyde for 1 h followed by staining in 0.5% crystal violet. All experiments were performed in parallel with baseline controls sera, in an established viral concentration to generate 60-120 plaques/well.

### Growth rate assay

Vero-E6 cells were cultured in Dulbecco’s Modified Eagle Medium (DMEM) with 5% heat-inactivated fetal bovine serum (FBS), and 1% Penicillin/Streptomycin. Calu-3 cells were cultured in Eagle’s minimal essential medium (EMEM) with 10% FBS and 1% Penicillin/Streptomycin.

To generate the viral stock of gamma used to test replication competence, 107 Vero-E6 cells overexpressing ACE2 and TMPRSS2 were infected with SARS-CoV-2 Isolate hCoV-19/Japan/TY7-503/2021 (BEI Resources #NR-54982) at an MOI of approximately 0.02. At 1 day post-infection (dpi), supernatant was collected, clarified by centrifugation (450 xg for 10 minutes), filtered through a 0.45-micron filter, and used to infect 5 × 107 Vero-E6 cells overexpressing ACE2 and TMPRSS2. At 1 dpi, supernatant was harvested, clarified, and filtered as above. Supernatant was concentrated ten-fold using Amicon Ultra-15 filters, then aliquoted for storage at -80°C. Viral stocks were titered by plaque assay in Vero-E6 cells stably overexpressing ACE2 and TMPRSS2. The cells were seeded at 5 × 105 cells/well in 12-well plates. The following day, the media was removed and replaced with 100 μl of 10-fold serial dilutions of virus. Plates were incubated at 37°C for 30 minutes with gentle rocking. Overlay media (DMEM, 2% FBS, 0.6% Avicel RC-581) was added to each well. At 2 days post-infection, plates were fixed with 10% formaldehyde for 30 minutes, stained with crystal violet solution (0.5% crystal violet in 20% ethanol) for 30 minutes, and then rinsed with deionized water to visualize plaques.

Calu-3 cells were used to test viral replication in a human cell culture model. Cells were seeded at 105 cells/well in 96-well plates. The following day, the media was removed and the cells were infected at a multiplicity of infection (MOI) of 0.1 (104 plaque forming units per well). At 1, 24, and 48 hours post-infection, 50 μl of supernatant was collected from each well and frozen at -80°C. Viable virus present in the supernatant was quantified in Vero-E6 cells stably overexpressing ACE2 and TMPRSS2 using the plaque assay described above. Three biological replicates and three technical replicates were performed.

All work with infectious virus was performed in a Biosafety Level 3 laboratory and approved by the Yale University Biosafety Committee.

## Supporting information

GISAID_Acknowledgement_Table

## Data Availability

All SARS-CoV-2 genomic data from this manuscript has been made publicly available on GISAID.

## ACKNOWLEDGEMENTS

The following reagent was obtained through BEI Resources, NIAID, NIH: SARS-Related Coronavirus 2, Isolate hCoV-19/Japan/TY7-503/2021 (Brazil P.1), NR-54982, contributed by National Institute of Infectious Diseases. This work was supported in part by a Fast Grant from Emergent Ventures at the Mercatus Center at George Mason University and the Centers for Disease Control and Prevention (CDC) Broad Agency Announcement # 75D30120C09570 awarded to NDG. CBW is funded by Burroughs Wellcome Fund and BM is funded through an NIH T32 HL 007974.

## SUPPLEMENT

**Supplementary Figure 1:**
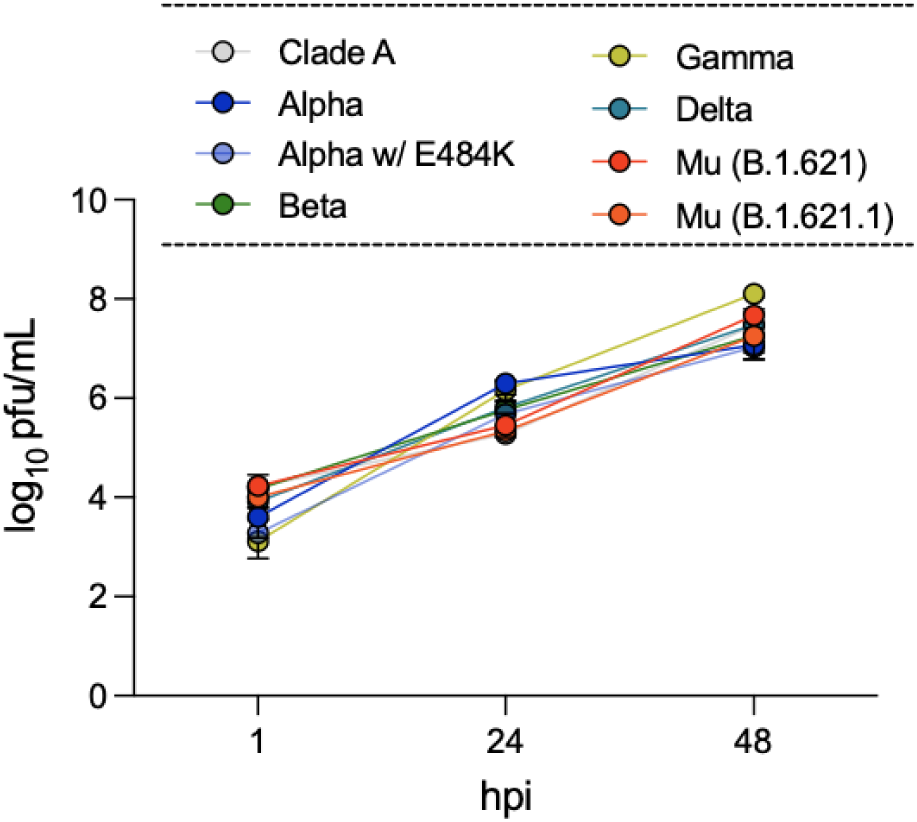
Intrinsic replication of SARS-CoV-2 VOCs in Calu-3 cells.

**Supplementary Table 1:** Lineage distribution for Figure 2a.

| Lineage | Count |
| --- | --- |
| Mu | 1957 |
| B.1.621 | 1373 |
| B.1.621.1 | 378 |
| B.1.621.2 | 62 |
| BB.2 | 144 |
| VOCs (Alpha, Beta, Gamma, Delta, Omicron) | 9 |
| B.1.546 | 211 |
| B.1.618 | 132 |
| B.1.625 | 167 |
| References | 2 |
| *Total* | *2478* |

**Supplementary Table 2:** Number of Mu and total SARS-CoV-2 genome sequences submitted to GISAID by geographic region.

| Region/Continent | No. Mu genomes reported | No. total genomes reported | Submitting labs per 1M population |
| --- | --- | --- | --- |
| North America | 6232 | 2511434 | 1.36 |
| Central America | 913 | 73699 | 0.45 |
| South America | 6676 | 162954 | 0.54 |
| Europe | 1197 | 3629925 | 1.95 |
| Africa | 1 | 70808 | 0.16 |
| Asia | 31 | 450875 | 0.12 |

**Supplementary Table 3:** Values for Fig. 2c.

| Region | Mean days to submission (Mu) | Mean days to submission (Delta) |
| --- | --- | --- |
| North America | 71.78709 | 30.72636 |
| Central America | 54.13363 | 34.0446 |
| South America | 42.30501 | 39.44826 |
| Colombia | 85.41713 | 57.31864 |
| Europe | 43.68212 | 16.52821 |
| Asia | 123.41935 | 57.74474 |

**Supplementary Figure 2:**
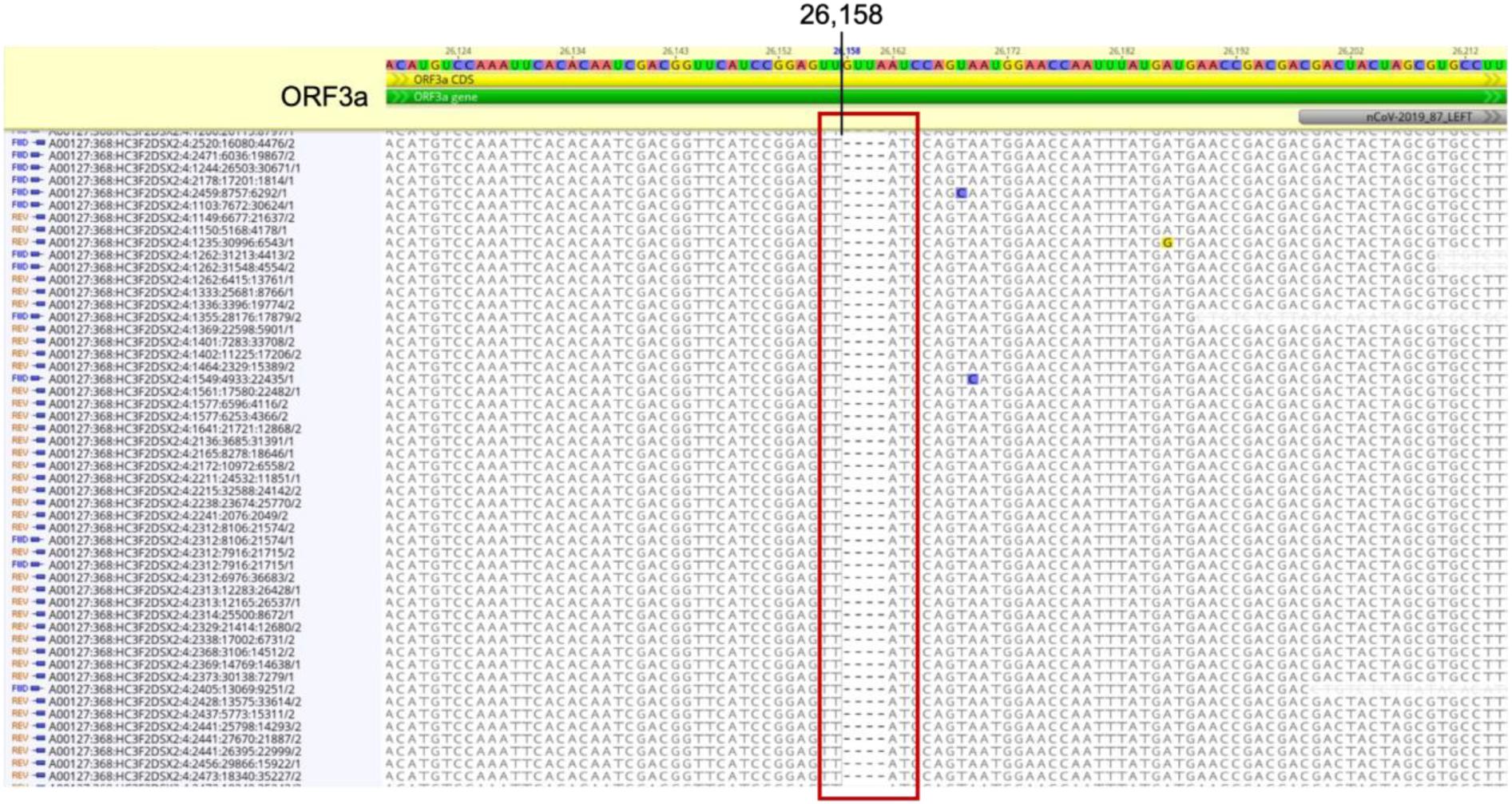
Frameshift in ORF3 results in a premature stop codon in the Mu genome.

**Supplementary Figure 3:**
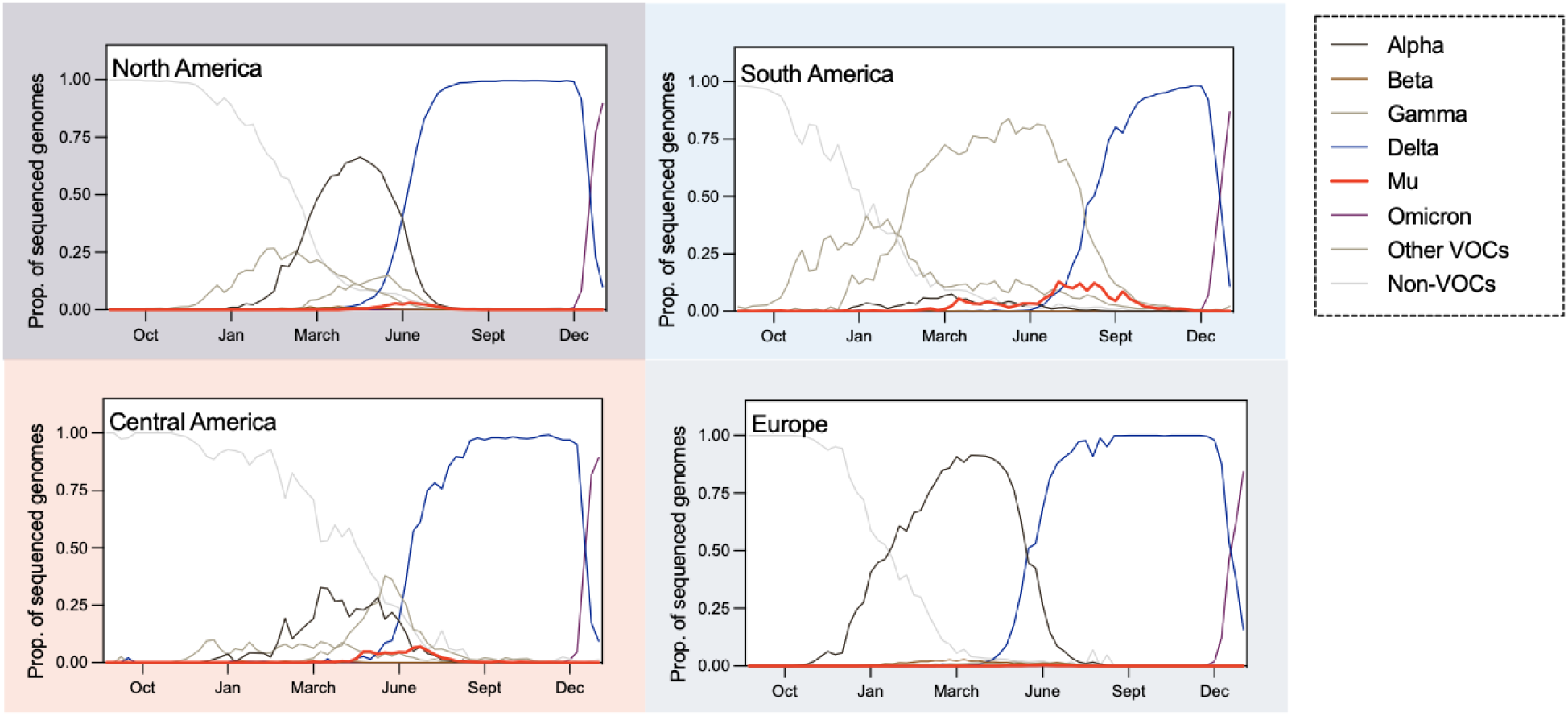
Time-dependent variant frequencies by continent. Variant frequencies between October 2020 and December 2021 based on genome sequences available on GISAID.org as of January 19, 2022. Graphs rendered in Prism v.9.3.1.

**Supplementary Table 4:** Lineages assigned to variant categories for frequency plots.

| Variant category | Lineages |
| --- | --- |
| Alpha | B.1.1.7, Q.X |
| Beta | B.1.351, B.1.351.X |
| Gamma | P.1, P.1.1, P.1.2 |
| Delta | B.1.617.2, AY.X |
| Mu | B.1.621, B.1.621.X, BB.X |
| Omicron | B.1.1.529, BA.X |
| other-VBMs | B.1.427, B.1.429, B.1.525, B.1.526,<br>B.1.617.1, B.1.617.3, P.2, C.37, C.37.1 |
| non-VBMs | Remaining lineages not listed above |

