## Supplementary material for "Insights into the limited global spread of the immune evasive SARS-CoV-2 variant Mu": GISAID_Acknowledgement_Table

We gratefully acknowledge the following Authors from the Originating laboratories responsible for obtaining the specimens, as well as the Submitting laboratories where the genome data were generated and shared via GISAID, on which this research is based.

All Submitters of data may be contacted directly via www.gisaid.org

Authors are sorted alphabetically.

Acknowledgement EPI_SET Identifier: EPI_SET_20220328ct

| Accession ID | Originating Laboratory | Submitting Laboratory | Authors |
|---|---|---|---|
| EPI_ISL_7987914 | 4Cyte Pathology | NSW Health Pathology - Institute of Clinical Pathology and Medical Research; Westmead Hospital; University of Sydney | Arnott A.; Draper J.; Gall M.; Martinez E.; Rockett R.; Sintchenko V.; on behalf of ICPMR |
| EPI_ISL_2345429 | AMBULATORIO DE ESPECIALIDADE V E MOGI MIRIM | Instituto Butantan / ESALQ-Piracicaba | Antonio Jorge Martins; Claudia Renata dos Santos Barros; David Schlesinger; Debora Botequio Moretti; Dimas Tadeu Covas; Elaine Cristina Marqueze; Elaine Vieira Santos; Evandra Strazza Rodrigues; Heidge Fukumasu; Jayme Augusto de Souza-Neto; José Salvatore Leister Patané; Luiz Alcantara; Luiz Lehmann Coutinho; Maria Carolina Elias; Maurício Lacerda Nogueira; Rafael dos Santos Bezerra; Raul Machado Neto; Rejane Maria Tommasini Grotto; Ricardo Haddad; Sandra Coccuzzo Sampaio Vessoni; Simone Kashima; Svetoslav Nanev Slavov; Vincent Louis Viala |
| EPI_ISL_1686547, EPI_ISL_1996455, EPI_ISL_2181206, EPI_ISL_2181383, EPI_ISL_3430367, EPI_ISL_3845629, EPI_ISL_3853010, EPI_ISL_4186127, EPI_ISL_4360363, EPI_ISL_4397844, EPI_ISL_8082202 | | | |
| see above | Aegis Sciences Corporation | Centers for Disease Control and Prevention Division of Viral Diseases, Pathogen Discovery | Adrian Paskey; Alec Vest; Benjamin Rambo-Martin; Christopher Gulvick; Clinton Paden; Clinton R. Paden; Cyndi Clark; Dakota Howard; Darlene Wagner; Dhwani Batra; Dillon Nall; Duncan MacCannell; Erisa Sula; Ethan Sanders; Holly Houdeshell; Jason Caravas; Kara Moser; Kristine Lacek; Matthew Hardison; Matthew Schmerer; Ola Kvalvaag; Patrick Campbell; Peter Cook; Peter W. Cook; Rob Case; Scott Sammons; Shatavia Morrison; Shaun Westlund; Tymeckia Kendall; Victoria Caban Figueroa; Vikramsinha Ghorpade; Yvette Unoarumhi |
| EPI_ISL_5433572 | Akademiska Sjukhuset, Clinical Microbiology and Hospital Hygiene | Uppsala University Hospital | Jonathan Haars; Julia Bergholm; Patrik Ellström; Rene Kaden; Steinar Mannsverk |
| EPI_ISL_1823038 | Altius Institute | Seattle Flu Study | Alex Nguyen; Amanda Adler; Andrew Meuser; Barry R. Lutz; Benjamin Pelle; Caitlin R. Wolf; Chris D. Frazar; Clem Green; Daniel Bates; Deborah A. Nickerson; Elisabeth Brandstetter; Erica Ryke; Hannah Petersen; Helen Y. Chu; Jacob Rodriguez; Janet A. Englund; Jay Shendure; Jessica Halow; John Stamatoyannopoulos; Joshua Richards; Jover Lee; Julia Wald; Kairsten Fay; Kirsten Lacombe; Kneshay Harper; Lea M. Starita; Mark J. Rieder; Matt Hartman; Matthew Richardson; Matthew Thompson; Melissa Truong; Michael Boeckh; Michael Famulare; Misja Ilcisin; Muhammad Halimun; Olivia Waltner; Peter D. Han; Rebecca Bruders; Ryan Alexander; Sadie Patraw; Sofia Olsson; Stephanie DeBaun; Thomas R. Sibley; Tobias Ragoczy; Trevor Bedford; Truong Nguyen |
| EPI_ISL_2438065 | Ayudas diagnosticas SURA | Universidad Nacional de Colombia - Laboratorio Genómico One Health | Andres F. Cardona-Rios; Carlos Franco-Muñoz; Carolina Muñoz-Arango; Celeny Ortiz; Daniel O. Maldonado-Perez; Diego A. Álvarez-Díaz; Hector Alejandro Ruiz-Moreno; Idabely Betancur Ortiz; Jorge E. Osorio; Juan P. Hernandez-Ortiz; Karl A Ciuoderis; Katherine Laiton-Donato; Laura Silvana Perez; Lina M. Hurtado; Marcela Mercado-Reyes; Maria Angélica Maya; Maria Stella López; Rita Almanza Payares; Sandra Ines Cano; Simón Villegas Velásquez |
| EPI_ISL_2528253, EPI_ISL_2721964 | BCCDC Public Health Laboratory | BCCDC Public Health Laboratory | Ana Pacagnella; Corrinne Ng; Dan Fornika; John Tyson; Kim Macdonald; Kimia Kamelian; Linda Hoang; Loretta Janz; Mel Krajden; Prystajecky Natalie; Robert Azana; Shannon Russell |
| EPI_ISL_5648490, EPI_ISL_7779341 | BIOFAST | Instituto Butantan | Antonio Jorge Martins; Claudia Renata dos Santos Barros; David Schlesinger; Debora Botequio Moretti; Dimas Tadeu Covas; Elaine Cristina Marqueze; Elaine Vieira Santos; Evandra Strazza Rodrigues; Heidge Fukumasu; Jayme Augusto de Souza-Neto; José Salvatore Leister Patané; Luiz Alcantara; Luiz Lehmann Coutinho; Maria Carolina Elias; Maurício Lacerda Nogueira; Rafael dos Santos Bezerra; Raul Machado Neto; Rejane Maria Tommasini Grotto; Ricardo Haddad; Sandra Coccuzzo Sampaio Vessoni; Simone Kashima; Svetoslav Nanev Slavov; Vincent Louis Viala |
| EPI_ISL_2933472 | BIOMNIS LYON | CNR Virus des Infections Respiratoires - France SUD | Antonin Bal; Bruno Lina; Gregory Destras; Gwendolyne Burfin; Hadrien Regue; Laurence Josset; Martine Valette; Quentin Semanas |
| EPI_ISL_1157306, EPI_ISL_1357210 | Bayerisches Landesamt für Gesundheit und Lebensmittelsicherheit (LGL) | Robert Koch Institute | |
| EPI_ISL_6619025, EPI_ISL_7893435, EPI_ISL_8086551 | Berkshire and Surrey Pathology Services Lighthouse Laboratory | Wellcome Sanger Institute for the COVID-19 Genomics UK (COG-UK) Consortium | Berkshire and Surrey Pathology Services Lighthouse Laboratory and Alex Alderton; Cordelia Langford; David K. Jackson; Dominic Kwiatkowski; Ewan Harrison; Ian Johnston; Jeffrey Barrett; John Sillitoe on behalf of the Wellcome Sanger Institute COVID-19 Surveillance Team; Roberto Amato; Sonia Goncalves |
| EPI_ISL_6809523 | BioneXt Lab | Laboratoire national de sante, Microbiology, Microbial Genomics Platform | Anke Wienecke-Baldacchino; Catherine Ragimbeau; Elodie Solarino; Fatu Djabi; Jessica Tapp; Lise Pignon; Raoul Salmon; Tamir Abdelrahman; Thibault Ferrandon; Virginie Jover |
| EPI_ISL_5502548 | Botswana Harvard HIV Reference Laboratory | Botswana Harvard HIV Reference Laboratory | Boitumelo J. L Zuze; Botshelo Radibe; Dorcas Maruapula; Joseph Makhema; Keoratile Ntshambiwa; Kgomotso Moruisi; Legodile Koopile; Letsibogo Gaoraelwe; Mosepele Mosepele; Mphaphi B. Mbulawa; Ontlametse T. Bareng; Pamela Smith-Lawrence; Roger Shapiro; Sefetogi Ramaologa; Shahin Lockman; Sikhulile Moyo; Simani Gaseitsiwe; Thela Tefelo; Thongbotho Mphoyakgosi; Wonderful T. Choga |
| EPI_ISL_3481365, EPI_ISL_3487617 | British Columbia Centre For Disease Control | BCCDC Public Health Laboratory | Ana Pacagnella; Corrinne Ng; Dan Fornika; John Tyson; Kim Macdonald; Kimia Kamelian; Linda Hoang; Loretta Janz; Mel Krajden; Prystajecky Natalie; Robert Azana; Shannon Russell |
| EPI_ISL_3007338, EPI_ISL_8214111 | Broad Institute Clinical Research Sequencing Platform | Infectious Disease Program, Broad Institute of Harvard and MIT | Adams, G.; B.L.; B.W.; Bauer, M.; Birren; Blumenstiel, B.; Brown, C.; Carter, A.; Chaluvadi, S.; D.J.; DeFelice, M.; DeRuff, K.; Dodge, S.; Gabriel, S.; Gallagher, G.; Gladden-Young, A.; Granger, B.; J.E.; K.J.; Lagerborg, K.; Larkin, K.; Lee, M.; Lemieux; Lennon, N.; Loreth, C.; Madoff, L.; McGovern, S.; Meldrim, J.; Normandin, E.; P.C.; Park; Pearlman, L.; Reilly, S.; Rudy, M.; Sabeti; Siddle; Smole, S.; Tomkins-Tinch, C.; Vicente, G.; and MacInnis |
| EPI_ISL_7747512 | CAPRISA_HILLCREST | CERI, Centre for Epidemic Response and Innvoation, Stellenbosch University and KRISP, KZN Research Innovation and Sequencing Platform, UKZN. | Aida Sivro; Arisha Maharaj; Giandhari J; Naidoo Y; Natasha Samsunder; Pillay S; Ramphal U; Ramphal Y; San JE; Tegally H; Tshiabuila D; Wilkinson E; de Oliveira T |
| EPI_ISL_5016681, EPI_ISL_7130811, EPI_ISL_7335880, EPI_ISL_7703912 | CDPH VBL | California Department of Public Health | Emily Smith on behalf of CDPH-COVIDNet |
| EPI_ISL_1628966 | CERBALLIANCE | UMR PIMIT | Dr Camille Lebarbenchon; Dr David A Wilkinson; Dr Patrick Mavingui; Magali Turpin |
| EPI_ISL_4605097 | CERBALLIANCE REUNION | CNR Virus des Infections Respiratoires - France SUD | Antonin Bal; Bruno Lina; Gregory Destras; Gwendolyne Burfin; Hadrien Regue; Laurence Josset; Martine Valette; Quentin Semanas |
| EPI_ISL_3347375 | CT Department of Public Health | Grubaugh Lab - Yale School of Public Health | Anderson Brito; Annie Watkins; Chaney Kalinich; Chantal Vogels; Claire Pearson; Isabel Ott; Jessica Rothman; Joseph Fauver; Kendall Billig; Mallery Breban; Mary Petrone; Nathan Grubaugh; Tara Alpert; Tobias Koch; Tu N. Nguyen |
| EPI_ISL_2688697 | Centre Hospitalier Eure Seine | Centre Hospitalier Universitaire de Rouen Laboratoire de Virologie | Alice Moisan; Fabienne De Oliveira; Marie Leoz |
| EPI_ISL_2467244, EPI_ISL_3242620, EPI_ISL_3242739 | Clinica INDISA | "Facultad de Ciencias de la Vida, UNAB" | "Claudio Meneses; Ariel Orellana"; Claudio Olmos; Daniel Leon; Dayan Sanhueza; Eduardo Castro; Gonzalo Campaña; Macarena Bastias; Paola Pidal; Ricardo Yusta; Sebastian Wolter; Susana Saez; Víctor Monreal; Waldo Diaz |
| EPI_ISL_8206945 | Clinical Microbiology Laboratory, Tel Aviv Sourasky Medical Center | Clinical Microbiology Laboratory, Tel Aviv Sourasky Medical Center | Alon Ziv; Amos Adler; Katya Levytskyi; Lior Handler; Ora Halutz |
| EPI_ISL_1664321 | Clinical Molecular Microbiology Laboratory, UNC Hospitals | Jeremy Wang | Alexander Rubinsteyn; Colleen Rice; Corbin Jones; Jason Smedberg; Jeremy Wang; Melissa Miller; Robert Hagan; Shawn Hawken |
| EPI_ISL_3261639 | Cliniques universitaires Saint-Luc | UCLouvain/IREC/MBLG | Benoit Kabamba Mukadi; Bertrand Bearzatto; Jean Ruelle |
| EPI_ISL_4634562 | Colorado Department of Public Health and Environment | Colorado Department of Public Health and Environment | Alexandria Rossheim; Diana Ir; Emily A. Travanty; Laura Bankers; Mandy Waters; Michael Martin; Molly C. Hetherington-Rauth; Sarah Elizabeth Totten; Shannon R. Matzinger |
| EPI_ISL_1817733 | Conville CDC wc CVC | NHLS/UCT | Arash Iranzadeh; Bruna Galvao; Carolyn Williamson; Deelan Doolabh; Diana Hardie; Innocent Mudau; Kruger Marais; Lynn Tyers; Marvin Hsiao; Stephen Korsman |
| EPI_ISL_4415852, EPI_ISL_4422568 | DASA | DASA | Adriano Bonaldi; Angelica Hristov; Annelise Lopes; Bianca Cota; Cristina Oliveira; Jose Levi; Lidia Yamamoto; Paulo Pierry; Rodrigo Guarischi; Rodrigo Salazar |
| EPI_ISL_3053801, EPI_ISL_8136866, EPI_ISL_8171714 | Department of Bacteria, Parasites and Fungi, Statens Serum Institut, Copenhagen, Denmark | Statens Serum Institut Bioinformatics and Microbial Genomics | Danish Covid-19 Genome Consortium |
| EPI_ISL_2811147, EPI_ISL_3872731 | Department of Infectious Diseases, Kobe Institute of Health | Department of Infectious Diseases, Kobe Institute of Health | Kentaro Itokawa; Makoto Kuroda; Masanori Hashino; Noriko Nakanishi; Rina Tanaka; Ryohei Nomoto; Tomotada Iwamoto; Tsuyoshi Sekizuka |
| EPI_ISL_1310632 | Department of Pathology, University of Cambridge | COVID-19 Genomics UK (COG-UK) Consortium | Aminu S. Jahun; Ian Goodfellow; Iliana Georgana; Martin D. Curran; Myra Hosmillo; Rhys Izuagbe; Surendra Parmar; William L. Hamilton; Yasmin Chaudhry |
| EPI_ISL_1876636 | Department of Virus and Microbiological Special Diagnostics, Statens Serum Institut, Copenhagen, Denmark | Aalborg University | Danish Covid-19 Genome Consortium |
| EPI_ISL_1820249 | Dept. of Medical Microbiology, Stavanger University Hospital, Helse Stavanger HF | Norwegian Institute of Public Health, Department of Virology | Atiya R Ali; Debech Nadia; Engebretsen Serina Beate; Garcia Llorente Ignacio; Hilde Elshaug; Hilde Vollan; Jon Bråte; Kamilla Heddeland Instefjord; Karoline Bragstad; Kathrine Stene-Johansen; Marie Paulsen Madsen; Olav Hungnes; Pedersen Benedikte Nevjen; Rasmus Riis Kopperud |
| EPI_ISL_1089695, EPI_ISL_1455691, EPI_ISL_1962271, | Dutch COVID-19 response team | National Institute for Public Health and the Environment (RIVM) | Adam Meijer; AnneMarie van den Brandt; Annelies Kroneman; Bas van der Veer; Chantal Reusken; Dennis Schmitz; Dirk Eggink; Euníce Then; Florian Zwagemaker; Harry Vennema; James Groot; Jeroen Cremer; Jolienke Hardeman; Karim Hajji; Kim Freriks; Linda van de Nes; Lisa Wijsman; Lynn Aarts; Melissa van Tuil; Robert Kohl; Ryanne Jaarsma; Sanne Bos; Sharon van den Brink; Sjoerd Kuiling; on behalf of the national COVID-19 response team |

| | | | |
|---|---|---|---|
| EPI_ISL_3057773 | | | |
| EPI_ISL_2370486 | ETHNIKO KENTRO AIMODOSIAS E.KE.A. | Greek Genome Center, Biomedical Research Foundation of the Academy of Athens (BRFAA) | Dimitrios Thanos; Efthimia Petinaki; Emmanouil Athanasiadis; Giannis Vatsellas; Katerina Zoi; Kostas Stamoulis; Theodoros Loupis |
| EPI_ISL_2477754 | Edmonton Provincial Lab | Public Health Agency of Canada (PHAC) National Microbiology Laboratory | Buss; Croxen M; Deo A; Dieu P; E; Ferrato C; Gill K; Khan F; Koleva P; Li V; Lloyd C; Lynch T; Ma R; Murphy S; Pabbaraju K; Shokoples S; Thayer J; Tipples G; Whitehouse M; Wong A; Yu C; Zelyas N |
| EPI_ISL_1571313 | Eurofins LifeCodexx GmbH | Robert Koch Institute | |
| EPI_ISL_4228722 | Eurofins MVZ Labor Gelsenkirchen | Robert Koch Institute | |
| EPI_ISL_1663231 | Fimlab Laboratories | Fimlab Laboratories | Bruno Luukinen; Leena Huhti; Mauri Keinänen; Minna Paloniemi; Sara Lehtinen; Tapio Seiskari |
| EPI_ISL_3100936, EPI_ISL_6049914 | Florida Bureau of Public Health Laboratories | Florida Bureau of Public Health Laboratories | Jason Blanton; Namratha Tarigopula; Sarah Schmedes; Tiffany Splatt |
| EPI_ISL_1570873 | Fraunhofer-Institut fÃ¼r Zelltherapie und Immunologie | Robert Koch Institute | |
| EPI_ISL_1667031, EPI_ISL_3305650, EPI_ISL_3609503, EPI_ISL_4369182, EPI_ISL_4369471, EPI_ISL_4371980, EPI_ISL_8081073 | | | |
| see above | Fulgent Genetics | Centers for Disease Control and Prevention Division of Viral Diseases, Pathogen Discovery | Adrian Paskey; Becky Tsai; Benafsh Sapra; Benjamin Rambo-Martin; Christopher Gulvick; Clinton Paden; Clinton R. Paden; Dakota Howard; Darlene Wagner; Dhwani Batra; Doreen Ng; Duncan MacCannell; Erisa Sula; Harry Gao; James Xie; Jason Caravas; John Gao; Joseph Fierro; Kara Moser; Kristine Lacek; Matthew Schmerer; Mickey Li; Peter Cook; Peter W. Cook; Scott Sammons; Shatavia Morrison; Tymeckia Kendall; Victoria Caban Figueroa; Yan Meng; Yvette Unoarumhi |
| EPI_ISL_5797270 | Fundacion Valle del Lili | Laboratorio de biotecnología, Universidad Icesi | "María I. Gutiérrez López; Adrián Camilo Rodríguez Ararat; Diana M. Florez Giraldo; Marcela Mercado; Maria F. Villegas Torres; Paola A. Caicedo Burbano"; Programa Nacional de Caracterización Genómica de SARS-CoV-2; Sara González Henao |
| EPI_ISL_4566162 | Gencore - Universidad de los Andes | Gencore - Universidad de los Andes | Cristian Barrera; David González; Gabriela Ariza; Luisa Sacristan; Marcela Guevara; Marcela Mercado; Silvia Restrepo |
| EPI_ISL_406798 | General Hospital of Central Theater Command of People's Liberation Army of China | BGI & Institute of Microbiology, Chinese Academy of Sciences & Shandong First Medical University & Shandong Academy of Medical Sciences & General Hospital of Central Theater Command of People's Liberation Army of China | Weifeng Shi and Zhenhong Hu; Weijun Chen; Yuhai Bi |
| EPI_ISL_1633527, EPI_ISL_2009197, EPI_ISL_2009624, EPI_ISL_2391368, EPI_ISL_2756096, EPI_ISL_2756124, EPI_ISL_2896151, EPI_ISL_3132696, EPI_ISL_3536127, EPI_ISL_4005118, EPI_ISL_4655970, EPI_ISL_5010817, EPI_ISL_5011403 | | | |
| see above | Genetica Molecular and Subdepartamento de Virologia ISP Chile | Instituto de Salud Publica de Chile | Andres Castillo; Barbara Parra; Constanza Campano; Gisselle Barra; Jaime Lagos; Javier Tognarelli; Jorge Fernandez; Karen Orostica; Loredana Arata; Patricia Bustos; Rodrigo Fasce; Soledad Ulloa |
| EPI_ISL_2002482 | Genomic Research Lab, BCSIR | Genomic Research Lab, BCSIR | Abu Sayeed Mohammad Mahmud; Barna Goswami; Eshrar Osman; Iffat Jahan; Md. Ahasan Habib; Md. Kamrul Islam; Md. Murshed Hasan Sarkar; Md. Saddam Hossain; Md. Salim Khan; Mohammad Mohi Uddin; Mohammad Samir Uzzaman; Shahina Akter; Tanjina Akhter Banu |
| EPI_ISL_5603127 | Glencore Rhovan mine | National Institute for Communicable Diseases of the National Health Laboratory Service | Amoako DG; Bhiman JN; Everatt J; Ismail A; Mahlangu B; Mnguni A; Mohale T; Ntuli N; Scheepers C |
| EPI_ISL_5853660 | H Divino Espirito Santo - Ponta Delgada | Instituto Nacional de Saude (INSA) | Borges et al |
| EPI_ISL_3102412 | HOSPITAL E MATERNIDADE DRA ZILDA ARNS NEUMANN | Analytical Competence Molecular Epidemiology Lab/ACME, Oswaldo Cruz Foundation, Ceara (FIOCRUZ CE) | Cleber Furtado Aksenen; Fabio Miyajima; Fernando Braga Stehling; Francisco Eder de Moura Lopes; Jamille Maria Mendes Bezerra; Joaquim César do Nascimento Sousa Junior; Pedro Miguel Carneiro Jeronimo; Suzana Porto Almeida e Lucas Delerino; Thais Ferreira de Oliveira; Thais de Oliveira Costa; Ticiane Cavalcante de Souza; Veridiana Pessoa Miyajima |
| EPI_ISL_6481085 | Helix | Centers for Disease Control and Prevention Division of Viral Diseases, Pathogen Discovery | Benjamin Rambo-Martin; Christopher Gulvick; Clinton Paden; Dakota Howard; Dhwani Batra; Duncan MacCannell; Erisa Sula; Helix CA; Jason Caravas; Kristine Lacek; Matthew Schmerer; Peter Cook; Scott Sammons; Shatavia Morrison; Tymeckia Kendall; Victoria Caban Figueroa; Yvette Unoarumhi |
| EPI_ISL_2320917, EPI_ISL_2599383, EPI_ISL_4346907, EPI_ISL_4347689 | Helix/Illumina | Centers for Disease Control and Prevention Division of Viral Diseases, Pathogen Discovery | Adrian Paskey; Alexandre Bolze; Ary Ascencio; Benjamin Rambo-Martin; Brad Sickler; Charlotte Rivera-Garcia; Christine Tran; Christopher Gulvick; Chrstine Tran; Clinton Paden; Clinton R. Paden; Dakota Howard; Darlene Wagner; David Becker; Dhwani Batra; Duncan MacCannell; Efren Sandoval; Eileen De Feo; Eileen de Feo; Elizabeth Cirulli; Eric Allen; Geraint Levan; James Lu; Jan Antico; Jason Caravas; Jason Nguyen; Jimmy Ramirez; Jingtao Liu; Kara Moser; Kelly Barrett; Kelly Schiabor Barrett; Kim Gietzen; Kristine Lacek; Magnus Isaksson; Marc Laurent; Matthew Schmerer; Matthew Tolentino; Nicole L. Washington; Nicole Washington; Peter Cook; Peter W. Cook; Phil Febbo; Ryan Cho; Scott Sammons; Shannon Wickline; Shatavia Morrison; Sherry Wang; Simon White; Tyler Cassens; William Lee; Yvette Unoarumhi |
| EPI_ISL_3505986 | Hospital General Universitario Gregorio Marañón | Hospital General Universitario Gregorio Marañón | Cristina Rodriguez-Grande; Darío García de Viedma; Julia Suárez; Laura Pérez-Lago; Marta Herranz Martin; Patricia Muñoz; Pedro Sola Campoy; Pilar Catalán; Sergio Buenestado Serrano; Victor Manuel de la Cueva |
| EPI_ISL_3216196 | Hospital General Universitario de Alicante - Instituto de Investigación Sanitaria y Biomédica de Alicante | SeqCOVID-SPAIN consortium/IBV(CSIC) | Carmen Molina Pardines and SeqCOVID-SPAIN consortium; Maripaz Ventero Martín |
| EPI_ISL_2600860 | Hospital Universitari i Politècnic La Fe de València | SeqCOVID-SPAIN consortium/IBV(CSIC) | Ana Gil Brusola; Eva González Barberá; José Luis López Hontangas and SeqCOVID-SPAIN consortium; María Dolores Gómez Ruiz; Salvador Giner Almaraz |
| EPI_ISL_7307141 | Hospital Universitario San Ignacio | Centro de Investigaciones en Microbiología y Biotecnología-UR (CIMBIUR), Facultad de Ciencias Naturales, Universidad del Rosario, Bogotá, Colombia | Alberto Paniz-Mondolfi; Angie Ramírez; Beatriz Ariza; Camilo A. Correa-Cárdenas; Carlos Gómez-Restrepo; Claudia Cardozo-Romero; Claudia Méndez; David-Santiago Quevedo; Guido España; Hernando Díaz; Juan David Ramírez; Juliana Cuervo-Rojas; Julie Pérez; Luz H. Patiño; Manuel-Antonio Franco; Maria-Clara Duque; Marina Muñoz; Nathalia Ballesteros; Nicolas Luna; Sergio Castañeda; Zulma M. Cucunubá |
| EPI_ISL_1755806 | Hôpital Avicenne | Department of Virology, Henri Mondor University Hospital, Assistance Publique Hôpitaux de Paris, Université Paris-Est Créteil, INSERM U955 | Alexandre Soulier; Christophe Rodriguez; Elisabeth Trawinski; Guillaume Gricourt; Jean-Michel Pawlotsky; Melissa N'Debi; Slim Fourati; Vanessa Demontant |
| EPI_ISL_2637420 | Hôpital Pitié-Salpêtrière | Department of Virology, Henri Mondor University Hospital, Assistance Publique Hôpitaux de Paris, Université Paris-Est Créteil, INSERM U955 | Alexandre Soulier; Christophe Rodriguez; Elisabeth Trawinski; Guillaume Gricourt; Jean-Michel Pawlotsky; Melissa N'Debi; Slim Fourati; Vanessa Demontant |
| EPI_ISL_7314430 | IDIME | Instituto Nacional de Salud- Dirección de Investigación en Salud Pública | Beatriz de Arco; Carlos Franco-Muñoz; Diego A. Álvarez-Díaz; Diego Andrés Prada; Dioselina Peláez-Carvajal; Gerardo Santamaría; Héctor Alejandro Ruiz-Moreno; Jhonnatan Reales-González; Jorge Rivera; Julián Naizaque; Katherine Laiton-Donato; Marcela Mercado-Reyes.; Martha Lucia Ospina Martínez; María T. Herrera-Sepúlveda; Paola Rojas-Estevez; Sheryll Corchuelo; Tatiana Cobos |
| EPI_ISL_7221763 | INSACOG-IGIB and Kerala State SARS-COV-2 Genome Surveillance Programme (GENESCOV2) | INSACOG and GENESCOV2 at CSIR Institute of Genomics and Integrative Biology | INSACOG-IGIB and Kerala State SARS-COV-2 Genome Surveillance Programme (GENESCOV2) |
| EPI_ISL_5647578, EPI_ISL_5653077 | INSIDE DIAGNOSTICOS | Instituto Butantan | Antonio Jorge Martins; Claudia Renata dos Santos Barros; David Schlesinger; Debora Botequio Moretti; Dimas Tadeu Covas; Elaine Cristina Marqueze; Elaine Vieira Santos; Evandra Strazza Rodrigues; Heidge Fukumasu; Jayme Augusto de Souza-Neto; José Salvatore Leister Patané; Luiz Alcantara; Luiz Lehmann Coutinho; Maria Carolina Elias; Maurício Lacerda Nogueira; Rafael dos Santos Bezerra; Raul Machado Neto; Rejane Maria Tommasini Grotto; Ricardo Haddad; Sandra Coccuzzo Sampaio Vessoni; Simone Kashima; Svetoslav Nanev Slavov; Vincent Louis Viala |
| EPI_ISL_6128872 | INSPI-CRN DE INFLUENZA Y OTROS VIRUS RESPIRATORIOS | INSPI-CRN DE INFLUENZA Y OTROS VIRUS RESPIRATORIOS | Alfredo Bruno; Daniel Ramos; Domenica de Mora; Fernando Llerena; Jimmy Garcés; Lizbeth Patiño; Marcela Mejia; Maria Angelica Becerra; Maritza Olmedo; Michelle Páez; Ruben Armas |
| EPI_ISL_4497083 | INSPI-CRN DE INFLUENZA Y OTROS VIRUS RESPIRATORIOS | NIC-INSPI | Alfredo Bruno; Carlos Chiluisa; Domenica de Mora.; Jimmy Garcés; Johanna Laines; Lizbeth Patiño; Manuel Gonzalez; Maritza Olmedo; Michelle Páez |
| EPI_ISL_8255195 | Illinois Department of Public Health - Springfield Lab | Illinois Department of Public Health - Springfield Lab | Bryan Sim; Gordon McCall |
| EPI_ISL_2186784, EPI_ISL_4139714, EPI_ISL_4384506, EPI_ISL_8032846 | Infinity Biologix | Centers for Disease Control and Prevention Division of Viral Diseases, Pathogen Discovery | Adrian Paskey; Benjamin Rambo-Martin; Chirayu Goswami; Christian Bixby; Christopher Gulvick; Clinton Paden; Clinton R. Paden; Dakota Howard; Darlene Wagner; Dhwani Batra; Duncan MacCannell; Erisa Sula; Jason Caravas; Jonathan Schultz; Kara Moser; Kristine Lacek; Matthew Schmerer; Peter Cook; Peter W. Cook; Robin Grimwood; Russ Hager; Scott Sammons; Shatavia Morrison; Tymeckia Kendall; Victoria Caban Figueroa; Yihe Wang; Yvette Unoarumhi |
| EPI_ISL_2828667 | Instituo Adolfo Lutz - Reginal de Ribeirao Preto | Instituto Adolfo Lutz, Interdiciplinary Procedures Center, Strategic Laboratory | Caio Vinicius Dias Lopes; Claudia Regina Gonçalves; Claudio Tavares Sacchi; Erica Valessa Ramos Gomes; Karoline Rodrigues Campos |
| EPI_ISL_6779106 | Institute for Infectious Diseases | Institute for Infectious Diseases | Alban Ramette; Christian Baumann; Cora Sägesser; Franziska Suter-Riniker; Loïc Borcard; Miguel A Terrazos Miani; Nicole Liechti; Pascal Bittel; Peter Keller; Sonja Gempeler; Stefan Neuenschwander; Stephen L Leib |
| EPI_ISL_7898712 | Instituto Adolfo Lutz - Regional de Santo Andre | Instituto Adolfo Lutz, Interdiciplinary Procedures Center, Strategic Laboratory | Ariadne Ferreira Amarante; Caio Vinicius Dias Lopes; Claudia Regina Gonçalves; Claudio Tavares Sacchi; Karoline Rodrigues Campos; Leonardo Tadeu de Araujo; Marlon Benedito Nascimento Santos |
| EPI_ISL_2609537 | Instituto Nacional de Investigação em Saúde | KRISP, KZN Research Innovation and Sequencing Platform | Afonso P; David K; Emmanuel SJ; Freitas RH; Giandhari J; Inglês L; Lutucuta S; Miranda J; Morais J; Mufinda M; Naidoo Y; Neto Z; Paulo A Carralero RR Paixão JP; Pereira A; Pillay S; Tegally H; Wilkinson E; de Oliveira T |
| EPI_ISL_2970402 | Instituto Nacional de Medicina Genomica | Instituto Nacional de Medicina | Blancas S; Cedro-Tanda A; Cisneros- Villanueva M; Cisneros-Villanueva M; Escobar-Arrazola; Gonzalez-Barrera D; Herrera-Montalvo LA.; Hidalgo-Miranda A; M; Mendoza-Vargas A; Munguia-Garza P; Orjuela-Rodríguez M; Ramirez-Vega O; Rangel-DeLeon D; Reyes-Grajeda JP |

| | | | |
|---|---|---|---|
| | | Genomica | |
| EPI_ISL_2295406 | Instituto de Biotecnologia - UNESP-Botucatu-SP | Instituto de Biotecnologia - UNESP-Botucatu-SP | Cecília Artico Banho; Cíntia Bittar; Fábio Sossai Possebon; Guilherme Campos; Helena Lage Ferreira; Jorge A. Petroli Marchesi; João Pessoa Araújo Jr.; Leila Sabrina Ullmann; Lívia Sacchetto; Maisa C. Pereira Parra; Marília Moraes; Maurício L. Nogueira; Paula Rahal; Paulo Inacio da Costa |
| EPI_ISL_2636203 | Israel Central Virology laboratory | Israel National Consortium for SARS-CoV-2 sequencing | Dana Bar-Ilan; Efrat Dahan Bucris; Efrat Glick-Saar; Ella Mendelson; Gideon Rechavi; Michal Mandelboim; Miranda Geva; Neta Zuckerman; Omri Nayshool; Oran Erster; Orna Mor |
| EPI_ISL_2331479 | Johns Hopkins Hospital Department of Pathology | Johns Hopkins Hospital Department of Pathology | Adannaya Amadi; C. Paul Morris; Chun Huai Luo; Heba H. Mostafa; Matthew Schwartz; Nicholas Gallagher |
| EPI_ISL_1382279, EPI_ISL_2833494, EPI_ISL_2877058 | KU Leuven, Rega Institute, Clinical and Epidemiological Virology | KU Leuven, Rega Institute, Clinical and Epidemiological Virology | Bert Vanmechelen; Joan Marti-Carerras; Piet Maes; Tony Wawina-Bokalanga |
| EPI_ISL_3825658 | Kaiser Permanente NW Reginal Lab | OHSU MM Lab | Yun Wu |
| EPI_ISL_2802073 | Klinika za infektivne bolesti "Dr. Fran Mihaljević" | Hrvatski zavod za javno zdravstvo | Irena Tabain; Ivana Ferenčak |
| EPI_ISL_5071120 | LABORATORIO ALIFE HEALTH | Instituto Nacional de Salud- Dirección de Investigación en Salud Pública | Carlos Franco-Muñoz; Carmen Osorio; Diana Malo; Diego A. Álvarez-Díaz; Diego Andrés Prada; Gerardo Santamaría; Hector Alejandro Ruiz-Moreno; Jhonnatan Reales-González; Jorge Rivera; Juan Camilo Martinez; Julian Naizaque; Katherine Laiton-Donato; Lisseth Pardo; Magdalena Wiesner; Marcela Mercado-Reyes; Maria T. Herrera-Sepúlveda; Marta Lopez Blanco; Martha Lucia Ospina Martinez; Paola Rojas-Estevez; Sergio Gomez; Sheryll Corchuelo; Ángela Alarcon Cruz |
| EPI_ISL_3385839 | LABORATORIO CLINICO CRISTIAN GRAM IPS SAS | Instituto Nacional de Salud- Dirección de Investigación en Salud Pública | Carlos Franco-Muñoz; Carmen Osorio; Diana Malo; Diego A. Álvarez-Díaz; Diego Andrés Prada; Gerardo Santamaría; Hector Alejandro Ruiz-Moreno; Jhonnatan Reales-González; Jorge Rivera; Juan Camilo Martinez; Julian Naizaque; Katherine Laiton-Donato; Lisseth Pardo; Magdalena Wiesner; Marcela Mercado-Reyes; Maria T. Herrera-Sepúlveda; Marta Lopez Blanco; Martha Lucia Ospina Martinez; Paola Rojas; Sergio Gomez; Sheryll Corchuelo; Ángela Alarcon Cruz |
| EPI_ISL_2345266 | LABORATORIO DE FRANCA | Instituto Butantan / ESALQ-Piracicaba | Antonio Jorge Martins; Claudia Renata dos Santos Barros; David Schlesinger; Debora Botequio Moretti; Dimas Tadeu Covas; Elaine Cristina Marqueze; Elaine Vieira Santos; Evandra Strazza Rodrigues; Heidge Fukumasu; Jayme Augusto de Souza-Neto; José Salvatore Leister Patané; Luiz Alcantara; Luiz Lehmann Coutinho; Maria Carolina Elias; Maurício Lacerda Nogueira; Rafael dos Santos Bezerra; Raul Machado Neto; Rejane Maria Tommasini Grotto; Ricardo Haddad; Sandra Coccuzzo Sampaio Vessoni; Simone Kashima; Svetoslav Nanev Slavov; Vincent Louis Viala |
| EPI_ISL_5055230 | LABORATORIO IMAT | Instituto Nacional de Salud- Dirección de Investigación en Salud Pública | Beatriz de Arco; Carlos Franco-Muñoz; Carmen Osorio; Diana Malo; Diego A. Álvarez-Díaz; Diego Andrés Prada; Gerardo Santamaría; Hector Alejandro Ruiz-Moreno; Jhonnatan Reales-González; Jorge Rivera; Juan Camilo Martinez; Julian Naizaque; Katherine Laiton-Donato; Lisseth Pardo; Magdalena Wiesner; Marcela Mercado-Reyes; Maria T. Herrera-Sepúlveda; Marta Lopez Blanco; Martha Lucia Ospina Martinez; Paola Rojas; Sergio Gomez; Sheryll Corchuelo; Tatiana Cobos; Ángela Alarcon Cruz |
| EPI_ISL_1643509 | Labor Dr. Wisplinghoff - KÃ¶ln | Robert Koch Institute | |
| EPI_ISL_2390702 | Labor ZOTZ\|KLIMAS; MVZ Düsseldorf-Centrum | Robert Koch Institute | |
| EPI_ISL_2111154 | Laborarztpraxis Dres. med. Walther Weindel & Kollegen | Robert Koch Institute | |
| EPI_ISL_2374084 | Laboratoire Virologie Saint Louis APHP | Laboratoire Virologie Saint Louis APHP | Constance Delauguerre; Jérome Le Goff; Linda Feghoul; Marie Laure Chaix; Marie Laure Néré; Maud Salmona; Severine Mercier Delarue; Sophia Achaibou |
| EPI_ISL_2975479, EPI_ISL_7707101 | Laboratoire de santé publique du Québec | Laboratoire de santé publique du Québec | Guillaume Bourque; Ioannis Ragoussis; Jesse Shapiro; Mark Lathrop and Judith Fafard on behalf of the CoVSeQ research group; Mark Lathrop and Michel Roger on behalf of the CoVSeQ research group; Sandrine Moreira |
| EPI_ISL_2401014 | Laboratoire national de sante, Microbiology, Virology | Laboratoire national de sante, Microbiology, Microbial Genomics Platform | Anke Wienecke-Baldacchino; Catherine Ragimbeau; Fatu Djabi; Jessica Tapp; Lise Pignon; Raoul Salmon; Tamir Abdelrahman; Trung Nguyen Nguyen |
| EPI_ISL_3148352 | Laboratoires d'analyses medicales - Ketterthill | Laboratoire national de sante, Microbiology, Microbial Genomics Platform | Anke Wienecke-Baldacchino; Caroline Scheiber; Catherine Ragimbeau; Elodie Solarino; Fatu Djabi; Jessica Tapp; Lise Pignon; Raoul Salmon; Serge Vedy; Tamir Abdelrahman; Virginie Jover |
| EPI_ISL_4946240 | Laboratorio Angel ( Synlab) | Instituto Nacional de Salud- Dirección de Investigación en Salud Pública | Carlos Franco-Muñoz; Carmen Osorio; Diana Malo; Diego A. Álvarez-Díaz; Diego Andrés Prada; Gerardo Santamaría; Hector Alejandro Ruiz-Moreno; Jhonnatan Reales-González; Jorge Rivera; Juan Camilo Martinez; Julian Naizaque; Katherine Laiton-Donato; Lisseth Pardo; Magdalena Wiesner; Marcela Mercado-Reyes; Maria T. Herrera-Sepúlveda; Marta Lopez Blanco; Martha Lucia Ospina Martinez; Paola Rojas; Sergio Gomez; Sheryll Corchuelo; Ángela Alarcon Cruz |
| EPI_ISL_2835946 | Laboratorio Central de Epidemiología (LCE) | Centro de Investigación en Enfermedades Infecciosas (CIENI), Instituto Nacional de Enfermedades Respiratorias (INER) | ; Alejandra Garcia-Gasca; Alejandra Hernández-Terán; Alejandro Sanchez-Flores; Alfredo Herrera-Estrella; Alicia Ocaña-Mondragón; Andreu Comas-García; Angel Gustavo Salas-Lais; Antonio Loza Román; Bernardo Martínez-Miguel; Blanca Taboada; Brenda Irasema Maldonado-Meza; Bruno Gomez-Gil; Carla Ivón Herrera-Najera; Carlos F. Arias; Celia Boukadida; Clara Esperanza Santacruz-Tinoco; Concepción Grajales-Muñiz; Consorcio Mexicano de Vigilancia Genómica (CoViGen-Mex). Authors (in alphabetical order): Julio Elias Alvarado-Yaah; Cristóbal Cháidez-Quiróz; Célida Duque Molina; Célida Martínez- Rodríguez; Daniel Fregoso-Rueda; Daniel Lira Morales; Eduardo Becerril-Vargas; Fernando Fontove-Herrera; Fidencio Mejía-Nepomuceno; Francisco Pulido; Gloria Elena Espinosa-Ayala; Gloria María Molina-Salinas; Gloria Vazquez; Hector Esteban Paz-Juárez; Hector Montoya-Fuentes; Helen Haydee Fernanda Ramírez-Plascencia; Irvin González-López; Jean Pierre González; Joel Armando Vázquez-Pérez.; Jorge Salas-Hernández; José Antonio Enciso-Moreno; José Arturo Martínez-Orozco; José Esteban Muñoz-Medina; José de Jesús Nuñez-Contreras; Juan Bautista Chale-Dzul; Julissa Enciso-Ibarra; Luis Alberto Ochoa-Carrera; Margarita Matías-Florentino; Mario Mújica-Sánchez; Marissa Perez-Garcia; María Guadalupe Santiago-Mauricio; María Guadalupe de Jesús Mireles-Rivera; Nelly Sélem-Mojica; Pavel Isa; Ricardo Ciria Merce; Ricardo Grande; Rosa María Gutierrez Rios; Santiago Ávila-Ríos; Selene Zárate; Susana Lopez; Victor Eduardo Garcia-Arias; Víctor Hugo Borja-Aburto |
| EPI_ISL_3434899 | Laboratorio Central de Saude Publica do Estado do Amapa (LACEN/AP) | Laboratory of Respiratory Viruses and Measles, Oswaldo Cruz Institute, FIOCRUZ | Agatha Cristinne Prudencio; Alice Sampaio Rocha; Ana Carolina Mendonca; Andreia Santos Costa; Anna Carolina Paixao; Anne Caroline da Silva Soledade; Elisa Cavalcante Pereira; Fernando Motta; Ighor Leonardo Arantes Gomes; Lindomar dos Anjos Silva; Luciana Appolinario; Marcia Socorro Pereira Cavalcante; Marilda Siqueira on behalf of the Fiocruz COVID-19 Genomic Surveillance Network; Paola Resende; Renata Serrano Lopes; Taina Venas |
| EPI_ISL_2196197 | Laboratorio Central de Saude Publica do Estado do Rio de Janeiro (LACEN-RJ) | Laboratory of Respiratory Viruses and Measles, Oswaldo Cruz Institute, FIOCRUZ | Alice Sampaio Rocha; Ana Carolina Mendonca; Andrea Cony Cavalcanti; Anna Carolina Paixao; Elisa Cavalcante Pereira; Fernando Motta; Luciana Appolinario; Marilda Siqueira on behalf of the Fiocruz COVID-19 Genomic Surveillance Network; Paola Resende; Renata Serrano Lopes; Taina Venas |
| EPI_ISL_5604607 | Laboratorio Masvida | Instituto Nacional de Salud- Dirección de Investigación en Salud Pública | Beatriz de Arco; Carlos Franco-Muñoz; Diego A. Álvarez-Díaz; Diego Andrés Prada; Dioselina Peláez-Carvajal; Gerardo Santamaría; Hector Alejandro Ruiz-Moreno; Jhonnatan Reales-González; Jorge Rivera; Julián Naizaque; Katherine Laiton-Donato; Marcela Mercado-Reyes.; Martha Lucia Ospina Martínez; María T. Herrera-Sepúlveda; Paola Rojas-Estevez; Sheryll Corchuelo; Tatiana Cobos |
| EPI_ISL_6703122 | Laboratorio SYNLAB Colombia | CIAT, Laboratorio de Virologia | Ana M. Leiva; Diana Lopez-Alvarez; Programa Nacional de Caracterización Genómica de SARS-CoV-2; WilmerJ. Cuellar |
| EPI_ISL_7961350 | Laboratorio de Biologia Molecular - Hospital Carlos Alberto Seguín Escobedo (ESSALUD) | Laboratorio de Genómica Microbiana, Universidad Peruana Cayetano Heredia | Brenes Hebleen; Camacho Erwin; Campos-Sánchez Rebeca; Ceballos Ana; Cordero Estela; Cristancho Marco; César Iván Chávez López; Diego Cuicapuza; Duarte Francisco; Erla Patricia Asencios Vales; Fernández-Do Porto Darío; Guillermo Salvatierra; Herrera-Estrella Alfredo; Iohana Nazarita Vera Núñez; Isabel Llerena Gamero; Janet Huancachoque / Fernando Sánchez Fragoso; Jiménez-Moraila Beatriz; Kreuze Jan; Lin Geraldin Zevallos Cuarite; Mireles-Rivera Guadalupe; Molina-Mora José; Muñoz-Medina José Esteban; Negri Tatiana; Nunes Gisele; Oliveira Guilherme; Oliveira Renato; Pedro E. Romero; Reales-González Jhonnatan; Remes-Lenicov Federico; Reyes Alejandro; Rosario Elsa Jesús; Segundo Fuentes; Sosa Ezequiel; Soto Claudio; Tatiana Shessira Chávez Arias; Tsukayama Pablo; Turjanski Adrián; Zúñiga Manrique; and Zayat Jonathan / Alejandra Dávila-Barclay |
| EPI_ISL_3376696 | Laboratorio de Biología Molecular - Universidad del Magdalena | Centro de Genética y Biología Molecular - Universidad del Magdalena | Andrea M. Ramírez Hernandez; Angel Oviedo Marquez; Daniel Bautista; Lyda R. Castro; Maria Teresa Mojica-Ortiz |
| EPI_ISL_3707390 | Laboratorio de Genómica Microbiana, Universidad Peruana Cayetano Heredia | Laboratorio de Genómica Microbiana, Universidad Peruana Cayetano Heredia | Alejandra Dávila-Barclay; Diego Cuicapuza; Guillermo Salvatierra; Janet Huancachoque; Luis González; Pablo Tsukayama; Pedro E. Romero; Pool Marcos |
| EPI_ISL_3133703, EPI_ISL_3758942 | Laboratorio de Infectologia y Virologia Molecular | Laboratory of Molecular Virology, School of Medicine, Pontificia Universidad Catolica de Chile | Ana Maria Contreras; Andres E. Munoz-Marcos; Carlos Palma; Catalina Pardo-Roa; Constanza Maldonado; Constanza Martinez-Valdevenito; Eileen Serrano; Erick Salinas; Estefany Poblete; Francisco Melo; Jennifer Angulo; Jorge Levican; Leonardo I. Almonacid; M. Belen Leyton; Marcela Ferres; Maria Jose Avendano; Rafael A. Medina; Tamara Garcia-Salum |
| EPI_ISL_3537555, EPI_ISL_3537600, EPI_ISL_3547031, EPI_ISL_3670583, EPI_ISL_3769581, EPI_ISL_3987841, EPI_ISL_4036755, EPI_ISL_4080203, EPI_ISL_4080280, EPI_ISL_4417204, EPI_ISL_4417382, EPI_ISL_4417405, EPI_ISL_4635258, EPI_ISL_4740164, EPI_ISL_5115887, EPI_ISL_5504530, EPI_ISL_6574110 | | | |
| see above | Laboratorio de Referencia Nacional de Virus Respiratorios. Centro Nacional de Salud Publica. Instituto Nacional de Salud Peru. | Laboratorio de Referencia Nacional de Virus Respiratorios. Centro Nacional de Salud Publica. Instituto Nacional de Salud Peru. | Alicia Nuñez Llanos; Carlos Padilla Rojas; Edwart Rogger Rivera Serrano; Henri Bailon Calderon; Iris Silva Molina; Joseph Huayra Niquen; Kelly Vanessa Izarra Rojas; Lely Solari Zerpa; Luis Barcena Flores; Marco Galarza Perez; Nancy Rojas Serrano; Nieves Sevilla Castañeda; Omar Caceres Rey; Orson Mestanza Millones; Princesa Medrano Alhuay; Priscila Lope Pari; Sandra Morales Ruiz; Sara Gordillo Vilchez; Steve Acedo Lazo; Veronica Hurtado Vela; Victor Jimenez Vasquez; Wendy Lizarraga Olivares |
| EPI_ISL_2536793, EPI_ISL_2536881, EPI_ISL_2921449, EPI_ISL_2921499, EPI_ISL_3023521, EPI_ISL_3375975, EPI_ISL_3376040 | | | |
| see above | Laboratorio de Referencial Nacional de Virus Respiratorios | Laboratorio de Referencial Nacional de Virus Respiratorios | Carlos Padilla Rojas; Henri Bailon Calderon; Iris Silva Molina; Joseph Huayra Niquen; Lely Solari Zerpa; Luis Barcena Flores; Marco Galarza Perez; Nancy Rojas Serrano; Omar Caceres Rey; Orson Mestanza Millones; Priscila Lope Pari; Sandra Morales Ruiz; Steve Acedo Lazo; Veronica Hurtado Vela |
| EPI_ISL_1548411, EPI_ISL_2184764, EPI_ISL_3687413, EPI_ISL_4375598, EPI_ISL_4376018, EPI_ISL_4381627, EPI_ISL_6079425, EPI_ISL_7904286, EPI_ISL_8156279 | | | |
| see above | Laboratory Corporation of America | Centers for Disease Control and Prevention Division of Viral Diseases, Pathogen Discovery | Adrian Paskey; Amanda Douglas; Amanda Suchanek; Andrea Throop; Ayla Burns; Benjamin Rambo-Martin; Bobbi Croy; Brian Krueger; Brian Norvell; Christopher Gulvick; Christos Petropoulos; Clinton Paden; Clinton R. Paden; Craig Lukasik; Dakota Howard; Darlene Wagner; Debbie Boles; Dhwani Batra; Duncan MacCannell; Eyad Almasri; Goran Stevovic; Howard Engler; Hrushikesh Deshmukh; Jake Humphrey; Jana Schroth; Jason Caravas; Joe Voshell; John Pruitt; Jonathan Meltzer; Jonathan Williams; Kara Moser; Kimberly Wagner; Kristine Lacek; Lax Iyer; Lisa Pfefferle; Lyndon Tilson; Manoj Jain; Marcia Eisenberg; Mary Ann Cristobal; Mary Cristobal; Mary Williamson; Matthew Robinson; Matthew Schmerer; Michael Levandoski; Mike Sapeta; Mindy Nye; Minoo Agarwal; Mohan Kolli; Nuthawin Charoensri; Oren Cohen; Peter Cook; Peter W. Cook; Prashant Gupta; Qian Zeng; Rama Ghatti; Scott Parker; Scott Ryan; Scott Sammons; Shatavia Morrison; Stanley Letovsky; Steven Ragan; Suresh Babu Selvaraju; Suresh Selvaraju; Susan Countryman; Susan Hicks; Suzanne Dale; Thomas Urban; Tim Kuphal; Tricia Zwiefelhofer; Tymeckia Kendall; Victoria Caban Figueroa; Vincent Drouillon; Yvette Unoarumhi |
| EPI_ISL_3050545, EPI_ISL_4516039 | Laboratório Central de Saúde Pública do Amazonas - LACEN-AM | Laboratorio de Ecologia de Doencas Transmissiveis na Amazonia, Instituto Leonidas e Maria Deane - Fiocruz Amazonia | André Corado; Felipe Naveca; Fernanda Nascimento; George Silva; Karina Pessoa; Luciana Gonçalves; Maria Júlia Brandão; Matilde Mejía; Valdinete Nascimento; Victor Souza; Ágatha Costa |
| EPI_ISL_4543295 | Laboratório de Biotecnologia Aplicada (LBA) - Laboratório de Biologia Molecular - Hospital das Clínicas, Faculdade de Medicina de Botucatu, Departamento de Bioprocessos e Biotecnologia - Faculdade de Ciências Agronômicas, UNESP – Botucatu/SP | Laboratory of Respiratory Viruses and Measles, Oswaldo Cruz Institute, FIOCRUZ | Alice Sampaio Rocha; Ana Carolina Mendonca; Anna Carolina Paixao; Elisa Cavalcante Pereira; Felipe Allan da Silva Costa; Fernando Motta; Jayme Augusto de Souza Neto; Leonardo Nazario de Moraes; Luciana Appolinario; Marilda Siqueira on behalf of the Fiocruz COVID-19 Genomic Surveillance Network; Paola Resende; Patricia Akemi Assato; Rejane Maria Tommasini; Renata Serrano Lopes; Taina Venas |
| EPI_ISL_5400838 | Landesamt für Verbraucherschutz des Landes Sachsen-Anhalt | Robert Koch Institute | |
| EPI_ISL_796926, EPI_ISL_1330351, EPI_ISL_1414295, EPI_ISL_1700095, EPI_ISL_3368302, EPI_ISL_4010236, EPI_ISL_5134344, EPI_ISL_5974027, EPI_ISL_7893732, EPI_ISL_7992921, EPI_ISL_8048904, EPI_ISL_8237543 | | | |
| see above | Lighthouse Lab in Alderley Park | Wellcome Sanger Institute for the COVID-19 Genomics UK (COG-UK) Consortium | Cordelia Langford; David K. Jackson; Dominic Kwiatkowski; Ewan Harrison; Ian Johnston; Jacquelyn Wynn; Jeffrey Barrett; John Sillitoe on behalf of the Wellcome Sanger Institute COVID-19 Surveillance Team; Mairead Hyland; Roberto Amato; Sonia Goncalves; The Lighthouse Lab in Alderley Park and Alex Alderton |
| EPI_ISL_7305684, | Lighthouse Lab in Glasgow | Wellcome Sanger Institute for the | Anna Dominiczak and Alex Alderton; Carol Clugston; Cordelia Langford; David Gray; David K. Jackson; Dominic Kwiatkowski; Ewan Harrison; Harper VanSteenhouse; Ian Johnston; Jeffrey Barrett; John Sillitoe on behalf of the Wellcome Sanger Institute COVID-19 Surveillance Team; Roberto Amato; Sonia |

| | | | |
|---|---|---|---|
| EPI_ISL_7630312, EPI_ISL_7867167 | | COVID-19 Genomics UK (COG-UK) Consortium | Goncalves; Yumi Kasai |
| EPI_ISL_873910, EPI_ISL_875020, EPI_ISL_2540099, EPI_ISL_3884142, EPI_ISL_5786901, EPI_ISL_6225882, EPI_ISL_7557476, EPI_ISL_7579656, EPI_ISL_7582935, EPI_ISL_7655401, EPI_ISL_7718518, EPI_ISL_7838025, EPI_ISL_7867480, EPI_ISL_7964979, EPI_ISL_7968329, EPI_ISL_7974757, EPI_ISL_8040219, EPI_ISL_8041334, EPI_ISL_8041571, EPI_ISL_8101978, EPI_ISL_8102137, EPI_ISL_8167447, EPI_ISL_8168656, EPI_ISL_8241909 | | | |
| see above | Lighthouse Lab in Milton Keynes | Wellcome Sanger Institute for the COVID-19 Genomics UK (COG-UK) Consortium | Cordelia Langford; David K. Jackson; Dominic Kwiatkowski; Ewan Harrison; Ian Johnston; Jeffrey Barrett; John Sillitoe on behalf of the Wellcome Sanger Institute COVID-19 Surveillance Team; Roberto Amato; Sonia Goncalves; The Lighthouse Lab in Milton Keynes and Alex Alderton |
| EPI_ISL_7862511, EPI_ISL_7955308, EPI_ISL_8235063 | Lighthouse Laboratory Plymouth | Wellcome Sanger Institute for the COVID-19 Genomics UK (COG-UK) Consortium | Cordelia Langford; David K. Jackson; Dominic Kwiatkowski; Ewan Harrison; Ian Johnston; Jeffrey Barrett; John Sillitoe on behalf of the Wellcome Sanger Institute COVID-19 Surveillance Team; Lighthouse Laboratory Plymouth and Alex Alderton; Roberto Amato; Sonia Goncalves |
| EPI_ISL_3152346 | MD PHL | Maryland Department of Health Laboratories Administration | Ami Patel; Eric N. Keller; Jillian Loomis; Kwang Low; Terence L. Moore; and Robert Myers |
| EPI_ISL_5659977 | MDI Limbach Berlin GmbH; MVZ Labor Berlin | Robert Koch Institute | |
| EPI_ISL_2689877 | MS Public Health Laboratory | Centers for Disease Control and Prevention Division of Viral Diseases, Pathogen Discovery | Alison Laufer Halpin; Ben L. Rambo-Martin; Clinton R. Paden; Dakota Howard; Darlene Wagner; Dave Wentworth; Dhwani Batra; Jasmine Padilla; Justin Lee; Katie Dillon; Krista Queen; Kristen Knipe; Kristine Lacek; Mark Burroughs; Matthew Schmerer; Mili Sheth; Peter Cook; Sam Shepard; Sarah Nobles; Shoshona Le; Suxiang Tong; Vivien Dugan; Yvette Unoarumhi |
| EPI_ISL_3427956, EPI_ISL_8126798, EPI_ISL_8262424 | Mako Medical | Centers for Disease Control and Prevention Division of Viral Diseases, Pathogen Discovery | Adrian Paskey; Benjamin Rambo-Martin; Christopher Gulvick; Clinton Paden; Clinton R. Paden; Dakota Howard; Darlene Wagner; Dhwani Batra; Duncan MacCannell; Erisa Sula; Jason Caravas; Kara Moser; Kristine Lacek; Lauren Moon; Matthew Schmerer; Matthew Tugwell; Peter Cook; Peter W. Cook; Scott Sammons; Shatavia Morrison; Tymeckia Kendall; Victoria Caban Figueroa; Yvette Unoarumhi |
| EPI_ISL_6963085 | Malawi Liverpool Wellcome Trust Clinical Research Program | Malawi Liverpool Wellcome Trust Clinical Research Program | Ben Morton; Catherine Anscombe; Kondwani Jambo; Philip Ashton; Sam Lissauer |
| EPI_ISL_1367749, EPI_ISL_7495079 | Michigan Department of Health and Human Services, Bureau of Laboratories | Michigan Department of Health and Human Services, Bureau of Laboratories | Blankenship HM; Riner D; Soehnlen MK |
| EPI_ISL_2140547 | Microbiology Department, Laboratori Clínic Metropolitana Nord. Hospital Universitari Germans Trias i Pujol | Can Ruti SARS-CoV-2 Sequencing Hub (HUGTiP/IrsiCaixa/IGTP) | Alba Sánchez; Anna Not; Antoni E Bordoy; Bonaventura Clotet; Cristina Casañ; Cristina Esteban; Francesc Catala-Moll; Gemma Clara; Ignacio Blanco; Marc Noguera-Julian; Maria Casadellà; Mariona Parera; Mercedes Guerrero; Montserrat Giménez; Pere-Joan Cardona; Pilar Armengol; Roger Paredes; Verónica Saludes; and Elisa Martró on behalf of the Can Ruti SARS-CoV-2 Sequencing Hub. |
| EPI_ISL_5331932 | Microbiology Department, University Hospital Donostia | Microbiology Department, University Hospital Donostia | Cilla G.; Gomez M; Marimon JM; Martin-Peñaranda T; Montes M; Piñeiro L; Sorarrain A |
| EPI_ISL_3470233 | Microbiology Department. Complexo Hospitalario Universitario de Vigo | Microbiology Department. Complexo Hospitalario Universitario de Vigo | Alfaya N; Alonso I; Alvarez M; Cabrera JJ; Carballo R; Cores O; Cortizo S; Davina C; Martinez L; Mediero G; Perez S; Potel C; Regueiro B; Rey S; Vasallo FJ; del-Campo V |
| EPI_ISL_935074, EPI_ISL_1911806, EPI_ISL_3589012, EPI_ISL_7874529 | Ministry of Health Turkey | Ministry of Health Turkey | Ayşe Başak Altaş; Fatma Bayrakdar; Gulay Korukluoglu; Gülay Korukluoğlu; Suleyman Yalcin; Süleyman Yalcin; Yasemin Cosgun; Yasemin Coşgun |
| EPI_ISL_2843164 | Ministry of Public Health / Hamad Medical Corporation | Biomedical Research Center (BRC), Qatar University / Qatar Genome Project (QGP) | Asmaa A. Al-Thani. MOPH and HMC: Abdullatif Al-Khal; BRC: Fatiha M. Benslimane; Chadi Saad; Dalia Al-Batesh; Dina Elgakhlab QGP: Fatima H. Al-Kuwari; Einas A. E. Al-Kuwari; Hadi M. Yassine; Hamad E. Al-Romaihi; Hamda Alromaihi; Heba A. Al-Khatib; Mashael A. Al-Bader; Mohammed Al-Thani; Muna A. S. Al-Maslamani; Oal Al-Jamal; Peter V. Coyle; Reham A. El-Kahlout. QBB: Tasneem Al-Hamad; Roberto Bertollini; Salih Al-Marri |
| EPI_ISL_8208476 | Montana Public Health Laboratory | Montana Public Health Laboratory | Ashley Ausman Deborah Gibson; Joy Goffena; Joy Ritter; Michelle Mozer |
| EPI_ISL_3704337 | NUPIT/UFPE | WallauLab on behalf of Fiocruz COVID-19 Genomic Surveillance Network | Alexandre Freitas da Silva; Cassia Docena; Constância Flávia Junqueira Ayres; Filipe Zimmer Dezordi; Gabriel Luz Wallau; Gustavo Barbosa de Lima; Lais Ceschini Machado; Lilian Caroliny Amorim Silva; Maira Galdino da Rocha Pitta; Marcelo Henrique dos Santos Paiva; Matheus Filgueira Bezerra; Michelly Cristiny Pereira; Rômulo Pessoa e Silva; Sinval Pinto Brandão Filho |
| EPI_ISL_2180060 | National Center of Infectious and Parasitic Diseases | National Center of Infectious and Parasitic Diseases | Alexiev et al |
| EPI_ISL_402125 | National Institute for Communicable Disease Control and Prevention (ICDC) Chinese Center for Disease Control and Prevention (China CDC) | National Institute for Communicable Disease Control and Prevention (ICDC) Chinese Center for Disease Control and Prevention (China CDC) | Chen; Dai; F.-H.; Hu, Y.; J.-H.; J.-J.; J.-L. and Zhu; Liu, Y.; Pei; Q.-M.; She; Song; T.-Y.; Tao; Tian; Wang; Wang, W.; Wu, F.; Xu, L.; Y.-L.; Y.-M.; Y.-Y.; Y.-Z.; Yu, B.; Z.-G.; Z.-W.; Zhang; Zhao, S.; Zheng |
| EPI_ISL_1827589 | National Institute for Communicable Diseases,National Health Laboratory Services, Gauteng, South Africa | National Institute for Communicable Diseases of the National Health Laboratory Service | Amoako DG; Bhiman JN; Ismail A; Mahlangu B; Mohale T; Ntuli N; Scheepers C |
| EPI_ISL_1665234 | National Platform bis UMONS/Jolimont | National Platform bis UMONS/Jolimont | François Dufrasne; Gautier Detry; Guillaume Bayon-Vicente; Ruddy Wattiez |
| EPI_ISL_1317637, EPI_ISL_1317646 | Nordland Hospital - Bodo, Laboratory Department, Molecular Biology Unit | Norwegian Institute of Public Health, Department of Virology | Atiya R Ali; Debech Nadia; Engebretsen Serina Beate; Garcia Llorente Ignacio; Hilde Elshaug; Hilde Vollan; Jon Bråte; Kamilla Heddeland Instefjord; Karoline Bragstad; Kathrine Stene-Johansen; Marie Paulsen Madsen; Olav Hungnes; Pedersen Benedikte Nevjen; Rasmus Riis Kopperud |
| EPI_ISL_2517869 | Northumbria University / South Tees Hospitals NHS Foundation Trust / North Cumbria Integrated Care NHS Foundation Trust / North Tees and Hartlepool NHS Foundation Trust / Newcastle Hospitals NHS Foundation Trust | COVID-19 Genomics UK (COG-UK) Consortium | Andrew Nelson; Brendan Payne; Clive Graham; Darren L Smith; Debra Padgett; Edward Barton; Emma Swindells; Garren Scott; Gary Black; Gary Eltringham; Giles S Holt; Greg R Young; Jane Greenaway; Jennifer Collins; John Allan; Joshua Loh; Lynn Dover; Matthew Bashton; Mohammad A Tariq; Paul Baker; Sarah Essex; Steve Liggett; Wen C Yew; Yusri Taha |
| EPI_ISL_7976245 | Northwestern Memorial Hospital | Northwestern University - Center for Pathogen Genomics and Microbial Evolution | Chad J. Achenbach; Chao Qi; Egon A. Ozer; Judd F. Hultquist; Lacy M. Simons; Lawrence J. Jennings; Michael G. Ison; Ramon Lorenzo-Redondo; Taylor J. Dean |
| EPI_ISL_2928905 | Oregon State Public Health Laboratory | Oregon State Public Health Laboratory | Eugene Yeboah; John Fontana and Shane Sevey; Laura Tsaknaridis; Rafia Razzaque; Vanda Makris |
| EPI_ISL_6294609, EPI_ISL_7350045, EPI_ISL_7465200 | Originating lab: Wales Specialist Virology Centre Sequencing lab: Pathogen Genomics Unit | Public Health Wales Microbiology Cardiff Wales Specialist Virology Centre | Alec Birchley; Alexander Adams; Amy Gaskin; Angela Marchbank; Bree Gatica-Wilcox; Catherine Moore; Jason Coombes; Joanne Watkins; Joel Southgate; Johnathan Evans; Laura Gifford; Lauren Gilbert; Lee Graham; Malorie Perry; Matthew Bull; Nicole Pacchiarini; Sally Corden; Sara Kumziene-Summerhayes; Sara Rey; Sarah Taylor; Simon Cottrell; Sophie Jones; Tom Connor |
| EPI_ISL_6002335 | Orlickoustecka nemocnice | University Hospital Hradec Kralove | Helena Kovarikova; Lenka Rysava; Marketa Gancarcikova; Monika Berankova |
| EPI_ISL_4119464 | Oslo University Hospital, Department of Microbiology | Norwegian Institute of Public Health, Department of Virology | Arvind Yegambaram Meenakshi Sundaram; Cathrine Fladeby; Garcia Llorente Ignacio; Gregor D. Gilfillan; Hilde Elshaug; Hilde Vollan; Jon Bråte; Kamilla Heddeland Instefjord; Karoline Bragstad; Kathrine Stene-Johansen; Line Victoria Moen; Lise Andresen; Mariann Nilsen; Mona Holberg-Petersen; Olav Hungnes; Pedersen Benedikte Nevjen; Pål Marius Bjørnstad; Rasmus Riis Kopperud; Teodora Plamenova Ribarska |
| EPI_ISL_3078829 | Oxford Viromics, NDM, University of Oxford; Oxford University Hospitals; Basingstoke and North Hampshire Hospital | COVID-19 Genomics UK (COG-UK) Consortium | Alex Mobbs; Amy Trebes; Anita Justice; Catrin Moore; Christophe Fraser; David Bonsall; David Buck; Emma Wise; George Macintyre; Jessica Lynch; John Todd; Mariateresa de Cesare; Matilde Mori; Monique Andersson; Nathan Moore; Nick Cortes; Robert Shaw; Stephen Kidd; Tanya Golubchik; Timothy Peto |
| EPI_ISL_5646133 | PSF DR ANTONIO PIRES DE ALMEIDA PORTO FELIZ | Instituto Butantan | Antonio Jorge Martins; Claudia Renata dos Santos Barros; David Schlesinger; Debora Botequio Moretti; Dimas Tadeu Covas; Elaine Cristina Marqueze; Elaine Vieira Santos; Evandra Strazza Rodrigues; Heidge Fukumasu; Jayme Augusto de Souza-Neto; José Salvatore Leister Patané; Luiz Alcantara; Luiz Lehmann Coutinho; Maria Carolina Elias; Maurício Lacerda Nogueira; Rafael dos Santos Bezerra; Raul Machado Neto; Rejane Maria Tommasini Grotto; Ricardo Haddad; Sandra Coccuzzo Sampaio Vessoni; Simone Kashima; Svetoslav Nanev Slavov; Vincent Louis Viala |
| EPI_ISL_3933315, EPI_ISL_4096617 | Pandemic Response Lab - NYC | Pandemic Response Lab, R&D | Cybill del Castillo; Dylan Law; Haiping Hao; Henry Lee; Isabel Fernandez Escapa; Jon Laurent; Katharine Nelson; Melissa Hopkins; Michael Hammerling; Pradeep Bugga; Shinyoung Clair Kang; Sol Rey; William Ward |
| EPI_ISL_3197396 | Pathogen Genomics Center, National Institute of Infectious Diseases | Pathogen Genomics Center, National Institute of Infectious Diseases | Kentaro Itokawa; Makoto Kuroda; Masanori Hashino; Rina Tanaka; Tsuyoshi Sekizuka |
| EPI_ISL_5306503 | Pathology West - NSW Health Pathology | NSW Health Pathology - Institute of Clinical Pathology and Medical Research; Westmead Hospital; University of Sydney | Arnott A.; Draper J.; Gall M.; Martinez E.; Rockett R.; Sintchenko V.; on behalf of ICPMR |
| EPI_ISL_2188122 | Philippine Red Cross Logistics and Multipurpose Center | Philippine Genome Center | Alethea R. de Guzman; Anna Ong-Lim; Arianne A. Zamora; Asia Louisa U. Chong; Benedict A. Maralit; Candice Francheska B. Tambaoan; Carlo M. Lapid; Celia Carlos; Devon Ray Pacial; Edsel Maurice Salvaña; El King D. Morado; Elcid Aaron R. Pangilinan; Eva Maria Cutiongco-de la Paz; Francis A. Tablizo; Irish Coleen A. Asin; Jaime C. Montoya; Jan Michael C. Yap; Jo-Hannah S. Llames; John Q. Wong; Joshua Gregor A. Dizon; Juan Antonio R. Magalang; Karol Sophia Agape R. Padilla; Kenneth M. Kim; Kris P. Punayan; Marc Edsel C. Ayes; Marc Jerrone R. Castro; Maria Rosario Singh-Vergeire and Cynthia P. Saloma; Maria Sofia L. Yangzon; Marissa Alejandria; Razel Nikka M. Hao; Rianna Patricia S. Cruz; Sheila Mae M. Araiza |
| EPI_ISL_5569914 | Philippine Red Cross- Batangas Chapter Molecular Laboratory | Philippine Genome Center | Alethea R. de Guzman; Alyssa Joyce E. Telles; Anna Ong-Lim; Arianne A. Zamora; Benedict A. Maralit; Carlo M. Lapid; Celia Carlos; Cynthia P. Saloma; Devon Ray Pacial; Diomedes A. Cariño; Edsel Maurice Salvana; El King D. Morado; Elcid Aaron R. Pangilinan; Eva Maria Cutiongco-de la Paz; Francis A. Tablizo; Henrietta Marie Rodriguez; Jaime C. Montoya; Jan Michael C. Yap; Jarvin E. Nipales; Jo-Hannah S. Llames; John Michael Egana; John Q. Wong; Joshua Gregor A. Dizon; Joshua Jose Endozo; Juan Antonio R. Magalang; Karol Sophia Agape R. Padilla; Kris P. Punayan; Kristina Patriz Dela Cruz; Lindsay Clare D.L. Carandang; Ma. Exanil Plantig; Marc Edsel C. Ayes; Maria Rosario Singh-Vergeire; Maria Sofia L. Yangzon; Marielle M. Gamboa; Marissa Alejandria; Niña Francesca Bustamante; Razel Nikka M. Hao; Renato Jacinto Q. Mantaring; Rianna Patricia S. Cruz; Shiela Mae M. Araiza; Yvonne Valerie Austria; Zipporah Mariebelle R. Enriquez; Zyrel V. Mollejon |
| EPI_ISL_3391074 | Plateau technique Biomer Metz Verriers | Department of Virology, Henri Mondor University Hospital, Assistance Publique Hôpitaux de Paris, Université | Alexandre Soulier; Christophe Rodriguez; Elisabeth Trawinski; Guillaume Gricourt; Jean-Michel Pawlotsky; Melissa N'Debi; Slim Fourati; Vanessa Demontant |

| | | | |
|---|---|---|---|
| | | Paris-Est Créteil, INSERM U955 | |
| EPI_ISL_1524087 | Public Health Authority of the Slovak Republic | Laboratory of Genomics and Bioinformatics, Comenius University Science Park | Anna Gičová; Diana Rusňáková; Jaroslav Budiš; Miroslav Böhmer; Tatiana Sedláčková; Tomáš Szemes |
| EPI_ISL_2698835 | Public Health Laboratory, Minnesota Department of Health | University of Minnesota Genomics Center | Corbin Dirkx; Daryl M. Gohl; Jaquelyn Kuriger-Laber; John Garbe; and Sean Wang |
| EPI_ISL_4369218, EPI_ISL_4372037, EPI_ISL_4372955, EPI_ISL_4373365 | Quest Diagnostics Incorporated | Centers for Disease Control and Prevention Division of Viral Diseases, Pathogen Discovery | A. Gerasimova; A. Perez; B. Anderson; Benjamin Rambo-Martin; Christopher Gulvick; Clinton Paden; Dakota Howard; Dhwani Batra; Duncan MacCannell; Erisa Sula; F. Lacbawan; I. Shlyakhter; Jason Caravas; K. Livingston; Kristine Lacek; L. Bernstein; M. Hua; Matthew Schmerer; P. Tanpaiboon; Peter Cook; R. Kagan; R. Owen; R. Rolando; S. Rosenthal; Scott Sammons; Shatavia Morrison; Tymeckia Kendall; Victoria Caban Figueroa; Y. Liu; Yvette Unoarumhi |
| EPI_ISL_2893914 | REUNILAB | CNR Virus des Infections Respiratoires - France SUD | Antonin Bal; Bruno Lina; Gregory Destras; Gwendolyne Burfin; Hadrien Regue; Laurence Josset; Martine Valette; Quentin Semanas |
| EPI_ISL_1628780 | REUNILAB | UMR PIMIT | Dr Camille Lebarbenchon; Dr David A Wilkinson; Dr Patrick Mavingui; Magali Turpin |
| EPI_ISL_1409540 | Randox Laboratories | Wellcome Sanger Institute for the COVID-19 Genomics UK (COG-UK) Consortium | Cordelia Langford; David K. Jackson; Dominic Kwiatkowski; Ewan Harrison; Ian Johnston; Jeffrey Barrett; John Sillitoe on behalf of the Wellcome Sanger Institute COVID-19 Surveillance Team; Randox Laboratories and Alex Alderton; Roberto Amato; Sonia Goncalves |
| EPI_ISL_2473932 | Reditus Laboratories | Reditus Laboratories | Alexa Eichelberger; Cassy Philips; Joshua J. Geltz; M.S.; Ph.D.; Rex Dyer; Robert M. Sgambelluri |
| EPI_ISL_8039057 | Regionalspital Emmental AG Burgdorf, Labor | Institute for Infectious Diseases, University of Bern | Alban Ramette; Christian Baumann; Cora Sägesser; Franziska Suter-Riniker; Loïc Borcard; Miguel A Terrazos Miani; Nicole Liechti; Pascal Bittel; Peter Keller; Sonja Gempeler; Stefan Neuenschwander; Stephen L Leib |
| EPI_ISL_2860081, EPI_ISL_4715142 | Research Institute for Tropical Medicine, Inc. (RITM) | Philippine Genome Center | Alethea R. de Guzman; Anna Ong-Lim; Arianne A. Zamora; Benedict A. Maralit; Carlo M. Lapid; Celia Carlos; Cynthia P. Saloma; Devon Ray Pacial; Diomedes A. Carino; Diomedes A. Cariño; Edsel Maurice Salvana; Edsel Maurice Salvaña; El King D. Morado; Elcid Aaron R. Pangilinan; Eva Maria Cutiongco-de la Paz; Francis A. Tablizo; Henrietta Marie Rodriguez; Jaime C. Montoya; Jan Michael C. Yap; Jarvin E. Nipales; Jo-Hannah S. Llames; John Q. Wong; Joshua Gregor A. Dizon; Juan Antonio R. Magalang; Karol Sophia Agape R. Padilla; Kenneth M. Kim; Kris P. Punayan; Krisitna Patriz Dela Cruz; Kristina Patriz Dela Cruz; Lindsay Claire D.L. Carandang; Lindsay Clare D.L. Carandang; Ma. Exanil Plantig; Marc Edsel C. Ayes; Maria Rosario Singh-Vergeire; Maria Rosario Singh-Vergeire and Cynthia P. Saloma; Maria Sofia L. Yangzon; Marielle M Gamboa; Marielle M. Gamboa; Marissa Alejandria; Nina Francesca Bustamante; Niña Francesca Bustamante; Razel Nikka M. Hao; Renato Jacinto Q. Mantaring; Rianna Patricia S. Cruz; Sheila Mae M. Araiza; Shiela Mae M. Araiza; Yvonne Valerie Austria; Zipporah Mariebelle R. Enriquez; Zyrel V. Mollejon |
| EPI_ISL_3080448, EPI_ISL_4309658 | Respiratory Virus Unit, Microbiology Services Colindale, Public Health England | COVID-19 Genomics UK (COG-UK) Consortium | PHE Covid Sequencing Team |
| EPI_ISL_7840805 | Rosalind Franklin Laboratory | Wellcome Sanger Institute for the COVID-19 Genomics UK (COG-UK) Consortium | Cordelia Langford; David K. Jackson; Dominic Kwiatkowski; Donald Fraser; Ewan Harrison; Ian Johnston; Jeffrey Barrett; John Sillitoe on behalf of the Wellcome Sanger Institute COVID-19 Surveillance Team; Rob Howes; Roberto Amato; Sonia Goncalves; Suki Lee; The Rosalind Franklin Laboratory and Alex Alderton |
| EPI_ISL_2765174, EPI_ISL_3195724, EPI_ISL_8097004 | SARS-CoV-2 testing team, National Institute of Infectious Diseases | Pathogen Genomics Center, National Institute of Infectious Diseases | Hazuka Y Furihata; Hiromizu Takahashi; Kentaro Itokawa; Makoto Kuroda; Masanori Hashino; Masumichi Saito; Naomi Nojiri; Nozomu Hanaoka; Rina Tanaka; Tsuguto Fujimoto; Tsuyoshi Sekizuka |
| EPI_ISL_3385800 | SECRETARIA DE SALUD DE SAN ANDRES ISLA | Instituto Nacional de Salud- Dirección de Investigación en Salud Pública | Carlos Franco-Muñoz; Carmen Osorio; Diana Malo; Diego A. Álvarez-Díaz; Diego Andrés Prada; Gerardo Santamaría; Hector Alejandro Ruiz-Moreno; Jhonnatan Reales-González; Jorge Rivera; Juan Camilo Martinez; Julian Naizaque; Katherine Laiton-Donato; Lisseth Pardo; Magdalena Wiesner; Marcela Mercado-Reyes; Maria T. Herrera-Sepúlveda; Marta Lopez Blanco; Martha Lucia Ospina Martinez; Paola Rojas; Sergio Gomez; Sheryll Corchuelo; Ángela Alarcon Cruz |
| EPI_ISL_3255757 | SK-Roy Romanow Provincial Laboratory | National Microbiology Laboratory (NML) | Alanna Senecal; Amanda Lang; Anna Majer; Anneliese Landgraff; CanCOGeN's metadata curation team; Darian Hole; Elsie Grudeski; Gary Van Domselaar; Grace Seo; Jennifer Tanner; Jessica Minion; Kara Loos; Keith MacKenzie; Kirsten Biggar; Madison Chapel; Meredith Faires; Morag Graham; Natalie Knox; Nathalie Bastien; Philip Mabon; Public Health Agency of Canada CanCOGeN team; Rachel DePaulo; Rhiannon Huzarewich; Russell Mandes; Ryan McDonald; Shari Tyson; Timothy Booth; Yan Li |
| EPI_ISL_1595690 | SYNLAB | GIGA Medical Genomics | Bouchra Boujemla; Cécile Meex; Keith Durkin; Maria Artesi; Marie-Pierre Hayette; Nathalie Renotte; Pierrette Melin; Raphaël Boreux; Sébastien Bontems; Vincent Bours |
| EPI_ISL_5202214 | SYNLAB | LSPSDS | Alejandro Gomez Lopez; Gabriela Delgado Murcia; Johana Hernandez Toloza; Marcela Castano Rodriguez; Marcela Mercado |
| EPI_ISL_4230406 | SYNLAB MVZ Hamburg | Robert Koch Institute | |
| EPI_ISL_1849542, EPI_ISL_2130997 | SYNLAB MVZ Heidelberg | Robert Koch Institute | |
| EPI_ISL_6467409 | SYNLAB MVZ Leinfelden-Echterdingen | Robert Koch Institute | |
| EPI_ISL_2111707 | SYNLAB MVZ Leverkusen | Robert Koch Institute | |
| EPI_ISL_6437701 | Salud Digna | Instituto Nacional de Medicina Genomica | Abraham Campos-Romero; Cedro-Tanda A; Cruz-Islas Jazmin; Escobar-Arrazola; Gonzalez-Barrera D; Herrera-Montalvo LA.; Hidalgo-Miranda A; Luna-Ruiz Marco; M; Mendoza-Vargas A; Moreno-Camacho José Luis; Munguia-Garza P; Ramirez-Vega O; Rangel-DeLeon D; Reyes-Grajeda JP; Rodriguez-Gallegos Jorge |
| EPI_ISL_5986683 | Scripps Medical Laboratory | Andersen lab at Scripps Research | Ellen Stefanski; Ian Mchardy; SEARCH Alliance San Diego with Michael Quigley |
| EPI_ISL_8185050 | Sema4 | Sema4 | Angela Yannes; Bisram Deocharan; Feras Hantash; Matthew Capozziello; Rui Gu; Victoria Parkington; Zdenek Markovic |
| EPI_ISL_7751354, EPI_ISL_7752987, EPI_ISL_7753061 | Servicio Virosis Respiratorias-Departamento Virologia-INEI | Instituto Nacional Enfermedades Infecciosas C.G.Malbran | Avaro M.; Baumeister E.; Benedetti E.; Campos J.; Cisterna D.; Dattero ME; De Belder D.; Haim MS.; Lorenzo F.; Molina V.; Perandones C.; Poklepovich T.; Pontoriero A.; Russo M.; Sanchez Loria J.; Tuduri E. |
| EPI_ISL_4083393, EPI_ISL_4083412, EPI_ISL_5159078, EPI_ISL_6694676, EPI_ISL_6694755 | Servicio Virosis Respiratorias-Departamento Virología-INEI | Instituto Nacional Enfermedades Infecciosas C.G.Malbran | Avaro M.; Baumeister E.; Benedetti E.; Campos J.; Cisterna D.; Dattero ME; De Belder D.; Haim MS.; Lorenzo F.; Molina V.; Perandones C.; Poklepovich T.; Pontoriero A.; Russo M.; Sanchez Loria J.; Tuduri E. |
| EPI_ISL_2934733 | Servicio de Microbiología Clínica (Complejo Hospitalario de Navarra, Pamplona) | Centro de Secuenciación NASERTIC | Ana Miqueleiz; Ana Navascués; Carmen Ezpeleta Baquedano |
| EPI_ISL_8136296 | Shamir Medical Center (Asaf Harofe) | Shamir Medical Center (Asaf Harofe) | Abu Hamad Ramzia; Adina Bar Chaim; Anna Vishnevsky; Chen Weiner; Nir Rainy; Patricia Benveniste-Lekovitz; Reut Sorek Abramovich; Yevgeni Yegorov |
| EPI_ISL_1420584, EPI_ISL_1602954, EPI_ISL_1606093, EPI_ISL_1617543, EPI_ISL_2211128, EPI_ISL_2256570, EPI_ISL_2418725 | | | |
| see above | Swedish national genomic surveillance program of SARS-CoV-2 | The Public Health Agency of Sweden | Alma Brolund; Maria Lind Karlberg; Maximilian Riess; Swedish national genomic surveillance program of SARS-CoV-2 |
| EPI_ISL_7952710 | Synlab Medilab | Karolinska University Hospital Huddinge | Annika Tiveljung Lindell; Henning Onsbring; Jan Albert; Karina Hentrich; Lynda Eneh; Martin Ekman; Natalija Gerasimcik; Robert Dyrdak; Sandra Broddesson; Shambhu Ganeshappa Aralaguppe; Tanja Normark; Tobias Allander; Valtteri Wirta; Zhibing Yun |
| EPI_ISL_6327824 | TSHEPONG LABORATORY | National Institute for Communicable Diseases of the National Health Laboratory Service | Amoako DG; Bhiman JN; Everatt J; Ismail A; Mahlangu B; Mnguni A; Mohale T; Ntuli N; Scheepers C |
| EPI_ISL_2678157, EPI_ISL_6250639 | The Caribbean Public Health Agency | Carrington Lab, Department of Preclinical Sciences, Faculty of Medical Sciences, The University of the West Indies, St Augustine Campus | Anushka Ramjag; Arianne Brown-Jordan; Avery Hinds; Christine V. F. Carrington; Christopher Oura; Gabriel Escobar; Nikita S. D. Sahadeo; Nuno Faria; Oliver Pybus; Risha Singh; Roshan Parasram; Sarah Hill; Simone Keizer-Beache; SueMin Nathaniel; Vernie Ramkissoon |
| EPI_ISL_1581433 | UAB InMedica | Vilnius University Hospital Santaros Klinikos, Center of Laboratory Medicine | Daniel Naumovas; Dovile Ezerskyte; Gytis Dudas; Ingrida Olendraite; Laimonas Griskevicius; Ligita Raugaite; Mindaugas Stoskus; Monika Katenaite; Rimvydas Norvilas |
| EPI_ISL_1966460 | UBS IV GUARARAPES | Instituto Butantan / Mendelics | Antonio Jorge Martins; Bianca Cechetto Carlos. Mendelics: Bibiana Santos; Claudia Renata dos Santos Barros; Cíntia Bittar; David Schlesinger. Hemocentro Ribeirão Preto: Simone Kashima; Debora Botequio Moretti; Elaine Cristina Marqueze; Elaine Vieira dos Santos; Elisangela Chicaroni Mattos; Erika Freitas; Evandra Strazza Rodrigues; Felipe Allan da Silva da Costa; Flavia Aburjaile; Fábio Sossai Possebon; Guilherme Campos; Guilherme Targino Valente; Heidge Fukumasu. USP-Botucatu: Rejane Maria Tommasini Grotto; Helena Lage Ferreira; Instituto Butantan: Dimas Tadeu Covas; Jardelina de Souza Todao Bernardino; Jayme A. Souza-Neto; Jessika Cristina Chagas Lesbon; Jorge A. Petroli Marchesi; José Salvatore Leister Patané; João Paulo Kitajima; João Pessoa Araújo Jr.; Leila Sabrina Ullmann; Loyze Paola Oliveira de Lima; Luiz Aurelio de Campos Crispin. Centro de Genômica Funcional da ESALQ: Luiz Lehmann Coutinho; Luiz Carlos Junior de Alcantara; Lívia Sacchetto; Maisa C. Pereira Parra; Maria Carolina Elias; Marta Giovanetti; Marília Moraes; Maurício Lacerda Nogueira. Prefeitura de Sao Paulo: Melissa Palmieri.; Patricia Akemi Assato; Paula Rahal; Paulo Inacio da Costa; Rafael dos Santos Bezerra; Raquel de Lello Rocha Campos Cassano. NGS Soluções Genômicas: Pilar Drummond Sampaio Corrêa Mariani. FZEA-USP Pirassununga: Mirele Daiana Poleti; Raul Machado Neto; Ricardo Augusto Brassaloti; Ricardo Haddad; Rodrigo Tocantins Calado. FAMERP-SJRP: Cecília Artico Banho; Sandra Coccuzzo Sampaio; Svetoslav Nanev Slavov; Vagner Fonseca; Vincent Louis Viala |
| EPI_ISL_5646922 | UNIDADE DE VIGILANCIA EM SAUDE | Instituto Butantan | Antonio Jorge Martins; Claudia Renata dos Santos Barros; David Schlesinger; Debora Botequio Moretti; Dimas Tadeu Covas; Elaine Cristina Marqueze; Elaine Vieira Santos; Evandra Strazza Rodrigues; Heidge Fukumasu; Jayme Augusto de Souza-Neto; José Salvatore Leister Patané; Luiz Alcantara; Luiz Lehmann Coutinho; Maria Carolina Elias; Maurício Lacerda Nogueira; Rafael dos Santos Bezerra; Raul Machado Neto; Rejane Maria Tommasini Grotto; Ricardo Haddad; Sandra Coccuzzo Sampaio Vessoni; Simone Kashima; Svetoslav Nanev Slavov; Vincent Louis Viala |
| EPI_ISL_5647737 | UNIDADE RESPIRATORIA NOVA HORTOLANDIA | Instituto Butantan | Antonio Jorge Martins; Claudia Renata dos Santos Barros; David Schlesinger; Debora Botequio Moretti; Dimas Tadeu Covas; Elaine Cristina Marqueze; Elaine Vieira Santos; Evandra Strazza Rodrigues; Heidge Fukumasu; Jayme Augusto de Souza-Neto; José Salvatore Leister Patané; Luiz Alcantara; Luiz Lehmann Coutinho; Maria Carolina Elias; Maurício Lacerda Nogueira; Rafael dos Santos Bezerra; Raul Machado Neto; Rejane Maria Tommasini Grotto; Ricardo Haddad; Sandra Coccuzzo Sampaio Vessoni; Simone Kashima; Svetoslav Nanev Slavov; Vincent Louis Viala |
| EPI_ISL_2208904 | UPA DR FRANCO DA ROCHA | Instituto Butantan / Mendelics | Antonio Jorge Martins; Claudia Renata dos Santos Barros; David Schlesinger; Debora Botequio Moretti; Dimas Tadeu Covas; Elaine Cristina Marqueze; Elaine Vieira Santos; Evandra Strazza Rodrigues; Heidge Fukumasu; Jayme Augusto de Souza-Neto; José Salvatore Leister Patané; Luiz Alcantara; Luiz Lehmann Coutinho; Maria Carolina Elias; Maurício Lacerda Nogueira; Rafael dos Santos Bezerra; Raul Machado Neto; Rejane Maria Tommasini Grotto; Ricardo Haddad; Sandra Coccuzzo Sampaio Vessoni; Simone Kashima; Svetoslav Nanev Slavov; Vincent Louis Viala |
| EPI_ISL_2693307 | UW Virology Lab | UW Virology Lab | Alexander Greninger; Hong Xie; Keith R Jerome; Lasata Shrestha; Meei-Li Huang; Michelle Lin; Nathan Breit; Noah R. Baker; Patrick Mathias; Pavitra Roychoudhury; Ricardo Perez; Robert J. Livingston; Sean Ellis; Shah Mohamed Bakhash; Tien V. Nguyen |
| EPI_ISL_3568784 | Unidad de Investigacion Medica de Yucatan (UIMY) | Unidad de Genomica Avanzada | ; Alejandra Garcia-Gasca; Alejandra Hernandez-Teran; Alejandro Sanchez-Flores; Alfredo Herrera-Estrella; Alicia OcaÃ±a-Mondragon; Andreu Comas-Garcia; Angel Gustavo Salas-Lais; Antonio Loza Roman; Bernardo Martinez-Miguel; Blanca Taboada; Brenda Irasema Maldonado-Meza; Bruno Gomez-Gil; Carla Ivon Herrera-Najera; Carlos F. Arias; Celia Boukadida; Celida Duque Molina; Celida Martinez- Rodriguez; Clara Esperanza Santacruz-Tinoco; Concepcion Grajales-MuÃ±iz; Consorcio Mexicano de Vigilancia Genomica (CoViGen-Mex). Authors (in alphabetical order): Julio Elias Alvarado-Yaah; Cristobal Chaidez-Quiroz; Daniel Fregoso-Rueda; Daniel Lira Morales; Eduardo Becerril-Vargas; Fernando Fontove-Herrera; Fidencio Mejia-Nepomuceno; Francisco Pulido; Gloria Elena Espinosa-Ayala; Gloria Maria Molina-Salinas; Gloria Vazquez; Hector Esteban Paz-Juarez; Hector Montoya-Fuentes; Helen Haydee Fernanda Ramirez-Plascencia; Irvin Gonzalez-Lopez; Jean Pierre Gonzalez; Jesus Hernandez; Joel Armando Vazquez-Perez.; Jorge Salas-Hernandez; Jose Antonio Enciso-Moreno; Jose Arturo Martinez-Orozco; Jose Esteban MuÃ±oz-Medina; Jose de Jesus NuÃ±ez-Contreras; Juan Bautista Chale-Dzul; Julissa Enciso-Ibarra; Luis Alberto Ochoa-Carrera; Margarita Matias-Florentino; Maria Guadalupe Santiago-Mauricio; Maria Guadalupe de Jesus Mireles-Rivera; Mario Mujica-Sanchez; Marissa Perez-Garcia; Nelly Selem-Mojica; Pavel Isa; Ricardo Ciria Merce; Ricardo Grande; Rosa Maria Gutierrez Rios; Santiago avila-Rios; Selene Zarate; Susana Lopez; Veronica Mata-Haro; Victor Eduardo Garcia-Arias; Victor Hugo Borja-Aburto |
| EPI_ISL_7780468 | Unidad de Investigación Médica de Yucatán (UIMY) | Centro de Investigación en Enfermedades Infecciosas (CIENI), | Alejandra García-Gasca; Alejandra Hernández-Terán; Alejandro Sánchez-Flores; Alfredo Herrera-Estrella; Alicia Ocaña-Mondragón; Andreu Comas-García; Angel Gustavo Salas-Lais; Antonio Loza Román; Bernardo Martínez-Miguel; Blanca Taboada; Brenda Irasema Maldonado-Meza; Bruno Gómez-Gil; Carla Ivón Herrera-Najera; Carlos F. Arias; Celia Boukadida; Clara Esperanza Santacruz-Tinoco; Concepción Grajales-Muñiz; Consorcio Mexicano de Vigilancia Genómica (CoViGen-Mex). Authors (in alphabetical order): Julio Elias Alvarado-Yaah; Cristóbal Cháidez-Quiróz; Célida Duque Molina; Célida |

| | | | |
|---|---|---|---|
| | | Instituto Nacional de Enfermedades Respiratorias (INER) | Martínez-Rodríguez; Daniel Fregoso-Rueda; Daniel Lira Morales; Eduardo Becerril-Vargas; Eduardo Rivera-Martínez; Fernando Fontove-Herrera; Fidencio Mejía-Nepomuceno; Francisco Pulido; Gabriel Chavira-Trujillo; Gloria Elena Espinosa-Ayala; Gloria María Molina-Salinas; Gloria Vazquez; Hector Montoya-Fuentes; Helen Haydee Fernanda Ramírez-Plascencia; Irvin González-López; Jean Pierre González; Jesús Hernández; Joel Armando Vázquez-Pérez.; Jorge Salas-Hernández; José Antonio Enciso-Moreno; José Arturo Martínez-Orozco; José Esteban Muñoz-Medina; José de Jesús Nuñez-Contreras; Juan Bautista Chale-Dzul; Julissa Enciso-Ibarra; Kathia Elizabeth Tapia-Diaz; Luis Alberto Ochoa-Carrera; Margarita Matías-Florentino; Mario Mújica-Sánchez; Marissa Perez-Garcia; María Eugenia Jiménez-Corona; María Guadalupe Santiago-Mauricio; María Guadalupe de Jesús Mireles-Rivera; Nelly Sélem-Mojica; Pavel Isa; Ricardo Ciria Merce; Ricardo Grande; Rosa María Gutiérrez Rios; Rosario Vazquez-Larios; Santiago Ávila-Ríos; Selene Zárate; Susana Lopez; Verónica Mata-Haro; Victor Eduardo García-Arias; Víctor Hugo Borja-Aburto |
| EPI_ISL_2898445 | Unilabs | Karolinska University Hospital | Annelie Bjerkner; Isak Sylvin; Jan Albert; Karolina Ininbergs; Lina Guerra Blomqvist; Lynda Eneh; Martin Ekman; Martina Wahlund; Robert Dyrdak; Sandra Broddesson; Tanja Normark; Tobias Allander; Valtteri Wirta; Zhibing Yun |
| EPI_ISL_3477572 | Universidad del Valle, LDAB-Laboratorio de Diagnostico de Agentes Biologicos | Universidad del Valle, TAO-Lab, VIREM, NEAS Network | Andres Castillo; Beatriz Parra & COVID-19 Team Univalle; Diana López-Alvarez; Nelson Rivera Franco |
| EPI_ISL_983044 | University Health Network/Mount Sinai Hospital Department of Microbiology | Ontario Institute for Cancer Research | Aimee Paterson; Allison McGeer; Angel Liu; Bernard Lam; Cassandra Bergwerff; Ilinca Lungu; Jared T. Simpson; Javier Hernandez; Jeff Wrana; Jessica Bourke; Kin Chan; Kuganya Nirmalarajah; Laurence Pelletier; Lawrence E. Heisler; Lubaina Kothari; Marc Mazzulli; Marie-Ming Aynaud; Michael Laszloffy; Patryk Aftanas; Paul Krzyzanowski; Richard de Borja; Samira Mubareka; Seda Barutcu; Tony Mazzulli |
| EPI_ISL_1241680 | University of Liège COVID-19 testing center | GIGA Medical Genomics | Bouchra Boujemla; Cécile Meex; Keith Durkin; Maria Artesi; Marie-Pierre Hayette; Nathalie Renotte; Pierrette Melin; Raphaël Boreux; Sébastien Bontems; Vincent Bours |
| EPI_ISL_4742149 | VITALAB Molecular Laboratory | Philippine Genome Center | Alethea R. de Guzman; Anna Ong-Lim; Arianne A. Zamora; Benedict A. Maralit; Carlo M. Lapid; Celia Carlos; Cynthia P. Saloma; Devon Ray Pacial; Diomedes A. Cariño; Edsel Maurice Salvana; El King D. Morado; Elcid Aaron R. Pangilinan; Eva Maria Cutiongco-de la Paz; Francis A. Tablizo; Henrietta Marie Rodriguez; Jaime C. Montoya; Jan Michael C. Yap; Jarvin E. Nipales; Jo-Hannah S. Llames; John Q. Wong; Joshua Gregor A. Dizon; Juan Antonio R. Magalang; Karol Sophia Agape R. Padilla; Kenneth M. Kim; Kris P. Punayan; Kristina Patriz Dela Cruz; Lindsay Clare D.L. Carandang; Ma. Exanil Plantig; Marc Edsel C. Ayes; Maria Rosario Singh-Vergeire; Maria Sofia L. Yangzon; Marielle M. Gamboa; Marissa Alejandria; Niña Francesca Bustamante; Razel Nikka M. Hao; Renato Jacinto Q. Mantaring; Rianna Patricia S. Cruz; Shiela Mae M. Araiza; Yvonne Valerie Austria; Zipporah Mariebelle R. Enriquez; Zyrel V. Mollejon |
| EPI_ISL_1371734 | Virginia Division of Consolidated Laboratory Services | Virginia Division of Consolidated Laboratory Services | Virginia DCLS |
| EPI_ISL_6563125 | WSSE w Gdansku | Wojewodzka Stacja Sanitarno-Epidemiologiczna w Olsztynie, Laboratorium Badan Epidemiologiczno-Klinicznych | Aleksandra Kobiatko; Barbara Dolinska; Emilia Tarabasz; Ewa Liszewska; Marta Lukian; Monika Czerminska; Patryk Bielecki; Paulina Rozycka; Sylwia Krzetowska; Tomasz Jakubczak |
| EPI_ISL_8209445 | Washington State Department of Health Public Health Laboratories | Washington State Department of Health Public Health Laboratories | Alex Lathum; Ardizon Valdez; Avi Singh; Claire Howell; Denny Russell; Drew MacKellar; Holly Halstead; JohnAric Peterson; Kathryn Sickles; Kristin Roche; Lisa Jones; Philip Dykema; Rebecca Cao |
| EPI_ISL_2534262 | Wichita State University - Molecular Diagnostics Lab | Kansas Health and Environmental Lab | Ben Olsen; Jonathan Barnell; Katherine Wiggins; Mike Grose; and Phil Adam |
| EPI_ISL_3556540 | Wisconsin State Laboratory of Hygiene Communicable Disease Division | Wisconsin State Laboratory of Hygiene Communicable Disease Division | Abigail C. Shockey; Alicia J. Mooney; Erika M. Hanson; Kelsey R. Florek; Richard Griesser; Sara Wagner; Tonya Danz |
| EPI_ISL_2034966 | Yale Clinical Virology Lab | Yale Center for Genomic Analysis | Brooke Sullivan; Curt Scharfe; Irina Tikhonova; Kaya Bilguvar; Shrikant Mane |
| EPI_ISL_2727263 | ZARV/NHLS, Department Medical Virology, University of Pretoria | KRISP, KZN Research Innovation and Sequencing Platform | Adriano Mendes; Amy Strydom; Emmanuel SJ; Giandhari J; Lessells R; Micheala Davids; Naidoo Y; Pillay S; Ramphal U; Sim Mayaphi and Marietjie Venter; Tegally H; Wilkinson E; de Oliveira T |
| EPI_ISL_6666338 | Zaklad Diagnostyki Laboratoryjnej Koagulologii i Mikrobiologii | Wojewodzka Stacja Sanitarno-Epidemiologiczna w Rzeszowie, Laboratorium Diagnostyki Medycznej | Anna Nowakowska; Karolina Ostrowska; Katarzyna Wilk; Marzena Baranowska |
| EPI_ISL_2360232 | Zentrallabor Zürich | Institute of Medical Virology | Alexandra Trkola; Annette Audigé; Cyril Shah; Gabriela Ziltener; Guido Bloemberg; Jon Huder; Jürg Böni; Kevin Steiner; Maria Grünberg; Maryam Zaheri; Michael Huber; Riccarda Capaul; Stefan Schmutz; Verena Kufner |
| EPI_ISL_4295792 | unknown | CNR Virus des Infections Respiratoires - France SUD | Antonin Bal; Bruno Lina; Gregory Destras; Gwendolyne Burfin; Hadrien Regue; Laurence Josset; Martine Valette; Quentin Semanas |

We gratefully acknowledge the following Authors from the Originating laboratories responsible for obtaining the specimens, as well as the Submitting laboratories where the genome data were generated and shared via GISAID, on which this research is based.

All Submitters of data may be contacted directly via www.gisaid.org

Authors are sorted alphabetically.

Acknowledgement EPI_SET Identifier: EPI_SET_20220328ge

| Accession ID | Originating Laboratory | Submitting Laboratory | Authors |
|---|---|---|---|
| EPI_ISL_3347206 | ADILAB | Laboratorio Departamental de Salud Publica de Antioquia | Andres F. Cardona-Rios; Gloria Isabel Escobar; Idabely Betancur Ortiz; Juan P. Hernandez-Ortiz; Maria Stella López |
| EPI_ISL_3276536, EPI_ISL_3276539, EPI_ISL_3276543 | ADILAB | Universidad Nacional de Colombia - Laboratorio Genómico One Health | Andres F. Cardona-Rios; Carlos Franco-Muñoz; Carolina Muñoz-Arango; Celeny Ortiz; Daniel O. Maldonado-Perez; Diego A. Álvarez-Díaz; Hector Alejandro Ruiz-Moreno; Idabely Betancur Ortiz; Jorge E. Osorio; Juan P. Hernandez-Ortiz; Karl A Ciuoderis; Katherine Laiton-Donato; Laura Silvana Perez; Lina M. Hurtado; Marcela Mercado-Reyes; Maria Angélica Maya; Maria Stella López; Rita Almanza Payares; Sandra Ines Cano; Simón Villegas Velásquez |
| EPI_ISL_3298351, EPI_ISL_3298358 | AREA DE SALUD ACOSTA | Inciensa, Instituto Costarricense de Investigación y Enseñanza en Nutrición y Salud | Adriana Godínez; Claudio Soto-Garita; Estela Cordero; Francisco Duarte; Hebleen Porras; José Luis Vargas; Mariela Gutiérrez & Joselyn Prado; Melany Calderón |
| EPI_ISL_3274361, EPI_ISL_3274362 | AREA DE SALUD ACOSTA [ACOSTA/SAN JOSE] | Inciensa, Instituto Costarricense de Investigación y Enseñanza en Nutrición y Salud | & Nazareth Ruiz; Adriana Godínez; Caterina Guzmán; Claudio Soto-Garita; Estela Cordero; Francisco Duarte; Hebleen Porras; José Luis Vargas; Mariela Gutiérrez Joselyn Prado; Melany Calderón |
| EPI_ISL_3274356 | AREA DE SALUD ATENAS | Inciensa, Instituto Costarricense de Investigación y Enseñanza en Nutrición y Salud | Adriana Godínez; Caterina Guzmán; Claudio Soto-Garita; Estela Cordero; Francisco Duarte; Hebleen Porras; José Luis Vargas; Mariela Gutiérrez Joselyn Prado; Melany Calderón; Nazareth Ruiz & Benito Vega |
| EPI_ISL_3298360 | AREA DE SALUD CORREDORES | Inciensa, Instituto Costarricense de Investigación y Enseñanza en Nutrición y Salud | Adriana Godínez; Claudio Soto-Garita; Estela Cordero; Francisco Duarte; Hebleen Porras; Joselyn Prado & Susana Mata; José Luis Vargas; Mariela Gutiérrez; Melany Calderón |
| EPI_ISL_3298355 | AREA DE SALUD COTO BRUS | Inciensa, Instituto Costarricense de Investigación y Enseñanza en Nutrición y Salud | Adriana Godínez; Claudio Soto-Garita; Estela Cordero; Francisco Duarte; Hebleen Porras; José Luis Vargas; Mariela Gutiérrez & Joselyn Prado; Melany Calderón |
| EPI_ISL_3274353, EPI_ISL_3274354, EPI_ISL_3274360 | AREA DE SALUD DESAMPARADOS 1 - CLINICA DR. MARCIAL FALLAS | Inciensa, Instituto Costarricense de Investigación y Enseñanza en Nutrición y Salud | & Nazareth Ruiz; Adriana Godínez; Caterina Guzmán; Claudio Soto-Garita; Estela Cordero; Francisco Duarte; Hebleen Porras; José Luis Vargas; Mariela Gutiérrez Joselyn Prado; Melany Calderón; Nazareth Ruiz & Juliana Mora Cortes |
| EPI_ISL_3274364 | AREA DE SALUD DESAMPARADOS 1 - CLINICA DR. MARCIAL FALLAS [GRIFO ALTO/DESAMPARADOS/SAN JOSE] | Inciensa, Instituto Costarricense de Investigación y Enseñanza en Nutrición y Salud | & Nazareth Ruiz; Adriana Godínez; Caterina Guzmán; Claudio Soto-Garita; Estela Cordero; Francisco Duarte; Hebleen Porras; José Luis Vargas; Mariela Gutiérrez Joselyn Prado; Melany Calderón |
| EPI_ISL_3298371 | AREA DE SALUD FORTUNA | Inciensa, Instituto Costarricense de Investigación y Enseñanza en Nutrición y Salud | Adriana Godínez; Claudio Soto-Garita; Estela Cordero; Francisco Duarte; Hebleen Porras; Joselyn Prado & Carolina Arrieta; José Luis Vargas; Mariela Gutiérrez; Melany Calderón |
| EPI_ISL_3638888 | AREA DE SALUD GOICOECHEA 2 - CLINICA DR. JIMENEZ NUÑEZ | Inciensa, Instituto Costarricense de Investigación y Enseñanza en Nutrición y Salud | Adriana Godínez; Claudio Soto-Garita; Estela Cordero; Francisco Duarte; Hebleen Porras; Joselyn Prado & Juan Carlos Cartes; José Luis Vargas; Mariela Gutiérrez; Melany Calderón |
| EPI_ISL_3274365, EPI_ISL_3274367 | AREA DE SALUD MATA REDONDA-HOSPITAL - CLINICA DR. MORENO CAÑAS [SAN JOSE/SAN JOSE] | Inciensa, Instituto Costarricense de Investigación y Enseñanza en Nutrición y Salud | & Nazareth Ruiz; Adriana Godínez; Caterina Guzmán; Claudio Soto-Garita; Estela Cordero; Francisco Duarte; Hebleen Porras; José Luis Vargas; Mariela Gutiérrez Joselyn Prado; Melany Calderón; Nazareth Ruiz & Ricardo González |
| EPI_ISL_3464535, EPI_ISL_3638898 | AREA DE SALUD OREAMUNO-PACAYAS-TIERRA BLANCA | Inciensa, Instituto Costarricense de Investigación y Enseñanza en Nutrición y Salud | Adriana Godínez; Claudio Soto-Garita; Estela Cordero; Francisco Duarte; Hebleen Porras; Joselyn Prado & Carolina Loría; Joselyn Prado & Carolina Loría Acosta; José Luis Vargas; Mariela Gutiérrez; Melany Calderón |
| EPI_ISL_3638807 | AREA DE SALUD PURISCAL-TURRUBARES | Inciensa, Instituto Costarricense de Investigación y Enseñanza en Nutrición y Salud | Adriana Godínez; Claudio Soto-Garita; Estela Cordero; Francisco Duarte; Hebleen Porras; José Luis Vargas; Mariela Gutiérrez & Joselyn Prado; Melany Calderón |
| EPI_ISL_3298349 | AREA DE SALUD SIQUIRRES | Inciensa, Instituto Costarricense de Investigación y Enseñanza en Nutrición y Salud | Adriana Godínez; Claudio Soto-Garita; Estela Cordero; Francisco Duarte; Hebleen Porras; Joselyn Prado & Ileana Chaves; José Luis Vargas; Mariela Gutiérrez; Melany Calderón |
| EPI_ISL_3639064 | AREA DE SALUD TIBAS-URUCA-MERCED - CLINICA DR. CLORITO PICADO | Inciensa, Instituto Costarricense de Investigación y Enseñanza en Nutrición y Salud | Adriana Godínez; Claudio Soto-Garita; Estela Cordero; Francisco Duarte; Hebleen Porras; Joselyn Prado & Johnny Villalobos; José Luis Vargas; Mariela Gutiérrez; Melany Calderón |
| EPI_ISL_3119719 | AULSS 3 Venezia | Istituto Zooprofilattico Sperimentale delle Venezie | Adelaide Milani; Alessia Schivo; Alice Fusaro; Ambra Pastori; Annalisa Salviato; Antonia Ricci; Calogero Terregino; Edoardo Giussani; Elisa Palumbo; Erika Giorgia Quaranta; Isabella Monne; Luca Tassoni |
| EPI_ISL_3127787, EPI_ISL_3446799 | AULSS 3 Venezia | UOSD Genetica e Citogenetica - Azienda ULSS 3 Serenissima; Istituto Zooprofilattico Sperimentale delle Venezie | Adelaide Milani; Alessia Schivo; Alice Fusaro; Ambra Pastori; Annalisa Salviato; Antonia Ricci; Calogero Terregino; Claudia Perini; Edoardo Giussani; Elisa Palumbo; Elisa Squarcina; Erika Giorgia Quaranta; Isabella Monne; Laura Bevilacqua; Laura Squarzon; Luca Sorino; Luca Tassoni; Mosé Favarato |
| EPI_ISL_3028236, EPI_ISL_3039672 | AULSS 6 Euganea | Istituto Zooprofilattico Sperimentale delle Venezie | Adelaide Milani; Alessia Schivo; Alice Fusaro; Ambra Pastori; Annalisa Salviato; Antonia Ricci; Calogero Terregino; Edoardo Giussani; Elisa Palumbo; Erika Giorgia Quaranta; Isabella Monne; Luca Tassoni |
| EPI_ISL_3430113, EPI_ISL_3430351, EPI_ISL_3430547 | Aegis Sciences Corporation | Centers for Disease Control and Prevention Division of Viral Diseases, Pathogen Discovery | Adrian Paskey; Alec Vest; Benjamin Rambo-Martin; Christopher Gulvick; Clinton R. Paden; Cyndi Clark; Dakota Howard; Darlene Wagner; Dhwani Batra; Dillon Nall; Duncan MacCannell; Ethan Sanders; Holly Houdeshell; Jason Caravas; Kara Moser; Matthew Hardison; Matthew Schmerer; Ola Kvalvaag; Patrick Campbell; Peter W. Cook; Rob Case; Scott Sammons; Shatavia Morrison; Shaun Westlund; Vikramsinha Ghorpade; Yvette Unoarumhi |
| EPI_ISL_3270474, EPI_ISL_3270475 | Alaska State Virology Laboratory | Alaska State Virology Laboratory | Elva House; Jack Chen; Jacob Zidek; Lisa Smith; Ph.D.; Stephanie DeRonde |
| EPI_ISL_3386397 | Arizona State University | Arizona State University | Ajeet Bains; Efrem S. Lim; Joshua LaBaer; LaRinda A. Holland; Matthew F. Smith; Nathaniel Johnson; Nicholas J. Mellor; Peter T. Skidmore; Rabia Maqsood; Vel Murugan |
| EPI_ISL_3276575 | Ayudas Diagnosticas Sura | Universidad Nacional de Colombia - Laboratorio Genómico One Health | Andres F. Cardona-Rios; Carlos Franco-Muñoz; Carolina Muñoz-Arango; Celeny Ortiz; Daniel O. Maldonado-Perez; Diego A. Álvarez-Díaz; Hector Alejandro Ruiz-Moreno; Idabely Betancur Ortiz; Jorge E. Osorio; Juan P. Hernandez-Ortiz; Karl A Ciuoderis; Katherine Laiton-Donato; Laura Silvana Perez; Lina M. Hurtado; Marcela Mercado-Reyes; Maria Angélica Maya; Maria Stella López; Rita Almanza Payares; Sandra Ines Cano; Simón Villegas Velásquez |
| EPI_ISL_3167149 | Basurto University Hospital: Clinical Microbiology Laboratory | Biocruces Bizkaia | Ana de la Hoz; Estibaliz Ugalde Zarraga; José Luis Díaz de Tuesta del Arco; Mikel Gallego Rodrigo; Mikel Urrutikoetxea-Gutiérrez; Mª Carmen Nieto Toboso |
| EPI_ISL_3042076, EPI_ISL_3043350, EPI_ISL_3109639, EPI_ISL_3169783, EPI_ISL_3225218 | Berkshire and Surrey Pathology Services Lighthouse Laboratory | Wellcome Sanger Institute for the COVID-19 Genomics UK (COG-UK) Consortium | Berkshire and Surrey Pathology Services Lighthouse Laboratory and Alex Alderton; Cordelia Langford; David K. Jackson; Dominic Kwiatkowski; Ewan Harrison; Ian Johnston; Jeffrey Barrett; John Sillitoe on behalf of the Wellcome Sanger Institute COVID-19 Surveillance Team; Roberto Amato; Sonia Goncalves |
| EPI_ISL_3145700 | BioneXt Lab | Laboratoire national de sante, Microbiology, Microbial Genomics Platform | Anke Wienecke-Baldacchino; Catherine Ragimbeau; Elodie Solarino; Fatu Djabi; Jessica Tapp; Lise Pignon; Raoul Salmon; Tamir Abdelrahman; Thibault Ferrandon; Virginie Jover |
| EPI_ISL_3602820 | British Columbia Centre For Disease Control | BCCDC Public Health Laboratory | Ana Pacagnella; Corrinne Ng; Dan Fornika; John Tyson; Kim Macdonald; Kimia Kamelian; Linda Hoang; Loretta Janz; Mel Krajden; Prystajecky Natalie; Robert Azana; Shannon Russell |
| EPI_ISL_3407886, EPI_ISL_3525509 | Broad Institute Clinical Research Sequencing Platform | Infectious Disease Program, Broad Institute of Harvard and MIT | Adams, G.; B.L.; B.W.; Bauer, M.; Birren; Blumenstiel, B.; Brown, C.; Carter, A.; Chaluvadi, S.; D.J.; DeFelice, M.; DeRuff, K.; Dodge, S.; Gabriel, S.; Gallagher, G.; Gladden-Young, A.; Granger, B.; J.E.; K.J.; Lagerborg, K.; Larkin, K.; Lee, M.; Lemieux; Lennon, N.; Loreth, C.; Madoff, L.; McGovern, S.; Meldrim, J.; Normandin, E.; P.C.; Park; Pearlman, L.; Reilly, S.; Rudy, M.; Sabeti; Siddle; Smole, S.; Tomkins-Tinch, C.; Vicente, G.; and MacInnis |
| EPI_ISL_3464547 | CENTRO PENAL LUIS PAULINO MORA | Inciensa, Instituto Costarricense de Investigación y Enseñanza en Nutrición y Salud | Adriana Godínez; Claudio Soto-Garita; Estela Cordero; Francisco Duarte; Hebleen Porras; Joselyn Prado & Juan Carlos Villalobos Ugalde; José Luis Vargas; Mariela Gutiérrez; Melany Calderón |
| EPI_ISL_3061861, EPI_ISL_3061862, | CH VALENCIENNES | CHU Lille - Laboratoire de Virologie | AIT YAHYA Emilie; ALIDJINOU Enagnon Kazali; BOCKET Laurence; CREPIN Michel; DEMAY Christophe; ENGELMANN Ilka; GEFFROY Sandrine; GUIGON Aurélie; LAMBERT Valérie; LAZREK Mouna; NOBILLIAUX Florian; PREVOST Brigitte; TCHANTCHOU NJOSSE YANICK; THUILLIER Caroline; TINEZ Claire |

| | | | |
|---|---|---|---|
| EPI_ISL_3061863, EPI_ISL_3061864 | | | |
| EPI_ISL_3432133 | CHTMAD | Instituto Nacional de Saude (INSA) | Borges et al |
| EPI_ISL_3383406 | CHU Purpan - Laboratoire de Virologie - Institut Fédératif de Biologie | CHU Purpan - Laboratoire de Virologie - Institut Fédératif de Biologie | Boyer P.; Carcenac R.; Ferrer V.; Harter A.; Izopet J.; Jeanne N.; Latour J.; Ranger N.; Tremeaux R. |
| EPI_ISL_3267818 | CHUV | Laboratory of genomics and metagenomics | Claire Bertelli; Damien Jacot; Gilbert Greub; Sébastien Aeby; Trestan Pillonel |
| EPI_ISL_3385855 | COLCAN | Instituto Nacional de Salud-Dirección de Investigación en Salud Pública | Carlos Franco-Muñoz; Carmen Osorio; Diana Malo; Diego A. Álvarez-Díaz; Diego Andrés Prada; Gerardo Santamaría; Hector Alejandro Ruiz-Moreno; Jhonnatan Reales-González; Jorge Rivera; Juan Camilo Martinez; Julian Naizaque; Katherine Laiton-Donato; Lisseth Pardo; Magdalena Wiesner; Marcela Mercado-Reyes; Maria T. Herrera-Sepúlveda; Marta Lopez Blanco; Martha Lucia Ospina Martinez; Paola Rojas; Sergio Gomez; Sheryll Corchuelo; Ángela Alarcon Cruz |
| EPI_ISL_3614327 | COVID LAB | Cayman Islands Forensic Science Lab | Brittany Balcewich; Jonathan Smellie; Karla Montes; Tanisha Gilbert |
| EPI_ISL_3347367 | CT Department of Public Health | Grubaugh Lab - Yale School of Public Health | Anderson Brito; Annie Watkins; Chaney Kalinich; Chantal Vogels; Claire Pearson; Isabel Ott; Jessica Rothman; Joseph Fauver; Kendall Billig; Mallery Breban; Mary Petrone; Nathan Grubaugh; Tara Alpert; Tobias Koch; Tu N. Nguyen |
| EPI_ISL_3355062 | CT-Dr. Katherine A. Kelley State Public Health Lab | Centers for Disease Control and Prevention Division of Viral Diseases, Pathogen Discovery | Alex Burgin; Ben Rambo-Martin; Clinton Paden; Dakota Howard; Dave Wentworth; Dhwani Batra; Jasmine Padilla; Justin Lee; Krista Queen; Kristen Knipe; Kristine Lacek; Mark Burroughs; Matthew Schmerer; Meghan Bentz; Mili Sheth; Peter Cook; Sam Shepard; Sarah Nobles; Suxiang Tong; Vivien Dugan; Yvette Unoarumhi |
| EPI_ISL_3242482, EPI_ISL_3242489, EPI_ISL_3242732 | Clinica INDISA | "Facultad de Ciencias de la Vida, UNAB" | "Claudio Meneses; Ariel Orellana"; Claudio Olmos; Daniel Leon; Dayan Sanhueza; Eduardo Castro; Gonzalo Campaña; Macarena Bastias; Paola Pidal; Ricardo Yusta; Sebastian Wolter; Susana Saez; Víctor Monreal; Waldo Diaz |
| EPI_ISL_3635412 | DYOMEDEA-LABORATOIRE DE LA SAUVEGARDE | CNR Virus des Infections Respiratoires - France SUD | Antonin Bal; Bruno Lina; Gregory Destras; Gwendolyne Burfin; Hadrien Regue; Laurence Josset; Martine Valette; Quentin Semanas |
| EPI_ISL_3041379, EPI_ISL_3340126 | Department of Bacteria, Parasites and Fungi, Statens Serum Institut, Copenhagen, Denmark | Statens Serum Institut Bioinformatics and Microbial Genomics | Danish Covid-19 Genome Consortium |
| EPI_ISL_3266210 | Department of Public Health Sibiu | National Institute of Infectious Diseases-Prof. Dr. Matei Bals Molecular Diagnostics Laboratory | Corina Casangiu; Dan Otelea; Leontina Banica; Marius Surleac; Ovidiu Vlaicu; Petre Milu; Robert Hohan; Simona Paraschiv |
| EPI_ISL_3152109, EPI_ISL_3298382, EPI_ISL_3298383 | Dipartimento di Medicina di Laboratorio, Azienda sanitaria universitaria Friuli Centrale (ASU FC) | Dipartimento di Medicina di Laboratorio, Azienda sanitaria universitaria Friuli Centrale (ASU FC) | Catia Mio; Chiara Dal Secco; Corrado Pipan; Francesco Curcio; Stefania Marzinotto |
| EPI_ISL_3052919 | Dirección regional de salud del Callao (DIRESA-CALLAO) | Centro de Investigaciones Tecnológicas, Biomédicas y Medioambientales (CITBM) | B; Huaman; J. Alarcon; M. Cuellar; M. Ramirez; M. Sovero |
| EPI_ISL_3024839 | Division of Emerging Infectious Diseases, Bureau of Infectious Diseases Diagnosis Control, Korea Disease Control and Prevention Agency | Division of Emerging Infectious Diseases, Bureau of Infectious Diseases Diagnosis Control, Korea Disease Control and Prevention Agency | Ae Kyung Park; Chae Young Lee; Eun-Jin Kim; Heui Man Kim; Il-Hwan Kim; Jeong-Ah Kim; Jeong-Min Kim |
| EPI_ISL_3389223, EPI_ISL_3578180, EPI_ISL_3578239, EPI_ISL_3578242 | Dr. Risch Ostschweiz AG | Microbiology, Dr. Risch | Dominique Fabien Hilti; Faina Wehrli; Lorenz Risch; Martin Risch; Nadia Wohlwend; Sinem Kas; Thomas Bodmer |
| EPI_ISL_3148575 | Dutch COVID-19 response team | Erasmus Medical Center | Anne van der Linden; Annemiek van der Eijk; Bas Oude Munnink; Corine GeurtsvanKessel; David Nieuwenhuijse; Emmanuelle Munger; Irina Chestakova; Marion Koopmans; Marjan Boter; Reina Sikkema; Richard Molenkamp; on behalf of the Dutch national COVID-19 response team. |
| EPI_ISL_3056750, EPI_ISL_3056849, EPI_ISL_3057362, EPI_ISL_3057834, EPI_ISL_3057842, EPI_ISL_3057849, EPI_ISL_3057855, EPI_ISL_3057864, EPI_ISL_3057871, EPI_ISL_3058099, EPI_ISL_3136585, EPI_ISL_3136617, EPI_ISL_3138350, EPI_ISL_3138360, EPI_ISL_3138368, EPI_ISL_3138370, EPI_ISL_3138405, EPI_ISL_3138419, EPI_ISL_3138424, EPI_ISL_3138429, EPI_ISL_3138430, EPI_ISL_3266858, EPI_ISL_3266859, EPI_ISL_3388928, EPI_ISL_3389111 | | | |
| see above | Dutch COVID-19 response team | National Institute for Public Health and the Environment (RIVM) | Adam Meijer; AnneMarie van den Brandt; Annelies Kroneman; Bas van der Veer; Chantal Reusken; Dennis Schmitz; Dirk Eggink; Euníce Then; Florian Zwagemaker; Harry Vennema; Jeroen Cremer; Karim Hajji; Kim Freriks; Lisa Wijsman; Lynn Aarts; Melissa van Tuil; Ryanne Jaarsma; Sanne Bos; Sharon van den Brink; Stijn van Rossum; on behalf of the national COVID-19 response team |
| EPI_ISL_3491507 | FL Bureau of Public Health Laboratories-Tampa | Centers for Disease Control and Prevention Division of Viral Diseases, Pathogen Discovery | Alex Burgin; Ben Rambo-Martin; Clinton Paden; Dakota Howard; Dave Wentworth; Dhwani Batra; Jasmine Padilla; Justin Lee; Krista Queen; Kristen Knipe; Kristine Lacek; Mark Burroughs; Matthew Schmerer; Meghan Bentz; Mili Sheth; Peter Cook; Sam Shepard; Sarah Nobles; Suxiang Tong; Vivien Dugan; Yvette Unoarumhi |
| EPI_ISL_3398814, EPI_ISL_3398815, EPI_ISL_3398819 | FUNDACION VALLE DE LILI | Instituto Nacional de Salud | Carlos Franco-Muñoz; Carmen Osorio; Diana Malo; Diego A. Álvarez-Díaz; Diego Andrés Prada; Gerardo Santamaría; Hector Alejandro Ruiz-Moreno; Jhonnatan Reales-González; Jorge Rivera; Juan Camilo Martinez; Julian Naizaque; Katherine Laiton-Donato; Lisseth Pardo; Magdalena Wiesner; Marcela Mercado-Reyes; Maria T.Herrera-Sepúlveda; Marta Lopez Blanco; Martha Lucia Ospina Martinez; Paola Rojas; Sergio Gomez; Sheryll Corchuelo; Ángela Alarcon Cruz |
| EPI_ISL_3369934 | FUNDACIÓN HOSPITAL SAN PEDRO | Corporacion CorpoGen-Universidad de los Andes-Universidad Central | Christian Romero; Jorge Duitama; Juan Manuel Anzola; Laura González; Maryam Chaib De Mares; María Mercedes Zambrano; Nelly Díaz; Patricia Del Portillo; Silvia Restrepo |
| EPI_ISL_3020727, EPI_ISL_3020836, EPI_ISL_3020842 | Fimlab Laboratoriot Oy Tampere | Expert Microbiology, National Institute for Health and Welfare | Carita Savolainen-Kopra; Erika Lindh; Haider al-Hello; Jani Halkilahti; Kirsi Liitsola; Niina Ikonen; Olli Vapalahti; Pekka Ellonen; Phuoc Truong; Päivi Laurila; Ravi Kant; Sari Hannula; Soile Blomqvist; Teemu Smura |
| EPI_ISL_3304939, EPI_ISL_3305727, EPI_ISL_3305743, EPI_ISL_3460963, EPI_ISL_3609634 | Fulgent Genetics | Centers for Disease Control and Prevention Division of Viral Diseases, Pathogen Discovery | Adrian Paskey; Becky Tsai; Benafsh Sapra; Benjamin Rambo-Martin; Christopher Gulvick; Clinton Paden; Clinton R. Paden; Dakota Howard; Darlene Wagner; Dhwani Batra; Doreen Ng; Duncan MacCannell; Harry Gao; James Xie; Jason Caravas; John Gao; Joseph Fierro; Kara Moser; Matthew Schmerer; Mickey Li; Peter Cook; Peter W. Cook; Scott Sammons; Shatavia Morrison; Yan Meng; Yvette Unoarumhi |
| EPI_ISL_3491809 | Gencore - Universidad de los Andes | Gencore - Universidad de los Andes | Catherine Jaller; Cristian Barrera; David González; Luisa Sacristan; Marcela Guevara; Silvia Restrepo |
| EPI_ISL_3185499, EPI_ISL_3185623, EPI_ISL_3185661, EPI_ISL_3185751, EPI_ISL_3185773, EPI_ISL_3185774, EPI_ISL_3185775, EPI_ISL_3185802 | | | |
| see above | Genetica Molecular and Subdepartamento de Virologia ISP Chile | Instituto de Salud Publica de Chile | Andres Castillo; Barbara Parra; Constanza Campano; Gisselle Barra; Javier Tognarelli; Jorge Fernandez; Karen Orostica; Loredana Arata; Patricia Bustos; Rodrigo Fasce; Soledad Ulloa |
| EPI_ISL_3464541 | HOSPITAL CIUDAD NEILY | Inciensa, Instituto Costarricense de Investigación y Enseñanza en Nutrición y Salud | Adriana Godínez; Claudio Soto-Garita; Estela Cordero; Francisco Duarte; Hebleen Porras; Joselyn Prado & Susana Mata Guerrero; José Luis Vargas; Mariela Gutiérrez; Melany Calderón |
| EPI_ISL_3274351 | HOSPITAL DR. CARLOS LUIS VALVERDE VEGA | Inciensa, Instituto Costarricense de Investigación y Enseñanza en Nutrición y Salud | Adriana Godínez; Caterina Guzmán; Claudio Soto-Garita; Estela Cordero; Francisco Duarte; Hebleen Porras; José Luis Vargas; Mariela Gutiérrez Joselyn Prado; Melany Calderón; Nazareth Ruiz & Charbel Vargas |
| EPI_ISL_3274363 | HOSPITAL DR. CARLOS LUIS VALVERDE VEGA [JESUS MARIA/SAN RAMON/ALAJUELA] | Inciensa, Instituto Costarricense de Investigación y Enseñanza en Nutrición y Salud | Adriana Godínez; Caterina Guzmán; Claudio Soto-Garita; Estela Cordero; Francisco Duarte; Hebleen Porras; José Luis Vargas; Mariela Gutiérrez Joselyn Prado; Melany Calderón; Nazareth Ruiz & Charbel Vargas |
| EPI_ISL_3639001 | HOSPITAL DR. FERNANDO ESCALANTE PRADILLA | Inciensa, Instituto Costarricense de Investigación y Enseñanza en Nutrición y Salud | Adriana Godínez; Claudio Soto-Garita; Estela Cordero; Francisco Duarte; Hebleen Porras; Joselyn Prado & María Fernanda Matamoros; José Luis Vargas; Mariela Gutiérrez; Melany Calderón |
| EPI_ISL_3274355, EPI_ISL_3298353 | HOSPITAL METROPOLITANO | Inciensa, Instituto Costarricense de Investigación y Enseñanza en Nutrición y Salud | Adriana Godínez; Caterina Guzmán; Claudio Soto-Garita; Estela Cordero; Francisco Duarte; Hebleen Porras; Joselyn Prado & Margarita Lee-Lui; José Luis Vargas; Mariela Gutiérrez; Mariela Gutiérrez Joselyn Prado; Melany Calderón; Nazareth Ruiz & Margarita Lee |
| EPI_ISL_3274358, EPI_ISL_3274359, EPI_ISL_3298342 | HOSPITAL SAN FRANCISCO DE ASIS | Inciensa, Instituto Costarricense de Investigación y Enseñanza en Nutrición y Salud | Adriana Godínez; Caterina Guzmán; Claudio Soto-Garita; Estela Cordero; Francisco Duarte; Hebleen Porras; Joselyn Prado & Adrián Fallas; José Luis Vargas; Mariela Gutiérrez; Mariela Gutiérrez Joselyn Prado; Melany Calderón; Nazareth Ruiz & Adrian Fallas Mora |
| EPI_ISL_3502573 | HOSPITAL UNIVERSITARIO 12 DE OCTUBRE | HOSPITAL UNIVERSITARIO 12 DE OCTUBRE | Carmen Martín-Higuera; Esther Viedma; Irene Múñoz-Gallego; M.ª Dolores Folgueira; Mar Aguilera; Noelia Moral; Rafael Delgado; Sagrario Zurita |
| EPI_ISL_3459404, EPI_ISL_3462744 | HOSPITAL UNIVERSITARIO HERNANDO MONCALEANO PERDOMO (HUHMP | Instituto Nacional de Salud | Carlos Franco-Muñoz; Carmen Osorio; Diana Malo; Diego A. Álvarez-Díaz; Diego Andrés Prada; Gerardo Santamaría; Hector Alejandro Ruiz-Moreno; Jhonnatan Reales-González; Jorge Rivera; Juan Camilo Martinez; Julian Naizaque; Katherine Laiton-Donato; Lisseth Pardo; Magdalena Wiesner; Marcela Mercado-Reyes; Maria T. Herrera-Sepúlveda; Marta Lopez Blanco; Martha Lucia Ospina Martinez; Paola Rojas; Sergio Gomez; Sheryll Corchuelo; Ángela Alarcon Cruz |

| | | | |
|---|---|---|---|
| EPI_ISL_3276551 | HPTU | Universidad Nacional de Colombia - Laboratorio Genómico One Health | Andres F. Cardona-Rios; Carlos Franco-Muñoz; Carolina Muñoz-Arango; Celeny Ortiz; Daniel O. Maldonado-Perez; Diego A. Álvarez-Díaz; Hector Alejandro Ruiz-Moreno; Idabely Betancur Ortiz; Jorge E. Osorio; Juan P. Hernandez-Ortiz; Karl A Ciuoderis; Katherine Laiton-Donato; Laura Silvana Perez; Lina M. Hurtado; Marcela Mercado-Reyes; Maria Angélica Maya; Maria Stella López; Rita Almanza Payares; Sandra Ines Cano; Simón Villegas Velásquez |
| EPI_ISL_3320721, EPI_ISL_3320726 | Hemato Oncologos | Instituto Nacional de Salud | Carlos Franco-Muñoz; Carmen Osorio; Diana Malo; Diego A. Álvarez-Díaz; Diego Andrés Prada; Gerardo Santamaría; Hector Alejandro Ruiz-Moreno; Jhonnatan Reales-González; Jorge Rivera; Juan Camilo Martinez; Julian Naizaque; Katherine Laiton-Donato; Lisseth Pardo; Magdalena Wiesner; Marcela Mercado-Reyes; Maria T. Herrera-Sepúlveda; Marta Lopez Blanco; Martha Lucia Ospina Martinez; Paola Rojas; Sergio Gomez; Sheryll Corchuelo; Ángela Alarcon Cruz |
| EPI_ISL_3061858, EPI_ISL_3061859 | Hospital | National Reference Center for Viruses of Respiratory Infections, Institut Pasteur, Paris | Alexandra Ducancelle; Angela Brisebarre; Camille Capel; Christophe Malabat; Corinne Maufrais; Etienne Simon-Lorière; Frédéric Lemoine; Hub de Bioinformatique et Biostatistique; Louise Lefrançois; Marion Barbet; Maud Vanpeene; Méline Bizard; Sylvie Behillil; Sylvie Van der Werf; Vincent Enouf |
| EPI_ISL_3050294 | Hospital Clínico Universitario Virgen de la Arrixaca | Hospital Clínico Universitario Virgen de la Arrixaca | Laura Moreno and Luis Gil-Gallardo; Marina Simón |
| EPI_ISL_3188867, EPI_ISL_3505956 | Hospital General Universitario Gregorio Marañón | Hospital General Universitario Gregorio Marañón | Cristina Rodriguez-Grande; Darío García de Viedma; Julia Suárez; Laura Pérez-Lago; Marta Herranz Martin; Patricia Muñoz; Pedro Sola Campoy; Pilar Catalán; Sergio Buenestado Serrano; Victor Manuel de la Cueva |
| EPI_ISL_3216188, EPI_ISL_3216194, EPI_ISL_3216216 | Hospital General Universitario de Alicante - Instituto de Investigación Sanitaria y Biomédica de Alicante | SeqCOVID-SPAIN consortium/IBV(CSIC) | Carmen Molina Pardines and SeqCOVID-SPAIN consortium; Maripaz Ventero Martín |
| EPI_ISL_3161803 | Hospital Universitari Vall d'Hebron - Vall d'Hebron Institut de Recerca | Hospital Universitari Vall d'Hebron - Vall d'Hebron Institut de Recerca | Alejandra González-Sánchez; Andrés Antón; Ariadna Rando; Carla Castillo; Cristina Andrés; Damir Garcia-Cehic; Josep Quer; Juliana Esperalba; Karen García; Maria Carmen Martin; Maria Gema Codina; Maria Piñana; Rodrigo Vásquez; Tomàs Pumarola |
| EPI_ISL_3276628, EPI_ISL_3276629 | ICMT-Apartado | Universidad Nacional de Colombia - Laboratorio Genómico One Health | Andres F. Cardona-Rios; Carlos Franco-Muñoz; Carolina Muñoz-Arango; Celeny Ortiz; Daniel O. Maldonado-Perez; Diego A. Álvarez-Díaz; Hector Alejandro Ruiz-Moreno; Idabely Betancur Ortiz; Jorge E. Osorio; Juan P. Hernandez-Ortiz; Karl A Ciuoderis; Katherine Laiton-Donato; Laura Silvana Perez; Lina M. Hurtado; Marcela Mercado-Reyes; Maria Angélica Maya; Maria Stella López; Rita Almanza Payares; Sandra Ines Cano; Simón Villegas Velásquez |
| EPI_ISL_3391981 | IMAT S.A.S. | Instituto Nacional de Salud-Dirección de Investigación en Salud Pública | Carlos Franco-Muñoz; Carmen Osorio; Diana Malo; Diego A. Álvarez-Díaz; Diego Andrés Prada; Gerardo Santamaría; Hector Alejandro Ruiz-Moreno; Jhonnatan Reales-González; Jorge Rivera; Juan Camilo Martinez; Julian Naizaque; Katherine Laiton-Donato; Lisseth Pardo; Magdalena Wiesner; Marcela Mercado-Reyes; Maria T. Herrera-Sepúlveda; Marta Lopez Blanco; Martha Lucia Ospina Martinez; Paola Rojas; Sergio Gomez; Sheryll Corchuelo; Ángela Alarcon Cruz |
| EPI_ISL_3104821, EPI_ISL_3104824, EPI_ISL_3579331 | INSIDE DIAGNÓSTICOS | Instituto Butantan | Antonio Jorge Martins; Claudia Renata dos Santos Barros; David Schlesinger; Debora Botequio Moretti; Dimas Tadeu Covas; Elaine Cristina Marqueze; Elaine Vieira Santos; Evandra Strazza Rodrigues; Heidge Fukumasu; Jayme Augusto de Souza-Neto; José Salvatore Leister Patané; Luiz Alcantara; Luiz Lehmann Coutinho; Maria Carolina Elias; Maurício Lacerda Nogueira; Rafael dos Santos Bezerra; Raul Machado Neto; Rejane Maria Tommasini Grotto; Ricardo Haddad; Sandra Coccuzzo Sampaio Vessoni; Simone Kashima; Svetoslav Nanev Slavov; Vincent Louis Viala |
| EPI_ISL_3031564 | INSIDE DIAGNÓSTICOS - UPA TATUAPÉ | Instituto Butantan | Antonio Jorge Martins; Claudia Renata dos Santos Barros; David Schlesinger; Debora Botequio Moretti; Dimas Tadeu Covas; Elaine Cristina Marqueze; Elaine Vieira Santos; Evandra Strazza Rodrigues; Heidge Fukumasu; Jayme Augusto de Souza-Neto; José Salvatore Leister Patané; Luiz Alcantara; Luiz Lehmann Coutinho; Maria Carolina Elias; Maurício Lacerda Nogueira; Rafael dos Santos Bezerra; Raul Machado Neto; Rejane Maria Tommasini Grotto; Ricardo Haddad; Sandra Coccuzzo Sampaio Vessoni; Simone Kashima; Svetoslav Nanev Slavov; Vincent Louis Viala |
| EPI_ISL_3133197, EPI_ISL_3274538 | INSPI-CRN DE INFLUENZA Y OTROS VIRUS RESPIRATORIOS | NIC-INSPI | Alfredo Bruno; Daniel Ramos; Domenica de Mora.; Jimmy Garcés; Johanna Laines; Lizbeth Patiño; Manuel Gonzalez; Maria Angelica Becerra; Maritza Olmedo; Mayra Wilca; Michelle Páez |
| EPI_ISL_3298687, EPI_ISL_3298689 | IVIC | Laboratorio de Virología Molecular | Carmen L Loureiro; CoViMol Group; Domingo J Garzaro; Esmeralda Vizzi; Flor H Pujol; Héctor R Rangel; José Luis Zambrano; Lieska Rodríguez; Mariana Hidalgo; Pierina D´Angelo; Rossana C Jaspe; Víctor Alarcón; Yoneira Sulbaran; Zoila Moros |
| EPI_ISL_3610849 | Infinity Biologix | Centers for Disease Control and Prevention Division of Viral Diseases, Pathogen Discovery | Adrian Paskey; Benjamin Rambo-Martin; Chirayu Goswami; Christian Bixby; Christopher Gulvick; Clinton Paden; Dakota Howard; Darlene Wagner; Dhwani Batra; Duncan MacCannell; Jason Caravas; Jonathan Schultz; Kara Moser; Matthew Schmerer; Peter Cook; Robin Grimwood; Russ Hager; Scott Sammons; Shatavia Morrison; Yihe Wang; Yvette Unoarumhi |
| EPI_ISL_3132358 | Institute of Microbiology, Universidad San Francisco de Quito | Institute of Microbiology, Universidad San Francisco de Quito | Alexandra Gonzalez; Belén Prado-Vivar; Bernardo Gutiérrez; Betty Angulo; Erika Muñoz; Fernanda Zurita; Francisco Guerra; Gabriel Trueba; Hugo Vergara; Jeaninna Peña; Juan José Guadalupe; Liu Yuqian; Luis Fuenmayor; Luz-Angelica Castillo; Marco Viteri Yanez; Michelle Grunauer; Michelle Jacome; Monica Becerra-Wong; Nancy Flores Lastra; Natalia Parra; Patricio Rojas-Silva; Paúl Cárdenas; Ronny Javier Pibaque; Stephanie Espín-Arroba; Sully Márquez; Valeria Armijos; Verónica Barragán |
| EPI_ISL_3316300 | Instituto Adolfo Lutz - Regional de Rio Claro | Instituto Adolfo Lutz, Interdiciplinary Procedures Center, Strategic Laboratory | Caio Vinicius Dias Lopes; Claudia Regina Gonçalves; Claudio Tavares Sacchi; Karoline Rodrigues Campos; Leonardo Tadeu de Araujo; Marlon Benedito Nascomento Santos |
| EPI_ISL_3507367, EPI_ISL_3507369, EPI_ISL_3507370 | Instituto Venezolano de Investigaciones Científicas | Laboratorio de Virología Molecular | Carmen L Loureiro; CoViVen Group; Domingo J Garzaro; Flor H Pujol; Héctor R Rangel; José Luis Zambrano; Lieska Rodríguez; Mariana Hidalgo; Pierina D´Angelo; Rossana C Jaspe; Víctor Alarcón; Yoneira Sulbaran; Zoila Moros |
| EPI_ISL_3188597, EPI_ISL_3188598, EPI_ISL_3188599, EPI_ISL_3188600, EPI_ISL_3188608, EPI_ISL_3188609, EPI_ISL_3236440, EPI_ISL_3236441, EPI_ISL_3236444, EPI_ISL_3236445, EPI_ISL_3236446, EPI_ISL_3236448, EPI_ISL_3236450, EPI_ISL_3236451 | | | |
| see above | Instituto de Medicina Tropical & Salud Global Universidad Iberoamericana | Grubaugh Lab - Yale School of Public Health | Alejandaro Vallejo Degaudenzi; Anderson Brito; Annie Watkins; Chaney Kalinich; Chantal Vogels; Elisa Contreras; Esperanza Mendoza; Isabel Ott; Jessica Rothman; Joseph Fauver; Kendall Billig; Mallery Breban; Mary Petrone; Nathan Grubaugh; Robert Paulino-Ramirez; Tara Alpert; Tobias Koch; Victor Virgilio Calderon |
| EPI_ISL_3071067 | Istituto Ortopedico Galeazzi | Laboratory of Clinical Microbiology, Virology and Bioemergencies, ASST Fatebenefratelli Sacco - Sacco Hospital | Valeria Micheli |
| EPI_ISL_3373121 | Johns Hopkins Hospital Department of Pathology | Johns Hopkins Hospital Department of Pathology | C. Paul Morris; Chun Huai Luo; David Gaston; Heba H. Mostafa; Julie M. Norton; Matthew Schwartz |
| EPI_ISL_3085775 | Karolinska University Hospital Solna | Karolinska University Hospital | Annelie Bjerkner; Isak Sylvin; Jan Albert; Karolina Ininbergs; Lina Guerra Blomqvist; Lynda Eneh; Martin Ekman; Martina Wahlund; Robert Dyrdak; Sandra Broddesson; Tanja Normark; Tobias Allander; Valtteri Wirta; Zhibing Yun |
| EPI_ISL_3462746 | LABORATORIO CLINICO BIO-TEST | Instituto Nacional de Salud | Carlos Franco-Muñoz; Carmen Osorio; Diana Malo; Diego A. Álvarez-Díaz; Diego Andrés Prada; Gerardo Santamaría; Hector Alejandro Ruiz-Moreno; Jhonnatan Reales-González; Jorge Rivera; Juan Camilo Martinez; Julian Naizaque; Katherine Laiton-Donato; Lisseth Pardo; Magdalena Wiesner; Marcela Mercado-Reyes; Maria T. Herrera-Sepúlveda; Marta Lopez Blanco; Martha Lucia Ospina Martinez; Paola Rojas; Sergio Gomez; Sheryll Corchuelo; Ángela Alarcon Cruz |
| EPI_ISL_3385791, EPI_ISL_3385842 | LABORATORIO CLINICO CRISTIAN GRAM IPS SAS | Instituto Nacional de Salud-Dirección de Investigación en Salud Pública | Carlos Franco-Muñoz; Carmen Osorio; Diana Malo; Diego A. Álvarez-Díaz; Diego Andrés Prada; Gerardo Santamaría; Hector Alejandro Ruiz-Moreno; Jhonnatan Reales-González; Jorge Rivera; Juan Camilo Martinez; Julian Naizaque; Katherine Laiton-Donato; Lisseth Pardo; Magdalena Wiesner; Marcela Mercado-Reyes; Maria T. Herrera-Sepúlveda; Marta Lopez Blanco; Martha Lucia Ospina Martinez; Paola Rojas; Sergio Gomez; Sheryll Corchuelo; Ángela Alarcon Cruz |
| EPI_ISL_3274357 | LABORATORIO CLINICO LABIN | Inciensa, Instituto Costarricense de Investigación y Enseñanza en Nutrición y Salud | Adriana Godínez; Caterina Guzmán; Claudio Soto-Garita; Estela Cordero; Francisco Duarte; Hebleen Porras; José Luis Vargas; Mariela Gutiérrez Joselyn Prado; Melany Calderón; Nazareth Ruiz & Pei Chan |
| EPI_ISL_3274352 | LABORATORIO CLINICO UNIVERSIDAD DE COSTA RICA (NUCLEO) | Inciensa, Instituto Costarricense de Investigación y Enseñanza en Nutrición y Salud | Adriana Godínez; Caterina Guzmán; Claudio Soto-Garita; Estela Cordero; Francisco Duarte; Hebleen Porras; José Luis Vargas; Mariela Gutiérrez Joselyn Prado; Melany Calderón; Nazareth Ruiz & Dimelsa Suarez |
| EPI_ISL_3246157 | LATE - Laboratório de Técnicas Especiais - Hospital Israelita Albert Einstein | LATE - Laboratório de Técnicas Especiais - Hospital Israelita Albert Einstein | Alexandre Hideaki Takara; Ana Paula Moreira Salles; Anelisie da Silva Santos; Deyvid Amgarten; Erick Gustavo Dorlass; Fernanda de Mello Malta; João Renato Rebello Pinho; Marcio Anunciacao Menezes; Pedro Henrique Sebe Rodrigues; Raquel Riyuzo |
| EPI_ISL_3459398 | LDSP CAQUETA | Instituto Nacional de Salud | Carlos Franco-Muñoz; Carmen Osorio; Diana Malo; Diego A. Álvarez-Díaz; Diego Andrés Prada; Gerardo Santamaría; Hector Alejandro Ruiz-Moreno; Jhonnatan Reales-González; Jorge Rivera; Juan Camilo Martinez; Julian Naizaque; Katherine Laiton-Donato; Lisseth Pardo; Magdalena Wiesner; Marcela Mercado-Reyes; Maria T. Herrera-Sepúlveda; Marta Lopez Blanco; Martha Lucia Ospina Martinez; Paola Rojas; Sergio Gomez; Sheryll Corchuelo; Ángela Alarcon Cruz |
| EPI_ISL_3459391 | LDSP CUNDINAMARCA | Instituto Nacional de Salud | Carlos Franco-Muñoz; Carmen Osorio; Diana Malo; Diego A. Álvarez-Díaz; Diego Andrés Prada; Gerardo Santamaría; Hector Alejandro Ruiz-Moreno; Jhonnatan Reales-González; Jorge Rivera; Juan Camilo Martinez; Julian Naizaque; Katherine Laiton-Donato; Lisseth Pardo; Magdalena Wiesner; Marcela Mercado-Reyes; Maria T. Herrera-Sepúlveda; Marta Lopez Blanco; Martha Lucia Ospina Martinez; Paola Rojas; Sergio Gomez; Sheryll Corchuelo; Ángela Alarcon Cruz |
| EPI_ISL_3127426 | Laboratoires d'analyses medicales - Ketterthill | Laboratoire national de sante, Microbiology, Microbial Genomics Platform | Anke Wienecke-Baldacchino; Caroline Scheiber; Catherine Ragimbeau; Elodie Solarino; Fatu Djabi; Jessica Tapp; Lise Pignon; Raoul Salmon; Serge Vedy; Tamir Abdelrahman; Virginie Jover |
| EPI_ISL_3045422 | Laboratorio Nacional de Salud Pública Dr. Defilló - LNSPDD | Laboratory of Respiratory Viruses and Measles, Oswaldo Cruz Institute, FIOCRUZ | Alice Sampaio Rocha; Ana Carolina Mendonca; Anna Carolina Paixao; Elisa Cavalcante Pereira; Fernando Motta; Grey Benoit Vasquez; Isaac Miguel Sanchez; Ivonne Imbert; Lucia de la Cruz; Luciana Appolinario; Marilda Siqueira on behalf of the Fiocruz COVID-19 Genomic Surveillance Network; Nurys de Castro; Paola Resende; Renata Serrano Lopes; Ronald Skewes; Taina Venas |
| EPI_ISL_3320742, EPI_ISL_3320747 | Laboratorio Olimpus | Instituto Nacional de Salud | Carlos Franco-Muñoz; Carmen Osorio; Diana Malo; Diego A. Álvarez-Díaz; Diego Andrés Prada; Gerardo Santamaría; Hector Alejandro Ruiz-Moreno; Jhonnatan Reales-González; Jorge Rivera; Juan Camilo Martinez; Julian Naizaque; Katherine Laiton-Donato; Lisseth Pardo; Magdalena Wiesner; Marcela Mercado-Reyes; Maria T. Herrera-Sepúlveda; Marta Lopez Blanco; Martha Lucia Ospina Martinez; Paola Rojas; Sergio Gomez; Sheryll Corchuelo; Ángela Alarcon Cruz |
| EPI_ISL_3376685, EPI_ISL_3376686, EPI_ISL_3376687, EPI_ISL_3376690, EPI_ISL_3376697, EPI_ISL_3376703, EPI_ISL_3376707, EPI_ISL_3376708, EPI_ISL_3376711, EPI_ISL_3376712, EPI_ISL_3376714, EPI_ISL_3376717, EPI_ISL_3376718, EPI_ISL_3376723, EPI_ISL_3376724, EPI_ISL_3376727 | | | |
| see above | Laboratorio de Biología Molecular - Universidad del Magdalena | Centro de Genética y Biología Molecular - Universidad del Magdalena | Andrea M. Ramírez Hernandez; Angel Oviedo Marquez; Daniel Bautista; Lyda R. Castro; Maria Teresa Mojica-Ortiz |
| EPI_ISL_3321115, EPI_ISL_3321116, EPI_ISL_3321309, EPI_ISL_3535716, EPI_ISL_3535748 | Laboratorio de Infectologia y Virologia Molecular | Laboratory of Molecular Virology, School of Medicine, Pontificia Universidad Catolica de Chile | Alejandro Bhrun; Ana Maria Contreras; Andres E. Munoz-Marcos; Carlos Palma; Catalina Pardo-Roa; Constanza Maldonado; Constanza Martinez-Valdevenito; Eileen Serrano; Erick Salinas; Estefany Poblete; Francisco Melo; Jennifer Angulo; Jorge Levican; Leonardo I. Almonacid; M. Belen Leyton; Magdalena Vera; Marcela Ferres; Maria Jose Avendano; Rafael A. Medina; Tamara Garcia-Salum |
| EPI_ISL_3654103, EPI_ISL_3654104, EPI_ISL_3654105, EPI_ISL_3654106, EPI_ISL_3654107, EPI_ISL_3655700 | Laboratorio de Referencia Nacional de Virus Respiratorios. Centro Nacional de Salud Publica. Instituto Nacional de Salud Peru. | Laboratorio de Referencia Nacional de Virus Respiratorios. Centro Nacional de Salud Publica. Instituto Nacional de Salud Peru. | Carlos Padilla Rojas; Henri Bailon Calderon; Iris Silva Molina; Joseph Huayra Niquen; Lely Solari Zerpa; Luis Barcena Flores; Marco Galarza Perez; Nancy Rojas Serrano; Nieves Sevilla Castañeda; Omar Caceres Rey; Orson Mestanza Millones; Princesa Medrano Alhuay; Priscila Lope Pari; Sandra Morales Ruiz; Sara Gordillo Vilchez; Steve Acedo Lazo; Veronica Hurtado Vela; Victor Jimenez Vasquez; Wendy Lizarraga Olivares |

| | | | |
|---|---|---|---|
| EPI_ISL_3055549 | Laboratorio de salud publica Puebla | LABOPAT | Cynthia Penaloza; Luis Mendoza; Silvia Montilla |
| EPI_ISL_3430864, EPI_ISL_3431052, EPI_ISL_3510253, EPI_ISL_3512112, EPI_ISL_3514445, EPI_ISL_3518226, EPI_ISL_3605427 | | | |
| see above | Laboratory Corporation of America | Centers for Disease Control and Prevention Division of Viral Diseases, Pathogen Discovery | Adrian Paskey; Amanda Douglas; Amanda Suchanek; Andrea Throop; Ayla Burns; Benjamin Rambo-Martin; Bobbi Croy; Brian Krueger; Brian Norvell; Christopher Gulvick; Christos Petropoulos; Clinton Paden; Clinton R. Paden; Craig Lukasik; Dakota Howard; Darlene Wagner; Debbie Boles; Dhwani Batra; Duncan MacCannell; Eyad Almasri; Goran Stevovic; Howard Engler; Hrushikesh Deshmukh; Jake Humphrey; Jana Schroth; Jason Caravas; Joe Voshell; John Pruitt; Jonathan Meltzer; Jonathan Williams; Kara Moser; Kimberly Wagner; Lax Iyer; Lisa Pfefferle; Lyndon Tilson; Manoj Jain; Marcia Eisenberg; Mary Ann Cristobal; Mary Cristobal; Mary Williamson; Matthew Robinson; Matthew Schmerer; Michael Levandoski; Mike Sapeta; Mindy Nye; Minoo Agarwal; Mohan Kolli; Nuthawin Charoensri; Oren Cohen; Peter Cook; Peter W. Cook; Prashant Gupta; Qian Zeng; Rama Ghatti; Scott Parker; Scott Ryan; Scott Sammons; Shatavia Morrison; Stanley Letovsky; Steven Ragan; Suresh Babu Selvaraju; Suresh Selvaraju; Susan Countryman; Susan Hicks; Suzanne Dale; Thomas Urban; Tim Kuphal; Tricia Zwiefelhofer; Vincent Drouillon; Yvette Unoarumhi |
| EPI_ISL_3583516 | Lighthouse Lab in Glasgow | Wellcome Sanger Institute for the COVID-19 Genomics UK (COG-UK) Consortium | Anna Dominiczak and Alex Alderton; Carol Clugston; Cordelia Langford; David Gray; David K. Jackson; Dominic Kwiatkowski; Ewan Harrison; Harper VanSteenhouse; Ian Johnston; Jeffrey Barrett; John Sillitoe on behalf of the Wellcome Sanger Institute COVID-19 Surveillance Team; Roberto Amato; Sonia Goncalves; Yumi Kasai |
| EPI_ISL_3042294, EPI_ISL_3095213, EPI_ISL_3095666, EPI_ISL_3096103, EPI_ISL_3108676, EPI_ISL_3171897 | Lighthouse Lab in Milton Keynes | Wellcome Sanger Institute for the COVID-19 Genomics UK (COG-UK) Consortium | Cordelia Langford; David K. Jackson; Dominic Kwiatkowski; Ewan Harrison; Ian Johnston; Jeffrey Barrett; John Sillitoe on behalf of the Wellcome Sanger Institute COVID-19 Surveillance Team; Roberto Amato; Sonia Goncalves; The Lighthouse Lab in Milton Keynes and Alex Alderton |
| EPI_ISL_3152228 | MD PHL | Maryland Department of Health Laboratories Administration | Ami Patel; Eric N. Keller; Jillian Loomis; Kwang Low; Terence L. Moore; and Robert Myers |
| EPI_ISL_3472780 | MIRIALIS CLUSES BECHET | CNR Virus des Infections Respiratoires - France SUD | Antonin Bal; Bruno Lina; Gregory Destras; Gwendolyne Burfin; Hadrien Regue; Laurence Josset; Martine Valette; Quentin Semanas |
| EPI_ISL_3085879, EPI_ISL_3086039 | Maine Health and Environmental Testing Laboratory | Tewhey Lab, The Jackson Laboratory | Barter, M.; Dewey, H.; H. and Tewhey, R.; Iosue, F.; Lynch, R.; Matluk, N.; Munger |
| EPI_ISL_3603488 | Mako Medical | Centers for Disease Control and Prevention Division of Viral Diseases, Pathogen Discovery | Adrian Paskey; Benjamin Rambo-Martin; Christopher Gulvick; Clinton Paden; Dakota Howard; Darlene Wagner; Dhwani Batra; Duncan MacCannell; Jason Caravas; Kara Moser; Lauren Moon; Matthew Schmerer; Matthew Tugwell; Peter Cook; Scott Sammons; Shatavia Morrison; Yvette Unoarumhi |
| EPI_ISL_3062529, EPI_ISL_3549736 | Maryland Genomics, Institute for Genome Sciences, University of Maryland School of Medicine | Maryland Genomics, Institute for Genome Sciences, University of Maryland School of Medicine | Claire M; Fraser; Hazen; Holly; Humphrys; Ivette; Jacques; Jonathan; Kranthi; Lim; Lisa D; Luke J; Mike; Ott; Ravel; Roussey; Sadzewicz; Sandra; Santana-Cruz; Tallon; Tracy; Vavikolanu |
| EPI_ISL_3332862 | Medyczne Laboratoria Diagnostyczne INVICTA | Wojewodzka Stacja Sanitarno-Epidemiologiczna w Lodzi, Oddzial Laboratoryjny Mikrobiologii i Parazytologii | Agnieszka Karnowska; Aleksandra Krawczyk; Danuta Bartczak; Elian Terpo; Katarzyna Kosowska; Lucyna Kochanowska; Marta Gawronska; Martyna Matera; Szymon Konkolewski |
| EPI_ISL_3251568 | Medyczne Laboratoria Diagnostyczne INVICTA | Wojewodzka Stacja Sanitarno-Epidemiologiczna w Rzeszowie, Laboratorium Diagnostyki Medycznej | Anna Nowakowska; Karolina Ostrowska; Katarzyna Wilk; Marzena Baranowska |
| EPI_ISL_3637925 | Michigan Department of Health and Human Services, Bureau of Laboratories | Michigan Department of Health and Human Services, Bureau of Laboratories | Blankenship HM; Riner D; Soehnlen MK |
| EPI_ISL_3241825, EPI_ISL_3470152, EPI_ISL_3470193, EPI_ISL_3470201 | Microbiology Department. Complexo Hospitalario Universitario de Vigo | Microbiology Department. Complexo Hospitalario Universitario de Vigo | Alfaya N; Alonso I; Alvarez M; Cabrera JJ; Carballo R; Cores O; Cortizo S; Davina C; Martinez L; Mediero G; Perez S; Potel C; Regueiro B; Rey S; Vasallo FJ; del-Campo V |
| EPI_ISL_3410989, EPI_ISL_3410990 | Ministry of Health Turkey | Ministry of Health Turkey | Fatma Bayrakdar; Gulay Korukluoglu; Suleyman Yalcin; Yasemin Cosgun |
| EPI_ISL_3319115 | NORTHWELL HEALTH LABORATORIES | Wadsworth Center, New York State Department of Health | Alexis Russell; Catharine Prussing; Daryl M. Lamson; Erasmus Schneider; Erica Lasek-Nesselquist; John Kelly; Jonathan Plitnick; Kirsten St. George; Matthew Shudt; Melissa A Leisner; Navjot Singh |
| EPI_ISL_3045698 | National Virus Reference Laboratory | National Virus Reference Laboratory | Charlene Bennett; Cillian F De Gascun; Gabriel Gonzalez; Jonathan Dean; Michael Carr; Zoe Yandle |
| EPI_ISL_3045076 | New Mexico Department of Health Scientific Laboratory | New Mexico Department of Health Scientific Laboratory | Anastacia Griego-Fisher; D'eldra Malone; Ellie Johnson; Jennifer Benoit; Linda Salazar; Ratheesh Rajan |
| EPI_ISL_3447229 | OLVZ Aalst | OLVZ Aalst | Astrid Holderbeke |
| EPI_ISL_3189600 | Orebro University Hospital, Dept Laboratory Medicine, Clinical Microbiology | Orebro University Hospital | Sundqvist M et al |
| EPI_ISL_3120108 | Ospedale di Vimercate | Laboratory of Clinical Microbiology, Virology and Bioemergencies, ASST Fatebenefratelli Sacco - Sacco Hospital | Valeria Micheli |
| EPI_ISL_3459407 | PREVIS - IPS | Instituto Nacional de Salud | Carlos Franco-Muñoz; Carmen Osorio; Diana Malo; Diego A. Álvarez-Díaz; Diego Andrés Prada; Gerardo Santamaría; Hector Alejandro Ruiz-Moreno; Jhonnatan Reales-González; Jorge Rivera; Juan Camilo Martinez; Julian Naizaque; Katherine Laiton-Donato; Lisseth Pardo; Magdalena Wiesner; Marcela Mercado-Reyes; Maria T. Herrera-Sepúlveda; Marta Lopez Blanco; Martha Lucia Ospina Martinez; Paola Rojas; Sergio Gomez; Sheryll Corchuelo; Ángela Alarcon Cruz |
| EPI_ISL_3188507, EPI_ISL_3339534, EPI_ISL_3394195 | Pandemic Response Lab - NYC | Pandemic Response Lab, R&D | Alex Carpio; Cybill del Castillo; Dylan Law; Haiping Hao; Henry Lee; Isabel Fernandez Escapa; Jon Laurent; Melissa Hopkins; Michael Hammerling; Pradeep Bugga; Shinyoung Clair Kang; Sol Rey; William Ward |
| EPI_ISL_3534151 | Pennsylvania Department of Health Bureau of Laboratories | Pennsylvania Department of Health Bureau of Laboratories | Dongxiang Xia |
| EPI_ISL_3233768 | Public Health Ontario Laboratory | Public Health Ontario Laboratory | Aimin Li; Alireza Eshaghi; Andre Villegas; Ashleigh Sullivan; Christine Frantz; Dean Maxwell; Esha Joshi; Jared Simpson; Jennifer L Guthrie; Jonathan B Gubbay; Karthikeyan Sivaraman; Lawrence Heisler; Matthew Watson; Michael CY Li; Michael Laszloffy; Nahuel Fittipaldi; Philip Banh; Richard de Borja; Samir N Patel; Sandeep Nagra; Sandra Zittermann; Sarah Teatero; Vanessa G Allen; Yao Chen; Yogi Sundaravadanam |
| EPI_ISL_3305019, EPI_ISL_3396975 | Quest Diagnostics Incorporated | Centers for Disease Control and Prevention Division of Viral Diseases, Pathogen Discovery | A. Gerasimova; A. Perez; Adrian Paskey; B. Anderson; Benjamin Rambo-Martin; Christopher Gulvick; Clinton R. Paden; Dakota Howard; Darlene Wagner; Dhwani Batra; Duncan MacCannell; F. Lacbawan; I. A. Shlyakhter; Jason Caravas; K.E. Livingston; Kara Moser; L.E. Bernstein; M. Hua; Matthew Schmerer; P. Tanpaiboon; Peter W. Cook; R. M. Kagan; R. Owen; R. V. Rolando; S. H. Rosenthal; Scott Sammons; Shatavia Morrison; Y. Liu; Yvette Unoarumhi |
| EPI_ISL_3080350, EPI_ISL_3080965, EPI_ISL_3081128, EPI_ISL_3179560, EPI_ISL_3179619, EPI_ISL_3291244, EPI_ISL_3291275, EPI_ISL_3291363, EPI_ISL_3291371, EPI_ISL_3291394, EPI_ISL_3291513, EPI_ISL_3292319, EPI_ISL_3292337, EPI_ISL_3292509, EPI_ISL_3292999, EPI_ISL_3573331, EPI_ISL_3573820, EPI_ISL_3573839, EPI_ISL_3573990, EPI_ISL_3574030, EPI_ISL_3574031, EPI_ISL_3574292, EPI_ISL_3574474, EPI_ISL_3574614 | | | |
| see above | Respiratory Virus Unit, Microbiology Services Colindale, Public Health England | COVID-19 Genomics UK (COG-UK) Consortium | PHE Covid Sequencing Team |
| EPI_ISL_3646351, EPI_ISL_3646354, EPI_ISL_3646358 | SARS-CoV-2 Sequencing Castilla y Leon-Spain Consortium | SARS-CoV-2 Sequencing Castilla y Leon-Spain Consortium | Antonio Orduña-Domingo; Carlos Fuster Foz; Carmen Aldea-Mansilla; Carmen Gimeno Crespo; David Abad; Gabriel March Rosello; Gregoria Megías Lobón; Jose María Eiros Bouza; M. Isabel Fernandez-Natal; Marta Dominguez-Gil; Marta Hernandez; María Antonia García Castro; Mª Fe Brezmes-Valdivieso; Noelia Arenal Andrés; Silvia Rojo; Sonsoles Garcinuño Pérez |
| EPI_ISL_3342048 | SC (UCO) Igiene e Sanità Pubblica, ASUGI, Trieste | ARGO Laboratorio Genomica ed Epigenomica | Barbone F; Braida C; Busetti M; D'Agaro P; Dal Monego S; Degasperi M; Licastro D; Maggiore A; Marcello A; Piscianz E; Segat L |
| EPI_ISL_3579329, EPI_ISL_3579330 | SECRETARIA MUNCIPAL DE SAUDE DE SANTA BARBARA D OESTE | Instituto Butantan | Antonio Jorge Martins; Claudia Renata dos Santos Barros; David Schlesinger; Debora Botequio Moretti; Dimas Tadeu Covas; Elaine Cristina Marqueze; Elaine Vieira Santos; Evandra Strazza Rodrigues; Heidge Fukumasu; Jayme Augusto de Souza-Neto; José Salvatore Leister Patané; Luiz Alcantara; Luiz Lehmann Coutinho; Maria Carolina Elias; Maurício Lacerda Nogueira; Rafael dos Santos Bezerra; Raul Machado Neto; Rejane Maria Tommasini Grotto; Ricardo Haddad; Sandra Coccuzzo Sampaio Vessoni; Simone Kashima; Svetoslav Nanev Slavov; Vincent Louis Viala |
| EPI_ISL_3369920, EPI_ISL_3369921, EPI_ISL_3369931, EPI_ISL_3369943 | SYNLAB - ANGEL DIAGNOSTICA | Corporacion CorpoGen-Universidad de los Andes-Universidad Central | Christian Romero; Jorge Duitama; Juan Manuel Anzola; Laura González; Maryam Chaib De Mares; María Mercedes Zambrano; Nelly Díaz; Patricia Del Portillo; Silvia Restrepo |
| EPI_ISL_3162242 | Salud Digna | Instituto Nacional de Medicina Genomica | Abraham Campos-Romero; Blancas S; Cedro-Tanda A; Cisneros-Villanueva M; Escobar-Arrazola; Gonzalez-Barrera D; Herrera-Montalvo LA.; Hidalgo-Miranda A; Luna-Ruiz Marco; M; Mendoza-Vargas A; Moreno-Camacho José Luis; Munguia-Garza P; Ramirez-Vega O; Rangel-DeLeon D; Reyes-Grajeda JP; Rodriguez-Gallegos Jorge |
| EPI_ISL_3373987 | Salud Digna, A.C | Andersen lab at Scripps Research | Abraham Garcia Gil; Jose Luis Moreno Camacho; Marco Antonio Luna Ruiz-Esparza; Miguel A. Fernandez Rojas; SEARCH Alliance with Abraham Campos Romero |
| EPI_ISL_3261726 | Scripps Medical Laboratory | Andersen lab at Scripps Research | Ellen Stefanski; Ian Mchardy; SEARCH Alliance San Diego with Michael Quigley |
| EPI_ISL_3391987 | Secretaria De Salud Del Guaviare | Instituto Nacional de Salud-Dirección de Investigación en Salud Pública | Carlos Franco-Muñoz; Carmen Osorio; Diana Malo; Diego A. Álvarez-Díaz; Diego Andrés Prada; Gerardo Santamaría; Hector Alejandro Ruiz-Moreno; Jhonnatan Reales-González; Jorge Rivera; Juan Camilo Martinez; Julian Naizaque; Katherine Laiton-Donato; Lisseth Pardo; Magdalena Wiesner; Marcela Mercado-Reyes; Maria T. Herrera-Sepúlveda; Marta Lopez Blanco; Martha Lucia Ospina Martinez; Paola Rojas; Sergio Gomez; Sheryll Corchuelo; Ángela Alarcon Cruz |

| | | | |
|---|---|---|---|
| EPI_ISL_3098720, EPI_ISL_3098721, EPI_ISL_3098722, EPI_ISL_3098723, EPI_ISL_3098724, EPI_ISL_3098725, EPI_ISL_3098726, EPI_ISL_3098727 | | | |
| see above | Section of Microbiology Department of Molecular and Translational Medicine, University of Brescia | Section of Microbiology Department of Molecular and Translational Medicine, University of Brescia | Alberto Zani; Anna Bertelli; Arnaldo Caruso; Francesca Caccuri; Giovanni Campisi; Serena Messali |
| EPI_ISL_3154873 | Shamir Medical Center (Asaf Harofe) | Shamir Medical Center (Asaf Harofe) | Abu Hamad Ramzia; Adina Bar Chaim; Anna Vishnevsky; Chen Weiner; Nir Rainy; Patricia Benveniste-Lekovitz; Reut Sorek Abramovich; Yevgeni Yegorov |
| EPI_ISL_3261400, EPI_ISL_3373781 | Sharp HealthCare Laboratory | Andersen lab at Scripps Research | Art Mendoza; Cathy Woerle; Jacquelyn Berumen; Liam McGinnis; Omid Bakhtar; SEARCH Alliance San Diego with Aaron Harding |
| EPI_ISL_3045364, EPI_ISL_3231558, EPI_ISL_3231561, EPI_ISL_3259958, EPI_ISL_3425894 | Stadtspital Triemli | Institute of Medical Virology | Alexandra Trkola; Annette Audigé; Catharine Aquino; Cyril Shah; Daniel Ehrsam; Gabriela Ziltener; Guido Bloemberg; Hubert Rehrauer; Isabel Stürmer; Joel Wirz; Jon Huder; Jürg Böni; Kevin Steiner; Maria Grünberg; Maryam Zaheri; Michael Huber; Riccarda Capaul; Stefan Schmutz; Verena Kufner; Weihong Qi |
| EPI_ISL_3326053 | TGen North | TGen North | Brett Van Tassel; Chris French; Darrin Lemmer; Dave Engelthaler; Hayley Yaglom; Heather Centner; Jolene Bowers |
| EPI_ISL_3383994, EPI_ISL_3384016 | TXDSHS | TXDSHS | Anita Pokharel; Bonnie Oh; Chun Wang; Grace Kubin; Jenny Zhang; Karen Bobier; Lorraine Rodriguez; Maliha Rahman; Mayela Pedrueza; Myong Koag; Rachel Lee; Rashmi Tuladhar |
| EPI_ISL_3373118 | The Jackson Laboratory | The Jackson Laboratory | Adams M; Kelly K; Li L; Long J; Omerza G; Renzette N |
| EPI_ISL_3321312, EPI_ISL_3370451 | UC-Christus Clinical Hospital Laboratory | Laboratory of Molecular Virology, School of Medicine, Pontificia Universidad Catolica de Chile | Ana Maira Guzman; Andres E. Munoz-Marcos; Catalina Pardo-Roa; Eileen Serrano; Erick Salinas; Estefany Poblete; Francisco Melo; Jorge Levican; Leonardo I. Almonacid; Maria Jose Avendano; Maria Patricia Vega; Rafael A. Medina; Ricardo Enrique de la Barra; Tamara Garcia-Salum |
| EPI_ISL_3105520 | UCC | Grubaugh Lab - Yale School of Public Health | Anderson Brito; Andrea Firpo; Annie Watkins; Chaney Kalinich; Chantal Vogels; David Rivera Aponte; Ernesto Vazquez; Fabiola Cruz-Lopez; Fabiola Fontanet; Isabel Ott; Jessica Rothman; Joseph Fauver; Karolane Gonzalez; Kendall Billig; Mallery Breban; Mary Petrone; Natalie Machargo; Nathan Grubaugh; Patricia Serrano; Tania Mitwalli; Tara Alpert; Tobias Koch; Vianca Aponte |
| EPI_ISL_3188574, EPI_ISL_3188576, EPI_ISL_3188577, EPI_ISL_3188578, EPI_ISL_3188591, EPI_ISL_3188612, EPI_ISL_3188613, EPI_ISL_3188614, EPI_ISL_3188615, EPI_ISL_3188618, EPI_ISL_3188619, EPI_ISL_3188620, EPI_ISL_3236562, EPI_ISL_3369783, EPI_ISL_3369786, EPI_ISL_3369787, EPI_ISL_3369788, EPI_ISL_3500991, EPI_ISL_3500998 | | | |
| see above | UCC (Universidad Central del Caribe) | Grubaugh Lab - Yale School of Public Health | Anderson Brito; Andrea Firpo; Annie Watkins; Chaney Kalinich; Chantal Vogels; David Rivera Aponte; Ernesto Vazquez; Fabiola Cruz-Lopez; Fabiola Fontanet; Isabel Ott; Jessica Rothman; Joseph Fauver; Karolane Gonzalez; Kendall Billig; Mallery Breban; Mary Petrone; Natalie Machargo; Nathan Grubaugh; Patricia Serrano; Tania Mitwalli; Tara Alpert; Tobias Koch; Vianca Aponte |
| EPI_ISL_3071377 | ULS Castelo Branco | Instituto Nacional de Saude (INSA) | Borges et al |
| EPI_ISL_3369936 | UNIVERSIDAD DEL QUINDIO | Corporacion CorpoGen-Universidad de los Andes-Universidad Central | Christian Romero; Jorge Duitama; Juan Manuel Anzola; Laura González; Maryam Chaib De Mares; María Mercedes Zambrano; Nelly Díaz; Patricia Del Portillo; Silvia Restrepo |
| EPI_ISL_3236599, EPI_ISL_3412244 | UW Virology Lab | UW Virology Lab | Alexander Greninger; Hong Xie; Keith R Jerome; Maria Lukes; Meei-Li Huang; Nathan Breit; Patrick Mathias; Pavitra Roychoudhury; Ricardo Perez; Robert J. Livingston; Sean Ellis; Shah Mohamed Bakhash; Tien V. Nguyen |
| EPI_ISL_3277390, EPI_ISL_3277391 | Unidad de Investigaciones Moleculares (UNIMOL) - Universidad de Cartagena | Centro de Genética y Biología Molecular - Universidad del Magdalena | Andrea M. Ramírez Hernandez; Angel Oviedo Marquez; Daniel Bautista; Lyda R. Castro; Maria Teresa Mojica-Ortiz |
| EPI_ISL_3072495 | Unidade de apoio ao diagnostico da COVID - UNADIG | Bioinformatics Laboratory / LNCC | Alessandra P Lamarca; Alexandra L Gerber; Amilcar Tanuri; Ana Paula de C Guimaraes; Ana Tereza R Vasconcelos; Andrea Cony Cavalcanti; Caio Luiz Pereira Ribeiro; Cassia Alves; Cintia Policarpo; Claudia Maria Braga de Mello; Cristiane Gomes da Silva; Diana Mariani; Douglas Terra Machado; Flavio Dias da Silva; Gleidson da Silva de Oliveira; Leandro Magalhaes de Souza; Liliane Cavalcante; Luiz G P de Almeida; Marcio Henrique de Oliveira Garcia; Mario Sergio Ribeiro; Ronaldo da Silva F Jr; Silvia Carvalho |
| EPI_ISL_3505613 | Universidad Estatal de Bolivar | Institute of Microbiology, Universidad San Francisco de Quito | Anita García; Belén Prado-Vivar; Bernardo Gutiérrez; Carlos Julio Tobar; Erika B. Muñoz; Fernanda Zurita; Gabriel Trueba; Juan José Guadalupe; Karina Barragán; Michelle Grunauer; Monica Becerra-Wong; Patricio Rojas-Silva; Paúl Cárdenas; René Bracho; Stephanie Arregui; Sully Márquez; Verónica Barragán |
| EPI_ISL_3276649, EPI_ISL_3276650 | Universidad Nacional de Colombia - Laboratorio Genómico One Health | Universidad Nacional de Colombia - Laboratorio Genómico One Health | Andres F. Cardona-Rios; Carlos Franco-Muñoz; Carolina Muñoz-Arango; Celeny Ortiz; Daniel O. Maldonado-Perez; Diego A. Álvarez-Díaz; Hector Alejandro Ruiz-Moreno; Idabely Betancur Ortiz; Jorge E. Osorio; Juan P. Hernandez-Ortiz; Karl A Ciuoderis; Katherine Laiton-Donato; Laura Silvana Perez; Lina M. Hurtado; Marcela Mercado-Reyes; Maria Angélica Maya; Maria Stella López; Rita Almanza Payares; Sandra Ines Cano; Simón Villegas Velásquez |
| EPI_ISL_3032253, EPI_ISL_3118424 | University of Liège COVID-19 testing center | GIGA Medical Genomics | Bouchra Boujemla; Claire Gourzonès; Cécile Meex; Keith Durkin; Laurent Gillet; Maria Artesi; Marie-Pierre Hayette; Nadine Cambisano; Nathalie Renotte; Olivier Ek; Sébastien Bontems; Vincent Bours |
| EPI_ISL_3259950 | Universität Zürich | Institute of Medical Virology | Alexandra Trkola; Annette Audigé; Catharine Aquino; Cyril Shah; Daniel Ehrsam; Gabriela Ziltener; Guido Bloemberg; Hubert Rehrauer; Isabel Stürmer; Joel Wirz; Jon Huder; Jürg Böni; Kevin Steiner; Maria Grünberg; Maryam Zaheri; Michael Huber; Riccarda Capaul; Stefan Schmutz; Verena Kufner; Weihong Qi |
| EPI_ISL_3236501, EPI_ISL_3347353, EPI_ISL_3369777, EPI_ISL_3369779, EPI_ISL_3369780, EPI_ISL_3369785 | VIDOH (Virgin Islands Department of Health) | Grubaugh Lab - Yale School of Public Health | Anderson Brito; Annie Watkins; Brett Ellis; Chaney Kalinich; Chantal Vogels; Esther Ellis; Isabel Ott; Jendai Richards; Jessica Rothman; Joseph Fauver; Kendall Billig; Mallery Breban; Marlon Lawrence; Mary Petrone; Nathan Grubaugh; TaLesa Aderohunmu; Tara Alpert; Tobias Koch |
| EPI_ISL_3123367, EPI_ISL_3123368 | WSSE w Gdansku | Wojewodzka Stacja Sanitarno-Epidemiologiczna w Olsztynie, Laboratorium Badan Epidemiologiczno-Klinicznych | Aleksandra Kobiatko; Barbara Dolinska; Emilia Tarabasz; Ewa Liszewska; Marta Lukian; Monika Czerminska; Patryk Bielecki; Paulina Rozycka; Sylwia Krzetowska; Tomasz Jakubczak |
| EPI_ISL_3587050 | Wisconsin State Laboratory of Hygiene Communicable Disease Division | Wisconsin State Laboratory of Hygiene Communicable Disease Division | Abigail C. Shockey; Alicia J. Mooney; Erika M. Hanson; Kelsey R. Florek; Richard Griesser; Sara Wagner; Tonya Danz |
| EPI_ISL_3188596 | Yale Clinical Virology Lab | Grubaugh Lab - Yale School of Public Health | Anderson Brito; Annie Watkins; Chaney Kalinich; Chantal Vogels; Isabel Ott; Jessica Rothman; Joseph Fauver; Kendall Billig; Mallery Breban; Marie L. Landry; Mary Petrone; Nathan Grubaugh; Tara Alpert; Tobias Koch |
| EPI_ISL_3539208 | ZLC Zentrallabor Chur | Microbiology, Dr. Risch | Dominique Fabien Hilti; Faina Wehrli; Lorenz Risch; Martin Risch; Nadia Wohlwend; Sinem Kas; Thomas Bodmer |
| EPI_ISL_3115647, EPI_ISL_3115672 | ZOTZ KLIMAS MVZ Düsseldorf-Centrum GbR ÜBAG für Labormedizin, Genetik, Zytologie, Pathologie | Center of Medical Microbiology, Virology, and Hospital Hygiene, University of Duesseldorf | Alexander Dilthey; Andreas Walker; Daniel Strelow; Jessica Nicolai; Jörg Timm; Katrin Hoffmann; Klaus Pfeffer; Lisanna Hülse; Malte Kohns Vasconcelos; Maximilian Damagnez; Nadine Lübke; Patrick Finzer; Rainer Zotz; Tobias Wienemann; Torsten Houwaart |

We gratefully acknowledge the following Authors from the Originating laboratories responsible for obtaining the specimens, as well as the Submitting laboratories where the genome data were generated and shared via GISAID, on which this research is based.

All Submitters of data may be contacted directly via www.gisaid.org

Authors are sorted alphabetically.

Acknowledgement EPI_SET Identifier: EPI_SET_20220328wg

| Accession ID | Originating Laboratory | Submitting Laboratory | Authors |
|---|---|---|---|
| EPI_ISL_3710325 | A. Krumbholz, Labor Dr. Krause und Kollegen MVZ GmbH, Kiel | Charité Universitätsmedizin Berlin, Institut für Virologie | Barbara Mühlemann; Christian Drosten; Julia Schneider; Jörn Beheim-Schwarzbach; Talitha Veith; Terry Jones; Victor M Corman |
| EPI_ISL_3878175 | AFIP | Instituto Butantan | Antonio Jorge Martins; Claudia Renata dos Santos Barros; David Schlesinger; Debora Botequio Moretti; Dimas Tadeu Covas; Elaine Cristina Marqueze; Elaine Vieira Santos; Evandra Strazza Rodrigues; Heidge Fukumasu; Jayme Augusto de Souza-Neto; José Salvatore Leister Patané; Luiz Alcantara; Luiz Lehmann Coutinho; Maria Carolina Elias; Maurício Lacerda Nogueira; Rafael dos Santos Bezerra; Raul Machado Neto; Rejane Maria Tommasini Grotto; Ricardo Haddad; Sandra Coccuzzo Sampaio Vessoni; Simone Kashima; Svetoslav Nanev Slavov; Vincent Louis Viala |
| EPI_ISL_4258609 | AREA DE SALUD ATENAS | Inciensa, Instituto Costarricense de Investigación y Enseñanza en Nutrición y Salud | Adriana Godínez; Claudio Soto-Garita; Estela Cordero; Francisco Duarte; Hebleen Porras; Joselyn Prado & Marilyn Alfaro Segura; José Luis Vargas; Mariela Gutiérrez; Melany Calderón |
| EPI_ISL_4258613 | AREA DE SALUD CORRALILLO | Inciensa, Instituto Costarricense de Investigación y Enseñanza en Nutrición y Salud | Adriana Godínez; Claudio Soto-Garita; Estela Cordero; Francisco Duarte; Hebleen Porras; Joselyn Prado & Natasha Tames Robles; José Luis Vargas; Mariela Gutiérrez; Melany Calderón |
| EPI_ISL_3948514 | AREA DE SALUD CORREDORES | Inciensa, Instituto Costarricense de Investigación y Enseñanza en Nutrición y Salud | Adriana Godínez; Claudio Soto-Garita; Estela Cordero; Francisco Duarte; Hebleen Porras; Joselyn Prado & Raul Zeledon; José Luis Vargas; Mariela Gutiérrez; Melany Calderón |
| EPI_ISL_3948546 | AREA DE SALUD DESAMPARADOS 2 (COOPESALUD) | Inciensa, Instituto Costarricense de Investigación y Enseñanza en Nutrición y Salud | Adriana Godínez; Claudio Soto-Garita; Estela Cordero; Francisco Duarte; Hebleen Porras; Joselyn Prado & Juliana Mora; José Luis Vargas; Mariela Gutiérrez; Melany Calderón |
| EPI_ISL_3948532 | AREA DE SALUD HEREDIA-VIRILLA | Inciensa, Instituto Costarricense de Investigación y Enseñanza en Nutrición y Salud | Adriana Godínez; Claudio Soto-Garita; Estela Cordero; Francisco Duarte; Hebleen Porras; José Luis Vargas; Mariela Gutiérrez & Joselyn Prado; Melany Calderón |
| EPI_ISL_3948547 | AREA DE SALUD PAVAS (COOPESALUD) | Inciensa, Instituto Costarricense de Investigación y Enseñanza en Nutrición y Salud | Adriana Godínez; Claudio Soto-Garita; Estela Cordero; Francisco Duarte; Hebleen Porras; Joselyn Prado & Juliana Mora; José Luis Vargas; Mariela Gutiérrez; Melany Calderón |
| EPI_ISL_3948539 | AREA DE SALUD PEREZ ZELEDON | Inciensa, Instituto Costarricense de Investigación y Enseñanza en Nutrición y Salud | Adriana Godínez; Claudio Soto-Garita; Estela Cordero; Francisco Duarte; Hebleen Porras; Joselyn Prado & Maria Fernanda Matamoros; José Luis Vargas; Mariela Gutiérrez; Melany Calderón |
| EPI_ISL_3753039, EPI_ISL_3816035, EPI_ISL_3819770, EPI_ISL_3819821, EPI_ISL_3819866, EPI_ISL_3820061, EPI_ISL_3820574, EPI_ISL_3820602, EPI_ISL_3822426, EPI_ISL_3822966, EPI_ISL_3843163, EPI_ISL_3843310, EPI_ISL_3844481, EPI_ISL_3849009, EPI_ISL_3851821, EPI_ISL_3852069, EPI_ISL_3852100, EPI_ISL_3855473, EPI_ISL_3857415, EPI_ISL_3857693, EPI_ISL_3860382, EPI_ISL_3860905, EPI_ISL_3863969, EPI_ISL_4186127, EPI_ISL_4186503, EPI_ISL_4186511, EPI_ISL_4360230, EPI_ISL_4360318, EPI_ISL_4361298, EPI_ISL_4361343, EPI_ISL_4361511, EPI_ISL_4361546, EPI_ISL_4361931, EPI_ISL_4362047, EPI_ISL_4362325, EPI_ISL_4362892, EPI_ISL_4362925, EPI_ISL_4363609, EPI_ISL_4364033, EPI_ISL_4364034, EPI_ISL_4364841, EPI_ISL_4365036, EPI_ISL_4365285, EPI_ISL_4365669, EPI_ISL_4366082, EPI_ISL_4366486 | | | |
| see above | Aegis Sciences Corporation | Centers for Disease Control and Prevention Division of Viral Diseases, Pathogen Discovery | Adrian Paskey; Alec Vest; Benjamin Rambo-Martin; Christopher Gulvick; Clinton Paden; Cyndi Clark; Dakota Howard; Darlene Wagner; Dhwani Batra; Dillon Nall; Duncan MacCannell; Erisa Sula; Ethan Sanders; Holly Houdeshell; Jason Caravas; Kara Moser; Kristine Lacek; Matthew Hardison; Matthew Schmerer; Ola Kvalvaag; Patrick Campbell; Peter Cook; Rob Case; Scott Sammons; Shatavia Morrison; Shaun Westlund; Tymeckia Kendall; Victoria Caban Figueroa; Vikramsinha Ghorpade; Yvette Unoarumhi |
| EPI_ISL_3668671, EPI_ISL_4072194, EPI_ISL_4199029 | Alaska State Virology Laboratory | Alaska State Virology Laboratory | Elva House; Jack Chen; Jacob Zidek; Lisa Smith; Ph.D.; Stephanie DeRonde |
| EPI_ISL_3721582 | Ayudas Diagnosticas Sura | Universidad Nacional de Colombia - Laboratorio Genómico One Health | Andres F. Cardona-Rios; Carlos Franco-Muñoz; Carolina Muñoz-Arango; Celeny Ortiz; Daniel O. Maldonado-Perez; Diego A. Álvarez-Díaz; Hector Alejandro Ruiz-Moreno; Idabely Betancur Ortiz; Jorge E. Osorio; Juan P. Hernandez-Ortiz; Karl A Ciuoderis; Katherine Laiton-Donato; Laura Silvana Perez; Lina M. Hurtado; Marcela Mercado-Reyes; Maria Angélica Maya; Maria Stella López; Rita Almanza Payares; Sandra Ines Cano; Simón Villegas Velásquez |
| EPI_ISL_3724153, EPI_ISL_4094266 | Broad Institute Clinical Research Sequencing Platform | Infectious Disease Program, Broad Institute of Harvard and MIT | Adams, G.; B.L.; B.W.; Bauer, M.; Birren; Blumenstiel, B.; Brown, C.; Carter, A.; Chaluvadi, S.; D.J.; DeFelice, M.; DeRuff, K.; Dodge, S.; Gabriel, S.; Gallagher, G.; Gladden-Young, A.; Granger, B.; J.E.; K.J.; Lagerborg, K.; Larkin, K.; Lee, M.; Lemieux; Lennon, N.; Loreth, C.; Madoff, L.; McGovern, S.; Meldrim, J.; Normandin, E.; P.C.; Park; Pearlman, L.; Reilly, S.; Rudy, M.; Sabeti; Siddle; Smole, S.; Tomkins-Tinch, C.; Vicente, G.; and MacInnis |
| EPI_ISL_3737595 | CHR Verviers | GIGA Medical Genomics | Bouchra Boujemla; Claire Gourzonès; Cécile Meex; Keith Durkin; Laurent Gillet; Maria Artesi; Marie-Pierre Hayette; Nadine Cambisano; Nathalie Renotte; Olivier Ek; Sébastien Bontems; Vincent Bours |
| EPI_ISL_3838194, EPI_ISL_3838195, EPI_ISL_3838196 | CHU NIMES | CHU NIMES | Agathe Boudet; Marie-Josée Carles; Sophie Bravo; Stephan Robin |
| EPI_ISL_3667141 | COVID LAB | Cayman Islands Forensic Science Lab | Brittany Balcewich; Jonathan Smellie; Karla Montes; Tanisha Gilbert |
| EPI_ISL_4299725 | Centro de Investigación Biomédica de Occidente (CIBO) | Centro de Investigación en Enfermedades Infecciosas (CIENI), Instituto Nacional de Enfermedades Respiratorias (INER) | Alejandra García-Gasca; Alejandra Hernández-Terán; Alejandro Sánchez-Flores; Alfredo Herrera-Estrella; Alicia Ocaña-Mondragón; Andreu Comas-García; Angel Gustavo Salas-Lais; Antonio Loza Román; Bernardo Martínez-Miguel; Blanca Taboada; Brenda Irasema Maldonado-Meza; Bruno Gómez-Gil; Carla Ivón Herrera-Najera; Carlos F. Arias; Celia Boukadida; Clara Esperanza Santacruz-Tinoco; Concepción Grajales-Muñiz; Consorcio Mexicano de Vigilancia Genómica (CoViGen-Mex). Authors (in alphabetical order): Julio Elias Alvarado-Yaah; Cristóbal Cháidez-Quiróz; Célida Duque Molina; Célida Martínez-Rodríguez; Daniel Fregoso-Rueda; Daniel Lira Morales; Eduardo Becerril-Vargas; Eduardo Rivera-Martínez; Fernando Fontove-Herrera; Fidencio Mejía-Nepomuceno; Francisco Pulido; Gabriel Chavira-Trujillo; Gloria Elena Espinosa-Ayala; Gloria María Molina-Salinas; Gloria Vazquez; Hector Montoya-Fuentes; Helen Haydee Fernanda Ramírez-Plascencia; Irvin González-López; Jean Pierre González; Jesús Hernández; Joel Armando Vázquez-Pérez.; Jorge Salas-Hernández; José Antonio Enciso-Moreno; José Arturo Martínez-Orozco; José Esteban Muñoz-Medina; José de Jesús Nuñez-Contreras; Juan Bautista Chale-Dzul; Julissa Enciso-Ibarra; Kathia Elizabeth Tapia-Diaz; Luis Alberto Ochoa-Carrera; Margarita Matías-Florentino; Mario Mújica-Sánchez; Marissa Perez-Garcia; María Eugenia Jiménez-Corona; María Guadalupe Santiago-Mauricio; María Guadalupe de Jesús Mireles-Rivera; Nelly Sélem-Mojica; Pavel Isa; Ricardo Ciria Merce; Ricardo Grande; Rosa María Gutiérrez Rios; Rosario Vazquez-Larios; Santiago Ávila-Ríos; Selene Zárate; Susana Lopez; Verónica Mata-Haro; Victor Eduardo García-Arias; Víctor Hugo Borja-Aburto |
| EPI_ISL_4006620, EPI_ISL_4006667 | Centro de Investigación Biomédica de Occidente (CIBO) | Instituto de Biotecnología de la UNAM | ; Alejandra García-Gasca; Alejandra Hernández-Terán; Alejandro Sánchez-Flores; Alfredo Herrera-Estrella; Alicia Ocaña-Mondragón; Andreu Comas-García; Angel Gustavo Salas-Lais; Antonio Loza Román; Bernardo Martínez-Miguel; Blanca Taboada; Brenda Irasema Maldonado-Meza; Bruno Gómez-Gil; Carla Ivón Herrera-Najera; Carlos F. Arias; Celia Boukadida; Clara Esperanza Santacruz-Tinoco; Concepción Grajales-Muñiz; Consorcio Mexicano de Vigilancia Genómica (CoViGen-Mex). Authors (in alphabetical order): Julio Elias Alvarado-Yaah; Cristóbal Cháidez-Quiróz; Célida Duque Molina; Célida Martínez- Rodríguez; Daniel Fregoso-Rueda; Daniel Lira Morales; Eduardo Becerril-Vargas; Fernando Fontove-Herrera; Fidencio Mejía-Nepomuceno; Francisco Pulido; Gloria Elena Espinosa-Ayala; Gloria María Molina-Salinas; Gloria Vazquez; Hector Esteban Paz-Juárez; Hector Montoya-Fuentes; Helen Haydee Fernanda Ramírez-Plascencia; Irvin González-López; Jean Pierre González; Jesús Hernández; Joel Armando Vázquez-Pérez).; Jorge Salas-Hernández; José Antonio Enciso-Moreno; José Arturo Martínez-Orozco; José Esteban Muñoz-Medina; José de Jesús Nuñez-Contreras; Juan Bautista Chale-Dzul; Julissa Enciso-Ibarra; Kathia Elizabeth Tapia-Diaz; Luis Alberto Ochoa-Carrera; Margarita Matías-Florentino; Mario Mújica-Sánchez; Marissa Perez-Garcia; María Guadalupe Santiago-Mauricio; María Guadalupe de Jesús Mireles-Rivera; Nelly Sélem-Mojica; Pavel Isa; Ricardo Ciria Merce; Ricardo Grande; Rosa María Gutiérrez Rios; Santiago Ávila-Ríos; Selene Zárate; Susana Lopez; Verónica Mata-Haro; Victor Eduardo García-Arias; Víctor Hugo Borja-Aburto |
| EPI_ISL_4006308 | Centro de Investigación Biomédica del Noreste (CIBIN) | Instituto de Biotecnología de la UNAM | ; Alejandra García-Gasca; Alejandra Hernández-Terán; Alejandro Sánchez-Flores; Alfredo Herrera-Estrella; Alicia Ocaña-Mondragón; Andreu Comas-García; Angel Gustavo Salas-Lais; Antonio Loza Román; Bernardo Martínez-Miguel; Blanca Taboada; Brenda Irasema Maldonado-Meza; Bruno Gómez-Gil; Carla Ivón Herrera-Najera; Carlos F. Arias; Celia Boukadida; Clara Esperanza Santacruz-Tinoco; Concepción Grajales-Muñiz; Consorcio Mexicano de Vigilancia Genómica (CoViGen-Mex). Authors (in alphabetical order): Julio Elias Alvarado-Yaah; Cristóbal Cháidez-Quiróz; Célida Duque Molina; Célida Martínez- Rodríguez; Daniel Fregoso-Rueda; Daniel Lira Morales; Eduardo Becerril-Vargas; Fernando Fontove-Herrera; Fidencio Mejía-Nepomuceno; Francisco Pulido; Gloria Elena Espinosa-Ayala; Gloria María Molina-Salinas; Gloria Vazquez; Hector Esteban Paz-Juárez; Hector Montoya-Fuentes; Helen Haydee Fernanda Ramírez-Plascencia; Irvin González-López; Jean Pierre González; Jesús Hernández; Joel Armando Vázquez-Pérez).; Jorge Salas-Hernández; José Antonio Enciso-Moreno; José Arturo Martínez-Orozco; José Esteban Muñoz-Medina; José de Jesús Nuñez-Contreras; Juan Bautista Chale-Dzul; Julissa Enciso-Ibarra; Kathia Elizabeth Tapia-Diaz; Luis Alberto Ochoa-Carrera; Margarita Matías-Florentino; Mario Mújica-Sánchez; Marissa Perez-Garcia; María Guadalupe Santiago-Mauricio; María Guadalupe de Jesús Mireles-Rivera; Nelly Sélem-Mojica; Pavel Isa; Ricardo Ciria Merce; Ricardo Grande; Rosa María Gutiérrez Rios; Santiago Ávila-Ríos; Selene Zárate; Susana Lopez; Verónica Mata-Haro; Victor Eduardo García-Arias; Víctor Hugo Borja-Aburto |
| EPI_ISL_4056032 | City of Milwaukee Health Department Laboratory | City of Milwaukee Health Department Laboratory | Amy Bauer; Manjeet Khubbar; Nandu Balakrishnan; Samantha Scott; Sanjib Bhattacharyya |
| EPI_ISL_3709760, EPI_ISL_3709761, | Department of Bacteria, Parasites and | Statens Serum Institut Bioinformatics and | Danish Covid-19 Genome Consortium |

| | | | |
|---|---|---|---|
| EPI_ISL_3709765 | Fungi, Statens Serum Institut, Copenhagen, Denmark | Microbial Genomics | |
| EPI_ISL_4006772 | Diagnóstico CIB | Corporación para Investigaciones Biológicas-CIB | Jeanneth Mosquera Rendon; Katterine Molina Hoyos; Marcela Mercado Reyes; Uriel A. Hurtado Paez |
| EPI_ISL_3771724, EPI_ISL_3771736, EPI_ISL_3771754, EPI_ISL_3771761, EPI_ISL_3771763, EPI_ISL_3771765, EPI_ISL_3771767, EPI_ISL_3771769, EPI_ISL_3771771, EPI_ISL_3771774, EPI_ISL_3771776, EPI_ISL_3917534, EPI_ISL_3917537, EPI_ISL_3917539, EPI_ISL_4076724, EPI_ISL_4077161, EPI_ISL_4077321, EPI_ISL_4077325 | | | |
| see above | Dutch COVID-19 response team | National Institute for Public Health and the Environment (RIVM) | Adam Meijer; AnneMarie van den Brandt; Annelies Kroneman; Bas van der Veer; Chantal Reusken; Dennis Schmitz; Dirk Eggink; Euníce Then; Florian Zwagemaker; Harry Vennema; Ivo van Walle; Jeroen Cremer; Karim Hajji; Kim Freriks; Lisa Wijsman; Lynn Aarts; Ryanne Jaarsma; Sanne Bos; Sharon van den Brink; Stijn van Rossum; on behalf of the national COVID-19 response team |
| EPI_ISL_3941383, EPI_ISL_3941388, EPI_ISL_3941390, EPI_ISL_3941391, EPI_ISL_3941392, EPI_ISL_3941393, EPI_ISL_3941394, EPI_ISL_3941395, EPI_ISL_3941396, EPI_ISL_3941397, EPI_ISL_3941402, EPI_ISL_3941405, EPI_ISL_3941406, EPI_ISL_3941407, EPI_ISL_3941409, EPI_ISL_3941413, EPI_ISL_3941418, EPI_ISL_3941422, EPI_ISL_3941424, EPI_ISL_3941425, EPI_ISL_3941426 | | | |
| see above | Edmonton Provincial Lab | Alberta Precision Labs (APL) | Buss; Croxen M; Deo A; Dieu P; E; Ferrato C; Gill K; Khan F; Koleva P; Li V; Lloyd C; Lynch T; Ma R; Murphy S; Pabbaraju K; Shokoples S; Thayer J; Tipples G; Whitehouse M; Wong A; Yu C; Zelyas N |
| EPI_ISL_3661081, EPI_ISL_3661421, EPI_ISL_3719766, EPI_ISL_4160689, EPI_ISL_4184283, EPI_ISL_4358179, EPI_ISL_4366972, EPI_ISL_4367467, EPI_ISL_4367935, EPI_ISL_4368833, EPI_ISL_4369438, EPI_ISL_4369471, EPI_ISL_4370473, EPI_ISL_4371167, EPI_ISL_4371547 | | | |
| see above | Fulgent Genetics | Centers for Disease Control and Prevention Division of Viral Diseases, Pathogen Discovery | Adrian Paskey; Becky Tsai; Benafsh Sapra; Benjamin Rambo-Martin; Christopher Gulvick; Clinton Paden; Dakota Howard; Darlene Wagner; Dhwani Batra; Doreen Ng; Duncan MacCannell; Erisa Sula; Harry Gao; James Xie; Jason Caravas; John Gao; Joseph Fierro; Kara Moser; Kristine Lacek; Matthew Schmerer; Mickey Li; Peter Cook; Scott Sammons; Shatavia Morrison; Tymeckia Kendall; Victoria Caban Figueroa; Yan Meng; Yvette Unoarumhi |
| EPI_ISL_4196877, EPI_ISL_4198483, EPI_ISL_4198494, EPI_ISL_4198499, EPI_ISL_4198500 | Gencore - Universidad de los Andes | Gencore - Universidad de los Andes | Cristian Barrera; David González; Gabriela Ariza; Luisa Sacristan; Marcela Guevara; Silvia Restrepo |
| EPI_ISL_3987681 | Genetica Molecular and Subdepartamento de Virologia ISP Chile | Instituto de Salud Publica de Chile | Andres Castillo; Barbara Parra; Constanza Campano; Gisselle Barra; Javier Tognarelli; Jorge Fernandez; Karen Orostica; Loredana Arata; Patricia Bustos; Rodrigo Fasce; Soledad Ulloa |
| EPI_ISL_3799749 | Gibraltar Health Authority Lab | Gibraltar Health Authority Covid-19 Laboratory | Dr Daniel Cassaglia; Dr Martyn Bell; Dr Nicholas Cortes; Dr Zoe Vincent |
| EPI_ISL_3761809, EPI_ISL_3761810 | HOSPITAL NACIONAL DE NIÑOS | Inciensa, Instituto Costarricense de Investigación y Enseñanza en Nutrición y Salud | Cristian Pérez-Corrales & Valeria Peralta-Barquero |
| EPI_ISL_4206000 | HOSPITAL UNIVERSITARIO 12 DE OCTUBRE | HOSPITAL UNIVERSITARIO 12 DE OCTUBRE | Carmen Martín-Higuera; Esther Viedma; Irene Múñoz-Gallego; M.ª Dolores Folgueira; Mar Aguilera; Noelia Moral; Rafael Delgado; Sagrario Zurita |
| EPI_ISL_3834651, EPI_ISL_3834652 | HOSPITAL UNIVERSITARIO CENTRAL DE ASTURIAS | Laboratorio de Virología HUCA | ; Alba L; Alvarez-Arguelles ME; Boga JA; Costales I; Coto E; González-Alba JM; Gómez de Oña J; Martín-Rodríguez G; Melón S; Perez-Martínez Z; Rojo S; Sandoval M |
| EPI_ISL_4345910, EPI_ISL_4346277, EPI_ISL_4346509, EPI_ISL_4347185, EPI_ISL_4347267, EPI_ISL_4347631 | Helix/Illumina | Centers for Disease Control and Prevention Division of Viral Diseases, Pathogen Discovery | Adrian Paskey; Alexandre Bolze; Ary Ascencio; Benjamin Rambo-Martin; Brad Sickler; Charlotte Rivera-Garcia; Christopher Gulvick; Chrstine Tran; Clinton Paden; Dakota Howard; Darlene Wagner; David Becker; Dhwani Batra; Duncan MacCannell; Efren Sandoval; Eileen De Feo; Elizabeth Cirulli; Eric Allen; Geraint Levan; James Lu; Jan Antico; Jason Caravas; Jason Nguyen; Jimmy Ramirez; Jingtao Liu; Kara Moser; Kelly Barrett; Kim Gietzen; Kristine Lacek; Magnus Isaksson; Marc Laurent; Matthew Schmerer; Matthew Tolentino; Nicole Washington; Peter Cook; Phil Febbo; Ryan Cho; Scott Sammons; Shannon Wickline; Shatavia Morrison; Sherry Wang; Simon White; Tyler Cassens; William Lee; Yvette Unoarumhi |
| EPI_ISL_3671012, EPI_ISL_3671057 | Hospital Fundacion San Vicente de Paul | Universidad Nacional de Colombia - Laboratorio Genómico One Health | Andres F. Cardona-Rios; Carlos Franco-Muñoz; Carolina Muñoz-Arango; Celeny Ortiz; Daniel O. Maldonado-Perez; Diego A. Álvarez-Díaz; Hector Alejandro Ruiz-Moreno; Idabely Betancur Ortiz; Jorge E. Osorio; Juan P. Hernandez-Ortiz; Karl A Ciuoderis; Katherine Laiton-Donato; Laura Silvana Perez; Lina M. Hurtado; Marcela Mercado-Reyes; Maria Angélica Maya; Maria Stella López; Rita Almanza Payares; Sandra Ines Cano; Simón Villegas Velásquez |
| EPI_ISL_4351984 | Hospital General Universitario Gregorio Marañón | Hospital General Universitario Gregorio Marañón | Cristina Rodriguez-Grande; Darío García de Viedma; Julia Suárez; Laura Pérez-Lago; Marta Herranz Martin; Patricia Muñoz; Pedro Sola Campoy; Pilar Catalán; Sergio Buenestado Serrano; Victor Manuel de la Cueva |
| EPI_ISL_3671065 | Hospital Pablo Tobon | Universidad Nacional de Colombia - Laboratorio Genómico One Health | Andres F. Cardona-Rios; Carlos Franco-Muñoz; Carolina Muñoz-Arango; Celeny Ortiz; Daniel O. Maldonado-Perez; Diego A. Álvarez-Díaz; Hector Alejandro Ruiz-Moreno; Idabely Betancur Ortiz; Jorge E. Osorio; Juan P. Hernandez-Ortiz; Karl A Ciuoderis; Katherine Laiton-Donato; Laura Silvana Perez; Lina M. Hurtado; Marcela Mercado-Reyes; Maria Angélica Maya; Maria Stella López; Rita Almanza Payares; Sandra Ines Cano; Simón Villegas Velásquez |
| EPI_ISL_4112985 | Hospital Universitari Arnau de Vilanova | Hospital Universitari Vall d'Hebron - Vall d'Hebron Institut de Recerca | Alejandra González-Sánchez; Andrés Antón; Ariadna Rando; Carla Castillo; Cristina Andrés; Damir Garcia-Cehic; Josep Quer; Juliana Esperalba; Karen García; Maria Carmen Martin; Maria Gema Codina; Maria Piñana; Rodrigo Vásquez; Tomàs Pumarola |
| EPI_ISL_3983941, EPI_ISL_3984119 | Hospital Universitari Vall d'Hebron - Vall d'Hebron Institut de Recerca | Hospital Universitari Vall d'Hebron - Vall d'Hebron Institut de Recerca | Alejandra González-Sánchez; Andrés Antón; Ariadna Rando; Carla Castillo; Cristina Andrés; Damir Garcia-Cehic; Josep Quer; Juliana Esperalba; Karen García; Maria Carmen Martin; Maria Gema Codina; Maria Piñana; Rodrigo Vásquez; Tomàs Pumarola |
| EPI_ISL_3721580, EPI_ISL_3721606 | ICMT-Apartado | Universidad Nacional de Colombia - Laboratorio Genómico One Health | Andres F. Cardona-Rios; Carlos Franco-Muñoz; Carolina Muñoz-Arango; Celeny Ortiz; Daniel O. Maldonado-Perez; Diego A. Álvarez-Díaz; Hector Alejandro Ruiz-Moreno; Idabely Betancur Ortiz; Jorge E. Osorio; Juan P. Hernandez-Ortiz; Karl A Ciuoderis; Katherine Laiton-Donato; Laura Silvana Perez; Lina M. Hurtado; Marcela Mercado-Reyes; Maria Angélica Maya; Maria Stella López; Rita Almanza Payares; Sandra Ines Cano; Simón Villegas Velásquez |
| EPI_ISL_4198473 | IDIME | Gencore - Universidad de los Andes | Cristian Barrera; David González; Gabriela Ariza; Luisa Sacristan; Marcela Guevara; Marcela Mercado; Silvia Restrepo |
| EPI_ISL_4344406 | Infinity Biologix | Centers for Disease Control and Prevention Division of Viral Diseases, Pathogen Discovery | Benjamin Rambo-Martin; Chirayu Goswami; Christian Bixby; Christopher Gulvick; Clinton Paden; Dakota Howard; Dhwani Batra; Duncan MacCannell; Erisa Sula; Jason Caravas; Jonathan Schultz; Kristine Lacek; Matthew Schmerer; Peter Cook; Robin Grimwood; Russ Hager; Scott Sammons; Shatavia Morrison; Tymeckia Kendall; Victoria Caban Figueroa; Yihe Wang; Yvette Unoarumhi |
| EPI_ISL_3721577 | Instituto Colombiano Medicina Tropical ICMT - Sede Sabaneta | Universidad Nacional de Colombia - Laboratorio Genómico One Health | Andres F. Cardona-Rios; Carlos Franco-Muñoz; Carolina Muñoz-Arango; Celeny Ortiz; Daniel O. Maldonado-Perez; Diego A. Álvarez-Díaz; Hector Alejandro Ruiz-Moreno; Idabely Betancur Ortiz; Jorge E. Osorio; Juan P. Hernandez-Ortiz; Karl A Ciuoderis; Katherine Laiton-Donato; Laura Silvana Perez; Lina M. Hurtado; Marcela Mercado-Reyes; Maria Angélica Maya; Maria Stella López; Rita Almanza Payares; Sandra Ines Cano; Simón Villegas Velásquez |
| EPI_ISL_3945527 | Instituto Nacional de Ciencias Médicas y Nutrición Salvador Zubirán (INCMNSZ) | Instituto Nacional de Medicina Genomica | Abraham Campos-Romero; Blancas S; Cedro-Tanda A; Cisneros-Villanueva M; Escobar-Arrazola MA; Herrera-Montalvo LA.; Hidalgo-Miranda A; Luna-Ruiz Marco; Mendoza-Vargas A; Moreno-Camacho José Luis; Ramirez-Vega O; Rangel-DeLeon D; Reyes-Grajeda JP; Rodriguez-Gallegos Jorge; Yair Alfaro-Mora |
| EPI_ISL_3982798 | Instituto Nacional de Enfermedades Respiratorias (INER) | Centro de Investigación en Enfermedades Infecciosas (CIENI), Instituto Nacional de Enfermedades Respiratorias (INER) | Alejandra García-Gasca; Alejandra Hernández-Terán; Alejandro Sánchez-Flores; Alfredo Herrera-Estrella; Alicia Ocaña-Mondragón; Andreu Comas-García; Angel Gustavo Salas-Lais; Antonio Loza Román; Bernardo Martínez-Miguel; Blanca Taboada; Brenda Irasema Maldonado-Meza; Bruno Gómez-Gil; Carla Ivón Herrera-Najera; Carlos F. Arias; Celia Boukadida; Clara Esperanza Santacruz-Tinoco; Concepción Grajales-Muñiz; Consorcio Mexicano de Vigilancia Genómica (CoViGen-Mex). Authors (in alphabetical order): Julio Elias Alvarado-Yaah; Cristóbal Cháidez-Quiróz; Célida Duque Molina; Célida Martínez-Rodríguez; Daniel Fregoso-Rueda; Daniel Lira Morales; Eduardo Becerril-Vargas; Eduardo Rivera-Martínez; Fernando Fontove-Herrera; Fidencio Mejía-Nepomuceno; Francisco Pulido; Gabriel Chavira-Trujillo; Gloria Elena Espinosa-Ayala; Gloria María Molina-Salinas; Gloria Vazquez; Hector Montoya-Fuentes; Helen Haydee Fernanda Ramírez-Plascencia; Irvin González-López; Jean Pierre González; Jesús Hernández; Joel Armando Vázquez-Pérez).; Jorge Salas-Hernández; José Antonio Enciso-Moreno; José Arturo Martínez-Orozco; José Esteban Muñoz-Medina; José de Jesús Nuñez-Contreras; Juan Bautista Chale-Dzul; Julissa Enciso-Ibarra; Kathia Elizabeth Tapia-Diaz; Luis Alberto Ochoa-Carrera; Margarita Matías-Florentino; Mario Mújica-Sánchez; Marissa Perez-Garcia; María Eugenia Jiménez-Corona; María Guadalupe Santiago-Mauricio; María Guadalupe de Jesús Mireles-Rivera; Nelly Sélem-Mojica; Pavel Isa; Ricardo Ciria Merce; Ricardo Grande; Rosa María Gutiérrez Rios; Rosario Vazquez-Larios; Santiago Ávila-Ríos; Selene Zárate; Susana Lopez; Verónica Mata-Haro; Victor Eduardo García-Arias; Víctor Hugo Borja-Aburto |
| EPI_ISL_4071007, EPI_ISL_4107171 | Instituto Nacional de Medicina Genomica | Instituto Nacional de Medicina Genomica | Blancas S; Cedro-Tanda A; Cisneros-Villanueva M; Escobar-Arrazola MA; Herrera-Montalvo LA.; Hidalgo-Miranda A; Mendoza-Vargas A; Ramirez-Vega O; Rangel-DeLeon D; Reyes-Grajeda JP; Yair Alfaro-Mora |
| EPI_ISL_3827499, EPI_ISL_3827558, EPI_ISL_3827767 | Instituto de Medicina Tropical & Salud Global, Universidad Iberoamericana (UNIBE) | Instituto de Medicina Tropical & Salud Global, Universidad Iberoamericana (UNIBE) | Alejandro; Calderon; Paulino-Ramirez; Robert; Vallejo Degaudenzi; Victor V. |
| EPI_ISL_3978876 | Integrated Covid Hub North East | Wellcome Sanger Institute for the COVID-19 Genomics UK (COG-UK) Consortium | Cordelia Langford; David K. Jackson; Dominic Kwiatkowski; Ewan Harrison; Ian Johnston; Integrated Covid Hub North East and Alex Alderton; Jeffrey Barrett; John Sillitoe on behalf of the Wellcome Sanger Institute COVID-19 Surveillance Team; Roberto Amato; Sonia Goncalves |
| EPI_ISL_4007284, EPI_ISL_4008105, EPI_ISL_4008108, EPI_ISL_4348705, EPI_ISL_4348711, EPI_ISL_4354278, EPI_ISL_4358318, EPI_ISL_4365206 | | | |
| see above | KU Leuven, Rega Institute, Clinical and | KU Leuven, Rega Institute, Clinical and | Bert Vanmechelen; Joan Marti-Carerras; Piet Maes; Tony Wawina-Bokalanga |

| | | | |
|---|---|---|---|
| | Epidemiological Virology | Epidemiological Virology | |
| EPI_ISL_3921333, EPI_ISL_3921335, EPI_ISL_3921336, EPI_ISL_3921341, EPI_ISL_3921343, EPI_ISL_3921357, EPI_ISL_3921365, EPI_ISL_3921376, EPI_ISL_3921379, EPI_ISL_3921386, EPI_ISL_3921389, EPI_ISL_3921391, EPI_ISL_3921395 | | | |
| see above | LDSP SAN ANDRES ISLA | Instituto Nacional de Salud- Dirección de Investigación en Salud Pública | Carlos Franco-Muñoz; Carmen Osorio; Diana Malo; Diego A. Álvarez-Díaz; Diego Andrés Prada; Gerardo Santamaría; Hector Alejandro Ruiz-Moreno; Jhonnatan Reales-González; Jorge Rivera; Juan Camilo Martinez; Julian Naizaque; Katherine Laiton-Donato; Lisseth Pardo; Magdalena Wiesner; Marcela Mercado-Reyes; Maria T. Herrera-Sepúlveda; Marta Lopez Blanco; Martha Lucia Ospina Martinez; Paola Rojas-Estevez; Sergio Gomez; Sheryll Corchuelo; Ángela Alarcon Cruz |
| EPI_ISL_4199478, EPI_ISL_4199479 | LESP Aguascalientes | Instituto de Diagnostico y Referencia Epidemiologicos (INDRE) | Abril Rodriguez-Maldonado; Ariadna Medina-Benitez; Claudia Wong-Arambula; Ernesto Ramirez-Gonzalez.; Gisela Barrera-Badillo; Irma Lopez-Martinez; Joaquin Quiroz-Mercado; Lucia Hernandez-Rivas; Maribel Gonzalez-Villa; Natividad Cruz-Ortiz; Sergio Rangel-Guerrero; Tatiana Nunez-Garcia; Vanessa Rivero-Arredondo |
| EPI_ISL_4299038 | LESP Michoacan | Instituto de Diagnostico y Referencia Epidemiologicos (INDRE) | Abril Rodriguez-Maldonado; Ariadna Medina-Benitez; Claudia Wong-Arambula; Ernesto Ramirez-Gonzalez.; Gisela Barrera-Badillo; Irma Lopez-Martinez; Joaquin Quiroz-Mercado; Lucia Hernandez-Rivas; Maribel Gonzalez-Villa; Natividad Cruz-Ortiz; Sergio Rangel-Guerrero; Tatiana Nunez-Garcia; Vanessa Rivero-Arredondo |
| EPI_ISL_4199471 | LESP Queretaro | Instituto de Diagnostico y Referencia Epidemiologicos (INDRE) | Abril Rodriguez-Maldonado; Ariadna Medina-Benitez; Claudia Wong-Arambula; Ernesto Ramirez-Gonzalez.; Gisela Barrera-Badillo; Irma Lopez-Martinez; Joaquin Quiroz-Mercado; Lucia Hernandez-Rivas; Maribel Gonzalez-Villa; Natividad Cruz-Ortiz; Sergio Rangel-Guerrero; Tatiana Nunez-Garcia; Vanessa Rivero-Arredondo |
| EPI_ISL_4196885, EPI_ISL_4196888, EPI_ISL_4196901, EPI_ISL_4212131, EPI_ISL_4212144, EPI_ISL_4212166 | LSP DISTRITAL BOGOTA | Gencore - Universidad de los Andes | Alejandro Gomez; Cristian Barrera; Gabriela Ariza; Gabriela Delgado; Johana Hernandez; Luisa Sacristan; Marcela Guevara; Marcela Mercado; Silvia Restrepo |
| EPI_ISL_3868377 | Lab voor klinische biologie | Lab voor klinische biologie | Bruno Verhasselt; Hannelore Hamerlinck; Marija Janevska |
| EPI_ISL_3998159 | Lab. Virología y Genética Universidad Simón Bolívar | Laboratorio de Biologia Molecular, Universidad Cooperativa de Colombia, Santa Marta | Andrew S. Muñoz-Gamba; Antonio Acosta; Daniel B. Ramírez-Osorio; José A. Usme-Ciro; Paula A. Quintero-Cortés; Yesid Bello |
| EPI_ISL_3801473, EPI_ISL_3999861 | Labo Analyses Med | National Reference Center for Viruses of Respiratory Infections, Institut Pasteur, Paris | Angela Brisebarre; Camille Capel; Christophe Malabat; Corinne Maufrais; Etienne Simon-Lorière; Frédéric Lemoine; Hub de Bioinformatique et Biostatistique; Julien Fumey; Louise Lefrançois; Marion Barbet; Maud Vanpeene; Méline Bizard; Slim El-Khiari; Sylvie Behillil; Sylvie Van der Werf; Vincent Enouf; Vincent Vieillefond |
| EPI_ISL_3947362 | Labor Dr. Fenner und Kollegen | Heinrich Pette Institute, Leibniz Institute for Experimental Virology | Adam Grundhoff; Alexis Robitaille; Johannes Knobloch; Martin Aepfelbacher; Nicole Fischer; Thomas Günther |
| EPI_ISL_4028962, EPI_ISL_4028964, EPI_ISL_4028966, EPI_ISL_4028973, EPI_ISL_4028977, EPI_ISL_4028981 | Laboratoire National de Santé Publique – LNSP | Genomics and Proteomics Departament, Gorgas Memorial Institute For Health Studies | Alexander A Martinez; Ambar Moreno; Claudia Estrada; Claudia Gonzalez V; César Roberto Conde Pereira; Jessica Gondola; Leyda Abrego; Marlene Castillo; Melissa Gaitan; Oris Chavarria |
| EPI_ISL_4006547 | Laboratorio Central de Epidemiología (LCE) | Instituto de Biotecnología de la UNAM | ; Alejandra García-Gasca; Alejandra Hernández-Terán; Alejandro Sánchez-Flores; Alfredo Herrera-Estrella; Alicia Ocaña-Mondragón; Andreu Comas-García; Angel Gustavo Salas-Lais; Antonio Loza Román; Bernardo Martínez-Miguel; Blanca Taboada; Brenda Irasema Maldonado-Meza; Bruno Gómez-Gil; Carla Ivón Herrera-Najera; Carlos F. Arias; Celia Boukadida; Clara Esperanza Santacruz-Tinoco; Concepción Grajales-Muñiz; Consorcio Mexicano de Vigilancia Genómica (CoViGen-Mex). Authors (in alphabetical order): Julio Elias Alvarado-Yaah; Cristóbal Cháidez-Quiróz; Célida Duque Molina; Célida Martínez- Rodríguez; Daniel Fregoso-Rueda; Daniel Lira Morales; Eduardo Becerril-Vargas; Fernando Fontove-Herrera; Fidencio Mejía-Nepomuceno; Francisco Pulido; Gloria Elena Espinosa-Ayala; Gloria María Molina-Salinas; Gloria Vazquez; Hector Esteban Paz-Juárez; Hector Montoya-Fuentes; Helen Haydee Fernanda Ramírez-Plascencia; Irvin González-López; Jean Pierre González; Jesús Hernández; Joel Armando Vázquez-Pérez).; Jorge Salas-Hernández; José Antonio Enciso-Moreno; José Arturo Martínez-Orozco; José Esteban Muñoz-Medina; José de Jesús Nuñez-Contreras; Juan Bautista Chale-Dzul; Julissa Enciso-Ibarra; Kathia Elizabeth Tapia-Diaz; Luis Alberto Ochoa-Carrera; Margarita Matías-Florentino; Mario Mújica-Sánchez; Marissa Perez-Garcia; María Guadalupe Santiago-Mauricio; María Guadalupe de Jesús Mireles-Rivera; Nelly Sélem-Mojica; Pavel Isa; Ricardo Ciria Merce; Ricardo Grande; Rosa María Gutiérrez Rios; Santiago Ávila-Ríos; Selene Zárate; Susana Lopez; Verónica Mata-Haro; Victor Eduardo García-Arias; Víctor Hugo Borja-Aburto |
| EPI_ISL_3671014, EPI_ISL_3671015 | Laboratorio Centrolab | Universidad Nacional de Colombia - Laboratorio Genómico One Health | Andres F. Cardona-Rios; Carlos Franco-Muñoz; Carolina Muñoz-Arango; Celeny Ortiz; Daniel O. Maldonado-Perez; Diego A. Álvarez-Díaz; Hector Alejandro Ruiz-Moreno; Idabely Betancur Ortiz; Jorge E. Osorio; Juan P. Hernandez-Ortiz; Karl A Ciuoderis; Katherine Laiton-Donato; Laura Silvana Perez; Lina M. Hurtado; Marcela Mercado-Reyes; Maria Angélica Maya; Maria Stella López; Rita Almanza Payares; Sandra Ines Cano; Simón Villegas Velásquez |
| EPI_ISL_4006785 | Laboratorio Clínico Citisalud | Corporación para Investigaciones Biológicas-CIB | Jeanneth Mosquera Rendon; Katterine Molina Hoyos; Marcela Mercado Reyes; Uriel A. Hurtado Paez |
| EPI_ISL_3998147, EPI_ISL_3998151 | Laboratorio Clínico Cristiam Gram | Laboratorio de Biologia Molecular, Universidad Cooperativa de Colombia, Santa Marta | Andrew S. Muñoz-Gamba; Daniel B. Ramírez-Osorio; Gloria A. Cuello; José A. Usme-Ciro; Katty Mindiola; Paula A. Quintero-Cortés; Yulieth L. Quiroz |
| EPI_ISL_4297520 | Laboratorio Clínico Nancy Florez García | Centro de Genética y Biología Molecular - Universidad del Magdalena | Andrea M. Ramírez Hernandez; Angel Oviedo Marquez; Daniel Bautista; Edison Lea-Ch; Lyda R. Castro; Maria Teresa Mojica-Ortiz |
| EPI_ISL_4297539, EPI_ISL_4297546 | Laboratorio Cristian Gram | Centro de Genética y Biología Molecular - Universidad del Magdalena | Andrea M. Ramírez Hernandez; Angel Oviedo Marquez; Daniel Bautista; Edison Lea-Ch; Lyda R. Castro; Maria Teresa Mojica-Ortiz |
| EPI_ISL_4219522, EPI_ISL_4219540, EPI_ISL_4219545 | Laboratorio IMAT Instituto médico de alta tecnología S.A.S | Molecular Genetics and Antimicrobial Resistance - UGRA, Universidad El Bosque | Catalina Espitia; Jinnethe Reyes; Lorena Diaz; Marcela Mercado; Mauricio Pacheco; Rafael Rios; Valentina Martinez |
| EPI_ISL_3671008, EPI_ISL_3671051 | Laboratorio Las Americas | Universidad Nacional de Colombia - Laboratorio Genómico One Health | Andres F. Cardona-Rios; Carlos Franco-Muñoz; Carolina Muñoz-Arango; Celeny Ortiz; Daniel O. Maldonado-Perez; Diego A. Álvarez-Díaz; Hector Alejandro Ruiz-Moreno; Idabely Betancur Ortiz; Jorge E. Osorio; Juan P. Hernandez-Ortiz; Karl A Ciuoderis; Katherine Laiton-Donato; Laura Silvana Perez; Lina M. Hurtado; Marcela Mercado-Reyes; Maria Angélica Maya; Maria Stella López; Rita Almanza Payares; Sandra Ines Cano; Simón Villegas Velásquez |
| EPI_ISL_3998114 | Laboratorio Lorena Vejarano sede Barranquilla | Laboratorio de Biologia Molecular, Universidad Cooperativa de Colombia, Santa Marta | Andrew S. Muñoz-Gamba; Daniel B. Ramírez-Osorio; Danis Lora; José A. Usme-Ciro; Lissette M. Lopez; Paula A. Quintero-Cortés |
| EPI_ISL_4297483, EPI_ISL_4297485 | Laboratorio Masvida de la Costa | Centro de Genética y Biología Molecular - Universidad del Magdalena | Andrea M. Ramírez Hernandez; Angel Oviedo Marquez; Daniel Bautista; Edison Lea-Ch; Lyda R. Castro; Maria Teresa Mojica-Ortiz |
| EPI_ISL_4220349, EPI_ISL_4220352, EPI_ISL_4220353, EPI_ISL_4220354, EPI_ISL_4220355, EPI_ISL_4220363, EPI_ISL_4220369, EPI_ISL_4220372, EPI_ISL_4220377, EPI_ISL_4220378, EPI_ISL_4220379 | | | |
| see above | Laboratorio Nacional de Salud Pública Dr. Defilló - LNSPDD | Laboratory of Respiratory Viruses and Measles, Oswaldo Cruz Institute, FIOCRUZ | ; Alice Sampaio Rocha; Ana Beatriz Machado Lima; Ana Carolina Mendonca; Anna Carolina Paixao; Elisa Cavalcante Pereira; Fernando Motta; Grey Benoit Vasquez; Isaac Miguel Sanchez; Ivonne Imbert; Lucia de la Cruz; Luciana Appolinario; Luiz Fernando Lopez Tort; Marilda Siqueira on behalf of the Fiocruz COVID-19 Genomic Surveillance Network; Mia Ferreira de Araujo; Natália Valente da Silva; Nurys de Castro; Paola Resende; Renata Serrano Lopes; Ronald Skewes; Taina Venas; Thayssa Keren Santos da Silva Neves |
| EPI_ISL_4029328, EPI_ISL_4029329 | Laboratorio Nacional de Salud, Ministerio de Salud Publica y Asistencia Social | Genomics and Proteomics Departament, Gorgas Memorial Institute For Health Studies | Alexander A Martinez; Ambar Moreno; Claudia Estrada; Claudia Gonzalez V; César Roberto Conde Pereira; Jessica Gondola; Leyda Abrego; Marlene Castillo; Melissa Gaitan; Oris Chavarria |
| EPI_ISL_4297505 | Laboratorio Rey-Fals | Centro de Genética y Biología Molecular - Universidad del Magdalena | Andrea M. Ramírez Hernandez; Angel Oviedo Marquez; Daniel Bautista; Edison Lea-Ch; Lyda R. Castro; Maria Teresa Mojica-Ortiz |
| EPI_ISL_3769570, EPI_ISL_3769571, EPI_ISL_3769572, EPI_ISL_3769573, EPI_ISL_3769576, EPI_ISL_3769577, EPI_ISL_3769581, EPI_ISL_3998033, EPI_ISL_3998034, EPI_ISL_3998035, EPI_ISL_3998038, EPI_ISL_3998180, EPI_ISL_3998184, EPI_ISL_3998195, EPI_ISL_3998199, EPI_ISL_3998201, EPI_ISL_3998202, EPI_ISL_4148466, EPI_ISL_4148467, EPI_ISL_4148476, EPI_ISL_4148484, EPI_ISL_4148485, EPI_ISL_4198084, EPI_ISL_4198096, EPI_ISL_4198098 | | | |
| see above | Laboratorio de Referencia Nacional de | Laboratorio de Referencia Nacional de | Carlos Padilla Rojas; Henri Bailon Calderon; Iris Silva Molina; Joseph Huayra Niquen; Lely Solari Zerpa; Luis Barcena Flores; Marco Galarza Perez; Nancy Rojas Serrano; Nieves Sevilla Castañeda; Omar Caceres Rey; Orson Mestanza Millones; Princesa Medrano Alhuay; Priscila Lope Pari; Sandra Morales Ruiz; Sara Gordillo Vilchez; Steve Acedo Lazo; Veronica Hurtado Vela; Victor Jimenez Vasquez; Wendy Lizarraga Olivares |

| | | | |
|---|---|---|---|
| | Virus Respiratorios. Centro Nacional de Salud Publica. Instituto Nacional de Salud Peru. | Virus Respiratorios. Centro Nacional de Salud Publica. Instituto Nacional de Salud Peru. | |
| EPI_ISL_3667219 | Laboratorio de Virología HUCA | Laboratorio de Virología HUCA | ; Alba L; Alvarez-Arguelles ME; Boga JA; Costales I; Coto E; González-Alba JM; Gómez de Oña J; Martín-Rodríguez G; Melón S; Perez-Martínez Z; Rojo S; Sandoval M |
| EPI_ISL_4196918, EPI_ISL_4197212, EPI_ISL_4212167, EPI_ISL_4212171 | Laboratorio de la Clínica Los Nogales | Gencore - Universidad de los Andes | Cristian Barrera; David González; Gabriela Ariza; Luisa Sacristan; Marcela Guevara; Marcela Mercado; Silvia Restrepo |
| EPI_ISL_3684751, EPI_ISL_3684971, EPI_ISL_3686276, EPI_ISL_3686480, EPI_ISL_3743419, EPI_ISL_3745925, EPI_ISL_3746461, EPI_ISL_4149236, EPI_ISL_4173053, EPI_ISL_4173473, EPI_ISL_4178156, EPI_ISL_4180800, EPI_ISL_4335425 | | | |
| see above | Laboratory Corporation of America | Centers for Disease Control and Prevention Division of Viral Diseases, Pathogen Discovery | Adrian Paskey; Amanda Douglas; Amanda Suchanek; Andrea Throop; Ayla Burns; Benjamin Rambo-Martin; Bobbi Croy; Brian Krueger; Brian Norvell; Christopher Gulvick; Christos Petropoulos; Clinton Paden; Craig Lukasik; Dakota Howard; Darlene Wagner; Debbie Boles; Dhwani Batra; Duncan MacCannell; Eyad Almasri; Goran Stevovic; Howard Engler; Hrushikesh Deshmukh; Jake Humphrey; Jana Schroth; Jason Caravas; Joe Voshell; John Pruitt; Jonathan Meltzer; Jonathan Williams; Kara Moser; Kimberly Wagner; Kristine Lacek; Lax Iyer; Lisa Pfefferle; Lyndon Tilson; Manoj Jain; Marcia Eisenberg; Mary Cristobal; Mary Williamson; Matthew Robinson; Matthew Schmerer; Michael Levandoski; Mike Sapeta; Mindy Nye; Minoo Agarwal; Mohan Kolli; Nuthawin Charoensri; Oren Cohen; Peter Cook; Prashant Gupta; Qian Zeng; Rama Ghatti; Scott Parker; Scott Ryan; Scott Sammons; Shatavia Morrison; Stanley Letovsky; Steven Ragan; Suresh Selvaraju; Susan Countryman; Susan Hicks; Suzanne Dale; Thomas Urban; Tim Kuphal; Tricia Zwiefelhofer; Tymeckia Kendall; Victoria Caban Figueroa; Vincent Drouillon; Yvette Unoarumhi |
| EPI_ISL_4060322 | MD DOH Laboratories Administration | Centers for Disease Control and Prevention Division of Viral Diseases, Pathogen Discovery | Alex Burgin; Ben Rambo-Martin; Clinton Paden; Dakota Howard; Dave Wentworth; Dhwani Batra; Jasmine Padilla; Justin Lee; Krista Queen; Kristen Knipe; Kristine Lacek; Mark Burroughs; Matthew Schmerer; Meghan Bentz; Mili Sheth; Peter Cook; Sam Shepard; Sarah Nobles; Suxiang Tong; Vivien Dugan; Yvette Unoarumhi |
| EPI_ISL_4083973 | Maine Health and Environmental Testing Laboratory | Tewhey Lab, The Jackson Laboratory | Barter, M.; Dewey, H.; H. and Tewhey, R.; Iosue, F.; Lynch, R.; Matluk, N.; Munger |
| EPI_ISL_4167559, EPI_ISL_4347742, EPI_ISL_4347822, EPI_ISL_4348028, EPI_ISL_4348090, EPI_ISL_4348107, EPI_ISL_4348124 | | | |
| see above | Mako Medical | Centers for Disease Control and Prevention Division of Viral Diseases, Pathogen Discovery | Adrian Paskey; Benjamin Rambo-Martin; Christopher Gulvick; Clinton Paden; Dakota Howard; Darlene Wagner; Dhwani Batra; Duncan MacCannell; Erisa Sula; Jason Caravas; Kara Moser; Kristine Lacek; Lauren Moon; Matthew Schmerer; Matthew Tugwell; Peter Cook; Scott Sammons; Shatavia Morrison; Tymeckia Kendall; Victoria Caban Figueroa; Yvette Unoarumhi |
| EPI_ISL_4178770 | Maryland Genomics, Institute for Genome Sciences, University of Maryland School of Medicine | Maryland Genomics, Institute for Genome Sciences, University of Maryland School of Medicine | Claire M; Fraser; Hazen; Holly; Humphrys; Ivette; Jacques; Jonathan; Kranthi; Lim; Lisa D; Luke J; Mike; Ott; Ravel; Roussey; Sadzewicz; Sandra; Santana-Cruz; Tallon; Tracy; Vavikolanu |
| EPI_ISL_4213081 | Michael E. DeBakery VA Medical Center | Genomics and Discovery, Respiratory Viruses Branch, Division of Viral Diseases, Centers for Disease Control and Prevention | Adam Retchless; Anna Kelleher; Anna Uehara; Brian Lynch; Clinton R. Paden; Dhwani Batra; Haibin Wang; Han Jia Justin Ng; Jasmine Padilla; Jing Zhang; Justin Lee; Mark Burroughs; Mili Sheth; Morgan Davis; Peter Cook; Rachel Marine; Sarah Nobles; Suxiang Tong; Tara Coalter; Yan Li; Ying Tao |
| EPI_ISL_3771045 | Microbiologia CATLAB | Can Ruti SARS-CoV-2 Sequencing Hub (HUGTiP/IrsiCaixa/IGTP) | Alba Sánchez; Alexia París; Anna Not; Antoni E Bordoy; Bonaventura Clotet; Cristina Casañ; David Panisello; Francesc Catala-Moll; Gemma Clara; Ignacio Blanco; Laia Soler; Lauro Sumoy; Marc Noguera-Julian; Maria Casadellà; Mariona Parera; Mercedes Guerrero; Montserrat Giménez; Pere-Joan Cardona; Pilar Armengol; Roger Paredes; Verónica Saludes; and Elisa Martró on behalf of the Can Ruti SARS-CoV-2 Sequencing Hub |
| EPI_ISL_3761813, EPI_ISL_4137465 | Microbiology Department, University Hospital Donostia | Microbiology Department, University Hospital Donostia | Cilla G.; Gomez M; Marimon JM; Martin-Peñaranda T; Montes M; Piñeiro L; Sorarrain A |
| EPI_ISL_4185684 | Microbiology Department. Complexo Hospitalario Universitario de Vigo | Microbiology Department. Complexo Hospitalario Universitario de Vigo | Alvarez M; Cabrera JJ; Carballo R; Cores O; Cortizo S; Davina C; Martinez L; Mediero G; Pena I; Perez S; Potel C; Regueiro B; Rey S; Vasallo FJ; del-Campo V |
| EPI_ISL_3914609 | Montana Public Health Laboratory | Montana Public Health Laboratory | Carrie Biskupiak; Deborah Gibson; Joy Ritter; Michael Dills; Michelle Mozer |
| EPI_ISL_4060407 | NYC Department of Health and Mental Hygiene | Centers for Disease Control and Prevention Division of Viral Diseases, Pathogen Discovery | Alex Burgin; Ben Rambo-Martin; Clinton Paden; Dakota Howard; Dave Wentworth; Dhwani Batra; Jasmine Padilla; Justin Lee; Krista Queen; Kristen Knipe; Kristine Lacek; Mark Burroughs; Matthew Schmerer; Meghan Bentz; Mili Sheth; Peter Cook; Sam Shepard; Sarah Nobles; Suxiang Tong; Vivien Dugan; Yvette Unoarumhi |
| EPI_ISL_3845461 | Naval Infectious Diseases Diagnostic Laboratory | Naval Medical Research Center Biological Defense Research Directorate | Andrea E. Luquette; Andrew J. Bennett; Catherine E. Arnold; Francisco Malagon; Gregory K. Rice; Kimberly A. Bishop-Lilly; Kyle A. Long; Lindsay A. Glang; Logan J. Voegtly; Megan A. Schilling; Michael V. Deschenes; Regina Z. Cer; Robin H. Miller; Victor A. Sugiharto |
| EPI_ISL_3825164 | Nevada State Public Health Laboratory | Nevada State Public Health Laboratory | Andrew Gorzalski; Mark Pandori |
| EPI_ISL_4221926 | Orange County Public Health Laboratory | Fulgent Genetics | Becky Tsai; Benafsh Sapra; Doreen Ng; Harry Gao; James Xie; John Gao; Joseph Fierro; Mickey Li; Yan Meng |
| EPI_ISL_3916157 | Oregon State Public Health Laboratory | Oregon State Public Health Laboratory | Eugene Yeboah; John Fontana and Shane Sevey; Laura Tsaknaridis; Rafia Razzaque; Vanda Makris |
| EPI_ISL_3933263, EPI_ISL_3933391, EPI_ISL_4096771 | Pandemic Response Lab - NYC | Pandemic Response Lab, R&D | Cybill del Castillo; Dylan Law; Haiping Hao; Henry Lee; Isabel Fernandez Escapa; Jon Laurent; Katharine Nelson; Melissa Hopkins; Michael Hammerling; Pradeep Bugga; Shinyoung Clair Kang; Sol Rey; William Ward |
| EPI_ISL_4182870 | Public Health Ontario Laboratory | Public Health Ontario Laboratory | Aimin Li; Alireza Eshaghi; Andre Villegas; Ashleigh Sullivan; Christine Frantz; Dean Maxwell; Esha Joshi; Jared Simpson; Jennifer L Guthrie; Jonathan B Gubbay; Karthikeyan Sivaraman; Lawrence Heisler; Matthew Watson; Michael CY Li; Michael Laszloffy; Nahuel Fittipaldi; Philip Banh; Richard de Borja; Samir N Patel; Sandeep Nagra; Sandra Zittermann; Sarah Teatero; Vanessa G Allen; Yao Chen; Yogi Sundaravadanam |
| EPI_ISL_4366811, EPI_ISL_4368810, EPI_ISL_4368977, EPI_ISL_4369780, EPI_ISL_4371143, EPI_ISL_4371445 | Quest Diagnostics Incorporated | Centers for Disease Control and Prevention Division of Viral Diseases, Pathogen Discovery | A. Gerasimova; A. Perez; B. Anderson; Benjamin Rambo-Martin; Christopher Gulvick; Clinton Paden; Dakota Howard; Dhwani Batra; Duncan MacCannell; Erisa Sula; F. Lacbawan; I. Shlyakhter; Jason Caravas; K. Livingston; Kristine Lacek; L. Bernstein; M. Hua; Matthew Schmerer; P. Tanpaiboon; Peter Cook; R. Kagan; R. Owen; R. Rolando; S. Rosenthal; Scott Sammons; Shatavia Morrison; Tymeckia Kendall; Victoria Caban Figueroa; Y. Liu; Yvette Unoarumhi |
| EPI_ISL_4060244 | Regional Hospital Liberec | Regional Hospital Liberec | Iva Dolinova; Katerina Arientova; Katerina Stillerova; Martin Kracik; Tomas Zajic |
| EPI_ISL_3775827, EPI_ISL_3777185 | Respiratory Virus Unit, Microbiology Services Colindale, Public Health England | COVID-19 Genomics UK (COG-UK) Consortium | PHE Covid Sequencing Team |
| EPI_ISL_3881334, EPI_ISL_3881812, EPI_ISL_3881832, EPI_ISL_3881841 | SARS-CoV-2 Sequencing Castilla y Leon-Spain Consortium | SARS-CoV-2 Sequencing Castilla y Leon-Spain Consortium | Antonio Orduña-Domingo; Carlos Fuster Foz; Carmen Aldea-Mansilla; Carmen Gimeno Crespo; David Abad; Gabriel March Rosello; Gregoria Megías Lobón; Jose María Eiros Bouza; M. Isabel Fernandez-Natal; Marta Dominguez-Gil; Marta Hernandez; María Antonia García Castro; Mª Fe Brezmes-Valdivieso; Noelia Arenal Andrés; Silvia Rojo; Sonsoles Garcinuño Pérez |
| EPI_ISL_3691311 | SYNLAB | GIGA Medical Genomics | Bouchra Boujemla; Claire Gourzonès; Cécile Meex; Keith Durkin; Laurent Gillet; Maria Artesi; Marie-Pierre Hayette; Nadine Cambisano; Nathalie Renotte; Olivier Ek; Sébastien Bontems; Vincent Bours |
| EPI_ISL_4071008, EPI_ISL_4071009, EPI_ISL_4199632 | Salud Digna | Instituto Nacional de Medicina Genomica | Abraham Campos-Romero; Blancas S; Cedro-Tanda A; Cisneros-Villanueva M; Cruz-Islas Jazmin; Escobar-Arrazola MA; Garnica-Lopez Dora; Herrera-Montalvo LA.; Hidalgo-Miranda A; Luna-Ruiz Marco; Mendoza-Vargas A; Moreno-Camacho José Luis; Ramirez-Vega O; Rangel-DeLeon D; Reyes-Grajeda JP; Rodriguez-Gallegos Jorge; Yair Alfaro-Mora |
| EPI_ISL_3913027 | Scripps Medical Laboratory | Andersen lab at Scripps Research | Ellen Stefanski; Ian Mchardy; SEARCH Alliance San Diego with Michael Quigley |
| EPI_ISL_4237131 | Stanford Health Care | Stanford University School of Medicine, Clinical Virology Laboratory | Becky Jiang; James Zehnder; Malaya K. Sahoo; Selamawit Bihon; and Benjamin A. Pinsky |
| EPI_ISL_3792632 | Swedish national genomic surveillance | The Public Health Agency of Sweden | Alma Brolund; Maria Lind Karlberg; Maximilian Riess; Swedish national genomic surveillance program of SARS-CoV-2 |

| | | | |
|---|---|---|---|
| | program of SARS-CoV-2 | | |
| EPI_ISL_4060851 | TXDSHS | TXDSHS | Anita Pokharel; Bonnie Oh; Chun Wang; Grace Kubin; Jenny Zhang; Karen Bobier; Lorraine Rodriguez; Maliha Rahman; Mayela Pedrueza; Myong Koag; Rachel Lee; Rashmi Tuladhar |
| EPI_ISL_3673913, EPI_ISL_3673914, EPI_ISL_3673915, EPI_ISL_3673916, EPI_ISL_3673917, EPI_ISL_3673918, EPI_ISL_3673919, EPI_ISL_3673920, EPI_ISL_3673921, EPI_ISL_3673922, EPI_ISL_3673923, EPI_ISL_3673924, EPI_ISL_3673925, EPI_ISL_3673926, EPI_ISL_3673927, EPI_ISL_3673928, EPI_ISL_3673929, EPI_ISL_3673930, EPI_ISL_3673931, EPI_ISL_3673932, EPI_ISL_3673933, EPI_ISL_3688253 | | | |
| see above | The Caribbean Public Health Agency | Carrington Lab, Department of Preclinical Sciences, Faculty of Medical Sciences, The University of the West Indies, St Augustine Campus | Anushka Ramjag; Arianne Brown-Jordan; Avery Hinds; Christine V. F. Carrington; Christopher Oura; Gabriel Escobar; Irad Potter; Nadia Ashwood; Nikita S. D. Sahadeo; Nuno Faria; Oliver Pybus; Risha Singh; Sarah Hill; SueMin Nathaniel; Vernie Ramkissoon |
| EPI_ISL_4169304 | US Air Force School of Aerospace Medicine | US Air Force School of Aerospace Medicine | Anthony Fries; Carol Garrett; Clarise Starr; Deanna Muehleman; Elizabeth Macias; Jennifer Meyer; Kelsey Lanter; Sarah Purves; William Gruner |
| EPI_ISL_4084898 | USC Clinical Lab | Los Angeles County Public Health Laboratories | P. Hemarajata et al. |
| EPI_ISL_3942173 | UW Virology Lab | UW Virology Lab | Alexander Greninger; Hong Xie; Keith R Jerome; Maria Lukes; Meei-Li Huang; Nathan Breit; Patrick Mathias; Pavitra Roychoudhury; Ricardo Perez; Robert J. Livingston; Sean Ellis; Shah Mohamed Bakhash; Tien V. Nguyen |
| EPI_ISL_4297514 | Universidad Simón Bolivar | Centro de Genética y Biología Molecular - Universidad del Magdalena | Andrea M. Ramírez Hernandez; Angel Oviedo Marquez; Daniel Bautista; Edison Lea-Ch; Lyda R. Castro; Maria Teresa Mojica-Ortiz |
| EPI_ISL_3721599 | Universidad de Caldas | Universidad Nacional de Colombia - Laboratorio Genómico One Health | Andres F. Cardona-Rios; Carlos Franco-Muñoz; Carolina Muñoz-Arango; Celeny Ortiz; Daniel O. Maldonado-Perez; Diego A. Álvarez-Díaz; Hector Alejandro Ruiz-Moreno; Idabely Betancur Ortiz; Jorge E. Osorio; Juan P. Hernandez-Ortiz; Karl A Ciuoderis; Katherine Laiton-Donato; Laura Silvana Perez; Lina M. Hurtado; Marcela Mercado-Reyes; Maria Angélica Maya; Maria Stella López; Rita Almanza Payares; Sandra Ines Cano; Simón Villegas Velásquez |
| EPI_ISL_3671047 | Universidad de Sucre | Universidad Nacional de Colombia - Laboratorio Genómico One Health | Andres F. Cardona-Rios; Carlos Franco-Muñoz; Carolina Muñoz-Arango; Celeny Ortiz; Daniel O. Maldonado-Perez; Diego A. Álvarez-Díaz; Hector Alejandro Ruiz-Moreno; Idabely Betancur Ortiz; Jorge E. Osorio; Juan P. Hernandez-Ortiz; Karl A Ciuoderis; Katherine Laiton-Donato; Laura Silvana Perez; Lina M. Hurtado; Marcela Mercado-Reyes; Maria Angélica Maya; Maria Stella López; Rita Almanza Payares; Sandra Ines Cano; Simón Villegas Velásquez |
| EPI_ISL_3998105, EPI_ISL_3998154 | Universidad del Atlántico Laboratorio de Investigación en Biología Molecular | Laboratorio de Biologia Molecular, Universidad Cooperativa de Colombia, Santa Marta | Andrew S. Muñoz-Gamba; Daniel B. Ramírez-Osorio; José A. Usme-Ciro; Paula A. Quintero-Cortés; Roberto Garcia |
| EPI_ISL_3914569, EPI_ISL_3914571, EPI_ISL_3914572 | Universidad del Valle, LDAB-Laboratorio de Diagnostico de Agentes Biologicos | Universidad del Valle | Programa Nacional de Caracterización Genómica de SARS-CoV-2 |
| EPI_ISL_3981839, EPI_ISL_3981840 | University of Liège COVID-19 testing center | GIGA Medical Genomics | Bouchra Boujemla; Claire Gourzonès; Cécile Meex; Keith Durkin; Laurent Gillet; Maria Artesi; Marie-Pierre Hayette; Nadine Cambisano; Nathalie Renotte; Olivier Ek; Sébastien Bontems; Vincent Bours |
| EPI_ISL_4348482 | Usansolo-Galdakao University Hospital | Cruces University Hospital | Ana Belén de la Hoz; Ana Gual-de-Torrella; Izaskun Alejo-Cancho; Mikel Gallego |
| EPI_ISL_3743387, EPI_ISL_3745405, EPI_ISL_4029470, EPI_ISL_4029482, EPI_ISL_4029605, EPI_ISL_4208554, EPI_ISL_4208573 | | | |
| see above | Utah Public Health Laboratory | Utah Public Health Laboratory | Erin L. Young; John Arnn; Kelly F. Oakeson; Olinto Linares-Perdomo; Pooja Gupta |
| EPI_ISL_3717248 | Valais Hospital, Central Institute | Valais Hospital, Central Institute | Alexis Dumoulin; Cedric Howald; Deborah Penet; Henri Pegeot; Ioannis Xenarios; Keith Harshman; Lorenzo Cerutti; Melyssa Elies |
| EPI_ISL_3673830 | Washington State Department of Health Public Health Laboratories | Washington State Department of Health Public Health Laboratories | Avi Singh; Darren Lucas; Denny Russell; Drew MacKellar; Geoff Melly; Hannah Gray; Joenice Gonzalez; JohnAric Peterson; Philip Dykema; Rebecca Cao; Vanessa De Los Santos |
| EPI_ISL_3742030 | Wyoming Public Health Laboratory | Wyoming Public Health Laboratory | Ashley Norberg; Brian Dominguez; Cari Sloma; Channing Weber; Chayse Rowley; Elliot Thomasson; Jim Mildenberger; Marley Goetz; Robert Petit; Sam Britz; Taylor Fearing; and Rob Christensen |
| EPI_ISL_4197547, EPI_ISL_4197999 | Yale Clinical Virology Lab | Grubaugh Lab - Yale School of Public Health | Anderson Brito; Annie Watkins; Chaney Kalinich; Chantal Vogels; Isabel Ott; Jessica Rothman; Joseph Fauver; Kendall Billig; Mallery Breban; Marie L. Landry; Mary Petrone; Nathan Grubaugh; Tara Alpert; Tobias Koch |

We gratefully acknowledge the following Authors from the Originating laboratories responsible for obtaining the specimens, as well as the Submitting laboratories where the genome data were generated and shared via GISAID, on which this research is based.

All Submitters of data may be contacted directly via www.gisaid.org

Authors are sorted alphabetically.

Acknowledgement EPI_SET Identifier: EPI_SET_20220328zg

| Accession ID | Originating Laboratory | Submitting Laboratory | Authors |
|---|---|---|---|
| EPI_ISL_5058060 | "SYNLAB ANGEL DIAGNOSTICA" | "Laboratorio de biotecnología, Universidad Icesi" | "María I. Gutiérrez López; Adrián Camilo Rodríguez Ararat; Diana M. Florez Giraldo; Marcela Mercado; Maria F. Villegas Torres; Paola A. Caicedo Burbano"; Programa Nacional de Caracterización Genómica de SARS-CoV-2; Sara González Henao |
| EPI_ISL_5058034, EPI_ISL_5058035, EPI_ISL_5058069 | "Unidad de Diagnóstico Hemato Oncológico - UDHO" | "Laboratorio de biotecnología, Universidad Icesi" | "María I. Gutiérrez López; Adrián Camilo Rodríguez Ararat; Diana M. Florez Giraldo; Marcela Mercado; Maria F. Villegas Torres; Paola A. Caicedo Burbano"; Programa Nacional de Caracterización Genómica de SARS-CoV-2; Sara González Henao |
| EPI_ISL_5797245 | "Unidad de Diagnóstico Hemato Oncológico - UDHO" | Laboratorio de biotecnología, Universidad Icesi | "María I. Gutiérrez López; Adrián Camilo Rodríguez Ararat; Diana M. Florez Giraldo; Marcela Mercado; Maria F. Villegas Torres; Paola A. Caicedo Burbano"; Programa Nacional de Caracterización Genómica de SARS-CoV-2; Sara González Henao |
| EPI_ISL_4960541, EPI_ISL_4984496, EPI_ISL_4984513, EPI_ISL_5428495 | ADILAB | Laboratorio Departamental de Salud Publica de Antioquia | Ana Victoria Valencia Duarte; Andres F. Cardona-Rios; Cristian Arbey Velarde Hoyos; Gloria Isabel Escobar; Idabely Betancur Ortiz; Juan P. Hernandez-Ortiz; Juan Pablo Isaza Agudelo; Maria Stella López |
| EPI_ISL_5914875 | AREA DE SALUD BARVA (COOPESIBA) | Inciensa, Instituto Costarricense de Investigación y Enseñanza en Nutrición y Salud | Adriana Godínez; Claudio Soto-Garita; Estela Cordero; Francisco Duarte; Hebleen Porras; José Luis Vargas; Mariela Gutiérrez; Melany Calderón; Sofia Herrera & Angélica Espinoza Fontana |
| EPI_ISL_4659127 | AUSTRAL-omics, UACh | AUSTRAL-omics, UACh | Andrea Silva; Carolina Encina; Cristian Molina; Daniela Plaza; Luis Guzmán; Suany Quesada |
| EPI_ISL_4850558, EPI_ISL_4850560, EPI_ISL_4850566, EPI_ISL_4850567 | AYUDAS DIAGNOSTICAS SURA | Instituto Nacional de Salud-Dirección de Investigación en Salud Pública | Carlos Franco-Muñoz; Carmen Osorio; Diana Malo; Diego A. Álvarez-Díaz; Diego Andrés Prada; Gerardo Santamaría; Hector Alejandro Ruiz-Moreno; Jhonnatan Reales-González; Jorge Rivera; Juan Camilo Martinez; Julian Naizaque; Katherine Laiton-Donato; Lisseth Pardo; Magdalena Wiesner; Marcela Mercado-Reyes; Maria T. Herrera-Sepúlveda; Marta Lopez Blanco; Martha Lucia Ospina Martinez; Paola Rojas; Sergio Gomez; Sheryll Corchuelo; Ángela Alarcon Cruz |
| EPI_ISL_5803646 | AYUDAS DIAGNOSTICAS SURA | Universidad Nacional de Colombia - Laboratorio Genómico One Health | Andres F. Cardona-Rios; Carlos Franco-Muñoz; Carolina Muñoz-Arango; Celeny Ortiz; Daniel O. Maldonado-Perez; Diego A. Álvarez-Díaz; Hector Alejandro Ruiz-Moreno; Idabely Betancur Ortiz; Jorge E. Osorio; Juan P. Hernandez-Ortiz; Karl A Ciuoderis; Katherine Laiton-Donato; Laura Silvana Perez; Lina M. Hurtado; Marcela Mercado-Reyes; Maria Angélica Maya; Maria Stella López; Rita Almanza Payares; Sandra Ines Cano; Simón Villegas Velásquez |
| EPI_ISL_4376769, EPI_ISL_5108824, EPI_ISL_5215412, EPI_ISL_5229610, EPI_ISL_5860239, EPI_ISL_6164570 | Aegis Sciences Corporation | Centers for Disease Control and Prevention Division of Viral Diseases, Pathogen Discovery | Alec Vest; Benjamin Rambo-Martin; Christopher Gulvick; Clinton Paden; Cyndi Clark; Dakota Howard; Dhwani Batra; Dillon Nall; Duncan MacCannell; Erisa Sula; Ethan Sanders; Holly Houdeshell; Jason Caravas; Kristine Lacek; Matthew Hardison; Matthew Schmerer; Ola Kvalvaag; Patrick Campbell; Peter Cook; Rob Case; Scott Sammons; Shatavia Morrison; Shaun Westlund; Tymeckia Kendall; Victoria Caban Figueroa; Vikramsinha Ghorpade; Yvette Unoarumhi |
| EPI_ISL_4574480 | Alaska State Virology Laboratory | Alaska State Virology Laboratory | Elva House; Jack Chen; Jacob Zidek; Lisa Smith; Ph.D.; Stephanie DeRonde |
| EPI_ISL_4960533 | BIORREFERENCIA | Laboratorio Departamental de Salud Publica de Antioquia | Cristian Arbey Velarde Hoyos; Gloria Isabel Escobar; Idabely Betancur Ortiz; Juan P. Hernandez-Ortiz; Maria Stella López |
| EPI_ISL_4520797 | Bienestar IPS | Universidad Nacional de Colombia - Laboratorio Genómico One Health | Andres F. Cardona-Rios; Carlos Franco-Muñoz; Carolina Muñoz-Arango; Celeny Ortiz; Daniel O. Maldonado-Perez; Diego A. Álvarez-Díaz; Hector Alejandro Ruiz-Moreno; Idabely Betancur Ortiz; Jorge E. Osorio; Juan P. Hernandez-Ortiz; Karl A Ciuoderis; Katherine Laiton-Donato; Laura Silvana Perez; Lina M. Hurtado; Marcela Mercado-Reyes; Maria Angélica Maya; Maria Stella López; Rita Almanza Payares; Sandra Ines Cano; Simón Villegas Velásquez |
| EPI_ISL_6572162 | BioMoLab | Molecular Genetics Laboratory, Instituto de Investigaciones Químicas, Universidad Mayor de San Andrés | Aneth Vasquez Michel; Carmen Delgado Barrera; Oscar M. Rollano-Peñaloza; Sandra Miranda Sardon |
| EPI_ISL_5152759, EPI_ISL_5152792 | CARVAJAL_LABORATORIO_IPS_SAS | UNAL-BOGOTA | Andrés Pinzón; Cristian Nicolás Rodríguez Pava; Emiliano Barreto Hernandez; Johana hernández Tolosa; Juan Germán Rodríguez Castillo; Lorena Alexandra Argoty Chamorro; Luis Fernando Cadavid; Marcela Castaño Rodríguez; Maria Andrea Angarita Rodriguez; Martha Isabel Murcia; María Irene Cerezo Cortes; Mishelle Cuello Mejia; Nicole Osorio Certuche |
| EPI_ISL_4509642, EPI_ISL_4510708, EPI_ISL_4511035, EPI_ISL_4511165, EPI_ISL_4680649, EPI_ISL_4959005, EPI_ISL_5015966, EPI_ISL_5016156, EPI_ISL_6175235, EPI_ISL_6175289 | | | |
| see above | CDPH VBL | California Department of Public Health | Emily Smith on behalf of CDPH-COVIDNet |
| EPI_ISL_6124117 | CHU Pontchaillou | CHU Pontchaillou | DE TAYRAC Marie; DENOUAL Florent; ETCHEVERRY Amandine; FEBREAU Christine; GALIBERT Marie Dominique; GROLHIER Claire; JAGLINE Steven; PRONIER Charlotte; QUENET Benjamin; SASSI Mohamed; THIBAULT Vincent |
| EPI_ISL_5071147, EPI_ISL_5071161, EPI_ISL_5071201, EPI_ISL_5072601 | COLCAN | Instituto Nacional de Salud-Dirección de Investigación en Salud Pública | Carlos Franco-Muñoz; Carmen Osorio; Diana Malo; Diego A. Álvarez-Díaz; Diego Andrés Prada; Gerardo Santamaría; Hector Alejandro Ruiz-Moreno; Jhonnatan Reales-González; Jorge Rivera; Juan Camilo Martinez; Julian Naizaque; Katherine Laiton-Donato; Lisseth Pardo; Magdalena Wiesner; Marcela Mercado-Reyes; Maria T. Herrera-Sepúlveda; Marta Lopez Blanco; Martha Lucia Ospina Martinez; Paola Rojas-Estevez; Sergio Gomez; Sheryll Corchuelo; Ángela Alarcon Cruz |
| EPI_ISL_5054472, EPI_ISL_5054477 | COLCAN - INS | Gencore - Universidad de los Andes | Cristian Barrera; David González; Gabriela Ariza; Luisa Sacristan; Marcela Guevara; Marcela Mercado; Silvia Restrepo |
| EPI_ISL_5071127 | CORPORACION CLINICA PRIMAVERA DE VILLAVICENCIO | Instituto Nacional de Salud-Dirección de Investigación en Salud Pública | Carlos Franco-Muñoz; Carmen Osorio; Diana Malo; Diego A. Álvarez-Díaz; Diego Andrés Prada; Gerardo Santamaría; Hector Alejandro Ruiz-Moreno; Jhonnatan Reales-González; Jorge Rivera; Juan Camilo Martinez; Julian Naizaque; Katherine Laiton-Donato; Lisseth Pardo; Magdalena Wiesner; Marcela Mercado-Reyes; Maria T. Herrera-Sepúlveda; Marta Lopez Blanco; Martha Lucia Ospina Martinez; Paola Rojas-Estevez; Sergio Gomez; Sheryll Corchuelo; Ángela Alarcon Cruz |
| EPI_ISL_5152809 | CPC-Synlab_Colombia | UNAL-BOGOTA | Andrés Pinzón; Cristian Nicolás Rodríguez Pava; Emiliano Barreto Hernandez; Johana hernández Tolosa; Juan Germán Rodríguez Castillo; Lorena Alexandra Argoty Chamorro; Luis Fernando Cadavid; Marcela Castaño Rodríguez; Maria Andrea Angarita Rodriguez; Martha Isabel Murcia; María Irene Cerezo Cortes; Mishelle Cuello Mejia; Nicole Osorio Certuche |
| EPI_ISL_5062464 | Centre Hospitalier Universitaire de Rouen Laboratoire de Virologie | Centre Hospitalier Universitaire de Rouen Laboratoire de Virologie | Alice Moisan; Fabienne De Oliveira; Marie Leoz |
| EPI_ISL_4560893 | Centro de Investigación Biomédica de La Rioja - Hospital San Pedro Logroño | SeqCOVID-SPAIN consortium/IBV(CSIC) | José Manuel Azcona Gutiérrez; María Pilar Bea Escudero; María de Toro; Miriam Blasco Alberdi and SeqCOVID-SPAIN consortium |
| EPI_ISL_5779882 | Clinique Saint-Pierre Ottignies | UCLouvain/IREC/MBLG | Benoit Kabamba Mukadi; Bertrand Bearzatto; Jean Ruelle |
| EPI_ISL_5603315, EPI_ISL_5603329 | Clínica Imbanaco | Laboratorio de Biología Molecular y Biotecnología - Universidad Tecnológica de Pereira | Augusto Zuluaga-Velez; Fredy A. Tabares-Villa; Juan C. Sepulveda; Juan D. Anacona-Montilla; Marcela Orjuela-Rodriguez |
| EPI_ISL_5264656 | Colorado Department of Public Health and Environment | Colorado Department of Public Health and Environment | Alexandria Rossheim; Diana Ir; Emily A. Travanty; Laura Bankers; Mandy Waters; Michael Martin; Molly C. Hetherington-Rauth; Sarah Elizabeth Totten; Shannon R. Matzinger |
| EPI_ISL_5603780 | Compensar Calle 26 | LSPSDS | Alejandro Gomez Lopez; Gabriela Delgado Murcia; Johana Hernandez Toloza; Marcela Castano Rodriguez; Marcela Mercado |
| EPI_ISL_5522795 | Curative Labs | Curative Labs | Elias L. Salfati; Eugenia Khorosheva; George Way; J.Cesar Ignacio-Espinoza; Janet Chen; Mikhail Hanewich-Hollatz; Nabjot Sandhu; Sophia Quasem; Vladimir Slepnev; Zhiyi Xie |
| EPI_ISL_4413569, EPI_ISL_4424795 | DASA | DASA | Adriano Bonaldi; Angelica Hristov; Annelise Lopes; Bianca Cota; Cristina Oliveira; Jose Levi; Lidia Yamamoto; Paulo Pierry; Rodrigo Guarischi; Rodrigo Salazar |
| EPI_ISL_4891956 | Diagnostyka. Laboratoria Medyczne. | 1. ViroGenetics - BSL3 Laboratory of Virology, Małopolska Centre of Biotechnology, Jagiellonian University; 2. genXone SA, Research & Development Laboratory | Aleksandra Gidlewicz; Anna Brylak; Gromowski, T.; Grzegorz Nowicki; Jakub Grabowski; Kowalski, M.; Labaj; Maciej Sykulski; Mazur-Panasiuk, N.; Michał Kaszuba; Natalia Drweska-Matelska; P.P.; Pyrc, K.; Ruslan Herasymenko; Sylwia Januszczak; Szulc, P.; Wydmanski, W.; Łukasz Krych |
| EPI_ISL_5645353, EPI_ISL_5645484, EPI_ISL_5645515, EPI_ISL_5645516, EPI_ISL_5645519 | Dirección Regional de Salud (DIRESA) Amazonas | Instituto de Enfermedades Tropicales - UNTRM | Alejandra Dávila Barclay; Carla Montenegro; Cecilia Pajuelo; Christian Campos; Diego Cuicapuza; Guillermo Salvatierra; Luis Rojas; Pablo Tsukayama; Pedro E. Romero; Rafael Tapia; Stella Chenet |
| EPI_ISL_4401998, EPI_ISL_4401999, EPI_ISL_4402000, EPI_ISL_4839317, | Dutch COVID-19 response team | National Institute for Public Health and the Environment (RIVM) | Adam Meijer; AnneMarie van den Brandt; Annelies Kroneman; Bas van der Veer; Chantal Reusken; Dennis Schmitz; Dirk Eggink; Euníce Then; Florian Zwagemaker; Harry Vennema; Ivo van Walle; Jeroen Cremer; Karim Hajji; Kim Freriks; Lisa Wijsman; Lynn Aarts; Ryanne Jaarsma; Sanne Bos; Sharon van den Brink; Stijn van Rossum; on behalf of the national COVID-19 response team |

| | | | |
|---|---|---|---|
| EPI_ISL_4846094 | | | |
| EPI_ISL_6512558 | ESCUELA DE MICROBIOLOGIA UNIVERSIDAD DE ANTIOQUIA | Laboratorio Departamental de Salud Publica de Antioquia | Ana Victoria Valencia Duarte; Cristian Arbey Velarde Hoyos; Gloria Isabel Escobar; Idabely Betancur Ortiz; Juan P. Hernandez-Ortiz; Juan Pablo Isaza Agudelo; Maria Stella López |
| EPI_ISL_5934856 | Fimlab Laboratoriot Oy Tampere | Expert Microbiology, National Institute for Health and Welfare | Carita Savolainen-Kopra; Erika Lindh; Haider al-Hello; Jani Halkilahti; Kirsi Liitsola; Niina Ikonen; Olli Vapalahti; Pekka Ellonen; Phuoc Truong; Päivi Laurila; Ravi Kant; Sari Hannula; Soile Blomqvist; Teemu Smura |
| EPI_ISL_5541948, EPI_ISL_5541982, EPI_ISL_5541997, EPI_ISL_5542170, EPI_ISL_5866225, EPI_ISL_5925008, EPI_ISL_5925291, EPI_ISL_5925292, EPI_ISL_5925297, EPI_ISL_6367519 | | | |
| see above | Florida Bureau of Public Health Laboratories | Florida Bureau of Public Health Laboratories | Jason Blanton; Namratha Tarigopula; Sarah Schmedes; Tiffany Splatt |
| EPI_ISL_4371757, EPI_ISL_4372077, EPI_ISL_5232587, EPI_ISL_5609058 | Fulgent Genetics | Centers for Disease Control and Prevention Division of Viral Diseases, Pathogen Discovery | Becky Tsai; Benafsh Sapra; Benjamin Rambo-Martin; Christopher Gulvick; Clinton Paden; Dakota Howard; Dhwani Batra; Doreen Ng; Duncan MacCannell; Erisa Sula; Harry Gao; James Xie; Jason Caravas; John Gao; Joseph Fierro; Kristine Lacek; Matthew Schmerer; Mickey Li; Peter Cook; Scott Sammons; Shatavia Morrison; Tymeckia Kendall; Victoria Caban Figueroa; Yan Meng; Yvette Unoarumhi |
| EPI_ISL_5736039, EPI_ISL_5797256, EPI_ISL_5797260, EPI_ISL_5797266 | Fundacion Valle del Lili | Laboratorio de biotecnología, Universidad Icesi | "María I. Gutiérrez López; Adrián Camilo Rodríguez Ararat; Diana M. Florez Giraldo; Marcela Mercado; Maria F. Villegas Torres; Paola A. Caicedo Burbano"; Programa Nacional de Caracterización Genómica de SARS-CoV-2; Sara González Henao |
| EPI_ISL_5365919 | Fundación Cardioinfantil | Centro de Investigaciones en Microbiología y Biotecnología-UR (CIMBIUR), Facultad de Ciencias Naturales, Universidad del Rosario, Bogotá, Colombia | Angie Ramírez; Juan David Ramírez; Luz H. Patiño; Marcela Mercado-Reyes; Marina Muñoz; Nathalia Ballesteros; Nicolas Niño; Sergio Castañeda |
| EPI_ISL_4566164, EPI_ISL_4566175, EPI_ISL_4566198, EPI_ISL_4740144, EPI_ISL_4740247, EPI_ISL_4740255, EPI_ISL_4740265, EPI_ISL_4891921 | | | |
| see above | Gencore - Universidad de los Andes | Gencore - Universidad de los Andes | Cristian Barrera; David González; Felipe Báez; Gabriela Ariza; Luisa Sacristan; Marcela Guevara; Marcela Mercado; Silvia Restrepo |
| EPI_ISL_4628790, EPI_ISL_4632314, EPI_ISL_4635740, EPI_ISL_4649141, EPI_ISL_4659826, EPI_ISL_5011668, EPI_ISL_5880326, EPI_ISL_6052412, EPI_ISL_6052417, EPI_ISL_6054318, EPI_ISL_6054328, EPI_ISL_6453513, EPI_ISL_6453958, EPI_ISL_6453978, EPI_ISL_6453989, EPI_ISL_6453993, EPI_ISL_6455293, EPI_ISL_6456070, EPI_ISL_6456092, EPI_ISL_6568026, EPI_ISL_6568041, EPI_ISL_6568043, EPI_ISL_6568047 | | | |
| see above | Genetica Molecular and Subdepartamento de Virologia ISP Chile | Instituto de Salud Publica de Chile | Andres Castillo; Barbara Parra; Constanza Campano; Gisselle Barra; Javier Tognarelli; Jorge Fernandez; Karen Orostica; Loredana Arata; Patricia Bustos; Rodrigo Fasce; Soledad Ulloa |
| EPI_ISL_5196317, EPI_ISL_5196318, EPI_ISL_5196319, EPI_ISL_5196320, EPI_ISL_5196321, EPI_ISL_5196322, EPI_ISL_5196323, EPI_ISL_5196324, EPI_ISL_5196325, EPI_ISL_5196326, EPI_ISL_5196327, EPI_ISL_5196328, EPI_ISL_5196329, EPI_ISL_5196330, EPI_ISL_5196331 | | | |
| see above | Gorgas Memorial Institute of Health Studies | Gorgas Memorial Institute of Health Studies | Castillo Jorge; Chen Maria; Franco Danilo; Gonzalez Claudia; Jessica Gondola; Leyda Abrego; Lopez-Verges Sandra; Marlenne Castillo; Martinez Alexander; Menacho Abdiel; Moreno Ambar; Moreno Brechla; Oris Chavarria; Ortiz Alma; Salazar Jacqueline |
| EPI_ISL_4744620 | Gorgas Memorial Laboratory of Health Studies | Gorgas Memorial Laboratory of Health Studies | Castillo Jorge; Chen Maria; Franco Danilo; Gonzalez Claudia; Jessica Gondola; Leyda Abrego; Lopez-Verges Sandra; Marlenne Castillo; Martinez Alexander; Menacho Abdiel; Moreno Ambar; Moreno Brechla; Oris Chavarria; Ortiz Alma; Salazar Jacqueline |
| EPI_ISL_5315111 | Gyncentrum | Gyncentrum | Adam Pudełko; Agnieszka Polak; Aleksandra Skubis-Sikora; Celina Kruszniewska-Rajs; Emilia Morawiec; Jolanta Bartosiewicz- Wąsik; Magdalena Samul; Maria Maklasińska-Majdanik; Paweł Czerwiński; Robert Wojtyczka; Tomasz Wąsik |
| EPI_ISL_5072527 | HOSPITAL DEPARTAMENTAL DE VILLAVICENCIO | Instituto Nacional de Salud-Dirección de Investigación en Salud Pública | Carlos Franco-Muñoz; Carmen Osorio; Diana Malo; Diego A. Álvarez-Díaz; Diego Andrés Prada; Gerardo Santamaría; Hector Alejandro Ruiz-Moreno; Jhonnatan Reales-González; Jorge Rivera; Juan Camilo Martinez; Julian Naizaque; Katherine Laiton-Donato; Lisseth Pardo; Magdalena Wiesner; Marcela Mercado-Reyes; Maria T. Herrera-Sepúlveda; Marta Lopez Blanco; Martha Lucia Ospina Martinez; Paola Rojas-Estevez; Sergio Gomez; Sheryll Corchuelo; Ángela Alarcon Cruz |
| EPI_ISL_4659468, EPI_ISL_6226805 | HOSPITAL UNIVERSITARIO 12 DE OCTUBRE | HOSPITAL UNIVERSITARIO 12 DE OCTUBRE | Carmen Martín-Higuera; Esther Viedma; Irene Múñoz-Gallego; M.ª Dolores Folgueira; Mar Aguilera; Noelia Moral; Rafael Delgado; Sagrario Zurita |
| EPI_ISL_5886781 | HOSPITAL UNIVERSITARIO CENTRAL DE ASTURIAS | Laboratorio de Virología HUCA | ; Alba L; Alvarez-Arguelles ME; Boga JA; Costales I; Coto E; González-Alba JM; Gómez de Oña J; Martín-Rodríguez G; Melón S; Perez-Martínez Z; Rojo S; Sandoval M |
| EPI_ISL_5263913 | HOSPITAL UNIVERSITARIO VIRGEN DE LA ARRIXACA | Instituto de Salud Carlos III | A. Monzón; F. Casas; I. Jiménez; I.MORENO PARRADO; LAURA; M. Sandonís; P. Zaballos; S. Cuesta; S. Iglesias-Caballero; S. Pozo; S. Varona; V. Camarero; Vázquez-Morón |
| EPI_ISL_4960508, EPI_ISL_4984735 | HPTU | Laboratorio Departamental de Salud Publica de Antioquia | Andres F. Cardona-Rios; Cristian Arbey Velarde Hoyos; Gloria Isabel Escobar; Idabely Betancur Ortiz; Juan P. Hernandez-Ortiz; Maria Stella López |
| EPI_ISL_5858260 | Hackensack Medical Center | New York Genome Center | Amy Baldwin; Andre Corvelo; Barry Kreiswirth; David Perlin; Dayna M. Oschwald; Jose Mediavilla; Kaelea Composto; Kar Chow; Liang Chen; Marcus Cunningham; Michael Zody; Samantha Fennessey; Tom Maniatis |
| EPI_ISL_5522122, EPI_ISL_5659018, EPI_ISL_5947562, EPI_ISL_6479524, EPI_ISL_6481761, EPI_ISL_6484453, EPI_ISL_6485137, EPI_ISL_6485232 | | | |
| see above | Helix | Centers for Disease Control and Prevention Division of Viral Diseases, Pathogen Discovery | Benjamin Rambo-Martin; Christopher Gulvick; Clinton Paden; Dakota Howard; Dhwani Batra; Duncan MacCannell; Erisa Sula; Helix CA; Jason Caravas; Kristine Lacek; Matthew Schmerer; Peter Cook; Scott Sammons; Shatavia Morrison; Tymeckia Kendall; Victoria Caban Figueroa; Yvette Unoarumhi |
| EPI_ISL_4962237 | Helix/Illumina | Centers for Disease Control and Prevention Division of Viral Diseases, Pathogen Discovery | Adrian Paskey; Alexandre Bolze; Ary Ascencio; Benjamin Rambo-Martin; Brad Sickler; Charlotte Rivera-Garcia; Christopher Gulvick; Chrstine Tran; Clinton Paden; Dakota Howard; Darlene Wagner; David Becker; Dhwani Batra; Duncan MacCannell; Efren Sandoval; Eileen De Feo; Elizabeth Cirulli; Eric Allen; Geraint Levan; James Lu; Jan Antico; Jason Caravas; Jason Nguyen; Jimmy Ramirez; Jingtao Liu; Kara Moser; Kelly Barrett; Kim Gietzen; Kristine Lacek; Magnus Isaksson; Marc Laurent; Matthew Schmerer; Matthew Tolentino; Nicole Washington; Peter Cook; Phil Febbo; Ryan Cho; Scott Sammons; Shannon Wickline; Shatavia Morrison; Sherry Wang; Simon White; Tyler Cassens; William Lee; Yvette Unoarumhi |
| EPI_ISL_4946245 | Hospital Civil Ipiales | Instituto Nacional de Salud-Dirección de Investigación en Salud Pública | Carlos Franco-Muñoz; Carmen Osorio; Diana Malo; Diego A. Álvarez-Díaz; Diego Andrés Prada; Gerardo Santamaría; Hector Alejandro Ruiz-Moreno; Jhonnatan Reales-González; Jorge Rivera; Juan Camilo Martinez; Julian Naizaque; Katherine Laiton-Donato; Lisseth Pardo; Magdalena Wiesner; Marcela Mercado-Reyes; Maria T. Herrera-Sepúlveda; Marta Lopez Blanco; Martha Lucia Ospina Martinez; Paola Rojas; Sergio Gomez; Sheryll Corchuelo; Ángela Alarcon Cruz |
| EPI_ISL_5587611, EPI_ISL_5587623 | Hospital General Plaza de la Salud | Instituto de Medicina Tropical & Salud Global, Universidad Iberoamericana (UNIBE) | A; Anel; Ann; Calderon; Cuevas; Cuevas Lantigua; Gabriella; Gilda; Guzman Marte; Jabier; Maridania; Paula; Paulino-Ramirez; Rita.; Robert; Rojas Fermin; Sanchez Marmolejos; Tolari Jacobo; Vallejo Degaudenzi; Victor Virgilio |
| EPI_ISL_5587633, EPI_ISL_5587645, EPI_ISL_5587651, EPI_ISL_5621195, EPI_ISL_5621494, EPI_ISL_5621499, EPI_ISL_5621502, EPI_ISL_5687696, EPI_ISL_5687945 | | | |
| see above | Hospital General Plaza de la Salud, Infectious Disease Department | Instituto de Medicina Tropical & Salud Global, Universidad Iberoamericana (UNIBE) | A; Anel; Ann; Calderon; Cuevas; Cuevas Lantigua; Gabriella; Gilda; Guzman Marte; Jabier; Maridania; Paula; Paulino-Ramirez; Rita.; Robert; Rojas Fermin; Sanchez Marmolejos; Tolari Jacobo; Vallejo Degaudenzi; Victor Virgilio |
| EPI_ISL_5587617 | Hospital General Plaza de la Salud, Infectious Disease Department | Instituto de Medicina Tropical & Salud Global, Universidad Iberoamericana (UNIBE), Santo Domingo, 22333, Dominican Republic | A; Anel; Ann; Calderon; Cuevas; Cuevas Lantigua; Gabriella; Gilda; Guzman Marte; Jabier; Maridania; Paula; Paulino-Ramirez; Rita.; Robert; Rojas Fermin; Sanchez Marmolejos; Tolari Jacobo; Vallejo Degaudenzi; Victor Virgilio |
| EPI_ISL_4987001, EPI_ISL_4987029, EPI_ISL_6135444, EPI_ISL_6135499, EPI_ISL_6135500, EPI_ISL_6135501, EPI_ISL_6157320 | | | |
| see above | Hospital General Universitario Gregorio Marañón | Hospital General Universitario Gregorio Marañón | Cristina Rodriguez-Grande; Darío García de Viedma; Jorge Rodríguez-Grande; Julia Suárez; Laura Pérez-Lago; Marta Herranz Martin; Patricia Muñoz; Pedro Sola Campoy; Pilar Catalán; Sergio Buenestado Serrano; Victor Manuel de la Cueva |
| EPI_ISL_4560905, EPI_ISL_4560920, EPI_ISL_4560973, EPI_ISL_4560982 | Hospital General Universitario de Alicante - Instituto de Investigación Sanitaria y Biomédica de Alicante | SeqCOVID-SPAIN consortium/IBV(CSIC) | Carmen Molina Pardines and SeqCOVID-SPAIN consortium; Maripaz Ventero Martín |
| EPI_ISL_5926925 | Hospital Herrera Llerandi | Asociación de Salud Integral / Clínica Familiar "Luis Ángel García" | Ana S. Gonzalez-Reiche; Claudia Rangel; Danicela Mercado; Eduardo Arathon; Hilda Ruiz; Luis Aguirre; Luis Rivas; Narda Medina; Oscar Bonilla; Osmar Gamboa. |
| EPI_ISL_4885353, EPI_ISL_4885355, EPI_ISL_4885356, EPI_ISL_6026625 | Hospital Universitari Joan XXIII | Hospital Universitari Joan XXIII | Carla Martín; Clara Benavent; Cristina Gutiérrez; Ester Picó; Gemma Recio; Margarida Terrón |
| EPI_ISL_5340043, EPI_ISL_5340961, EPI_ISL_5341127, EPI_ISL_5341826, EPI_ISL_5344149, EPI_ISL_5375470 | Houston Methodist Hospital | Houston Methodist Hospital | Ilya J. Finkelstein; James J. Davis; Jessica Cambric; Jimmy Gollihar; Kristina Reppond; Layne Pruitt; Madison N. Shyer; Marcus Nguyen; Matthew Ojeda Saavedra; Paul A. Christensen; Prasanti Yerramilli; Randall J. Olsen; Robert Olson; Ryan Gadd; S. Wesley Long; Sishir Subedi; and James M. Musser |
| EPI_ISL_4984530 | ICMT APARTADO | Laboratorio Departamental de Salud Publica de Antioquia | Andres F. Cardona-Rios; Cristian Arbey Velarde Hoyos; Gloria Isabel Escobar; Idabely Betancur Ortiz; Juan P. Hernandez-Ortiz; Maria Stella López |
| EPI_ISL_5778137 | INCMNSZ | Instituto Nacional de Medicina Genomica | Cedro-Tanda A; Cruz-Islas J; Escobar-Arrazola MA; Herrera-Montalvo LA.; Hidalgo-Miranda A; Mendoza-Vargas A; Ramirez-Vega O; Rangel-DeLeon D; Reyes-Grajeda JP; Yair Alfaro-Mora |
| EPI_ISL_5650473 | INSIDE DIAGNOSTICOS | Instituto Butantan | Antonio Jorge Martins; Claudia Renata dos Santos Barros; David Schlesinger; Debora Botequio Moretti; Dimas Tadeu Covas; Elaine Cristina Marqueze; Elaine Vieira Santos; Evandra Strazza Rodrigues; Heidge Fukumasu; Jayme Augusto de Souza-Neto; José Salvatore Leister Patané; Luiz Alcantara; Luiz Lehmann |

Coutinho; Maria Carolina Elias; Maurício Lacerda Nogueira; Rafael dos Santos Bezerra; Raul Machado Neto; Rejane Maria Tommasini Grotto; Ricardo Haddad; Sandra Coccuzzo Sampaio Vessoni; Simone Kashima; Svetoslav Nanev Slavov; Vincent Louis Viala

EPI_ISL_6129002, EPI_ISL_6129027, EPI_ISL_6208167, EPI_ISL_6208176, EPI_ISL_6208197, EPI_ISL_6208214, EPI_ISL_6208237, EPI_ISL_6208247, EPI_ISL_6208263, EPI_ISL_6208293, EPI_ISL_6208306, EPI_ISL_6208311, EPI_ISL_6208319, EPI_ISL_6208320, EPI_ISL_6507608, EPI_ISL_6507610, EPI_ISL_6507611, EPI_ISL_6507612, EPI_ISL_6507613, EPI_ISL_6507614, EPI_ISL_6507691, EPI_ISL_6526278, EPI_ISL_6526284, EPI_ISL_6526285, EPI_ISL_6569586, EPI_ISL_6569593, EPI_ISL_6569599, EPI_ISL_6569609, EPI_ISL_6569625, EPI_ISL_6569673

| | | | |
|---|---|---|---|
| see above | INSPI-CRN DE INFLUENZA Y OTROS VIRUS RESPIRATORIOS | INSPI-CRN DE INFLUENZA Y OTROS VIRUS RESPIRATORIOS | Alfredo Bruno; Carlos Chiluiza; Daniel Ramos; Domenica de Mora; Fernando Llerena; Jimmy Garcés; Lizbeth Patiño; Marcela Mejia; Maria Angelica Becerra; Maritza Olmedo; Michelle Páez; Ruben Armas |
| EPI_ISL_4497025 | INSPI-CRN DE INFLUENZA Y OTROS VIRUS RESPIRATORIOS | NIC-INSPI | Alfredo Bruno; Domenica de Mora.; Jimmy Garcés; Johanna Laines; Lizbeth Patiño; Manuel Gonzalez; Maritza Olmedo; Michelle Páez |
| EPI_ISL_4382923, EPI_ISL_4384702, EPI_ISL_4386123, EPI_ISL_4387287, EPI_ISL_6102799 | Infinity Biologix | Centers for Disease Control and Prevention Division of Viral Diseases, Pathogen Discovery | Benjamin Rambo-Martin; Chirayu Goswami; Christian Bixby; Christopher Gulvick; Clinton Paden; Dakota Howard; Dhwani Batra; Duncan MacCannell; Erisa Sula; Jason Caravas; Jonathan Schultz; Kristine Lacek; Matthew Schmerer; Peter Cook; Robin Grimwood; Russ Hager; Scott Sammons; Shatavia Morrison; Tymeckia Kendall; Victoria Caban Figueroa; Yihe Wang; Yvette Unoarumhi |
| EPI_ISL_4740197, EPI_ISL_5094648, EPI_ISL_6368039 | Institute of Microbiology, Universidad San Francisco de Quito | Institute of Microbiology, Universidad San Francisco de Quito | Belén Prado-Vivar; Bernardo Gutiérrez; Daniela Villalva; Erika B. Muñoz; Fernanda Zurita; Gabriel Trueba; Hugo Navarrete; Jaime Costales; Juan José Guadalupe; Mateo Carvajal; Michelle Grunauer; Monica Becerra-Wong; Nelson Montalvan; Patricio Rojas-Silva; Paúl Cárdenas; Sully Márquez; Verónica Barragán |
| EPI_ISL_5778127 | Instituto Nacional de Medicina Genomica | Instituto Nacional de Medicina Genomica | Cedro-Tanda A; Cruz-Islas J; Escobar-Arrazola MA; Herrera-Montalvo LA.; Hidalgo-Miranda A; Mendoza-Vargas A; Ramirez-Vega O; Rangel-DeLeon D; Reyes-Grajeda JP; Yair Alfaro-Mora |
| EPI_ISL_5393528, EPI_ISL_5424306 | KU Leuven, Rega Institute, Clinical and Epidemiological Virology | KU Leuven, Rega Institute, Clinical and Epidemiological Virology | Bert Vanmechelen; Joan Marti-Carerras; Piet Maes; Tony Wawina-Bokalanga |
| EPI_ISL_5421700 | Kaiser Permanente NW Reginal Lab | OHSU MM Lab | Amber Halse; Jeannine Lama; Xuan Qin; Yun Wu |
| EPI_ISL_5071132, EPI_ISL_5071190, EPI_ISL_5072505, EPI_ISL_5072515, EPI_ISL_5072592 | LABORATORIO ALIFE HEALTH | Instituto Nacional de Salud-Dirección de Investigación en Salud Pública | Carlos Franco-Muñoz; Carmen Osorio; Diana Malo; Diego A. Álvarez-Díaz; Diego Andrés Prada; Gerardo Santamaría; Hector Alejandro Ruiz-Moreno; Jhonnatan Reales-González; Jorge Rivera; Juan Camilo Martinez; Julian Naizaque; Katherine Laiton-Donato; Lisseth Pardo; Magdalena Wiesner; Marcela Mercado-Reyes; Maria T. Herrera-Sepúlveda; Marta Lopez Blanco; Martha Lucia Ospina Martinez; Paola Rojas-Estevez; Sergio Gomez; Sheryll Corchuelo; Ángela Alarcon Cruz |
| EPI_ISL_5055255, EPI_ISL_5055261, EPI_ISL_5055262, EPI_ISL_5055269, EPI_ISL_5055281, EPI_ISL_5055300 | LABORATORIO CLINICO COMPENSAR | Instituto Nacional de Salud-Dirección de Investigación en Salud Pública | Beatriz de Arco; Carlos Franco-Muñoz; Carmen Osorio; Diana Malo; Diego A. Álvarez-Díaz; Diego Andrés Prada; Gerardo Santamaría; Hector Alejandro Ruiz-Moreno; Jhonnatan Reales-González; Jorge Rivera; Juan Camilo Martinez; Julian Naizaque; Katherine Laiton-Donato; Lisseth Pardo; Magdalena Wiesner; Marcela Mercado-Reyes; Maria T. Herrera-Sepúlveda; Marta Lopez Blanco; Martha Lucia Ospina Martinez; Paola Rojas; Sergio Gomez; Sheryll Corchuelo; Tatiana Cobos; Ángela Alarcon Cruz |
| EPI_ISL_4960511, EPI_ISL_4960512 | LABORATORIO CLINICO HEMATOLOGICO | Laboratorio Departamental de Salud Publica de Antioquia | Cristian Arbey Velarde Hoyos; Gloria Isabel Escobar; Idabely Betancur Ortiz; Juan P. Hernandez-Ortiz; Maria Stella López |
| EPI_ISL_5334482 | LABORATORIO CLINICO HIGUERA ESCALANTE | Instituto Nacional de Salud-Dirección de Investigación en Salud Pública | Beatriz de Arco; Carlos Franco-Muñoz; Diego A. Álvarez-Díaz; Diego Andrés Prada; Dioselina Peláez-Carvajal; Gerardo Santamaría; Héctor Alejandro Ruiz-Moreno; Jhonnatan Reales-González; Jorge Rivera; Julián Naizaque; Katherine Laiton-Donato; Marcela Mercado-Reyes.; Martha Lucia Ospina Martínez; María T. Herrera-Sepúlveda; Paola Rojas; Sheryll Corchuelo; Tatiana Cobos |
| EPI_ISL_4984816, EPI_ISL_5428461 | LABORATORIO ECHAVARRIA | Laboratorio Departamental de Salud Publica de Antioquia | Ana Victoria Valencia Duarte; Andres F. Cardona-Rios; Cristian Arbey Velarde Hoyos; Gloria Isabel Escobar; Idabely Betancur Ortiz; Juan P. Hernandez-Ortiz; Juan Pablo Isaza Agudelo; Maria Stella López |
| EPI_ISL_5055131, EPI_ISL_5055132, EPI_ISL_5055141, EPI_ISL_5055174, EPI_ISL_5055216 | LABORATORIO IMAT | Instituto Nacional de Salud-Dirección de Investigación en Salud Pública | Beatriz de Arco; Carlos Franco-Muñoz; Carmen Osorio; Diana Malo; Diego A. Álvarez-Díaz; Diego Andrés Prada; Gerardo Santamaría; Hector Alejandro Ruiz-Moreno; Jhonnatan Reales-González; Jorge Rivera; Juan Camilo Martinez; Julian Naizaque; Katherine Laiton-Donato; Lisseth Pardo; Magdalena Wiesner; Marcela Mercado-Reyes; Maria T. Herrera-Sepúlveda; Marta Lopez Blanco; Martha Lucia Ospina Martinez; Paola Rojas; Sergio Gomez; Sheryll Corchuelo; Tatiana Cobos; Ángela Alarcon Cruz |
| EPI_ISL_5055268 | LABORATORIO LORENA VEJARANO S.A.S. | Instituto Nacional de Salud-Dirección de Investigación en Salud Pública | Beatriz de Arco; Carlos Franco-Muñoz; Carmen Osorio; Diana Malo; Diego A. Álvarez-Díaz; Diego Andrés Prada; Gerardo Santamaría; Hector Alejandro Ruiz-Moreno; Jhonnatan Reales-González; Jorge Rivera; Juan Camilo Martinez; Julian Naizaque; Katherine Laiton-Donato; Lisseth Pardo; Magdalena Wiesner; Marcela Mercado-Reyes; Maria T. Herrera-Sepúlveda; Marta Lopez Blanco; Martha Lucia Ospina Martinez; Paola Rojas; Sergio Gomez; Sheryll Corchuelo; Tatiana Cobos; Ángela Alarcon Cruz |
| EPI_ISL_4965717, EPI_ISL_4965728, EPI_ISL_4965734, EPI_ISL_4965742 | LDSP | Laboratorio Departamental de Salud Publica de Antioquia | Andres F. Cardona-Rios; Cristian Arbey Velarde Hoyos; Gloria Isabel Escobar; Idabely Betancur Ortiz; Juan P. Hernandez-Ortiz; Maria Stella López |
| EPI_ISL_5055250 | LDSP DEL CESAR | Instituto Nacional de Salud-Dirección de Investigación en Salud Pública | Beatriz de Arco; Carlos Franco-Muñoz; Carmen Osorio; Diana Malo; Diego A. Álvarez-Díaz; Diego Andrés Prada; Gerardo Santamaría; Hector Alejandro Ruiz-Moreno; Jhonnatan Reales-González; Jorge Rivera; Juan Camilo Martinez; Julian Naizaque; Katherine Laiton-Donato; Lisseth Pardo; Magdalena Wiesner; Marcela Mercado-Reyes; Maria T. Herrera-Sepúlveda; Marta Lopez Blanco; Martha Lucia Ospina Martinez; Paola Rojas; Sergio Gomez; Sheryll Corchuelo; Tatiana Cobos; Ángela Alarcon Cruz |
| EPI_ISL_4984610 | LDSP de Antioquia | Laboratorio Departamental de Salud Publica de Antioquia | Andres F. Cardona-Rios; Cristian Arbey Velarde Hoyos; Gloria Isabel Escobar; Idabely Betancur Ortiz; Juan P. Hernandez-Ortiz; Maria Stella López |
| EPI_ISL_4917852 | LESP Tabasco | Instituto de Diagnostico y Referencia Epidemiologicos (INDRE) | Abril Rodriguez-Maldonado; Ariadna Medina-Benitez; Claudia Wong-Arambula; Ernesto Ramirez-Gonzalez.; Fernando Gonzalez-Dominguez; Gisela Barrera-Badillo; Irma Lopez-Martinez; Joaquin Quiroz-Mercado; Lucia Hernandez-Rivas; Maribel Gonzalez-Villa; Natividad Cruz-Ortiz; Sergio Rangel-Guerrero; Tatiana Nunez-Garcia; Vanessa Rivero-Arredondo |
| EPI_ISL_4984862, EPI_ISL_4984875, EPI_ISL_5923355 | LIME | Laboratorio Departamental de Salud Publica de Antioquia | Ana Victoria Valencia Duarte; Andres F. Cardona-Rios; Cristian Arbey Velarde Hoyos; Gloria Isabel Escobar; Idabely Betancur Ortiz; Juan P. Hernandez-Ortiz; Juan Pablo Isaza Agudelo; Maria Stella López |
| EPI_ISL_5020128 | LSP Valle del Cauca | Molecular Genetics and Antimicrobial Resistance - UGRA, Universidad El Bosque | Catalina Espitia; Jinnethe Reyes; Lorena Diaz; Marcela Mercado; Mauricio Pacheco; Rafael Rios; Valentina Martinez |
| EPI_ISL_4417574, EPI_ISL_4417588, EPI_ISL_4417653 | LSP de Atlantico | Centro de Investigaciones en Microbiología y Biotecnología-UR (CIMBIUR), Facultad de Ciencias Naturales, Universidad del Rosario, Bogotá, Colombia | Angie Ramírez; Juan David Ramírez; Luz H. Patiño; Marcela Mercado-Reyes; Marina Muñoz; Nathalia Ballesteros; Nicolas Niño; Sergio Castañeda |
| EPI_ISL_4419177 | Lab Christus Sinergia Salud - Clinica Farallones | Universidad del Valle | Programa Nacional de Caracterización Genómica de SARS-CoV-2 |
| EPI_ISL_5887479 | Lab. Hospital clinico Universidad de Chile | "Facultad de Ciencias de la Vida, UNAB" | "Claudio Meneses; Ariel Orellana"; Claudio Olmos; Daniel Leon; Dayan Sanhueza; Eduardo Castro; Gonzalo Campaña; Macarena Bastias; Paola Pidal; Ricardo Yusta; Sebastian Wolter; Susana Saez; Víctor Monreal; Waldo Diaz |
| EPI_ISL_4412226, EPI_ISL_4412229, EPI_ISL_4412231 | Labo Analyses Med | National Reference Center for Viruses of Respiratory Infections, Institut Pasteur, Paris | Angela Brisebarre; Camille Capel; Christophe Malabat; Corinne Chauvet; Corinne Maufrais; Etienne Simon-Lorière; Frédéric Lemoine; Julien Fumey; Louise Lefrançois; Marion Barbet; Maud Vanpeene; Méline Bizard; Slim El Khiari; Sylvie Behillil; Sylvie Van der Werf; Vincent Enouf |
| EPI_ISL_5422240, EPI_ISL_5897949, EPI_ISL_5897950, EPI_ISL_5897953, EPI_ISL_5898098 | Laboratoire de santé publique du Québec | Laboratoire de santé publique du Québec | Guillaume Bourque; Ioannis Ragoussis; Jesse Shapiro; Mark Lathrop and Michel Roger on behalf of the CoVSeQ research group; Sandrine Moreira |
| EPI_ISL_5395466 | Laboratoires Reunis | Laboratoire national de sante, Microbiology, Microbial Genomics Platform | Anke Wienecke-Baldacchino; Bernard Weber; Catherine Ragimbeau; Elodie Solarino; Fatu Djabi; Jessica Tapp; Lise Pignon; Raoul Salmon; Tamir Abdelrahman; Virginie Jover |
| EPI_ISL_4634221 | Laboratori de Referencia de Catalunya | Laboratori de Referencia de Catalunya | Bellosillo B.; Canal M.; Hernandez JJ.; Padilla E.; Ramirez A.; Vilas A. |
| EPI_ISL_4946220, EPI_ISL_4946239, EPI_ISL_4946241 | Laboratorio Angel ( Synlab) | Instituto Nacional de Salud-Dirección de Investigación en Salud Pública | Carlos Franco-Muñoz; Carmen Osorio; Diana Malo; Diego A. Álvarez-Díaz; Diego Andrés Prada; Gerardo Santamaría; Hector Alejandro Ruiz-Moreno; Jhonnatan Reales-González; Jorge Rivera; Juan Camilo Martinez; Julian Naizaque; Katherine Laiton-Donato; Lisseth Pardo; Magdalena Wiesner; Marcela Mercado-Reyes; Maria T. Herrera-Sepúlveda; Marta Lopez Blanco; Martha Lucia Ospina Martinez; Paola Rojas; Sergio Gomez; Sheryll Corchuelo; Ángela Alarcon Cruz |
| EPI_ISL_6062547, EPI_ISL_6155758, EPI_ISL_6155762, EPI_ISL_6155769, EPI_ISL_6155775 | Laboratorio Central, Ministerio de Salud Córdoba | Instituto de Patologia Vegetal (CIAP-INTA) on behalf of 'Proyecto Argentino Interinstitucional de genomica de SARS-CoV-2' (PAIS Consortium) | A; Amadio; Barbas, G.; Castro, G.; Debat; Debat, HJ.; FD; Fernández; Irazoqui; M; M.B.; Marquez, N.; Pisano; Re, V. |

EPI_ISL_4417560, EPI_ISL_4417604, EPI_ISL_4417606, EPI_ISL_4417608, EPI_ISL_4417609, EPI_ISL_4417610, EPI_ISL_4417619, EPI_ISL_4417628, EPI_ISL_4417642, EPI_ISL_4417645, EPI_ISL_4417648

| | | | |
|---|---|---|---|
| see above | Laboratorio Clinica Iberoamericana (Sanitas) | Centro de Investigaciones en Microbiología y Biotecnología-UR (CIMBIUR), Facultad de Ciencias Naturales, Universidad del Rosario, Bogotá, Colombia | Angie Ramírez; Juan David Ramírez; Luz H. Patiño; Marcela Mercado-Reyes; Marina Muñoz; Nathalia Ballesteros; Nicolas Niño; Sergio Castañeda |
| EPI_ISL_4703869, EPI_ISL_5395975 | Laboratorio Clinico CITISALUD | Laboratorio de Biologia Molecular, Universidad Cooperativa de Colombia, Santa Marta | Andrew S. Muñoz-Gamba; Camila Gonzalez; Daniel B. Ramírez-Osorio; Johana Zúñiga; José A. Usme-Ciro; Maria J. Capmartin; Paula A. Quintero-Cortés; Robinson Alvarez |
| EPI_ISL_5103737 | Laboratorio Clínico BioReferencia | Corporación para Investigaciones Biológicas-CIB | Jeanneth Mosquera Rendon; Jenny Santiago Cuesta; Marcela Mercado Reyes; Uriel A. Hurtado Paez |
| EPI_ISL_4878266, EPI_ISL_4878270, EPI_ISL_4878303 | Laboratorio Direccion Sanidad Policia Nacional | Instituto Nacional de Salud-Dirección de Investigación en Salud Pública | Carlos Franco-Muñoz; Carmen Osorio; Diana Malo; Diego A. Álvarez-Díaz; Diego Andrés Prada; Gerardo Santamaría; Hector Alejandro Ruiz-Moreno; Jhonnatan Reales-González; Jorge Rivera; Juan Camilo Martinez; Julian Naizaque; Katherine Laiton-Donato; Lisseth Pardo; Magdalena Wiesner; Marcela Mercado-Reyes; Maria T. Herrera-Sepúlveda; Marta Lopez Blanco; Martha Lucia Ospina Martinez; Paola Rojas; Sergio Gomez; Sheryll Corchuelo; Ángela Alarcon Cruz |
| EPI_ISL_5063050, EPI_ISL_5063065 | Laboratorio Direccion Sanidad Policia Nacional | Centro de Investigaciones en Microbiología y Biotecnología-UR (CIMBIUR), Facultad de Ciencias Naturales, Universidad del Rosario, Bogotá, Colombia | Angie Ramírez; Juan David Ramírez; Luz H. Patiño; Marcela Mercado-Reyes; Marina Muñoz; Nathalia Ballesteros; Nicolas Niño; Sergio Castañeda |
| EPI_ISL_4703599 | Laboratorio Las Americas | Laboratorio de Biologia Molecular, Universidad Cooperativa de Colombia, Santa Marta | Adriana Espitia; Andrew S. Muñoz-Gamba; Daniel B. Ramírez-Osorio; Javier Martinez; José A. Usme-Ciro; Paula A. Quintero-Cortés |
| EPI_ISL_5103733, EPI_ISL_5103746, EPI_ISL_5103747, EPI_ISL_5103765, EPI_ISL_5103773, EPI_ISL_5103784, EPI_ISL_5103787, EPI_ISL_5103823, EPI_ISL_5103846 | | | |
| see above | Laboratorio SYNLAB Colombia | Corporación para Investigaciones Biológicas-CIB | Jeanneth Mosquera Rendon; Jenny Santiago Cuesta; Marcela Mercado Reyes; Uriel A. Hurtado Paez |
| EPI_ISL_5199330, EPI_ISL_5199349 | Laboratorio SYNLAB Colombia | Instituto Nacional de Salud-Dirección de Investigación en Salud Pública | Beatriz de Arco; Carlos Franco-Muñoz; Carmen Osorio; Diana Malo; Diego A. Álvarez-Díaz; Diego Andrés Prada; Gerardo Santamaría; Hector Alejandro Ruiz-Moreno; Jhonnatan Reales-González; Jorge Rivera; Juan Camilo Martinez; Julian Naizaque; Katherine Laiton-Donato; Lisseth Pardo; Magdalena Wiesner; Marcela Mercado-Reyes; Maria T. Herrera-Sepúlveda; Marta Lopez Blanco; Martha Lucia Ospina Martinez; Paola Rojas; Sergio Gomez; Sheryll Corchuelo; Tatiana Cobos; Ángela Alarcon Cruz |
| EPI_ISL_5020133 | Laboratorio clínico Angeles Biología molecular | Molecular Genetics and Antimicrobial Resistance - UGRA, Universidad El Bosque | Catalina Espitia; Jinnethe Reyes; Lorena Diaz; Marcela Mercado; Mauricio Pacheco; Rafael Rios; Valentina Martinez |
| EPI_ISL_4703643, EPI_ISL_4703763, EPI_ISL_4703808 | Laboratorio de Biologia Molecular, Universidad Cooperativa de Colombia, Santa Marta | Laboratorio de Biologia Molecular, Universidad Cooperativa de Colombia, Santa Marta | Andres Rojas-Gulloso; Andrew S. Muñoz-Gamba; Beatriz Maestre; Camila Gonzalez; Daniel B. Ramírez-Osorio; Emy S. Torres; Johanna Zúñiga; José A. Usme-Ciro; Paula A. Quintero-Cortés |
| EPI_ISL_5065711, EPI_ISL_5065721, EPI_ISL_5065730 | Laboratorio de Biología Molecular - Universidad del Magdalena | Centro de Genética y Biología Molecular - Universidad del Magdalena | Andrea M. Ramírez Hernandez; Angel Oviedo Marquez; Daniel Bautista; Edison Lea-Ch; Lyda R. Castro; Maria Teresa Mojica-Ortiz |
| EPI_ISL_4578878, EPI_ISL_4578886 | Laboratorio de Biología molecular del Hospital General de Agudos Dr. Carlos G. Durand | Área de Secuenciación del Laboratorio de Virología del Hospital de Niños Dr. Ricardo Gutierrez on behalf of 'Proyecto Argentino Interinstitucional de genomica de SARS-CoV-2' (PAIS Consortium) | A; Acuña; Anzorena; B; C; Carrón; Castro; Cañellas; Chiussi; Colina; Costa; D; Dahinten; Dima; Domínguez; E; Elsegood; F; Frisone; G; Gatica; Goya; Irrazábal; Jurado; L; LE; Leivas; Loayza; Lusso; M; MB; MF; MI; MS; Marchissio; Marina; Matillas; N; Nabaes Jodar; Narduzzi; Natale; Notaristéfano; R; Rivarola; Rodríguez Cardozo; Rodríguez Saá; Rozo; S; Theaux; Valinotto; Viegas, M.; W; Warszatska; Y; Yaunguzian |
| EPI_ISL_4890344, EPI_ISL_4890345, EPI_ISL_4890346, EPI_ISL_4890359 | Laboratorio de Biología molecular y Biotecnología UTP | Laboratorio de Biología Molecular y Biotecnología - Universidad Tecnológica de Pereira | Augusto Zuluaga-Velez; Fredy A. Tabares-Villa; Juan C. Sepulveda; Juan D. Anacona-Montilla; Marcela Orjuela-Rodriguez |
| EPI_ISL_5147062 | Laboratorio de Diagnóstico e Investigación BIOSCIENCE SRL | Laboratorio de Genómica Microbiana, Universidad Peruana Cayetano Heredia | Alejandra Dávila-Barclay; Diego Cuicapuza; Esdenka Pérez; Freddy Tinajeros; Guillermo Salvatierra; Janet Huancachoque; Luis González; Mauricio Prado; Pablo Tsukayama; Paula Carballo; Pedro E. Romero; Pool Marcos |
| EPI_ISL_5490873, EPI_ISL_5490878, EPI_ISL_5490886 | Laboratorio de Ecologia de Doencas Transmissiveis na Amazonia, Instituto Leonidas e Maria Deane - Fiocruz Amazonia | Laboratorio de Ecologia de Doencas Transmissiveis na Amazonia, Instituto Leonidas e Maria Deane - Fiocruz Amazonia | Adele Schwartz Benzaken; André Corado; Felipe Naveca; Fernanda Nascimento; George Silva; José Joaquín Carvajal Cortés; Juan Camilo Grisales Nieto; Kelly Natalia Romero Vesga; Luciana Gonçalves; Maria Júlia Brandão; Matilde Mejía; Valdinete Nascimento; Victor Souza |
| EPI_ISL_6208403, EPI_ISL_6208578, EPI_ISL_6208579, EPI_ISL_6208610, EPI_ISL_6208698, EPI_ISL_6574128 | Laboratorio de Referencia Nacional de Virus Respiratorios. Centro Nacional de Salud Publica. Instituto Nacional de Salud Peru. | Laboratorio de Referencia Nacional de Virus Respiratorios. Centro Nacional de Salud Publica. Instituto Nacional de Salud Peru. | Alicia Nuñez Llanos; Carlos Padilla Rojas; Edwart Rogger Rivera Serrano; Henri Bailon Calderon; Iris Silva Molina; Joseph Huayra Niquen; Kelly Vanessa Izarra Rojas; Lely Solari Zerpa; Luis Barcena Flores; Marco Galarza Perez; Nancy Rojas Serrano; Nieves Sevilla Castañeda; Omar Caceres Rey; Orson Mestanza Millones; Princesa Medrano Alhuay; Priscila Lope Pari; Sara Gordillo Vilchez; Steve Acedo Lazo; Veronica Hurtado Vela; Victor Jimenez Vasquez; Wendy Lizarraga Olivares |
| EPI_ISL_4578879, EPI_ISL_4578882 | Laboratorio de Virología del Hospital de Niños Dr. Ricardo Gutierrez | Área de Secuenciación del Laboratorio de Virología del Hospital de Niños Dr. Ricardo Gutierrez on behalf of 'Proyecto Argentino Interinstitucional de genomica de SARS-CoV-2' (PAIS Consortium) | AS; Acevedo; Acuña; Alexay; Alvarez Lopez; C; D; E; Goya; Gravis; Jacques; LE; Lusso; ME; MI; MS; Mistchenko; Nabaes Jodar; Natale; O; S; Valinotto; Viegas, M. |
| EPI_ISL_6368859 | Laboratorio nacional de salud publica | Viral Special Pathogens Branch, Centers for Disease Control and Prevention | Emir Talundzic; Isaac Miguel; Joel Montgomery; John Klena; Justin Lee; Melissa Mobley; Ronald Skewes-Ramm; Shannon Whitmer |
| EPI_ISL_4372847, EPI_ISL_4373371, EPI_ISL_4373672, EPI_ISL_4375408, EPI_ISL_4375456, EPI_ISL_4376827, EPI_ISL_4377577, EPI_ISL_4379164, EPI_ISL_4381338, EPI_ISL_4381935, EPI_ISL_4382021, EPI_ISL_4382170, EPI_ISL_4484580, EPI_ISL_4488844, EPI_ISL_4488845, EPI_ISL_4811830, EPI_ISL_4814264, EPI_ISL_4963077, EPI_ISL_4965554, EPI_ISL_4966220, EPI_ISL_5214209, EPI_ISL_5222609, EPI_ISL_5223528, EPI_ISL_5223668, EPI_ISL_5223669, EPI_ISL_5223812, EPI_ISL_5223907, EPI_ISL_5562250, EPI_ISL_5573377, EPI_ISL_6079093 | | | |
| see above | Laboratory Corporation of America | Centers for Disease Control and Prevention Division of Viral Diseases, Pathogen Discovery | Amanda Douglas; Amanda Suchanek; Andrea Throop; Ayla Burns; Benjamin Rambo-Martin; Bobbi Croy; Brian Krueger; Brian Norvell; Christopher Gulvick; Christos Petropoulos; Clinton Paden; Craig Lukasik; Dakota Howard; Debbie Boles; Dhwani Batra; Duncan MacCannell; Eyad Almasri; Goran Stevovic; Howard Engler; Hrushikesh Deshmukh; Jake Humphrey; Jana Schroth; Jason Caravas; Joe Voshell; John Pruitt; Jonathan Meltzer; Jonathan Williams; Kimberly Wagner; Kristine Lacek; Lax Iyer; Lisa Pfefferle; Lyndon Tilson; Manoj Jain; Marcia Eisenberg; Mary Cristobal; Mary Williamson; Matthew Robinson; Matthew Schmerer; Michael Levandoski; Mike Sapeta; Mindy Nye; Minoo Agarwal; Mohan Kolli; Nuthawin Charoensri; Oren Cohen; Peter Cook; Prashant Gupta; Qian Zeng; Rama Ghatti; Scott Parker; Scott Ryan; Scott Sammons; Shatavia Morrison; Stanley Letovsky; Steven Ragan; Suresh Selvaraju; Susan Countryman; Susan Hicks; Suzanne Dale; Thomas Urban; Tim Kuphal; Tricia Zwiefelhofer; Tymeckia Kendall; Victoria Caban Figueroa; Vincent Drouillon; Yvette Unoarumhi |
| EPI_ISL_4516039 | Laboratório Central de Saúde Pública do Amazonas - LACEN-AM | Laboratorio de Ecologia de Doencas Transmissiveis na Amazonia, Instituto Leonidas e Maria Deane - Fiocruz Amazonia | André Corado; Felipe Naveca; Fernanda Nascimento; George Silva; Karina Pessoa; Luciana Gonçalves; Maria Júlia Brandão; Matilde Mejía; Valdinete Nascimento; Victor Souza |
| EPI_ISL_6492839, EPI_ISL_6493133 | MSHS Clinical Microbiology Laboratories | MSHS Pathogen Surveillance Program | Adolfo García-Sastre; Adriana van de Guchte; Ajay Obla; Alberto Paniz-Mondolfi; Ana S. Gonzalez-Reiche; Angela Amoako; Ashley Salimbangon; Betsaida Salom Melo; Bremy Alburquerque; Brianne Ciferri; Charles Gleason; Daniel Floda; Deena R. Altman; Denise Jurczyszak; Emilia Mia Sordillo; Gintaras Deikus; Giulio Kleiner; Gopi Patel; Hala Alshammary; Harm van Bakel; Irina Oussenko; Jayeeta Dutta; Juan Soto; Julia Matthews; Katherine Beach; Kathryn Twyman; Kayla Russo; Komal Srivastava; Levy Sominsky; Mahmoud Awawda; Marta Luksza; Matthew M. Hernandez; Melissa Gitman; Michael D. Nowak; Mitchell J. Sullivan; Nancy Francoeur; Robert Sebra; Sarah Schaefer; Shelcie Fabre; Shwetha Hara Sridhar; Viviana Simon; Ying-Chih Wang; Zain Khalil; Zenab Khan |
| EPI_ISL_5471871 | Maasstad Ziekenhuis | Maasstad ziekenhuis | Pieter Smit |
| EPI_ISL_5366551 | Maine Health and Environmental Testing Laboratory | Tewhey Lab, The Jackson Laboratory | Barter, M.; Dewey, H.; H. and Tewhey, R.; Iosue, F.; Lynch, R.; Matluk, N.; Munger |
| EPI_ISL_4463015, EPI_ISL_4463016 | Maryland Genomics, Institute for Genome Sciences, University of Maryland School of Medicine | Maryland Genomics, Institute for Genome Sciences, University of Maryland School of Medicine | Claire M; Fraser; Hazen; Holly; Humphrys; Ivette; Jacques; Jonathan; Kranthi; Lim; Lisa D; Luke J; Mike; Ott; Ravel; Roussey; Sadzewicz; Sandra; Santana-Cruz; Tallon; Tracy; Vavikolanu |
| EPI_ISL_5199359, EPI_ISL_5438634 | Massachusetts State Public Health Laboratory | Massachusetts State Public Health Laboratory | Andrew Lang; Glen Gallagher; Sandra Smole; Timelia Fink |
| EPI_ISL_4552962 | Michigan Department of Health and Human Services, Bureau of Laboratories | Michigan Department of Health and Human Services, Bureau of Laboratories | Blankenship HM; Riner D; Soehnlen MK |
| EPI_ISL_4458487 | Microbiology Department. Complexo Hospitalario Universitario de Vigo | Microbiology Department. Complexo Hospitalario Universitario de Vigo | Alvarez M; Cabrera JJ; Carballo R; Cores O; Cortizo S; Davina C; Martinez L; Mediero G; Pena I; Perez S; Potel C; Regueiro B; Rey S; Vasallo FJ; del-Campo V |

| | | | |
|---|---|---|---|
| EPI_ISL_4936740 | NJDOH, Public Health and Environmental Laboratories | NJ PHEL | Byeong Jeong; Chelsea San Filippo; Dana Woell; Lindsey Bodnar; Maria-Magdalene Pugliese; Mohammad M. Ali; Ryan Pachucki; Shiv K. Verma |
| EPI_ISL_5429195 | National Public Health Laboratory, Ministry of Health & Wellness | Viral Special Pathogens Branch, Centers for Disease Control and Prevention | Antoni Comrie; Carl Bruce; Emir Talundzic; Joel Montgomery; John Klena; Justin Lee; Karen Webster-Kerr; Michelle Brown; Michelle Hamilton; Monica Smikle; Nathlee McMorris; Sandra Jackson; Shannon Whitmer; Suwanie Lewis |
| EPI_ISL_4984903, EPI_ISL_4984954, EPI_ISL_4984955 | PECET UDEA | Laboratorio Departamental de Salud Publica de Antioquia | Andres F. Cardona-Rios; Cristian Arbey Velarde Hoyos; Gloria Isabel Escobar; Idabely Betancur Ortiz; Juan P. Hernandez-Ortiz; Maria Stella López |
| EPI_ISL_4458156 | PR Public Health Lab | Centers for Disease Control and Prevention Division of Viral Diseases, Pathogen Discovery | Becky Tsai; Benafsh Sapra; Benjamin Rambo-Martin; Christopher Gulvick; Clinton Paden; Dakota Howard; Dhwani Batra; Doreen Ng; Duncan MacCannell; Erisa Sula; Harry Gao; James Xie; Jason Caravas; John Gao; Joseph Fierro; Kristine Lacek; Matthew Schmerer; Mickey Li; Peter Cook; Scott Sammons; Shatavia Morrison; Tymeckia Kendall; Victoria Caban Figueroa; Yan Meng; Yvette Unoarumhi |
| EPI_ISL_5540381, EPI_ISL_6037383 | Pandemic Response Lab - NYC | Pandemic Response Lab, R&D | Alex Carpio; Cybill del Castillo; Dylan Law; Haiping Hao; Henry Lee; Isabel Fernandez Escapa; Jon Laurent; Melissa Hopkins; Michael Hammerling; Pradeep Bugga; Shinyoung Clair Kang; Sol Rey; William Ward |
| EPI_ISL_4473154 | Public Health Authority of the Slovak Republic | Public Health Authority of the Slovak Republic | Anna Gičová; Barbora Kotvasová; Elena Tichá; Lucia Ševčíková; Miroslav Böhmer; Pavol Mišenko; Terézia Vrabľová; Tomáš Szemes |
| EPI_ISL_4409524, EPI_ISL_4409728, EPI_ISL_4410095, EPI_ISL_4564064, EPI_ISL_4564065, EPI_ISL_6339154 | Public Health Ontario Laboratory | Public Health Ontario Laboratory | Aimin Li; Alireza Eshaghi; Andre Villegas; Ashleigh Sullivan; Christine Frantz; Dean Maxwell; Esha Joshi; Jared Simpson; Jennifer L Guthrie; Jonathan B Gubbay; Karthikeyan Sivaraman; Lawrence Heisler; Matthew Watson; Michael CY Li; Michael Laszloffy; Nahuel Fittipaldi; Philip Banh; Richard de Borja; Samir N Patel; Sandeep Nagra; Sandra Zittermann; Sarah Teatero; Vanessa G Allen; Yao Chen; Yogi Sundaravadanam |
| EPI_ISL_5104527 | Puerto Rico Department of Health | Centers for Disease Control and Prevention, Dengue Branch | Betzabel Flores; Gabriela Paz-Bailey; Gilberto A. Santiago; Glenda Gonzalez; Jorge L. Munoz-Jordan; Keyla Charriez |
| EPI_ISL_4372352, EPI_ISL_4372365, EPI_ISL_4372480, EPI_ISL_4372681, EPI_ISL_4372933, EPI_ISL_4372986, EPI_ISL_4373162, EPI_ISL_5068063, EPI_ISL_5846287, EPI_ISL_5929268 | | | |
| see above | Quest Diagnostics Incorporated | Centers for Disease Control and Prevention Division of Viral Diseases, Pathogen Discovery | A. Gerasimova; A. Perez; B. Anderson; Benjamin Rambo-Martin; Christopher Gulvick; Clinton Paden; Dakota Howard; Dhwani Batra; Duncan MacCannell; Erisa Sula; F. Lacbawan; I. Shlyakhter; Jason Caravas; K. Livingston; Kristine Lacek; L. Bernstein; M. Hua; Matthew Schmerer; P. Tanpaiboon; Peter Cook; R. Kagan; R. Owen; R. Rolando; S. Rosenthal; Scott Sammons; Shatavia Morrison; Tymeckia Kendall; Victoria Caban Figueroa; Y. Liu; Yvette Unoarumhi |
| EPI_ISL_4531131, EPI_ISL_5810934 | Respiratory Virus Unit, Microbiology Services Colindale, Public Health England | COVID-19 Genomics UK (COG-UK) Consortium | PHE Covid Sequencing Team |
| EPI_ISL_4520808 | Rey Fals | Universidad Nacional de Colombia - Laboratorio Genómico One Health | Andres F. Cardona-Rios; Carlos Franco-Muñoz; Carolina Muñoz-Arango; Celeny Ortiz; Daniel O. Maldonado-Perez; Diego A. Álvarez-Díaz; Hector Alejandro Ruiz-Moreno; Idabely Betancur Ortiz; Jorge E. Osorio; Juan P. Hernandez-Ortiz; Karl A Ciuoderis; Katherine Laiton-Donato; Laura Silvana Perez; Lina M. Hurtado; Marcela Mercado-Reyes; Maria Angélica Maya; Maria Stella López; Rita Almanza Payares; Sandra Ines Cano; Simón Villegas Velásquez |
| EPI_ISL_4542967 | Rush University Medical Center | RIPHL at Rush University Medical Center | Diane Springer; Edith Perez; Felix Araujo Perez; Joyce Houlihan; Kevin Kunstman; Laura Furtado; Marieta Hyde; Mary Hayden; Sofiya Bobrovska; Stefan Green |
| EPI_ISL_5031027 | SALUD DIGNA | Instituto Nacional de Medicina Genomica | Abraham Campos-Romero; Cedro-Tanda A; Cruz-Islas Jazmin; Escobar-Arrazola MA; Garnica-Lopez Dora; Herrera-Montalvo LA.; Hidalgo-Miranda A; Luna-Ruiz Marco; Mendoza-Vargas A; Moreno-Camacho José Luis; Ramirez-Vega O; Rangel-DeLeon D; Reyes-Grajeda JP; Rodriguez-Gallegos Jorge; Yair Alfaro-Mora |
| EPI_ISL_5143473, EPI_ISL_5143483, EPI_ISL_5143486, EPI_ISL_5143505, EPI_ISL_6512535 | SOMER | Laboratorio Departamental de Salud Publica de Antioquia | Ana Victoria Valencia Duarte; Cristian Arbey Velarde Hoyos; Gloria Isabel Escobar; Idabely Betancur Ortiz; Juan P. Hernandez-Ortiz; Juan Pablo Isaza Agudelo; Maria Stella López |
| EPI_ISL_4740147 | SURA | Gencore - Universidad de los Andes | Cristian Barrera; Felipe Báez; Gabriela Ariza; Luisa Sacristan; Marcela Guevara; Marcela Mercado; Silvia Restrepo |
| EPI_ISL_4965818, EPI_ISL_4984622, EPI_ISL_5143457, EPI_ISL_5143458, EPI_ISL_5143504 | SURA | Laboratorio Departamental de Salud Publica de Antioquia | Andres F. Cardona-Rios; Cristian Arbey Velarde Hoyos; Gloria Isabel Escobar; Idabely Betancur Ortiz; Juan P. Hernandez-Ortiz; Maria Stella López |
| EPI_ISL_4878272 | SURA Antioquia | Instituto Nacional de Salud-Dirección de Investigación en Salud Pública | Carlos Franco-Muñoz; Carmen Osorio; Diana Malo; Diego A. Álvarez-Díaz; Diego Andrés Prada; Gerardo Santamaría; Hector Alejandro Ruiz-Moreno; Jhonnatan Reales-González; Jorge Rivera; Juan Camilo Martinez; Julian Naizaque; Katherine Laiton-Donato; Lisseth Pardo; Magdalena Wiesner; Marcela Mercado-Reyes; Maria T. Herrera-Sepúlveda; Marta Lopez Blanco; Martha Lucia Ospina Martinez; Paola Rojas; Sergio Gomez; Sheryll Corchuelo; Ángela Alarcon Cruz |
| EPI_ISL_4878283 | SURA Bogota | Instituto Nacional de Salud-Dirección de Investigación en Salud Pública | Carlos Franco-Muñoz; Carmen Osorio; Diana Malo; Diego A. Álvarez-Díaz; Diego Andrés Prada; Gerardo Santamaría; Hector Alejandro Ruiz-Moreno; Jhonnatan Reales-González; Jorge Rivera; Juan Camilo Martinez; Julian Naizaque; Katherine Laiton-Donato; Lisseth Pardo; Magdalena Wiesner; Marcela Mercado-Reyes; Maria T. Herrera-Sepúlveda; Marta Lopez Blanco; Martha Lucia Ospina Martinez; Paola Rojas; Sergio Gomez; Sheryll Corchuelo; Ángela Alarcon Cruz |
| EPI_ISL_5152646, EPI_ISL_5152661 | SYNLAB | CIAT, Laboratorio de Virologia | Ana M. Leiva; Diana Lopez; Programa Nacional de Caracterización Genómica de SARS/CoV/2; WilmerJ. Cuellar |
| EPI_ISL_4878281, EPI_ISL_4878293 | SYNLAB | Instituto Nacional de Salud-Dirección de Investigación en Salud Pública | Carlos Franco-Muñoz; Carmen Osorio; Diana Malo; Diego A. Álvarez-Díaz; Diego Andrés Prada; Gerardo Santamaría; Hector Alejandro Ruiz-Moreno; Jhonnatan Reales-González; Jorge Rivera; Juan Camilo Martinez; Julian Naizaque; Katherine Laiton-Donato; Lisseth Pardo; Magdalena Wiesner; Marcela Mercado-Reyes; Maria T. Herrera-Sepúlveda; Marta Lopez Blanco; Martha Lucia Ospina Martinez; Paola Rojas; Sergio Gomez; Sheryll Corchuelo; Ángela Alarcon Cruz |
| EPI_ISL_5202113, EPI_ISL_5202128, EPI_ISL_5202151, EPI_ISL_5202240, EPI_ISL_5202241, EPI_ISL_5202248, EPI_ISL_5202354, EPI_ISL_5202382, EPI_ISL_5202421, EPI_ISL_5202610, EPI_ISL_5202611, EPI_ISL_5202693 | | | |
| see above | SYNLAB | LSPSDS | Alejandro Gomez Lopez; Gabriela Delgado Murcia; Johana Hernandez Toloza; Marcela Castano Rodriguez; Marcela Mercado |
| EPI_ISL_4984548, EPI_ISL_4984717, EPI_ISL_4984831 | SYNLAB | Laboratorio Departamental de Salud Publica de Antioquia | Andres F. Cardona-Rios; Cristian Arbey Velarde Hoyos; Gloria Isabel Escobar; Idabely Betancur Ortiz; Juan P. Hernandez-Ortiz; Maria Stella López |
| EPI_ISL_4878527 | SYNLAB | Universidad Nacional de Colombia - Laboratorio Genómico One Health | Andres F. Cardona-Rios; Carlos Franco-Muñoz; Carolina Muñoz-Arango; Celeny Ortiz; Daniel O. Maldonado-Perez; Diego A. Álvarez-Díaz; Hector Alejandro Ruiz-Moreno; Idabely Betancur Ortiz; Jorge E. Osorio; Juan P. Hernandez-Ortiz; Karl A Ciuoderis; Katherine Laiton-Donato; Laura Silvana Perez; Lina M. Hurtado; Marcela Mercado-Reyes; Maria Angélica Maya; Maria Stella López; Rita Almanza Payares; Sandra Ines Cano; Simón Villegas Velásquez |
| EPI_ISL_4410699 | Salud Digna | Instituto Nacional de Medicina Genomica | Abraham Campos-Romero; Cedro-Tanda A; Cisneros-Villanueva M; Cruz-Islas Jazmin; Escobar-Arrazola MA; Garnica-Lopez Dora; Herrera-Montalvo LA.; Hidalgo-Miranda A; Luna-Ruiz Marco; Mendoza-Vargas A; Moreno-Camacho José Luis; Ramirez-Vega O; Rangel-DeLeon D; Reyes-Grajeda JP; Rodriguez-Gallegos Jorge; Yair Alfaro-Mora |
| EPI_ISL_4920267 | Servicio Microbiología H.U. Dr. Negrín | Servicio Microbiología H.U. Dr. Negrín | Ana Bordes Benítez; Bartolomé Gómez Arroyo; Francisco Javier Chamizo López |
| EPI_ISL_5655844 | Servicio de Microbiología Clínica (Complejo Hospitalario de Navarra, Pamplona) | Centro de Secuenciación NASERTIC | Ana Miqueleiz; Ana Navascués; Carmen Ezpeleta Baquedano |
| EPI_ISL_6080358 | Shamir Medical Center (Asaf Harofe) | Shamir Medical Center (Asaf Harofe) | Abu Hamad Ramzia; Adina Bar Chaim; Anna Vishnevsky; Chen Weiner; Nir Rainy; Patricia Benveniste-Lekovitz; Reut Sorek Abramovich; Yevgeni Yegorov |
| EPI_ISL_6402371 | TGen North | TGen North | Brett Van Tassel; Chris French; Darrin Lemmer; Dave Engelthaler; Hayley Yaglom; Heather Centner; Jolene Bowers |
| EPI_ISL_5334484 | UNIDAD HEMATOLOGICA ESPECIALIZADA | Instituto Nacional de Salud-Dirección de Investigación en Salud Pública | Beatriz de Arco; Carlos Franco-Muñoz; Diego A. Álvarez-Díaz; Diego Andrés Prada; Dioselina Peláez-Carvajal; Gerardo Santamaría; Héctor Alejandro Ruiz-Moreno; Jhonnatan Reales-González; Jorge Rivera; Julián Naizaque; Katherine Laiton-Donato; Marcela Mercado-Reyes.; Martha Lucia Ospina Martínez; María T. Herrera-Sepúlveda; Paola Rojas; Sheryll Corchuelo; Tatiana Cobos |
| EPI_ISL_6012788 | UNILABS | Instituto Nacional de Saude (INSA) | Borges et al |
| EPI_ISL_5055405 | UNIVERSIDAD DE ANTIOQUIA - LIME | Instituto Nacional de Salud-Dirección de Investigación en Salud Pública | Beatriz de Arco; Carlos Franco-Muñoz; Carmen Osorio; Diana Malo; Diego A. Álvarez-Díaz; Diego Andrés Prada; Gerardo Santamaría; Hector Alejandro Ruiz-Moreno; Jhonnatan Reales-González; Jorge Rivera; Juan Camilo Martinez; Julian Naizaque; Katherine Laiton-Donato; Lisseth Pardo; Magdalena Wiesner; Marcela Mercado-Reyes; Maria T. Herrera-Sepúlveda; Marta Lopez Blanco; Martha Lucia Ospina Martinez; Paola Rojas; Sergio Gomez; Sheryll Corchuelo; Tatiana Cobos; Ángela Alarcon Cruz |
| EPI_ISL_4472792 | UW Virology Lab | UW Virology Lab | Alexander Greninger; Hong Xie; Keith R Jerome; Maria Lukes; Meei-Li Huang; Nathan Breit; Patrick Mathias; Pavitra Roychoudhury; Ricardo Perez; Robert J. Livingston; Sean Ellis; Shah Mohamed Bakhash; Tien V. Nguyen |
| EPI_ISL_4879059 | Unidad de Investigación Médica de Yucatán (UIMY) | Instituto de Biotecnología de la UNAM | ; Alejandra García-Gasca; Alejandra Hernández-Terán; Alejandro Sánchez-Flores; Alfredo Herrera-Estrella; Alicia Ocaña-Mondragón; Andreu Comas-García; Angel Gustavo Salas-Lais; Antonio Loza Román; Bernardo Martínez-Miguel; Blanca Taboada; Brenda Irasema Maldonado-Meza; Bruno Gómez-Gil; Carla Ivón Herrera-Najera; Carlos F. Arias; Celia Boukadida; Clara Esperanza Santacruz-Tinoco; Concepción Grajales-Muñiz; Consorcio Mexicano de Vigilancia Genómica (CoViGen-Mex). Authors (in alphabetical order): Julio Elias Alvarado-Yaah; Cristóbal Cháidez-Quiróz; Célida Duque Molina; Célida Martínez- Rodríguez; Daniel Fregoso-Rueda; Daniel Lira Morales; Eduardo Becerril-Vargas; Fernando Fontove-Herrera; Fidencio Mejía-Nepomuceno; Francisco Pulido; Gloria Elena Espinosa-Ayala; Gloria María Molina-Salinas; Gloria Vazquez; Hector Esteban Paz-Juárez; Hector Montoya-Fuentes; Helen Haydee Fernanda Ramírez-Plascencia; Irvin González-López; Jean Pierre González; Jesús Hernández; Joel Armando Vázquez-Pérez).; Jorge Salas-Hernández; José Antonio Enciso-Moreno; José Arturo Martínez-Orozco; José Esteban Muñoz-Medina; José de Jesús Nuñez-Contreras; Juan Bautista Chale-Dzul; Julissa Enciso-Ibarra; Kathia Elizabeth Tapia-Diaz; Luis Alberto Ochoa-Carrera; Margarita Matías-Florentino; Mario Mújica-Sánchez; Marissa Perez-Garcia; María Guadalupe Santiago-Mauricio; María Guadalupe de Jesús Mireles-Rivera; Nelly Sélem-Mojica; Pavel Isa; Ricardo Ciria Merce; Ricardo Grande; Rosa María Gutiérrez Rios; Santiago Ávila-Ríos; Selene Zárate; Susana Lopez; Verónica Mata-Haro; Victor Eduardo García-Arias; Víctor Hugo Borja-Aburto |
| EPI_ISL_5887477 | Universidad San Sebastian | "Facultad de Ciencias de la Vida, UNAB" | "Claudio Meneses; Ariel Orellana"; Claudio Olmos; Daniel Leon; Dayan Sanhueza; Eduardo Castro; Gonzalo Campaña; Macarena Bastias; Paola Pidal; Ricardo Yusta; Sebastian Wolter; Susana Saez; Víctor Monreal; Waldo Diaz |
| EPI_ISL_5065684, EPI_ISL_5065695, EPI_ISL_5065713, | Universidad del Sinú - Sede Cartagena | Centro de Genética y Biología Molecular - Universidad del Magdalena | Andrea M. Ramírez Hernandez; Angel Oviedo Marquez; Daniel Bautista; Edison Lea-Ch; Lyda R. Castro; Maria Teresa Mojica-Ortiz |

| | | | |
|---|---|---|---|
| EPI_ISL_5065725 | | | |
| EPI_ISL_4419161, EPI_ISL_4419165, EPI_ISL_4419174 | Universidad del Valle, LDAB-Laboratorio de Diagnostico de Agentes Biologicos | Universidad del Valle | Programa Nacional de Caracterización Genómica de SARS-CoV-2 |
| EPI_ISL_5397463 | Universitätsklinikum Köln; Institut für Virologie | Robert Koch Institute | |
| EPI_ISL_5421948 | Usansolo-Galdakao University Hospital | Cruces University Hospital | Ana Belén de la Hoz; Ana Gual-de-Torrella; Izaskun Alejo-Cancho; Mikel Gallego |
| EPI_ISL_4494213, EPI_ISL_5334261, EPI_ISL_5334279 | Utah Public Health Laboratory | Utah Public Health Laboratory | Erin L. Young; John Arnn; Kelly F. Oakeson; Olinto Linares-Perdomo; Pooja Gupta |
| EPI_ISL_4760533 | V20 | Y20 | Amy Bauer; Jenny Lentz; Manjeet Khubbar; Nandhukumar Balakrishnan; Samantha Scott; Sanjib Bhattacharyya |
| EPI_ISL_5511723 | Wyoming Public Health Laboratory | Wyoming Public Health Laboratory | Ashley Norberg; Brian Dominguez; Cari Sloma; Channing Weber; Chayse Rowley; Elliot Thomasson; Jim Mildenberger; Marley Goetz; Robert Petit; Sam Britz; Taylor Fearing; and Rob Christensen |
| EPI_ISL_4520825 | Yamina Cumplido Romero | Universidad Nacional de Colombia - Laboratorio Genómico One Health | Andres F. Cardona-Rios; Carlos Franco-Muñoz; Carolina Muñoz-Arango; Celeny Ortiz; Daniel O. Maldonado-Perez; Diego A. Álvarez-Díaz; Hector Alejandro Ruiz-Moreno; Idabely Betancur Ortiz; Jorge E. Osorio; Juan P. Hernandez-Ortiz; Karl A Ciuoderis; Katherine Laiton-Donato; Laura Silvana Perez; Lina M. Hurtado; Marcela Mercado-Reyes; Maria Angélica Maya; Maria Stella López; Rita Almanza Payares; Sandra Ines Cano; Simón Villegas Velásquez |

We gratefully acknowledge the following Authors from the Originating laboratories responsible for obtaining the specimens, as well as the Submitting laboratories where the genome data were generated and shared via GISAID, on which this research is based.

All Submitters of data may be contacted directly via www.gisaid.org

Authors are sorted alphabetically.

Acknowledgement EPI_SET Identifier: EPI_SET_20220328wc

| Accession ID | Originating Laboratory | Submitting Laboratory | Authors |
|---|---|---|---|
| EPI_ISL_7660301 | ADILAB | Laboratorio Departamental de Salud Publica de Antioquia | Ana Victoria Valencia Duarte; Cristian Arbey Velarde Hoyos; Gloria Isabel Escobar; Idabely Betancur Ortiz; Juan P. Hernandez-Ortiz; Juan Pablo Isaza Agudelo; Maria Stella López |
| EPI_ISL_7809039 | AFIP | Instituto Butantan | Antonio Jorge Martins; Claudia Renata dos Santos Barros; David Schlesinger; Debora Botequio Moretti; Dimas Tadeu Covas; Elaine Cristina Marqueze; Elaine Vieira Santos; Evandra Strazza Rodrigues; Heidge Fukumasu; Jayme Augusto de Souza-Neto; José Salvatore Leister Patané; Luiz Alcantara; Luiz Lehmann Coutinho; Maria Carolina Elias; Maurício Lacerda Nogueira; Rafael dos Santos Bezerra; Raul Machado Neto; Rejane Maria Tommasini Grotto; Ricardo Haddad; Sandra Coccuzzo Sampaio Vessoni; Simone Kashima; Svetoslav Nanev Slavov; Vincent Louis Viala |
| EPI_ISL_6649094, EPI_ISL_6786693 | Aegis Sciences Corporation | Centers for Disease Control and Prevention Division of Viral Diseases, Pathogen Discovery | Alec Vest; Benjamin Rambo-Martin; Christopher Gulvick; Clinton Paden; Cyndi Clark; Dakota Howard; Dhwani Batra; Dillon Nall; Duncan MacCannell; Erisa Sula; Ethan Sanders; Holly Houdeshell; Jason Caravas; Kristine Lacek; Matthew Hardison; Matthew Schmerer; Ola Kvalvaag; Patrick Campbell; Peter Cook; Rob Case; Scott Sammons; Shatavia Morrison; Shaun Westlund; Tymeckia Kendall; Victoria Caban Figueroa; Vikramsinha Ghorpade; Yvette Unoarumhi |
| EPI_ISL_7711703 | Area De Salud Cartago | Universidad de Costa Rica e Instituto Costarricense de Investigacion y Ensenanza en Nutricion y Salud | Brenes Hebleen; Camacho Erwin; Campos-Sanchez Rebeca; Ceballos Ana; Cordero Estela; Cristancho Marco; Duarte Francisco; Fernandez-Do Porto Dario; Herrera-Estrella Alfredo; Jimenez-Moraila Beatriz; Kreuze Jan; Mireles-Rivera Guadalupe; Molina-Mora Jose; Munoz-Medina Jose Esteban; Negri Tatiana; Nunes Gisele; Oliveira Guilherme; Oliveira Renato; Reales-Gonzalez Jhonnatan; Remes-Lenicov Federico; Reyes Alejandro; Sosa Ezequiel; Soto Claudio; Tsukayama Pablo; Turjanski Adrian; and Zayat Jonathan |
| EPI_ISL_7711707 | Area De Salud Heredia Cubujuchi | Universidad de Costa Rica e Instituto Costarricense de Investigacion y Ensenanza en Nutricion y Salud | Brenes Hebleen; Camacho Erwin; Campos-Sanchez Rebeca; Ceballos Ana; Cordero Estela; Cristancho Marco; Duarte Francisco; Fernandez-Do Porto Dario; Herrera-Estrella Alfredo; Jimenez-Moraila Beatriz; Kreuze Jan; Mireles-Rivera Guadalupe; Molina-Mora Jose; Munoz-Medina Jose Esteban; Negri Tatiana; Nunes Gisele; Oliveira Guilherme; Oliveira Renato; Reales-Gonzalez Jhonnatan; Remes-Lenicov Federico; Reyes Alejandro; Sosa Ezequiel; Soto Claudio; Tsukayama Pablo; Turjanski Adrian; and Zayat Jonathan |
| EPI_ISL_8482343, EPI_ISL_8482347, EPI_ISL_8482372, EPI_ISL_8482721, EPI_ISL_8482723, EPI_ISL_8482739, EPI_ISL_8482740, EPI_ISL_8482746, EPI_ISL_8482750, EPI_ISL_8482922, EPI_ISL_8482926, EPI_ISL_8482928, EPI_ISL_8482929, EPI_ISL_8482930, EPI_ISL_8482931, EPI_ISL_8482933, EPI_ISL_8482939, EPI_ISL_8482941, EPI_ISL_8482944, EPI_ISL_8482952, EPI_ISL_8482955, EPI_ISL_8482957, EPI_ISL_8482958, EPI_ISL_8482965, EPI_ISL_8482966, EPI_ISL_8482974, EPI_ISL_8482977, EPI_ISL_8482979, EPI_ISL_8482997 | | | |
| see above | Biotechnology Core Facility Branch, Centers for Disease Control and Prevention | Centers for Disease Control and Prevention Division of Viral Diseases, Pathogen Discovery | ; Alex Burgin; Alison Nicholson; Anir Enkhbat; Antoni Comrie; Ariuntuya Ganbat; Battur Lkhagvaa; Ben Rambo-Martin; Bilegtsaikhan Sukhee; Carl Bruce; Clinton Paden; Dakota Howard; Davaasukh Battumur; Dave Wentworth; Dhwani Batra; Ganbold Dalantai; Gulnar Genden; Jasmine Padilla; Joseph Madden; Justin Lee; Karen Webster-Kerr; Kristen Knipe; Kristine Lacek; Mark Burroughs; Matthew Schmerer; Meghan Bentz; Michelle Brown; Michelle Hamilton; Mili Sheth; Nathlee McMorris; Peter Cook; Sam Shepard; Sandra Jackson; Sarah Nobles; Suwanie Lewis; Vivien Dugan; Yvette Unoarumhi |
| EPI_ISL_7308809, EPI_ISL_8395542, EPI_ISL_8395951, EPI_ISL_8396009, EPI_ISL_8396231, EPI_ISL_8397336 | CDPH VBL | California Department of Public Health | CDPH-COVIDNet; Emily Smith on behalf of CDPH-COVIDNet |
| EPI_ISL_7507357 | Cedars-Sinai Medical Center, Molecular Pathology Laboratory of Department of Pathology & Laboratory Medicine and Genomic Core | Cedars-Sinai Medical Center, Molecular Pathology Laboratory of Department of Pathology & Laboratory Medicine and Genomic Core | Brian Davis; Eric Vail; Jasmine T Plummer; Jorge Mario Sincuir Martinez; Stephanie Chen; Wenjuan Zhang |
| EPI_ISL_7809634 | Colorado Department of Public Health and Environment | Colorado Department of Public Health and Environment | Alexandria Rossheim; Diana Ir; Emily A. Travanty; Laura Bankers; Mandy Waters; Michael A. Martin; Molly C. Hetherington-Rauth; Sarah Elizabeth Totten; Shannon R. Matzinger |
| EPI_ISL_8357381, EPI_ISL_8357385, EPI_ISL_8357389 | Compensar Calle 26 | LSPSDS | Alejandro Gomez Lopez; Gabriela Delgado Murcia; Johana Hernandez Toloza; Marcela Mercado |
| EPI_ISL_8357369 | Compensar Calle 63 | LSPSDS | Alejandro Gomez Lopez; Gabriela Delgado Murcia; Johana Hernandez Toloza; Marcela Mercado |
| EPI_ISL_6941250, EPI_ISL_7039253 | DASA | DASA | Adriano Bonaldi; Angelica Hristov; Annelise Lopes; Bianca Cota; Camila Romano; Cristina Oliveira; Jose Levi; Lidia Yamamoto; Luciane Sussuchi; Paulo Pierry; Rodrigo Guarischi; Rodrigo Salazar |
| EPI_ISL_7808713 | DeRisi Lab, University of California, San Francisco | DeRisi Lab, University of California, San Francisco | Amy Kistler; Anthea Mitchell; Carina Marquez; Diane Havlir; Diane Jones; Douglas Black; Eric Chow; Gabriel Chamie; Genay Pilarowski; Grace Wang; IDseq Team; Jackie Martinez; James Peng; Jamin Liu; John Schrom; Jon Jacobo; Lucy Li; Luis Rubio; Manu Vanaerschot; Matthew Laurie; Matthias Hapte-Selassie; Maya Petersen; Patrick Ayscue; Sabrina Mann; Sara Sunshine; Susana Rojas; Susy Rojas; Valerie Tulier-Laiwa; and Joseph DeRisi |
| EPI_ISL_7908699, EPI_ISL_7912756 | Department of Bacteria, Parasites and Fungi, Statens Serum Institut, Copenhagen, Denmark | Statens Serum Institut Bioinformatics and Microbial Genomics | Danish Covid-19 Genome Consortium |
| EPI_ISL_7747681 | Emory Molecular Diagnostics Laboratory, Emory Healthcare | Piantadosi Lab, Emory Department of Pathology | Ahmed Babiker; Anne Piantadosi; Dara Khosravi |
| EPI_ISL_7319328, EPI_ISL_7319464 | Florida Bureau of Public Health Laboratories | Florida Bureau of Public Health Laboratories | Jason Blanton; Namratha Tarigopula; Sarah Schmedes; Tiffany Splatt |
| EPI_ISL_6620229 | Fulgent Genetics | Centers for Disease Control and Prevention Division of Viral Diseases, Pathogen Discovery | Becky Tsai; Benafsh Sapra; Benjamin Rambo-Martin; Christopher Gulvick; Clinton Paden; Dakota Howard; Dhwani Batra; Doreen Ng; Duncan MacCannell; Erisa Sula; Harry Gao; James Xie; Jason Caravas; John Gao; Joseph Fierro; Kristine Lacek; Matthew Schmerer; Mickey Li; Peter Cook; Scott Sammons; Shatavia Morrison; Tymeckia Kendall; Victoria Caban Figueroa; Yan Meng; Yvette Unoarumhi |
| EPI_ISL_6675476, EPI_ISL_6675503, EPI_ISL_6675612, EPI_ISL_6675615, EPI_ISL_6675619, EPI_ISL_6675620, EPI_ISL_6675624, EPI_ISL_6675673, EPI_ISL_6675694, EPI_ISL_6675857, EPI_ISL_7778190, EPI_ISL_7778208, EPI_ISL_8187693 | | | |
| see above | Genetica Molecular and Subdepartamento de Virologia ISP Chile | Instituto de Salud Publica de Chile | Andres Castillo; Barbara Parra; Constanza Campano; Gisselle Barra; Javier Tognarelli; Jorge Fernandez; Karen Orostica; Loredana Arata; Patricia Bustos; Rodrigo Fasce; Soledad Ulloa |
| EPI_ISL_7307270, EPI_ISL_7476228, EPI_ISL_7476314, EPI_ISL_7476330, EPI_ISL_7476368, EPI_ISL_7476423, EPI_ISL_7476449, EPI_ISL_7476483, EPI_ISL_7476510, EPI_ISL_7476523, EPI_ISL_7476597, EPI_ISL_7476651, EPI_ISL_7476741, EPI_ISL_7476758, EPI_ISL_7476784, EPI_ISL_7476794, EPI_ISL_7476796, EPI_ISL_7476801 | | | |
| see above | Grupo de Investigación en Enfermedades Tropicales del Ejército (GINETEJ), Laboratorio de Referencia e Investigación, Dirección de Sanidad Ejército, Bogotá, Colombia | Centro de Investigaciones en Microbiología y Biotecnología-UR (CIMBIUR), Facultad de Ciencias Naturales, Universidad del Rosario, Bogotá, Colombia | Alberto E. Paniz-Mondolfi; Alberto Paniz-Mondolfi; Angie L. Ramírez; Angie Ramírez; Beatriz Ariza; Camilo A. Correa-Cárdenas; Camilo Correa-Cardenas; Carlos Gómez-Restrepo; Claudia Cardozo-Romero; Claudia Mendez; Claudia Méndez; David-Santiago Quevedo; Enzo Guerrero-Araya; Guido España; Hernando Díaz; Juan David Ramírez; Juliana Cuervo-Rojas; Julie Perez; Julie Pérez; Luz H. Patiño; Manuel-Antonio Franco; Maria Clara Duque; Maria-Clara Duque; Marina Muñoz; Nathalia Ballesteros; Nicolas Luna; Nicolás Luna; Sergio Castañeda; Zulma M. Cucunubá |
| EPI_ISL_6945580 | HOSPITAL UNIVERSITARIO SAN IGNACIO | Centro de Genética y Biología Molecular - Universidad del Magdalena | Andrea M. Ramírez Hernandez; Angel Oviedo Marquez; Daniel Bautista; Edison Lea-Ch; Lyda R. Castro; Maria Teresa Mojica-Ortiz |
| EPI_ISL_8181151 | Helix | Centers for Disease Control and Prevention Division of Viral Diseases, Pathogen Discovery | Benjamin Rambo-Martin; Christopher Gulvick; Clinton Paden; Dakota Howard; Dhwani Batra; Duncan MacCannell; Erisa Sula; Helix CA; Jason Caravas; Kristine Lacek; Matthew Schmerer; Peter Cook; Scott Sammons; Shatavia Morrison; Tymeckia Kendall; Victoria Caban Figueroa; Yvette Unoarumhi |
| EPI_ISL_7711705 | Hospital Dr. Carlos Luis Valverde Vega | Universidad de Costa Rica e Instituto Costarricense de Investigacion y Ensenanza en Nutricion y Salud | Brenes Hebleen; Camacho Erwin; Campos-Sanchez Rebeca; Ceballos Ana; Cordero Estela; Cristancho Marco; Duarte Francisco; Fernandez-Do Porto Dario; Herrera-Estrella Alfredo; Jimenez-Moraila Beatriz; Kreuze Jan; Mireles-Rivera Guadalupe; Molina-Mora Jose; Munoz-Medina Jose Esteban; Negri Tatiana; Nunes Gisele; Oliveira Guilherme; Oliveira Renato; Reales-Gonzalez Jhonnatan; Remes-Lenicov Federico; Reyes Alejandro; Sosa Ezequiel; Soto Claudio; Tsukayama Pablo; Turjanski Adrian; and Zayat Jonathan |
| EPI_ISL_7711704 | Hospital Mexico | Universidad de Costa Rica e Instituto Costarricense de Investigacion y Ensenanza en Nutricion y Salud | Brenes Hebleen; Camacho Erwin; Campos-Sanchez Rebeca; Ceballos Ana; Cordero Estela; Cristancho Marco; Duarte Francisco; Fernandez-Do Porto Dario; Herrera-Estrella Alfredo; Jimenez-Moraila Beatriz; Kreuze Jan; Mireles-Rivera Guadalupe; Molina-Mora Jose; Munoz-Medina Jose Esteban; Negri Tatiana; Nunes Gisele; Oliveira Guilherme; Oliveira Renato; Reales-Gonzalez Jhonnatan; Remes-Lenicov Federico; Reyes Alejandro; Sosa Ezequiel; Soto Claudio; Tsukayama Pablo; Turjanski Adrian; and Zayat Jonathan |
| EPI_ISL_7606117, EPI_ISL_7606119 | Hospital Nacional de Niños | Inciensa, Instituto Costarricense de Investigación y Enseñanza en Nutrición y Salud | Cristian Pérez-Corrales |
| EPI_ISL_7711706 | Hospital San Francisco De Asis [San Isidro/Grecia/Alajuela] | Universidad de Costa Rica e Instituto Costarricense de Investigacion y Ensenanza en Nutricion y Salud | Brenes Hebleen; Camacho Erwin; Campos-Sanchez Rebeca; Ceballos Ana; Cordero Estela; Cristancho Marco; Duarte Francisco; Fernandez-Do Porto Dario; Herrera-Estrella Alfredo; Jimenez-Moraila Beatriz; Kreuze Jan; Mireles-Rivera Guadalupe; Molina-Mora Jose; Munoz-Medina Jose Esteban; Negri Tatiana; Nunes Gisele; Oliveira Guilherme; Oliveira Renato; Reales-Gonzalez Jhonnatan; Remes-Lenicov Federico; Reyes Alejandro; Sosa Ezequiel; Soto Claudio; Tsukayama Pablo; Turjanski Adrian; and Zayat Jonathan |
| EPI_ISL_7211378 | Hospital Universitari Dr. Josep Trueta | Institut d'Investigació Biomèdica | Bernat del Olmo; Mel·lina Pinsach; Meritxell Deulofeu; Nuria Esther Neto; Paula Costa |

| | | | |
|---|---|---|---|
| | | de Girona \| Hospital Universitari Dr. Josep Trueta | |
| EPI_ISL_7307162, EPI_ISL_7307166, EPI_ISL_7307182, EPI_ISL_7307194, EPI_ISL_7307198, EPI_ISL_7307202, EPI_ISL_7307215, EPI_ISL_7307223, EPI_ISL_7307227, EPI_ISL_7307233, EPI_ISL_7307280, EPI_ISL_7307292, EPI_ISL_7339454, EPI_ISL_7339455, EPI_ISL_7339458, EPI_ISL_7339470, EPI_ISL_7829667 | | | |
| see above | Hospital Universitario San Ignacio | Centro de Investigaciones en Microbiología y Biotecnología-UR (CIMBIUR), Facultad de Ciencias Naturales, Universidad del Rosario, Bogotá, Colombia | Alberto Paniz-Mondolfi; Angie Ramírez; Beatriz Ariza; Beatriz Elena Ariza; Camilo A. Correa-Cárdenas; Carlos Gómez-Restrepo; Claudia Cardozo-Romero; Claudia Cecilia Cardozo; Claudia Méndez; David-Santiago Quevedo; Guido España; Hernando Díaz; Juan David Ramírez; Juliana Cuervo-Rojas; Juliana Maria Cuervo; Julie Pérez; Luz H. Patiño; Manuel Antonio Franco; Manuel-Antonio Franco; Maria-Clara Duque; Marina Muñoz; Nathalia Ballesteros; Nicolas Luna; Sandra Liliana Valderrama; Sergio Castañeda; Zulma M. Cucunubá; Zulma Milena Cucunubá |
| EPI_ISL_8033762 | IHU Mediterranee Infection | IHU Mediterranee Infection | Bernard La Scola; Philippe Colson; et al. |
| EPI_ISL_8097176, EPI_ISL_8097183, EPI_ISL_8097185 | IIPSI SOL WAYUU | Instituto Nacional de Salud-Dirección de Investigación en Salud Pública | Beatriz de Arco; Carlos Franco-Muñoz; Diego A. Álvarez-Díaz; Diego Andrés Prada; Dioselina Peláez-Carvajal; Gerardo Santamaría; Héctor Alejandro Ruiz-Moreno; Jhonnatan Reales-González; Jorge Rivera; Julián Naizaque; Katherine Laiton-Donato; Marcela Mercado-Reyes.; Martha Lucia Ospina Martínez; María T. Herrera-Sepúlveda; Paola Rojas-Estevez; Sheryll Corchuelo; Tatiana Cobos |
| EPI_ISL_6976265, EPI_ISL_6989642, EPI_ISL_6990137, EPI_ISL_6990142, EPI_ISL_7829649, EPI_ISL_7829658, EPI_ISL_7831126, EPI_ISL_7831406, EPI_ISL_7831409 | | | |
| see above | IVIC | Laboratorio de Virología Molecular | Carmen L Loureiro; CoViMol Group; CoViVen Group; Domingo J Garzaro; Flor H Pujol; Héctor R Rangel; José Luis Zambrano; Lieska Rodríguez; Mariana Hidalgo; Pierina D´Angelo; Rossana C Jaspe; Víctor Alarcón; Yoneira Sulbaran; Zoila Moros |
| EPI_ISL_7709464, EPI_ISL_7709473, EPI_ISL_7709474, EPI_ISL_8183444, EPI_ISL_8183483 | Institute of Microbiology, Universidad San Francisco de Quito | Institute of Microbiology, Universidad San Francisco de Quito | Belén Prado-Vivar; Bernardo Gutiérrez; Erika B. Muñoz; Fernanda Zurita; Gabriel Trueba; Juan Jose Villacís; Juan José Guadalupe; Mateo Carvajal; Michelle Grunauer; Monica Becerra-Wong; Nelson Montalvan; Patricio Rojas-Silva; Paúl Cárdenas; Sully Márquez; Verónica Barragán; Verónica Pacheco |
| EPI_ISL_7813104 | Instituto Mexicano del Seguro Social (IMSS) | CABANA: Unidad de Genómica Avanzada del Centro de Investigación y de Estudios Avanzados (UGA-LANGEBIO, CINVESTAV) | Adrián Turjanski; Alejandro Reyes; Alfredo Herrera-Estrella; Ana Ceballos; Beatriz Jiménez-Moraila; Claudio Soto; Darío Fernández-Do Porto; Erwin Camacho; Estela Cordero; Ezequiel Sosa; Federico Remes-Lenicov; Francisco Duarte; Gisele Nunes; Guadalupe Mireles-Rivera; Guilherme Oliveira; Hebleen Brenes; Jan Kreuze; Jhonnatan Reales-González; Jonathan Zayat; José Esteban Muñoz-Medina; José Molina-Mora; Marco Cristancho; Pablo Tsukayama; Rebeca Campos-Sánchez; Renato Oliveira; Tatiana Negri |
| EPI_ISL_7747261 | Instituto Nacional de Enfermedades Respiratorias (INER) | Centro de Investigación en Enfermedades Infecciosas (CIENI), Instituto Nacional de Enfermedades Respiratorias (INER) | Alejandra García-Gasca; Alejandra Hernández-Terán; Alejandro Sánchez-Flores; Alfredo Herrera-Estrella; Alicia Ocaña-Mondragón; Andreu Comas-García; Angel Gustavo Salas-Lais; Antonio Loza Román; Bernardo Martínez-Miguel; Blanca Taboada; Brenda Irasema Maldonado-Meza; Bruno Gómez-Gil; Carla Ivón Herrera-Najera; Carlos F. Arias; Celia Boukadida; Clara Esperanza Santacruz-Tinoco; Concepción Grajales-Muñiz; Consorcio Mexicano de Vigilancia Genómica (CoViGen-Mex). Authors (in alphabetical order): Julio Elias Alvarado-Yaah; Cristóbal Cháidez-Quiróz; Célida Duque Molina; Célida Martínez-Rodríguez; Daniel Fregoso-Rueda; Daniel Lira Morales; Eduardo Becerril-Vargas; Eduardo Rivera-Martínez; Fernando Fontove-Herrera; Fidencio Mejía-Nepomuceno; Francisco Pulido; Gabriel Chavira-Trujillo; Gloria Elena Espinosa-Ayala; Gloria María Molina-Salinas; Gloria Vazquez; Hector Montoya-Fuentes; Helen Haydee Fernanda Ramírez-Plascencia; Irvin González-López; Jean Pierre González; Jesús Hernández; Joel Armando Vázquez-Pérez.; Jorge Salas-Hernández; José Antonio Enciso-Moreno; José Arturo Martínez-Orozco; José Esteban Muñoz-Medina; José de Jesús Nuñez-Contreras; Juan Bautista Chale-Dzul; Julissa Enciso-Ibarra; Kathia Elizabeth Tapia-Diaz; Luis Alberto Ochoa-Carrera; Margarita Matías-Florentino; Mario Mújica-Sánchez; Marissa Perez-Garcia; María Eugenia Jiménez-Corona; María Guadalupe Santiago-Mauricio; María Guadalupe de Jesús Mireles-Rivera; Nelly Sélem-Mojica; Pavel Isa; Ricardo Ciria Merce; Ricardo Grande; Rosa María Gutiérrez Rios; Rosario Vazquez-Larios; Santiago Ávila-Ríos; Selene Zárate; Susana Lopez; Verónica Mata-Haro; Victor Eduardo García-Arias; Víctor Hugo Borja-Aburto |
| EPI_ISL_7951686 | LABORATORIO CLINICO COMPENSAR LOS COBOS | Instituto Nacional de Salud-Dirección de Investigación en Salud Pública | Beatriz de Arco; Carlos Franco-Muñoz; Diego A. Álvarez-Díaz; Diego Andrés Prada; Dioselina Peláez-Carvajal; Gerardo Santamaría; Héctor Alejandro Ruiz-Moreno; Jhonnatan Reales-González; Jorge Rivera; Julián Naizaque; Katherine Laiton-Donato; Marcela Mercado-Reyes.; Martha Lucia Ospina Martínez; María T. Herrera-Sepúlveda; Paola Rojas-Estevez; Sheryll Corchuelo; Tatiana Cobos |
| EPI_ISL_8357393, EPI_ISL_8357394, EPI_ISL_8357396, EPI_ISL_8357403 | LABORATORIO LORENA VEJARANO SAS | LSPSDS | Alejandro Gomez Lopez; Gabriela Delgado Murcia; Johana Hernandez Toloza; Marcela Mercado |
| EPI_ISL_8357466 | LSP Distrital Bogota | LSPSDS | Alejandro Gomez Lopez; Gabriela Delgado Murcia; Johana Hernandez Toloza; Marcela Mercado |
| EPI_ISL_6769061 | Lab. Virología y Genética Universidad Simón Bolívar | Laboratorio de Biologia Molecular, Universidad Cooperativa de Colombia, Santa Marta | Andrew S. Muñoz-Gamba; Antonio Acosta; Camila Gonzalez; Daniel B. Ramírez-Osorio; Johana Zuñiga; José A. Usme-Ciro; Paula A. Quintero-Cortés; Yesid Bello |
| EPI_ISL_8178768 | Laboratoire de Recherche et d'Analyses Médicales de la Gendarmerie Royale | Laboratoire de Recherche et d'Analyses Médicales de la Gendarmerie Royale | Amal SOUIRI; Hajar LEMRISS; Mohamed LABIOUI; Mohamed Mouatakid; Nabil LEMZAOUI; Sanaâ LEMRISS; Saâd EL KABBAJ |
| EPI_ISL_7662410, EPI_ISL_7662503, EPI_ISL_7662506, EPI_ISL_7664072, EPI_ISL_7664074, EPI_ISL_7664076, EPI_ISL_7664078 | | | |
| see above | Laboratoire de santé publique du Québec | Laboratoire de santé publique du Québec | Guillaume Bourque; Ioannis Ragoussis; Jesse Shapiro; Mark Lathrop and Judith Fafard on behalf of the CoVSeQ research group; Sandrine Moreira |
| EPI_ISL_8408362, EPI_ISL_8408363, EPI_ISL_8408365, EPI_ISL_8408370, EPI_ISL_8408371, EPI_ISL_8408373 | Laboratorio Central, Ministerio de Salud Córdoba | Fundacion para el Progreso de la Medicina and Instituto de Patologia Vegetal (IPAVE-CIAP-INTA) on behalf of 'Proyecto Argentino Interinstitucional de genomica de SARS-CoV-2' (PAIS Consortium) | Barbas, G.; Castro, G.; Debat, H.; F.D.; Fernández; Goya, S.; Lucca, A.; M.B.; Marquez, N.; Pisano; Re, V.; Sicilia, P.; Zeballos, M. |
| EPI_ISL_7961381, EPI_ISL_7961386 | Laboratorio Referencial Regional en Salud Pública - GERESA Lambayeque | Laboratorio de Genómica Microbiana, Universidad Peruana Cayetano Heredia | Alejandra Dávila-Barclay; Cristian Díaz Vélez; Diego Cuicapuza; Guillermo Salvatierra; Gustavo Adolfo Sandoval Peña; Janet Huancachoque; Luis Miguel Serquén Lopez; Pablo Tsukayama; Richard Montalvo Aguirre; Ronald Milian Pérez; Sergio Luis Aguilar Martínez |
| EPI_ISL_8217527 | Laboratorio SYNLAB Colombia | "Laboratorio de biotecnología, Universidad Icesi" | "María I. Gutiérrez López; Adrián Camilo Rodríguez Ararat; Diana M. Florez Giraldo; Marcela Mercado; Maria F. Villegas Torres; Paola A. Caicedo Burbano"; Programa Nacional de Caracterización Genómica de SARS-CoV-2 |
| EPI_ISL_6703229, EPI_ISL_6703248 | Laboratorio SYNLAB Colombia | CIAT, Laboratorio de Virologia | Ana M. Leiva; Diana Lopez-Alvarez; Programa Nacional de Caracterización Genómica de SARS-CoV-2; WilmerJ. Cuellar |
| EPI_ISL_7961384, EPI_ISL_7961385 | Laboratorio de Genómica Microbiana - Universidad Peruana Cayetano Heredia | Laboratorio de Genómica Microbiana, Universidad Peruana Cayetano Heredia | Brenes Hebleen; Camacho Erwin; Campos-Sánchez Rebeca; Ceballos Ana; Cordero Estela; Cristancho Marco; Diego Cuicapuza; Duarte Francisco; Fernández-Do Porto Darío; Guillermo Salvatierra; Herrera-Estrella Alfredo; Janet Huancachoque; Jiménez-Moraila Beatriz; Kreuze Jan; Mireles-Rivera Guadalupe; Molina-Mora José; Muñoz-Medina José Esteban; Negri Tatiana; Nunes Gisele; Oliveira Guilherme; Oliveira Renato; Pedro E. Romero; Reales-González Jhonnatan; Remes-Lenicov Federico; Reyes Alejandro; Segundo Fuentes; Sosa Ezequiel; Soto Claudio; Tsukayama Pablo; Turjanski Adrián; and Zayat Jonathan / Alejandra Dávila-Barclay |
| EPI_ISL_6574179, EPI_ISL_6574229, EPI_ISL_6574318, EPI_ISL_6689772, EPI_ISL_6689773, EPI_ISL_6689774, EPI_ISL_6689775, EPI_ISL_6689842, EPI_ISL_6946002, EPI_ISL_6946040, EPI_ISL_6946056, EPI_ISL_7494925, EPI_ISL_7494926, EPI_ISL_7494931, EPI_ISL_7494932, EPI_ISL_7494933, EPI_ISL_7494934, EPI_ISL_7494935, EPI_ISL_7494936, EPI_ISL_7494937, EPI_ISL_7846229, EPI_ISL_7846327, EPI_ISL_7846353, EPI_ISL_8143835 | | | |
| see above | Laboratorio de Referencia Nacional de Virus Respiratorios. Centro Nacional de Salud Publica. Instituto Nacional de Salud Peru. | Laboratorio de Referencia Nacional de Virus Respiratorios. Centro Nacional de Salud Publica. Instituto Nacional de Salud Peru. | Alicia Nuñez Llanos; Carlos Padilla Rojas; Edwart Rogger Rivera Serrano; Henri Bailon Calderon; Iris Silva Molina; Joseph Huayra Niquen; Kelly Vanessa Izarra Rojas; Lely Solari Zerpa; Luis Barcena Flores; Marco Galarza Perez; Nancy Rojas Serrano; Nieves Sevilla Castañeda; Omar Caceres Rey; Orson Mestanza Millones; Princesa Medrano Alhuay; Priscila Lope Pari; Sara Gordillo Vilchez; Steve Acedo Lazo; Veronica Hurtado Vela; Victor Jimenez Vasquez; Wendy Lizarraga Olivares |
| EPI_ISL_7747330, EPI_ISL_7747336, EPI_ISL_7747351, EPI_ISL_7747353, EPI_ISL_7747355, EPI_ISL_7747361, EPI_ISL_7747363, EPI_ISL_7747364, EPI_ISL_7747366, EPI_ISL_7747370, EPI_ISL_7747372, EPI_ISL_7747374, EPI_ISL_7747377, EPI_ISL_7747381, EPI_ISL_7747385, EPI_ISL_7747386, EPI_ISL_7747389, EPI_ISL_7747391, EPI_ISL_7747392, EPI_ISL_7747394, EPI_ISL_7747418, EPI_ISL_7747440, EPI_ISL_7835287, EPI_ISL_7835297 | | | |
| see above | Laboratorio de Salud Pública de Bogotá | Gencore - Universidad de los Andes | Alejandro Gomez; Cristian Barrera; Gabriela Ariza; Gabriela Delgado; Johana Hernandez; Luisa Sacristan; Marcela Guevara; Silvia Restrepo |
| EPI_ISL_7719094 | Laboratorio di Microbiologia e Virologia, Università Vita-Salute San Raffaele, Milano | Laboratorio di Microbiologia e Virologia, Università Vita-Salute San Raffaele, Milano | Elena Criscuolo; Enzo Boeri; Massimo Clementi; Massimo Locatelli; Matteo Castelli; Michela Sampaolo; Nicasio Mancini; Nicola Clementi; Roberta Antonia Diotti; Roberto Ferrarese |
| EPI_ISL_8264091, EPI_ISL_8367942, EPI_ISL_8469117, EPI_ISL_8469736 | Laboratory Corporation of America | Centers for Disease Control and Prevention Division of Viral Diseases, Pathogen Discovery | Amanda Douglas; Amanda Suchanek; Andrea Throop; Ayla Burns; Benjamin Rambo-Martin; Bobbi Croy; Brian Krueger; Brian Norvell; Christopher Gulvick; Christos Petropoulos; Clinton Paden; Craig Lukasik; Dakota Howard; Debbie Boles; Dhwani Batra; Duncan MacCannell; Eyad Almasri; Goran Stevovic; Howard Engler; Hrushikesh Deshmukh; Jake Humphrey; Jana Schroth; Jason Caravas; Joe Voshell; John Pruitt; Jonathan Meltzer; Jonathan Williams; Kimberly Wagner; Kristine Lacek; Lax Iyer; Lisa Pfefferle; Lyndon Tilson; Manoj Jain; Marcia Eisenberg; Mary Cristobal; Mary Williamson; Matthew Robinson; Matthew Schmerer; Michael Levandoski; Mike Sapeta; Mindy Nye; Minoo Agarwal; Mohan Kolli; Nuthawin Charoensri; Oren Cohen; Peter Cook; Prashant Gupta; Qian Zeng; Rama Ghatti; Scott Parker; Scott Ryan; Scott Sammons; Shatavia Morrison; Stanley Letovsky; Steven Ragan; Suresh Selvaraju; Susan Countryman; Susan Hicks; Suzanne Dale; Thomas Urban; Tim Kuphal; Tricia Zwiefelhofer; Tymeckia Kendall; Victoria Caban Figueroa; Vincent Drouillon; Yvette Unoarumhi |
| EPI_ISL_7613697 | Maasstad Ziekenhuis | Maasstad ziekenhuis | Pieter Smit |
| EPI_ISL_7462685 | Molecular Virology Unit, Microbiology and Virology Depertment, Fondazione IRCCS Policlinico San Matteo, Pavia | Molecular Virology Unit, Microbiology and Virology Depertment, Fondazione IRCCS Policlinico San Matteo, Pavia | Antonio Piralla; Fausto Baldanti; Federica Giardina; Guglielmo Ferrari; Stefano Gaiarsa |
| EPI_ISL_7711154 | NJDOH, Public Health and Environmental Laboratories | NJ_PHEL | Allison Roder; Byeong Jeong; Chelsea San Filippo; Dana Woell; Jacquelyn Deverell; Lindsey Bodnar; Maria-Magdalene Pugliese; Mohammad M. Ali; Ryan Pachucki; Shiv K. Verma |
| EPI_ISL_7263474 | National Platform bis COVID ULB-IBC | National Platform bis COVID ULB-IBC | Arnaud Marchant; Benoit Haerlingen; Coralie Henin; Marie-Luce Delforge; Ricardo De Mendonça |
| EPI_ISL_6631981, EPI_ISL_6632036, EPI_ISL_6632044, EPI_ISL_6632077, EPI_ISL_6632100, EPI_ISL_6632101, EPI_ISL_6632107, EPI_ISL_6632112, EPI_ISL_6632116, EPI_ISL_6632119, EPI_ISL_6632127, EPI_ISL_6632131, EPI_ISL_6632132, EPI_ISL_6632133, EPI_ISL_6632134, EPI_ISL_6632135, EPI_ISL_6632142, EPI_ISL_6632149, EPI_ISL_6632161, EPI_ISL_6632166, EPI_ISL_6632172, EPI_ISL_6632179, EPI_ISL_6632185 | | | |
| see above | National Public Health Laboratory, Ministry of Health & Wellness | Viral Special Pathogens Branch, Centers for Disease Control and | Antoni Comrie; Carl Bruce; Emir Talundzic; Joel Montgomery; John Klena; Justin Lee; Karen Webster-Kerr; Melissa Mobley; Michelle Brown; Michelle Hamilton; Monica Smikle; Nathlee McMorris; Sandra Jackson; Shannon Whitmer; Suwanie Lewis |

| | | | |
|---|---|---|---|
| | | Prevention | |
| EPI_ISL_8133617 | Northumbria University / South Tees Hospitals NHS Foundation Trust / North Cumbria Integrated Care NHS Foundation Trust / North Tees and Hartlepool NHS Foundation Trust / Newcastle Hospitals NHS Foundation Trust | COVID-19 Genomics UK (COG-UK) Consortium | Andrew Nelson; Brendan Payne; Clive Graham; Darren L Smith; Debra Padgett; Edward Barton; Emma Swindells; Garren Scott; Gary Black; Gary Eltringham; Giles S Holt; Greg R Young; Jane Greenaway; Jennifer Collins; John Allan; Joshua Loh; Lynn Dover; Matthew Bashton; Mohammad A Tariq; Paul Baker; Sarah Essex; Steve Liggett; Wen C Yew; Yusri Taha |
| EPI_ISL_7623977 | OHSU Lab Services Molecular Microbiology Lab | Oregon SARS-CoV-2 Genome Sequencing Center | Alec J. Hirsch; Andrew C. Adey; Benjamin N. Bimber; Brendan L. O'Connell; Brian J. O'Roak; Cierra LaBlanc; Daniel N. Streblow; Destine Krenik; Donna Hansel; Guang Fan; Rabeka Ali; Ruth V. Nichols; Sonia Acharya; William B. Messer; Xuan Qin |
| EPI_ISL_7602560, EPI_ISL_7851219, EPI_ISL_8094966 | Pandemic Response Lab - NYC | Pandemic Response Lab, R&D | Alex Carpio; Cybill del Castillo; Dylan Law; Haiping Hao; Henry Lee; Isabel Fernandez Escapa; Jon Laurent; Melissa Hopkins; Michael Hammerling; Pradeep Bugga; Shinyoung Clair Kang; Sol Rey; William Ward |
| EPI_ISL_8323023, EPI_ISL_8323077 | Public Health Ontario Laboratory | Public Health Ontario Laboratory | Aimin Li; Alireza Eshaghi; Andre Villegas; Ashleigh Sullivan; Christine Frantz; Dean Maxwell; Esha Joshi; Jared Simpson; Jennifer L Guthrie; Jonathan B Gubbay; Karthikeyan Sivaraman; Lawrence Heisler; Matthew Watson; Michael CY Li; Michael Laszloffy; Nahuel Fittipaldi; Philip Banh; Richard de Borja; Samir N Patel; Sandeep Nagra; Sandra Zittermann; Sarah Teatero; Vanessa G Allen; Yao Chen; Yogi Sundaravadanam |
| EPI_ISL_8044419 | Quadram Institute Bioscience | COVID-19 Genomics UK (COG-UK) Consortium | Alexander J Trotter; Alison E. Mather; Alp Aydin; Ana P. Tedim; Anastasia Kolyva; Andrew Bell; Andrew J. Page; Christopher Jeanes; Claire Stuart; Dave J. Baker; Ebenezer Foster-Nyarko; Gemma L. Kay; John Wain; Justin O'Grady; Leonardo de Oliveira Martins; Lewis G. Spurgin; Lindsay Coupland; Lizzie Meadows; Luke Bedford; Maria Diaz; Mark Webber; Martin Lott; Michaela Matthews; Muhammed Yasir; Nabil-Fareed Alikhan; Ngozi Elumogo; Nicholas M. Thomson; Rachael Stanley; Rachel Gilroy; Reenesh Prakash; Rose K Davidson; Samir Dervisevic; Samuel Bloomfield; Sophie J. Prosolek; Steven Rudder; Thanh Le-Viet |
| EPI_ISL_6783400, EPI_ISL_7936987 | Quest Diagnostics Incorporated | Centers for Disease Control and Prevention Division of Viral Diseases, Pathogen Discovery | A. Gerasimova; A. Perez; B. Anderson; Benjamin Rambo-Martin; Christopher Gulvick; Clinton Paden; Dakota Howard; Dhwani Batra; Duncan MacCannell; Erisa Sula; F. Lacbawan; I. Shlyakhter; Jason Caravas; K. Livingston; Kristine Lacek; L. Bernstein; M. Hua; Matthew Schmerer; P. Tanpaiboon; Peter Cook; R. Kagan; R. Owen; R. Rolando; S. Rosenthal; Scott Sammons; Shatavia Morrison; Tymeckia Kendall; Victoria Caban Figueroa; Y. Liu; Yvette Unoarumhi |
| EPI_ISL_7166193 | Respiratory Virus Unit, Microbiology Services Colindale, Public Health England | COVID-19 Genomics UK (COG-UK) Consortium | PHE Covid Sequencing Team |
| EPI_ISL_7979718 | SURA | Gencore - Universidad de los Andes | Cristian Barrera; David González; Gabriela Ariza; Luisa Sacristan; Marcela Guevara; Marcela Mercado; Silvia Restrepo |
| EPI_ISL_6811664 | Salud Digna | Instituto Nacional de Medicina Genomica | Abraham Campos-Romero; Cedro-Tanda A; Cruz-Islas Jazmin; Escobar-Arrazola MA; Garnica-Lopez Dora; Herrera-Montalvo LA.; Hidalgo-Miranda A; Luna-Ruiz Marco; Mendoza-Vargas A; Moreno-Camacho José Luis; Ramirez-Vega O; Rangel-DeLeon D; Reyes-Grajeda JP; Rodriguez-Gallegos Jorge; Yair Alfaro-Mora |
| EPI_ISL_7751221, EPI_ISL_7751429, EPI_ISL_7751639, EPI_ISL_7753089, EPI_ISL_7753141, EPI_ISL_7753763 | Servicio Virosis Respiratorias-Departamento Virologia-INEI | Instituto Nacional Enfermedades Infecciosas C.G.Malbran | Avaro M.; Baumeister E.; Benedetti E.; Campos J.; Cisterna D.; Dattero ME; De Belder D.; Haim MS.; Lorenzo F.; Molina V.; Perandones C.; Poklepovich T.; Pontoriero A.; Russo M.; Sanchez Loria J.; Tuduri E. |
| EPI_ISL_6694518, EPI_ISL_6694533, EPI_ISL_6694694, EPI_ISL_6694854, EPI_ISL_6694858, EPI_ISL_6694884, EPI_ISL_6694888, EPI_ISL_6694956, EPI_ISL_6695030, EPI_ISL_6695103 | | | |
| see above | Servicio Virosis Respiratorias-Departamento Virología-INEI | Instituto Nacional Enfermedades Infecciosas C.G.Malbran | Avaro M.; Baumeister E.; Benedetti E.; Campos J.; Cisterna D.; Dattero ME; De Belder D.; Haim MS.; Lorenzo F.; Molina V.; Perandones C.; Poklepovich T.; Pontoriero A.; Russo M.; Sanchez Loria J.; Tuduri E. |
| EPI_ISL_7710663, EPI_ISL_7710667, EPI_ISL_7938382, EPI_ISL_7938405 | Servicios Medicos Olimpus | Laboratorio de Biologia Molecular, Universidad Cooperativa de Colombia, Santa Marta | Andrew S. Muñoz-Gamba; Daniel B. Ramírez-Osorio; José A. Usme-Ciro; Mayerlin Vasquez Cardenas; Paula A. Quintero-Cortés; Pricelis Paulin Polanco Fontalvo |
| EPI_ISL_6941451, EPI_ISL_6941452, EPI_ISL_6941460, EPI_ISL_6941461, EPI_ISL_6941462, EPI_ISL_6941463, EPI_ISL_6941464, EPI_ISL_6941465, EPI_ISL_6941466, EPI_ISL_6941467, EPI_ISL_6941468, EPI_ISL_6941469, EPI_ISL_6941470, EPI_ISL_6941471, EPI_ISL_6941472, EPI_ISL_6941493, EPI_ISL_6941494, EPI_ISL_6941495, EPI_ISL_6941496, EPI_ISL_6941497, EPI_ISL_6941498, EPI_ISL_6941499, EPI_ISL_6941501, EPI_ISL_6941502, EPI_ISL_6941503, EPI_ISL_6941506 | | | |
| see above | The Caribbean Public Health Agency | Carrington Lab, Department of Preclinical Sciences, Faculty of Medical Sciences, The University of the West Indies, St Augustine Campus | Anushka Ramjag; Arianne Brown-Jordan; Avery Hinds; Christine V. F. Carrington; Christopher Oura; Gabriel Escobar; Irad Potter; Jacquiline Bisesor-McKenzie; Nikita S. D. Sahadeo; Nuno Faria; Oliver Pybus; Risha Singh; Sarah Hill; Simone Keizer-Beache; Soren Nicholls; SueMin Nathaniel; Vernie Ramkissoon |

We gratefully acknowledge the following Authors from the Originating laboratories responsible for obtaining the specimens, as well as the Submitting laboratories where the genome data were generated and shared via GISAID, on which this research is based.

All Submitters of data may be contacted directly via www.gisaid.org

Authors are sorted alphabetically.

Acknowledgement EPI_SET Identifier: EPI_SET_20220328wx

| Accession ID | Originating Laboratory | Submitting Laboratory | Authors |
|---|---|---|---|
| EPI_ISL_2621218 | ADILAB | Universidad Nacional de Colombia - Laboratorio Genómico One Health | Andres F. Cardona-Rios; Carlos Franco-Muñoz; Carolina Muñoz-Arango; Celeny Ortiz; Daniel O. Maldonado-Perez; Diego A. Álvarez-Díaz; Hector Alejandro Ruiz-Moreno; Idabely Betancur Ortiz; Jorge E. Osorio; Juan P. Hernandez-Ortiz; Karl A Ciuoderis; Katherine Laiton-Donato; Laura Silvana Perez; Lina M. Hurtado; Marcela Mercado-Reyes; Maria Angélica Maya; Maria Stella López; Rita Almanza Payares; Sandra Ines Cano; Simón Villegas Velásquez |
| EPI_ISL_2828007, EPI_ISL_2828008 | AREA DE SALUD SAN RAMON | Inciensa, Instituto Costarricense de Investigación y Enseñanza en Nutrición y Salud | Adriana Godínez; Claudio Soto-Garita; Estela Cordero; Francisco Duarte; Hebleen Porras; José Luis Vargas; Mariela Gutiérrez & Joselyn Prado; Melany Calderón |
| EPI_ISL_2975142, EPI_ISL_2975350, EPI_ISL_2975351, EPI_ISL_2975352, EPI_ISL_2975353 | AULSS 6 Euganea | Istituto Zooprofilattico Sperimentale delle Venezie | Adelaide Milani; Alessia Schivo; Alice Fusaro; Ambra Pastori; Annalisa Salviato; Antonia Ricci; Calogero Terregino; Edoardo Giussani; Elisa Palumbo; Erika Giorgia Quaranta; Isabella Monne; Luca Tassoni |
| EPI_ISL_2927986 | AULSS 7 Pedemontana | Istituto Zooprofilattico Sperimentale delle Venezie | Adelaide Milani; Alessia Schivo; Alice Fusaro; Ambra Pastori; Annalisa Salviato; Antonia Ricci; Calogero Terregino; Edoardo Giussani; Elisa Palumbo; Erika Giorgia Quaranta; Isabella Monne; Luca Tassoni |
| EPI_ISL_2975200 | AULSS 8 Berica | Istituto Zooprofilattico Sperimentale delle Venezie | Adelaide Milani; Alessia Schivo; Alice Fusaro; Ambra Pastori; Annalisa Salviato; Antonia Ricci; Calogero Terregino; Edoardo Giussani; Elisa Palumbo; Erika Giorgia Quaranta; Isabella Monne; Luca Tassoni |
| EPI_ISL_2651228 | AYUDAS DIAGNOSTICAS SURA | Universidad Nacional de Colombia - Laboratorio Genómico One Health | Andres F. Cardona-Rios; Carlos Franco-Muñoz; Carolina Muñoz-Arango; Celeny Ortiz; Daniel O. Maldonado-Perez; Diego A. Álvarez-Díaz; Hector Alejandro Ruiz-Moreno; Idabely Betancur Ortiz; Jorge E. Osorio; Juan P. Hernandez-Ortiz; Karl A Ciuoderis; Katherine Laiton-Donato; Laura Silvana Perez; Lina M. Hurtado; Marcela Mercado-Reyes; Maria Angélica Maya; Maria Stella López; Rita Almanza Payares; Sandra Ines Cano; Simón Villegas Velásquez |
| EPI_ISL_2438080 | Adilab | Universidad Nacional de Colombia - Laboratorio Genómico One Health | Andres F. Cardona-Rios; Carlos Franco-Muñoz; Carolina Muñoz-Arango; Celeny Ortiz; Daniel O. Maldonado-Perez; Diego A. Álvarez-Díaz; Hector Alejandro Ruiz-Moreno; Idabely Betancur Ortiz; Jorge E. Osorio; Juan P. Hernandez-Ortiz; Karl A Ciuoderis; Katherine Laiton-Donato; Laura Silvana Perez; Lina M. Hurtado; Marcela Mercado-Reyes; Maria Angélica Maya; Maria Stella López; Rita Almanza Payares; Sandra Ines Cano; Simón Villegas Velásquez |
| EPI_ISL_1562674 | Aegis Sciences Corporation | Centers for Disease Control and Prevention Division of Viral Diseases, Pathogen Discovery | Adrian Paskey; Alec Vest; Benjamin Rambo-Martin; Christopher Gulvick; Clinton R. Paden; Cyndi Clark; Dakota Howard; Darlene Wagner; Dhwani Batra; Dillon Nall; Duncan MacCannell; Ethan Sanders; Holly Houdeshell; Jason Caravas; Kara Moser; Matthew Hardison; Matthew Schmerer; Ola Kvalvaag; Patrick Campbell; Peter W. Cook; Rob Case; Scott Sammons; Shatavia Morrison; Shaun Westlund; Vikramsinha Ghorpade; Yvette Unoarumhi |
| EPI_ISL_2694880 | Affidea | Instituto Nacional de Saude (INSA) | Borges et al |
| EPI_ISL_2158266, EPI_ISL_2473689, EPI_ISL_2504002, EPI_ISL_2969595 | Alaska State Virology Laboratory | Alaska State Virology Laboratory | Elva House; Jack Chen; Jacob Zidek; Lisa Smith; Ph.D.; Stephanie DeRonde |
| EPI_ISL_2086307 | Arizona State Public Health Laboratory | Arizona State Public Health Laboratory | Jessica Escobar; Katherine Fullerton; Linda Getsinger; Nobuko Fukushima; Stacy White; Trung Huynh; Victor Waddell |
| EPI_ISL_2617632, EPI_ISL_2617633, EPI_ISL_2617634, EPI_ISL_2617635, EPI_ISL_2617636, EPI_ISL_2617637, EPI_ISL_2617638, EPI_ISL_2617639, EPI_ISL_2617640, EPI_ISL_2617641, EPI_ISL_2617642, EPI_ISL_2617643, EPI_ISL_2617644, EPI_ISL_2617645, EPI_ISL_2617646, EPI_ISL_2617647, EPI_ISL_2617648, EPI_ISL_2617649, EPI_ISL_2617650, EPI_ISL_2617651, EPI_ISL_2617652, EPI_ISL_2617653, EPI_ISL_2617654, EPI_ISL_2617655, EPI_ISL_2617656, EPI_ISL_2617657, EPI_ISL_2617658, EPI_ISL_2617659 | | | |
| see above | Austrian Agency for Health and Food Safety (AGES) | Bergthaler laboratory, CeMM Research Center for Molecular Medicine of the Austrian Academy of Sciences | Andreas Bergthaler; Anna Schedl; Bekir Erguner; Benedikt Agerer; Christoph Bock; Fabian Amman; Jan Laine; Lukas Endler; Maelle Le Moing; Martin Senekowitsch; Matthew Thornton; Michael Schuster; Petr Triska; Thomas Penz |
| EPI_ISL_2438009 | Ayudas diagnosticas SURA | Universidad Nacional de Colombia - Laboratorio Genómico One Health | Andres F. Cardona-Rios; Carlos Franco-Muñoz; Carolina Muñoz-Arango; Celeny Ortiz; Daniel O. Maldonado-Perez; Diego A. Álvarez-Díaz; Hector Alejandro Ruiz-Moreno; Idabely Betancur Ortiz; Jorge E. Osorio; Juan P. Hernandez-Ortiz; Karl A Ciuoderis; Katherine Laiton-Donato; Laura Silvana Perez; Lina M. Hurtado; Marcela Mercado-Reyes; Maria Angélica Maya; Maria Stella López; Rita Almanza Payares; Sandra Ines Cano; Simón Villegas Velásquez |
| EPI_ISL_2872589, EPI_ISL_2872590 | Azienda Ospedaliero Universitaria di Sassari | AMES Centro Polidiagnostico Strumentale S.r.l. | Anna Puggioni; Bianca Paglietti; Caterina Serra; Claudia Piu; Elena Rimini; Erica Mura; Flavia Angioj; Gabriele Ibba; Giulia Rocca; Laura Firino; Rosalba Govoni; Salvatore Rubino; Sergio Uzzau; Vincenzo Lai |
| EPI_ISL_2372852, EPI_ISL_2550664, EPI_ISL_2550696 | Azienda Sanitaria dell'Alto Adige Laboratorio Aziendale di Microbiologia e Virologia | Istituto di Genomica Applicata | Davide Scaglione; Eleonora Paparelli; Elisa Masi; Elisabetta Giacobazzi; Elisabetta Pagani; Gabriele Magris; Irena Jurman; Irene Bianconi; Michele Morgante; Stefanie Wieser; Vera Vendramin |
| EPI_ISL_431154 | BIOMNIS EUROFINS IVRY | Department of Virology, Henri Mondor University Hospital, Assistance Publique Hôpitaux de Paris, Université Paris-Est Créteil, INSERM U955 | Alexandre Soulier; Christophe Rodriguez; Elisabeth Trawinski; Guillaume Gricourt; Jean-Michel Pawlotsky; Melissa N'Debi; Slim Fourati; Vanessa Demontant |
| EPI_ISL_2321586 | BIOREFERENCIA | Universidad Nacional de Colombia - Laboratorio Genómico One Health | Andres F. Cardona-Rios; Carlos Franco-Muñoz; Carolina Muñoz-Arango; Celeny Ortiz; Daniel O. Maldonado-Perez; Diego A. Álvarez-Díaz; Hector Alejandro Ruiz-Moreno; Idabely Betancur Ortiz; Jorge E. Osorio; Juan P. Hernandez-Ortiz; Karl A Ciuoderis; Katherine Laiton-Donato; Laura Silvana Perez; Lina M. Hurtado; Marcela Mercado-Reyes; Maria Angélica Maya; Maria Stella López; Rita Almanza Payares; Sandra Ines Cano; Simón Villegas Velásquez |
| EPI_ISL_2999927 | Berkshire and Surrey Pathology Services Lighthouse Laboratory | Wellcome Sanger Institute for the COVID-19 Genomics UK (COG-UK) Consortium | Berkshire and Surrey Pathology Services Lighthouse Laboratory and Alex Alderton; Cordelia Langford; David K. Jackson; Dominic Kwiatkowski; Ewan Harrison; Ian Johnston; Jeffrey Barrett; John Sillitoe on behalf of the Wellcome Sanger Institute COVID-19 Surveillance Team; Roberto Amato; Sonia Goncalves |
| EPI_ISL_2536144 | Bundeswehrkrankenhaus Ulm | Bundeswehr Institute of Microbiology | Alexandra Rehn; Enrico Georgi; Malena Bestehorn-Willmann; Markus Antwerpen; Mathias Walter; Mike Pillukat; Roman Wölfel; Sabine Zange |
| EPI_ISL_2621207 | CENTROLAB | Universidad Nacional de Colombia - Laboratorio Genómico One Health | Andres F. Cardona-Rios; Carlos Franco-Muñoz; Carolina Muñoz-Arango; Celeny Ortiz; Daniel O. Maldonado-Perez; Diego A. Álvarez-Díaz; Hector Alejandro Ruiz-Moreno; Idabely Betancur Ortiz; Jorge E. Osorio; Juan P. Hernandez-Ortiz; Karl A Ciuoderis; Katherine Laiton-Donato; Laura Silvana Perez; Lina M. Hurtado; Marcela Mercado-Reyes; Maria Angélica Maya; Maria Stella López; Rita Almanza Payares; Sandra Ines Cano; Simón Villegas Velásquez |
| EPI_ISL_2695346 | CHEDV | Instituto Nacional de Saude (INSA) | Borges et al |
| EPI_ISL_2796193 | CHU Sao Joao, Porto | Instituto Nacional de Saude (INSA) | Borges et al |
| EPI_ISL_2835967 | CHUGA-IBP-laboratoire de Virologie | IBP-laboratoire de virologie | Alban Caporossi; Anne Signori -Schmuck; Anne-Karen Faure; Aurélie Truffot; Benjamin Nemoz; Hugo Jardin; Julien Andréani; Julien Lupo; Léa Ponderand; Pascal Poignard; Patrice Morand; Raphaële Germi; Sylvie Larrat |
| EPI_ISL_2695100, EPI_ISL_2695101, EPI_ISL_2695102 | CHULN - H Santa Maria | Instituto Nacional de Saude (INSA) | Borges et al |
| EPI_ISL_2500506, EPI_ISL_2500516, EPI_ISL_2622106 | CHUV | Laboratory of genomics and metagenomics | Claire Bertelli; Damien Jacot; Gilbert Greub; Sébastien Aeby; Trestan Pillonel |
| EPI_ISL_3008830 | Center of Advanced Studies and Technology, Molecular Genetics Laboratory | Center of Advanced Studies and Technology, Molecular Genetics Laboratory | Anaclerio Federico; De Fabritiis Simone |
| EPI_ISL_1970836, EPI_ISL_1970837 | Centro de Investigación en Enfermedades Infecciosas (CIENI), Instituto Nacional de Enfermedades Respiratorias (INER) | Centro de Investigación en Enfermedades Infecciosas (CIENI), Instituto Nacional de Enfermedades Respiratorias (INER) | Alejandra Hernández-Terán; Alejandro Sanchez-Flores; Blanca Taboada; Carlos F. Arias; Celia Boukadida; Eduardo Becerril-Vargas; Fidencio Mejía-Nepomuceno; Francisco Pulido; Hector Esteban Paz-Juárez; Joel Armando Vázquez-Pérez and Consorcio Mexicano de Vigilancia Genómica (CoViGen-Mex); Jorge Salas-Hernández; José Arturo Martínez-Orozco; Margarita Matías-Florentino; Mario Mújica-Sánchez; Pavel Isa; Ricardo Grande; Santiago Ávila-Ríos |
| EPI_ISL_2434137, EPI_ISL_2927083, EPI_ISL_2966323 | Cliniques universitaires Saint-Luc | UCLouvain/IREC/MBLG | Benoit Kabamba Mukadi; Bertrand Bearzatto; Jean Ruelle; Ophélie Simon |
| EPI_ISL_2784135 | Curative Labs | Curative Labs | Elias L. Salfati; Eugenia Khorosheva; George Way; J.Cesar Ignacio-Espinoza; Janet Chen; Mikhail Hanewich-Hollatz; Nabjot Sandhu; Sophia Quasem; Vladimir Slepnev; Zhiyi Xie |
| EPI_ISL_1678824 | Department of Medical Microbiology & Infection prevention, Amsterdam University Medical Centers location AMC | Department of Medical Microbiology & Infection prevention, Amsterdam University Medical Centers location AMC | Fokla Zorgdrager; Janke Schinkel; Marcel Jonges; Matthijs Welkers; Menno de Jong; Robin van Houdt; Sebastien Matamoros; Sjoerd Rebers |
| EPI_ISL_1868200, EPI_ISL_1890559, EPI_ISL_2025772, | Department of Virus and Microbiological Special Diagnostics, Statens Serum | Aalborg University | Danish Covid-19 Genome Consortium |

| | | | |
|---|---|---|---|
| EPI_ISL_2296939, EPI_ISL_2298986 | Institut, Copenhagen, Denmark | | |
| EPI_ISL_2694746 | Diagnostyka. Laboratoria Medyczne. | 1. ViroGenetics - BSL3 Laboratory of Virology, Małopolska Centre of Biotechnology, Jagiellonian University; 2. Diagtron Laboratoria Łukasz Rąbalski | Gromowski, T.; Kowalski, M.; Labaj; Maciej Kosinski; Mazur-Panasiuk, N.; Natalia Derewonko; P.P.; Pyrc, K.; Rabalski L.; Rogalska-Kupiec M.; Swadzba J.; Sylwia Januszczak; Szulc, P.; Wydmanski, W. |
| EPI_ISL_2955372 | Dianalabs SA | Genesupport | Geraldine Jost; Katia Jaton; Nadia Liassine; Tanguy ARAUD |
| EPI_ISL_2613602, EPI_ISL_2613603, EPI_ISL_2613604, EPI_ISL_2613605, EPI_ISL_2613606, EPI_ISL_2613607, EPI_ISL_2613608, EPI_ISL_2613609, EPI_ISL_2613610, EPI_ISL_2613611, EPI_ISL_2966247 | | | |
| see above | Dipartimento di Medicina di Laboratorio, Azienda sanitaria universitaria Friuli Centrale (ASU FC) | Dipartimento di Medicina di Laboratorio, Azienda sanitaria universitaria Friuli Centrale (ASU FC) | Catia Mio; Chiara Dal Secco; Corrado Pipan; Francesco Curcio; Stefania Marzinotto |
| EPI_ISL_1970344, EPI_ISL_1970345, EPI_ISL_1970346, EPI_ISL_2483545, EPI_ISL_2483546, EPI_ISL_2483547, EPI_ISL_2887845 | | | |
| see above | Dutch COVID-19 response team | Erasmus Medical Center | Anne van der Linden; Annemiek van der Eijk; Bas Oude Munnink; Corine GeurtsvanKessel; David Nieuwenhuijse; Emmanuelle Munger; Irina Chestakova; Marion Koopmans; Marjan Boter; Reina Sikkema; Richard Molenkamp; on behalf of the Dutch national COVID-19 respo; on behalf of the Dutch national COVID-19 response team. |
| EPI_ISL_1962951, EPI_ISL_1962952, EPI_ISL_2094359, EPI_ISL_2094360, EPI_ISL_2094361, EPI_ISL_2094362, EPI_ISL_2094363, EPI_ISL_2094364, EPI_ISL_2094365, EPI_ISL_2094366, EPI_ISL_2094367, EPI_ISL_2094371, EPI_ISL_2220450, EPI_ISL_2220453, EPI_ISL_2220456, EPI_ISL_2220458, EPI_ISL_2220460, EPI_ISL_2220461, EPI_ISL_2303941, EPI_ISL_2303942, EPI_ISL_2303943, EPI_ISL_2303944, EPI_ISL_2303945, EPI_ISL_2303946, EPI_ISL_2303947, EPI_ISL_2405403, EPI_ISL_2405469, EPI_ISL_2405492, EPI_ISL_2405505, EPI_ISL_2405516, EPI_ISL_2405527, EPI_ISL_2405550, EPI_ISL_2405596, EPI_ISL_2406319, EPI_ISL_2406427, EPI_ISL_2476271, EPI_ISL_2476273, EPI_ISL_2476276, EPI_ISL_2609616, EPI_ISL_2609687, EPI_ISL_2610006, EPI_ISL_2610152, EPI_ISL_2610357, EPI_ISL_2610368, EPI_ISL_2610471, EPI_ISL_2610607, EPI_ISL_2610618, EPI_ISL_2610630, EPI_ISL_2610681, EPI_ISL_2610905, EPI_ISL_2672446, EPI_ISL_2672828, EPI_ISL_2672859, EPI_ISL_2672922, EPI_ISL_2673226, EPI_ISL_2673250, EPI_ISL_2673263, EPI_ISL_2673286, EPI_ISL_2788181, EPI_ISL_2788182, EPI_ISL_2788183, EPI_ISL_2788185, EPI_ISL_2788186, EPI_ISL_2788187, EPI_ISL_2788188, EPI_ISL_2862953, EPI_ISL_2862955, EPI_ISL_2862957, EPI_ISL_2981128, EPI_ISL_2981262, EPI_ISL_2981334, EPI_ISL_2981446, EPI_ISL_2981465, EPI_ISL_2981466, EPI_ISL_2981467, EPI_ISL_2981888, EPI_ISL_2981891, EPI_ISL_2981893, EPI_ISL_2981894, EPI_ISL_2981896, EPI_ISL_2981901 | | | |
| see above | Dutch COVID-19 response team | National Institute for Public Health and the Environment (RIVM) | Adam Meijer; AnneMarie van den Brandt; Annelies Kroneman; Bas van der Veer; Chantal Reusken; Dennis Schmitz; Dirk Eggink; Euníce Then; Florian Zwagemaker; Harry Vennema; James Groot; Jeroen Cremer; Karim Hajji; Kim Freriks; Linda van de Nes; Lisa Wijsman; Lynn Aarts; Melissa van Tuil; Ryanne Jaarsma; Sanne Bos; Sharon van den Brink; Sjoerd Kuiling; on behalf of the national COVID-19 response team |
| EPI_ISL_1582978, EPI_ISL_1582980 | E.S.E. HOSPITAL SAN JOSE DE MAICAO | Instituto Nacional de Salud- Dirección de Investigación en Salud Pública | Carlos Franco-Muñoz; Carmen Osorio; Diana Malo; Diego A. Álvarez-Díaz; Diego Andrés Prada; Gerardo Santamaría; Hector Alejandro Ruiz-Moreno; Jhonnatan Reales-González; Juan Camilo Martinez; Julian Naizaque; Katherine Laiton-Donato; Lisseth Pardo; Magdalena Wiesner; Marcela Mercado-Reyes; Maria T. Herrera-Sepúlveda; Marta Lopez Blanco; Martha Lucia Ospina Martinez; Paola Rojas; Sergio Gomez; Sheryll Corchuelo; Ángela Alarcon Cruz |
| EPI_ISL_2686819 | EHNV | Laboratory of genomics and metagenomics | Claire Bertelli; Damien Jacot; Gilbert Greub; Sébastien Aeby; Trestan Pillonel |
| EPI_ISL_2617660, EPI_ISL_2617661, EPI_ISL_2617662, EPI_ISL_2617663, EPI_ISL_2617664, EPI_ISL_2617665, EPI_ISL_2617666, EPI_ISL_2617667, EPI_ISL_2617668, EPI_ISL_2617669, EPI_ISL_2617670, EPI_ISL_2617671, EPI_ISL_2617672, EPI_ISL_2617673, EPI_ISL_2617674, EPI_ISL_2617675, EPI_ISL_2617676, EPI_ISL_2617677, EPI_ISL_2617678, EPI_ISL_2617679 | | | |
| see above | Elling group, Institute of Molecular Biotechnology (IMBA) | Bergthaler laboratory, CeMM Research Center for Molecular Medicine of the Austrian Academy of Sciences | Andreas Bergthaler; Anna Schedl; Bekir Erguner; Benedikt Agerer; Christoph Bock; Fabian Amman; Jan Laine; Lukas Endler; Maelle Le Moing; Martin Senekowitsch; Matthew Thornton; Michael Schuster; Petr Triska; Thomas Penz |
| EPI_ISL_2634614 | Eurofins LifeCodexx GmbH | Robert Koch Institute | |
| EPI_ISL_2975271 | Fimlab Laboratories | Fimlab Laboratories | Bruno Luukinen; Leena Huhti; Mauri Keinänen; Minna Paloniemi; Sara Lehtinen; Tapio Seiskari |
| EPI_ISL_2860417 | Florida Bureau of Public Health Laboratories | Florida Bureau of Public Health Laboratories | Jason Blanton; Sarah Schmedes |
| EPI_ISL_2365426 | GERENCIA DE ASISITENCIA SANITARIA DE SORIA | Instituto de Salud Carlos III | A. Monzón; CARMEN; F. Casas; I. Jiménez; I.ALDEA MANSILLA; M. Sandonís; P. Zaballos; S. Cuesta; S. Iglesias-Caballero; S. Pozo; S. Varona; V. Camarero; Vázquez-Morón |
| EPI_ISL_2659196 | Genetica Molecular and Subdepartamento de Virologia ISP Chile | Instituto de Salud Publica de Chile | Andres Castillo; Barbara Parra; Constanza Campano; Gisselle Barra; Javier Tognarelli; Jorge Fernandez; Karen Orostica; Loredana Arata; Patricia Bustos; Rodrigo Fasce; Soledad Ulloa |
| EPI_ISL_2339796, EPI_ISL_2339814, EPI_ISL_2339855, EPI_ISL_2584533, EPI_ISL_2584563, EPI_ISL_2584574 | Grupo de Investigación en Enfermedades Tropicales del Ejército (GINETEJ), Laboratorio de Referencia e Investigación, Dirección de Sanidad Ejército, Bogotá, Colombia | Centro de Investigaciones en Microbiología y Biotecnología-UR (CIMBIUR), Facultad de Ciencias Naturales, Universidad del Rosario, Bogotá, Colombia | Camilo A. Correa-Cárdenas; Carolina Oliveros; Claudia Méndez; Elizabeth K. Márquez; Frank de los Santos Ortiz; Juan David Ramírez; Julie Pérez; Lorena Albarracín; Luz H. Patiño; Maria Clara Duque; Marina Muñoz; María Teresa Alvarado; Nathalia Ballesteros; Sergio Castañeda; Sergio Gutíerrez-Riveros; Yanira Romero; Zulma Cucunubá |
| EPI_ISL_2694913, EPI_ISL_2694919, EPI_ISL_2694924, EPI_ISL_2895188 | H Garcia de Orta | Instituto Nacional de Saude (INSA) | Borges et al |
| EPI_ISL_2695087 | H Vila Franca Xira | Instituto Nacional de Saude (INSA) | Borges et al |
| EPI_ISL_1821073 | HIC | Instituto Nacional de Salud- Dirección de Investigación en Salud Pública | Carlos Franco-Muñoz; Carmen Osorio; Diana Malo; Diego A. Álvarez-Díaz; Diego Andrés Prada; Gerardo Santamaría; Hector Alejandro Ruiz-Moreno; Jhonnatan Reales-González; Jorge Rivera; Juan Camilo Martinez; Julian Naizaque; Katherine Laiton-Donato; Lisseth Pardo; Magdalena Wiesner; Marcela Mercado-Reyes; Maria T. Herrera-Sepúlveda; Marta Lopez Blanco; Martha Lucia Ospina Martinez; Paola Rojas; Sergio Gomez; Sheryll Corchuelo; Ángela Alarcon Cruz |
| EPI_ISL_1821071 | HIGUERA ESCALANTE | Instituto Nacional de Salud- Dirección de Investigación en Salud Pública | Carlos Franco-Muñoz; Carmen Osorio; Diana Malo; Diego A. Álvarez-Díaz; Diego Andrés Prada; Gerardo Santamaría; Hector Alejandro Ruiz-Moreno; Jhonnatan Reales-González; Jorge Rivera; Juan Camilo Martinez; Julian Naizaque; Katherine Laiton-Donato; Lisseth Pardo; Magdalena Wiesner; Marcela Mercado-Reyes; Maria T. Herrera-Sepúlveda; Marta Lopez Blanco; Martha Lucia Ospina Martinez; Paola Rojas; Sergio Gomez; Sheryll Corchuelo; Ángela Alarcon Cruz |
| EPI_ISL_2500969 | HOSPITAL DEPARTAMENTAL MARIA INMACULADA | Instituto Nacional de Salud- Dirección de Investigación en Salud Pública | Carlos Franco-Muñoz; Carmen Osorio; Diana Malo; Diego A. Álvarez-Díaz; Diego Andrés Prada; Gerardo Santamaría; Hector Alejandro Ruiz-Moreno; Jhonnatan Reales-González; Jorge Rivera; Juan Camilo Martinez; Julian Naizaque; Katherine Laiton-Donato; Lisseth Pardo; Magdalena Wiesner; Marcela Mercado-Reyes; Maria T. Herrera-Sepúlveda; Marta Lopez Blanco; Martha Lucia Ospina Martinez; Paola Rojas; Sergio Gomez; Sheryll Corchuelo; Ángela Alarcon Cruz |
| EPI_ISL_2657688, EPI_ISL_2827987 | HOSPITAL DR. CARLOS LUIS VALVERDE VEGA | Inciensa, Instituto Costarricense de Investigación y Enseñanza en Nutrición y Salud | Adriana Godínez; Claudio Soto-Garita; Estela Cordero; Francisco Duarte; Hebleen Porras; Joselyn Prado & Yahaira Ramos; José Luis Vargas; Mariela Gutiérrez; Melany Calderón |
| EPI_ISL_2178400, EPI_ISL_2178401 | HOSPITAL GENERAL UNIVERSITARIO REINA SOFIA | Instituto de Salud Carlos III | A. Monzón; F. Casas; I. Jiménez; I.NUÑEZ TRIGUEROS; M. Sandonís; Mª LUZ; P. Zaballos; S. Cuesta; S. Iglesias-Caballero; S. Pozo; S. Varona; V. Camarero; Vázquez-Morón |
| EPI_ISL_2828015 | HOSPITAL MEXICO | Inciensa, Instituto Costarricense de Investigación y Enseñanza en Nutrición y Salud | Adriana Godínez; Claudio Soto-Garita; Estela Cordero; Francisco Duarte; Hebleen Porras; Joselyn Prado & Juan Carlos Villalobos; José Luis Vargas; Mariela Gutiérrez; Melany Calderón |
| EPI_ISL_2674346, EPI_ISL_2674348 | HOSPITAL REGIONAL DE LA ORINOQUIA | Instituto Nacional de Salud- Dirección de Investigación en Salud Pública | Carlos Franco-Muñoz; Carmen Osorio; Diana Malo; Diego A. Álvarez-Díaz; Diego Andrés Prada; Gerardo Santamaría; Hector Alejandro Ruiz-Moreno; Jhonnatan Reales-González; Jorge Rivera; Juan Camilo Martinez; Julian Naizaque; Katherine Laiton-Donato; Lisseth Pardo; Magdalena Wiesner; Marcela Mercado-Reyes; Maria T. Herrera-Sepúlveda; Marta Lopez Blanco; Martha Lucia Ospina Martinez; Paola Rojas; Sergio Gomez; Sheryll Corchuelo; Ángela Alarcon Cruz |
| EPI_ISL_2674347 | HOSPITAL REGIONAL ORINOQUIA | Instituto Nacional de Salud- Dirección de Investigación en Salud Pública | Carlos Franco-Muñoz; Carmen Osorio; Diana Malo; Diego A. Álvarez-Díaz; Diego Andrés Prada; Gerardo Santamaría; Hector Alejandro Ruiz-Moreno; Jhonnatan Reales-González; Jorge Rivera; Juan Camilo Martinez; Julian Naizaque; Katherine Laiton-Donato; Lisseth Pardo; Magdalena Wiesner; Marcela Mercado-Reyes; Maria T. Herrera-Sepúlveda; Marta Lopez Blanco; Martha Lucia Ospina Martinez; Paola Rojas; Sergio Gomez; Sheryll Corchuelo; Ángela Alarcon Cruz |
| EPI_ISL_2178393, EPI_ISL_2178394, EPI_ISL_2178395, EPI_ISL_2178396, EPI_ISL_2178397, EPI_ISL_2178398, EPI_ISL_2178399, EPI_ISL_2178402, EPI_ISL_2178403, EPI_ISL_2178404, EPI_ISL_2178405, EPI_ISL_2227874, EPI_ISL_2227889 | | | |
| see above | HOSPITAL UNIVERSITARIO VIRGEN DE LA ARRIXACA | Instituto de Salud Carlos III | A. Monzón; F. Casas; I. Jiménez; I. MORENO PARRADO; I.MORENO PARRADO; Iglesias-Caballero; LAURA; M. Camarero; M. Sandonís; P. Zaballos; S. Camarero; S. Cuesta; S. Iglesias-Caballero; S. Pozo; S. Varona; Sandonís; V. Camarero; V. Vázquez-Morón; Vázquez-Morón |
| EPI_ISL_1581369 | Helix/Illumina | Centers for Disease Control and Prevention Division of Viral Diseases, Pathogen Discovery | Adrian Paskey; Alexandre Bolze; Ary Ascencio; Benjamin Rambo-Martin; Brad Sickler; Charlotte Rivera-Garcia; Christine Tran; Christopher Gulvick; Clinton R. Paden; Dakota Howard; Darlene Wagner; David Becker; Dhwani Batra; Duncan MacCannell; Efren Sandoval; Eileen de Feo; Elizabeth Cirulli; Eric Allen; Geraint Levan; James Lu; Jan Antico; Jason Caravas; Jason Nguyen; Jimmy Ramirez; Jingtao Liu; Kara Moser; Kelly Schiabor Barrett; Kim Gietzen; Magnus Isaksson; Marc Laurent; Matthew Schmerer; Matthew Tolentino; Nicole L. Washington; Peter W. Cook; Phil Febbo; Ryan Cho; Scott Sammons; Shannon Wickline; Shatavia Morrison; Sherry Wang; Simon White; Tyler Cassens; William Lee; Yvette Unoarumhi |
| EPI_ISL_1401468, EPI_ISL_1401469, EPI_ISL_1647948, EPI_ISL_1647949, EPI_ISL_1647956 | Hospital General Universitario Gregorio Marañón | Hospital General Universitario Gregorio Marañón | Cristina Rodriguez-Grande; Darío García de Viedma; Laura Pérez-Lago; Patricia Muñoz; Pedro Sola Campoy; Pilar Catalán; Sergio Buenestado Serrano |
| EPI_ISL_1908878 | Hospital Universitari Arnau de Vilanova | Hospital Universitari Vall d'Hebron - Vall d'Hebron Institut de Recerca | Andrés Antón; Ariadna Rando; Carla Castillo; Cristina Andrés; Damir Garcia-Cehic; Josep Quer; Juliana Esperalba; Maria Carmen Martin; Maria Gema Codina; Maria Piñana; Tomàs Pumarola |
| EPI_ISL_2466275 | Hospital Universitari Vall d'Hebron - Vall d'Hebron Institut de Recerca | Hospital Universitari Vall d'Hebron - Vall d'Hebron Institut de Recerca | Alejandra González-Sánchez; Andrés Antón; Ariadna Rando; Carla Castillo; Cristina Andrés; Damir Garcia-Cehic; Josep Quer; Juliana Esperalba; Maria Carmen Martin; Maria Gema Codina; Maria Piñana; Tomàs Pumarola |
| EPI_ISL_2621160, EPI_ISL_2651224 | ICMT-SABANETA | Universidad Nacional de Colombia - Laboratorio Genómico One Health | Andres F. Cardona-Rios; Carlos Franco-Muñoz; Carolina Muñoz-Arango; Celeny Ortiz; Daniel O. Maldonado-Perez; Diego A. Álvarez-Díaz; Hector Alejandro Ruiz-Moreno; Idabely Betancur Ortiz; Jorge E. Osorio; Juan P. Hernandez-Ortiz; Karl A Ciuoderis; Katherine Laiton-Donato; Laura Silvana Perez; Lina M. Hurtado; Marcela Mercado-Reyes; Maria Angélica Maya; Maria Stella López; Rita Almanza Payares; Sandra Ines Cano; Simón Villegas Velásquez |
| EPI_ISL_2458116, EPI_ISL_2488730, EPI_ISL_2897538, EPI_ISL_2897539, EPI_ISL_2897542, EPI_ISL_2897545, EPI_ISL_2897555, EPI_ISL_2897557, EPI_ISL_3010424, EPI_ISL_3010434, EPI_ISL_3010438, EPI_ISL_3010439, EPI_ISL_3010440, EPI_ISL_3010448, EPI_ISL_3010452, EPI_ISL_3010456, EPI_ISL_3010463 | | | |
| see above | INSPI-CRN DE INFLUENZA Y OTROS VIRUS RESPIRATORIOS | NIC-INSPI | Alfredo Bruno; Domenica de Mora.; Jimmy Garcés; Johanna Laines; Lizbeth Patiño; Manuel Gonzalez; Maritza Olmedo; Michelle Páez |
| EPI_ISL_2786928 | Inovie AS LABOSUD | Inovie AS GenBio | Florian VERCRUYSSEN / Pauline LAURENT |
| EPI_ISL_2100436, EPI_ISL_2603836 | Institute of Microbiology, Universidad San Francisco de | Institute of Microbiology, Universidad San Francisco de Quito | Belén Prado-Vivar; Bernardo Gutiérrez; Fernanda Zurita; Gabriel Trueba; Juan José Guadalupe; Luis Morales; Michelle Grunauer; Monica Becerra-Wong; Nelson Montalvan; Patricio Rojas-Silva; Paúl Cárdenas; Sully Márquez; Verónica Barragán |

| | | | |
|---|---|---|---|
| | Quito | | |
| EPI_ISL_2562149 | Institute of Virology, Biomedical Research Center of the Slovak Academy of Sciences, Bratislava | Faculty of Natural Sciences, Comenius University, Bratislava | Boris Klempa; Brona Brejova; Jozef Nosek; Juraj Kopacek; Kristina Borsova; Lubomira Lukacikova; Martina Lickova; Martina Nebohacova; Monika Slavikova; Sabina Fumacova Havlikova; Tomas Vinar; Viktoria Cabanova; Viktoria Hodorova |
| EPI_ISL_2500936, EPI_ISL_2500944 | Instituto Nacional de Salud | Instituto Nacional de Salud- Dirección de Investigación en Salud Pública | Carlos Franco-Muñoz; Carmen Osorio; Diana Malo; Diego A. Álvarez-Díaz; Diego Andrés Prada; Gerardo Santamaría; Hector Alejandro Ruiz-Moreno; Jhonnatan Reales-González; Jorge Rivera; Juan Camilo Martinez; Julian Naizaque; Katherine Laiton-Donato; Lisseth Pardo; Magdalena Wiesner; Marcela Mercado-Reyes; Maria T. Herrera-Sepúlveda; Marta Lopez Blanco; Martha Lucia Ospina Martinez; Paola Rojas; Sergio Gomez; Sheryll Corchuelo; Ángela Alarcon Cruz |
| EPI_ISL_2642004, EPI_ISL_2642005, EPI_ISL_2642006, EPI_ISL_2642008, EPI_ISL_2642009, EPI_ISL_2642010 | Istituto Zooprofilattico Sperimentale del Mezzogiorno | Telethon Institute of Genetics and Medicine (TIGEM) | Antonio Grimaldi Patrizia Annunziata Francesco Panariello Biancamaria Pierri Claudia Tiberio Teresa Giuliano Valentina Bouche Chiara Colantuono Maria Concetta Cuomo Denise Di Concilio Lucio Di Filippo Anna Manfredi Marcello Salvi Antonio Limone Luigi Atripaldi Pellegrino Cerino Andrea Ballabio Davide Cacchiarelli |
| EPI_ISL_2864200, EPI_ISL_2877584, EPI_ISL_2886222, EPI_ISL_2886223, EPI_ISL_2886224, EPI_ISL_2886225, EPI_ISL_2886227, EPI_ISL_2886228 | | | |
| see above | KU Leuven, Rega Institute, Clinical and Epidemiological Virology | KU Leuven, Rega Institute, Clinical and Epidemiological Virology | Bert Vanmechelen; Joan Marti-Carerras; Piet Maes; Tony Wawina-Bokalanga |
| EPI_ISL_2898054 | Karolinska University Hospital Huddinge | Karolinska University Hospital | Annelie Bjerkner; Isak Sylvin; Jan Albert; Karolina Ininbergs; Lina Guerra Blomqvist; Lynda Eneh; Martin Ekman; Martina Wahlund; Robert Dyrdak; Sandra Broddesson; Tanja Normark; Tobias Allander; Valtteri Wirta; Zhibing Yun |
| EPI_ISL_2657873 | LABORATORIO BILOGIA MOLECULAR IMAT SAS | Instituto Nacional de Salud- Dirección de Investigación en Salud Pública | Carlos Franco-Muñoz; Carmen Osorio; Diana Malo; Diego A. Álvarez-Díaz; Diego Andrés Prada; Gerardo Santamaría; Hector Alejandro Ruiz-Moreno; Jhonnatan Reales-González; Jorge Rivera; Juan Camilo Martinez; Julian Naizaque; Katherine Laiton-Donato; Lisseth Pardo; Magdalena Wiesner; Marcela Mercado-Reyes; Maria T. Herrera-Sepúlveda; Marta Lopez Blanco; Martha Lucia Ospina Martinez; Paola Rojas; Sergio Gomez; Sheryll Corchuelo; Ángela Alarcon Cruz |
| EPI_ISL_2674283 | LABORATORIO CONTINENTAL | Instituto Nacional de Salud- Dirección de Investigación en Salud Pública | Carlos Franco-Muñoz; Carmen Osorio; Diana Malo; Diego A. Álvarez-Díaz; Diego Andrés Prada; Gerardo Santamaría; Hector Alejandro Ruiz-Moreno; Jhonnatan Reales-González; Jorge Rivera; Juan Camilo Martinez; Julian Naizaque; Katherine Laiton-Donato; Lisseth Pardo; Magdalena Wiesner; Marcela Mercado-Reyes; Maria T. Herrera-Sepúlveda; Marta Lopez Blanco; Martha Lucia Ospina Martinez; Paola Rojas; Sergio Gomez; Sheryll Corchuelo; Ángela Alarcon Cruz |
| EPI_ISL_1820934 | LABORATORIO DE INVESTIGACION HORMONAL | Instituto Nacional de Salud- Dirección de Investigación en Salud Pública | Carlos Franco-Muñoz; Carmen Osorio; Christian Romero; Diana Malo; Diego A. Álvarez-Díaz; Diego Andrés Prada; Gerardo Santamaría; Hector Alejandro Ruiz-Moreno; Jhonnatan Reales-González; Jorge Rivera; Juan Camilo Martinez; Julian Naizaque; Katherine Laiton-Donato; Lisseth Pardo; Magdalena Wiesner; Marcela Mercado-Reyes; Maria T. Herrera-Sepúlveda; Marta Lopez Blanco; Martha Lucia Ospina Martinez; Paola Rojas; Patricia del Portillo; Sergio Gomez; Sheryll Corchuelo; Ángela Alarcon Cruz |
| EPI_ISL_2621868 | LABORATORIO ECHAVARRIA | Universidad Nacional de Colombia - Laboratorio Genómico One Health | Andres F. Cardona-Rios; Carlos Franco-Muñoz; Carolina Muñoz-Arango; Celeny Ortiz; Daniel O. Maldonado-Perez; Diego A. Álvarez-Díaz; Hector Alejandro Ruiz-Moreno; Idabely Betancur Ortiz; Jorge E. Osorio; Juan P. Hernandez-Ortiz; Karl A Ciuoderis; Katherine Laiton-Donato; Laura Silvana Perez; Lina M. Hurtado; Marcela Mercado-Reyes; Maria Angélica Maya; Maria Stella López; Rita Almanza Payares; Sandra Ines Cano; Simón Villegas Velásquez |
| EPI_ISL_2362580 | LABORATORIO NACY FLOREZ GARCIA | Instituto Nacional de Salud- Dirección de Investigación en Salud Pública | Carlos Franco-Muñoz; Carmen Osorio; Diana Malo; Diego A. Álvarez-Díaz; Diego Andrés Prada; Gerardo Santamaría; Hector Alejandro Ruiz-Moreno; Jhonnatan Reales-González; Jorge Rivera; Juan Camilo Martinez; Julian Naizaque; Katherine Laiton-Donato; Lisseth Pardo; Magdalena Wiesner; Marcela Mercado-Reyes; Maria T. Herrera-Sepúlveda; Marta Lopez Blanco; Martha Lucia Ospina Martinez; Paola Rojas; Sergio Gomez; Sheryll Corchuelo; Ángela Alarcon Cruz |
| EPI_ISL_2896232 | LACEN do Estado de Mato Grosso | Instituto Adolfo Lutz, Interdiciplinary Procedures Center, Strategic Laboratory | Caio Vinicius Dias Lopes; Claudia Regina Gonçalves; Claudio Tavares Sacchi; Karoline Rodrigues Campos |
| EPI_ISL_1960057 | LDSP | Universidad Nacional de Colombia - Laboratorio Genómico One Health | Andres F. Cardona-Rios; Carlos Franco-Muñoz; Carolina Muñoz-Arango; Celeny Ortiz; Daniel O. Maldonado-Perez; Diego A. Álvarez-Díaz; Hector Alejandro Ruiz-Moreno; Idabely Betancur Ortiz; Jorge E. Osorio; Juan P. Hernandez-Ortiz; Karl A Ciuoderis; Katherine Laiton-Donato; Laura Silvana Perez; Lina M. Hurtado; Marcela Mercado-Reyes; Maria Angélica Maya; Maria Stella López; Rita Almanza Payares; Sandra Ines Cano; Simón Villegas Velásquez |
| EPI_ISL_2827782 | LDSP - LAB CLINICO ESPECIALIZADO AIDA ASCENCIO | Instituto Nacional de Salud- Dirección de Investigación en Salud Pública | Carlos Franco-Muñoz; Carmen Osorio; Diana Malo; Diego A. Álvarez-Díaz; Diego Andrés Prada; Gerardo Santamaría; Hector Alejandro Ruiz-Moreno; Jhonnatan Reales-González; Jorge Rivera; Juan Camilo Martinez; Julian Naizaque; Katherine Laiton-Donato; Lisseth Pardo; Magdalena Wiesner; Marcela Mercado-Reyes; Maria T. Herrera-Sepúlveda; Marta Lopez Blanco; Martha Lucia Ospina Martinez; Paola Rojas; Sergio Gomez; Sheryll Corchuelo; Ángela Alarcon Cruz |
| EPI_ISL_1582996 | LDSP BARRANQUILLA | Instituto Nacional de Salud- Dirección de Investigación en Salud Pública | Carlos Franco-Muñoz; Carmen Osorio; Diana Malo; Diego A. Álvarez-Díaz; Diego Andrés Prada; Gerardo Santamaría; Hector Alejandro Ruiz-Moreno; Jhonnatan Reales-González; Juan Camilo Martinez; Julian Naizaque; Katherine Laiton-Donato; Lisseth Pardo; Magdalena Wiesner; Marcela Mercado-Reyes; Maria T. Herrera-Sepúlveda; Marta Lopez Blanco; Martha Lucia Ospina Martinez; Paola Rojas; Sergio Gomez; Sheryll Corchuelo; Ángela Alarcon Cruz |
| EPI_ISL_2158315, EPI_ISL_2158316, EPI_ISL_2158317 | LESP Ciudad de Mexico | Instituto de Diagnostico y Referencia Epidemiologicos (INDRE) | Abril Rodriguez-Maldonado; Ariadna Medina-Benitez; Claudia Wong-Arambula; Ernesto Ramirez-Gonzalez.; Gisela Barrera-Badillo; Irma Lopez-Martinez; Joaquin Quiroz-Mercado; Lucia Hernandez-Rivas; Natividad Cruz-Ortiz; Sergio Rangel-Guerrero; Tatiana Nunez-Garcia; Vanessa Rivero-Arredondo |
| EPI_ISL_2601972 | LKO | Jessa | Berden et al. on behalf of the Jessa_cmdLab |
| EPI_ISL_2657876 | Lab microbiologia FVL | Instituto Nacional de Salud- Dirección de Investigación en Salud Pública | Carlos Franco-Muñoz; Carmen Osorio; Diana Malo; Diego A. Álvarez-Díaz; Diego Andrés Prada; Gerardo Santamaría; Hector Alejandro Ruiz-Moreno; Jhonnatan Reales-González; Jorge Rivera; Juan Camilo Martinez; Julian Naizaque; Katherine Laiton-Donato; Lisseth Pardo; Magdalena Wiesner; Marcela Mercado-Reyes; Maria T. Herrera-Sepúlveda; Marta Lopez Blanco; Martha Lucia Ospina Martinez; Paola Rojas; Sergio Gomez; Sheryll Corchuelo; Ángela Alarcon Cruz |
| EPI_ISL_2844837 | Lab voor klinische biologie | Lab voor klinische biologie | Bruno Verhasselt; Hannelore Hamerlinck; Marija Janevska |
| EPI_ISL_2382623, EPI_ISL_2466775, EPI_ISL_2466824, EPI_ISL_2694812 | Labor Berlin Charité Vivantes GmbH / Institut für Virologie | Charité Universitätsmedizin Berlin, Institut für Virologie/Labor Berlin | Barbara Mühlemann; Christian Drosten; Christine Stephan; Peter Menzel; Rolf Schwarzer; Terry Jones; Victor M Corman |
| EPI_ISL_2111528, EPI_ISL_2111598 | Labor ZOTZ\|KLIMAS; MVZ Dusseldorf-Centrum | Robert Koch Institute | |
| EPI_ISL_2636058, EPI_ISL_2678752 | Labor ZOTZ\|KLIMAS; MVZ Düsseldorf-Centrum | Robert Koch Institute | |
| EPI_ISL_2361500, EPI_ISL_2361501 | Laboratorio HUB -Azienda Ospedaliero Universitaria - AOU – Cagliari | Laboratorio SPOKE Biologia Molecolare -Azienda Ospedaliero Universitaria - AOU – Cagliari | Alessandra Scano; Ferdinando Coghe; Germano Orrù; Miriam Loddo; Riccardo Cappai; Sara Fais; Valentina Medda |
| EPI_ISL_2674362 | Laboratorio Salud Publica San Andres Islas | Instituto Nacional de Salud- Dirección de Investigación en Salud Pública | Carlos Franco-Muñoz; Carmen Osorio; Diana Malo; Diego A. Álvarez-Díaz; Diego Andrés Prada; Gerardo Santamaría; Hector Alejandro Ruiz-Moreno; Jhonnatan Reales-González; Jorge Rivera; Juan Camilo Martinez; Julian Naizaque; Katherine Laiton-Donato; Lisseth Pardo; Magdalena Wiesner; Marcela Mercado-Reyes; Maria T. Herrera-Sepúlveda; Marta Lopez Blanco; Martha Lucia Ospina Martinez; Paola Rojas; Sergio Gomez; Sheryll Corchuelo; Ángela Alarcon Cruz |
| EPI_ISL_1582988, EPI_ISL_1582989 | Laboratorio de Biología Molecular – Universidad del Magdalena | Instituto Nacional de Salud- Dirección de Investigación en Salud Pública | Carlos Franco-Muñoz; Carmen Osorio; Diana Malo; Diego A. Álvarez-Díaz; Diego Andrés Prada; Gerardo Santamaría; Hector Alejandro Ruiz-Moreno; Jhonnatan Reales-González; Juan Camilo Martinez; Julian Naizaque; Katherine Laiton-Donato; Lisseth Pardo; Magdalena Wiesner; Marcela Mercado-Reyes; Maria T. Herrera-Sepúlveda; Marta Lopez Blanco; Martha Lucia Ospina Martinez; Paola Rojas; Sergio Gomez; Sheryll Corchuelo; Ángela Alarcon Cruz |
| EPI_ISL_2544236, EPI_ISL_2708977 | Laboratorio di Microbiologia | Laboratorio di Microbiologia | Martinetti Lucchini Gladys; Valeria Spina |
| EPI_ISL_2614407 | Lifebrain Covid Labor GmbH | Dept. of Laboratory Medicine | Anna Gschaider; Claudia Weber; Fabian Konig; Harald Esterbauer; Oswald Wagner; Petra Jurkowitsch; Robert Strassl; Sabina Plumer |
| EPI_ISL_2682959 | Lighthouse Lab in Glasgow | Wellcome Sanger Institute for the COVID-19 Genomics UK (COG-UK) Consortium | Anna Dominiczak and Alex Alderton; Carol Clugston; Cordelia Langford; David Gray; David K. Jackson; Dominic Kwiatkowski; Ewan Harrison; Harper VanSteenhouse; Ian Johnston; Jeffrey Barrett; John Sillitoe on behalf of the Wellcome Sanger Institute COVID-19 Surveillance Team; Roberto Amato; Sonia Goncalves; Yumi Kasai |
| EPI_ISL_2392230, EPI_ISL_2651559, EPI_ISL_2729283, EPI_ISL_2911018 | Lighthouse Lab in Milton Keynes | Wellcome Sanger Institute for the COVID-19 Genomics UK (COG-UK) Consortium | Cordelia Langford; David K. Jackson; Dominic Kwiatkowski; Ewan Harrison; Ian Johnston; Jeffrey Barrett; John Sillitoe on behalf of the Wellcome Sanger Institute COVID-19 Surveillance Team; Roberto Amato; Sonia Goncalves; The Lighthouse Lab in Milton Keynes and Alex Alderton |
| EPI_ISL_2849837 | Lighthouse Laboratory Plymouth | Wellcome Sanger Institute for the COVID-19 Genomics UK (COG-UK) Consortium | Cordelia Langford; David K. Jackson; Dominic Kwiatkowski; Ewan Harrison; Ian Johnston; Jeffrey Barrett; John Sillitoe on behalf of the Wellcome Sanger Institute COVID-19 Surveillance Team; Lighthouse Laboratory Plymouth and Alex Alderton; Roberto Amato; Sonia Goncalves |
| EPI_ISL_2689085 | MD PHL | MD PHL | Maryland Department of Health Laboratories Administration |
| EPI_ISL_2626466 | MIRIALIS CLUSES BECHET | CNR Virus des Infections Respiratoires - France SUD | Antonin Bal; Bruno Lina; Bruno Simon; Gregory Destras; Gwendolyne Burfin; Hadrien Regue; Laurence Josset; Martine Valette; Quentin Semanas |
| EPI_ISL_2918865 | Medica | Institute of Medical Virology | Alexandra Trkola; Annette Audigé; Cyril Shah; Gabriela Ziltener; Guido Bloemberg; Jon Huder; Jürg Böni; Kevin Steiner; Maria Grünberg; Maryam Zaheri; Michael Huber; Riccarda Capaul; Stefan Schmutz; Verena Kufner |
| EPI_ISL_2650339 | Microbiologia CATLAB | Can Ruti SARS-CoV-2 Sequencing Hub (HUGTiP/IrsiCaixa/IGTP) | Alba Sánchez; Anna Not; Antoni E Bordoy; Bonaventura Clotet; Cristina Casañ; Cristina Esteban; Francesc Catala-Moll; Gemma Clara; Ignacio Blanco; Marc Noguera-Julian; Maria Casadellà; Mariona Parera; Mercedes Guerrero; Montserrat Giménez; Pere-Joan Cardona; Pilar Armengol; Roger Paredes; Verónica Saludes; and Elisa Martró on behalf of the Can Ruti SARS-CoV-2 Sequencing Hub |
| EPI_ISL_2926201 | Microbiology Department, Laboratori Clínic Metropolitana Nord. Hospital Universitari Germans Trias i Pujol | Can Ruti SARS-CoV-2 Sequencing Hub (HUGTiP/IrsiCaixa/IGTP) | Alba Sánchez; Alexia París; Anna Not; Antoni E Bordoy; Bonaventura Clotet; Cristina Casañ; David Panisello; Francesc Catala-Moll; Gemma Clara; Ignacio Blanco; Laia Soler; Lauro Sumoy; Marc Noguera-Julian; Maria Casadellà; Mariona Parera; Mercedes Guerrero; Montserrat Giménez; Pere-Joan Cardona; Pilar Armengol; Roger Paredes; Verónica Saludes; and Elisa Martró on behalf of the Can Ruti SARS-CoV-2 Sequencing Hub |
| EPI_ISL_1820898, EPI_ISL_1820899, EPI_ISL_2259146, EPI_ISL_2259158, EPI_ISL_2259204, EPI_ISL_2259205, EPI_ISL_2420759 | | | |
| see above | Microvida | Microvida | Jaco Verweij; Joep Stohr; Joep Stöhr; Suzan D. Pas |
| EPI_ISL_2854655 | Molecular Diagnostics Pathology Department Mater Dei Hospital Malta | Molecular Diagnostics Pathology Department Mater Dei Hospital Malta | C Cilia; G Zahra; L Grech; M Briffa; R Borg |
| EPI_ISL_2784724 | National Platform bis UMONS/Jolimont | National Platform bis UMONS/Jolimont | Eric Tarantino; Florian Juszczak; François Dufrasne; Gautier Detry; Guillaume Bayon-Vicente; Ruddy Wattiez |

| | | | |
|---|---|---|---|
| EPI_ISL_2932231, EPI_ISL_2932234, EPI_ISL_3011367 | National Virus Reference Laboratory | National Virus Reference Laboratory | Charlene Bennett; Cillian F De Gascun; Gabriel Gonzalez; Jonathan Dean; Michael Carr; Zoe Yandle |
| EPI_ISL_2876172 | Ohio Department of Health Laboratory | Ohio Department of Health Laboratory | Allison Black; Brent Lee; Caitlin McDonnell; Eric Brandt; Erica Leasure; Glen McGillivary; Heather Blankenship; Holmes; Jade Mowery; Jennifer; Kelsey Florek; Keoni Omura; Kirtana Ramadugu; Quanta Brown; Stephanie Mccracken; Tyler Payne; and Tammy Bannerman |
| EPI_ISL_2905966 | Originating lab: Wales Specialist Virology Centre Sequencing lab: Pathogen Genomics Unit | Public Health Wales Microbiology Cardiff Wales Specialist Virology Centre | Alec Birchley; Alexander Adams; Amy Gaskin; Angela Marchbank; Bree Gatica-Wilcox; Catherine Moore; Jason Coombes; Joanne Watkins; Joel Southgate; Johnathan Evans; Laura Gifford; Lauren Gilbert; Lee Graham; Malorie Perry; Matthew Bull; Nicole Pacchiarini; Sally Corden; Sara Kumziene-Summerhayes; Sara Rey; Sarah Taylor; Simon Cottrell; Sophie Jones; Tom Connor |
| EPI_ISL_2621179 | PRIME DIAGNOSTICS | Universidad Nacional de Colombia - Laboratorio Genómico One Health | Andres F. Cardona-Rios; Carlos Franco-Muñoz; Carolina Muñoz-Arango; Celeny Ortiz; Daniel O. Maldonado-Perez; Diego A. Álvarez-Díaz; Hector Alejandro Ruiz-Moreno; Idabely Betancur Ortiz; Jorge E. Osorio; Juan P. Hernandez-Ortiz; Karl A Ciuoderis; Katherine Laiton-Donato; Laura Silvana Perez; Lina M. Hurtado; Marcela Mercado-Reyes; Maria Angélica Maya; Maria Stella López; Rita Almanza Payares; Sandra Ines Cano; Simón Villegas Velásquez |
| EPI_ISL_1385573, EPI_ISL_2965056 | Pandemic Response Lab - NYC | Pandemic Response Lab, R&D | Cybill del Castillo; Dylan Law; Haiping Hao; Henry Lee; Isabel Fernandez Escapa; Jon Laurent; Melissa Hopkins; Michael Hammerling; Pradeep Bugga; Shinyoung Clair Kang; Sol Rey; William Ward |
| EPI_ISL_2455384 | Premier Medical Lab | Minnesota Department of Health, Public Health Laboratory | Alexandra Lorentz; Jacob Garfin; Matt Plumb; and Xiong Wang |
| EPI_ISL_2611881, EPI_ISL_2673694 | Public Health Authority of the Slovak Republic | Laboratory of Genomics and Bioinformatics, Comenius University Science Park | Anna Gičová; Diana Rusňáková; Jakub Styk; Jaroslav Budiš; Miroslav Böhmer; Tatiana Sedláčková; Tomáš Szemes |
| EPI_ISL_2818170, EPI_ISL_2818299, EPI_ISL_2904293 | Respiratory Virus Unit, Microbiology Services Colindale, Public Health England | COVID-19 Genomics UK (COG-UK) Consortium | PHE Covid Sequencing Team |
| EPI_ISL_2933813, EPI_ISL_2933815 | SARS-CoV-2 testing team, National Institute of Infectious Diseases | Pathogen Genomics Center, National Institute of Infectious Diseases | Hazuka Y Furihata; Hiromizu Takahashi; Kentaro Itokawa; Makoto Kuroda; Masanori Hashino; Masumichi Saito; Naomi Nojiri; Nozomu Hanaoka; Rina Tanaka; Tsuguto Fujimoto; Tsuyoshi Sekizuka |
| EPI_ISL_2566238, EPI_ISL_2566240 | SC (UCO) Igiene e Sanità Pubblica, ASUGI, Trieste | ARGO Laboratorio Genomica ed Epigenomica | Barbone F; Braida C; Busetti M; D'Agaro P; Dal Monego S; Degasperi M; Licastro D; Maggiore A; Marcello A; Piscianz E; Segat L |
| EPI_ISL_2178749, EPI_ISL_2178755 | Salud Digna | Instituto Nacional de Medicina Genomica | Abraham Campos-Romero; Cedro-Tanda A; Cisneros-Villanueva M; Escobar-Arrazola; Gonzalez-Barrera D; Herrera-Montalvo LA.; Hidalgo-Miranda A; Luna-Ruiz Marco; M; Mendoza-Vargas A; Moreno-Camacho José Luis; Munguia-Garza P; Orjuela-Rodríguez M; Ramirez-Vega O; Rangel-DeLeon D; Reyes-Grajeda JP; Rodriguez-Gallegos Jorge |
| EPI_ISL_2179709 | Servicio Microbiología Hospital La Paz | Servicio Microbiología Hospital La Paz | Elie Dahdouh; Fernando Lázaro; Jesús Mingorance Cruz; Rubén Cáceres |
| EPI_ISL_2925507, EPI_ISL_2931128 | Servicio de Microbiología Clínica (Complejo Hospitalario de Navarra, Pamplona) | Centro de Secuenciación NASERTIC | Ana Miqueleiz; Ana Navascués; Carmen Ezpeleta Baquedano |
| EPI_ISL_2438026, EPI_ISL_2438034, EPI_ISL_2621190, EPI_ISL_2621192, EPI_ISL_2651211 | Somer | Universidad Nacional de Colombia - Laboratorio Genómico One Health | Andres F. Cardona-Rios; Carlos Franco-Muñoz; Carolina Muñoz-Arango; Celeny Ortiz; Daniel O. Maldonado-Perez; Diego A. Álvarez-Díaz; Hector Alejandro Ruiz-Moreno; Idabely Betancur Ortiz; Jorge E. Osorio; Juan P. Hernandez-Ortiz; Karl A Ciuoderis; Katherine Laiton-Donato; Laura Silvana Perez; Lina M. Hurtado; Marcela Mercado-Reyes; Maria Angélica Maya; Maria Stella López; Rita Almanza Payares; Sandra Ines Cano; Simón Villegas Velásquez |
| EPI_ISL_2875838 | Spital Limmattal | Institute of Medical Virology | Alexandra Trkola; Annette Audigé; Catharine Aquino; Cyril Shah; Daniel Ehrsam; Gabriela Ziltener; Guido Bloemberg; Hubert Rehrauer; Isabel Stürmer; Joel Wirz; Jon Huder; Jürg Böni; Kevin Steiner; Maria Grünberg; Maryam Zaheri; Michael Huber; Riccarda Capaul; Stefan Schmutz; Verena Kufner; Weihong Qi |
| EPI_ISL_2769123, EPI_ISL_2769124 | Spital Männedorf AG | Institute of Medical Virology | Alexandra Trkola; Annette Audigé; Cyril Shah; Gabriela Ziltener; Guido Bloemberg; Jon Huder; Jürg Böni; Kevin Steiner; Maria Grünberg; Maryam Zaheri; Michael Huber; Riccarda Capaul; Stefan Schmutz; Verena Kufner |
| EPI_ISL_2438017, EPI_ISL_2621151 | Synlab | Universidad Nacional de Colombia - Laboratorio Genómico One Health | Andres F. Cardona-Rios; Carlos Franco-Muñoz; Carolina Muñoz-Arango; Celeny Ortiz; Daniel O. Maldonado-Perez; Diego A. Álvarez-Díaz; Hector Alejandro Ruiz-Moreno; Idabely Betancur Ortiz; Jorge E. Osorio; Juan P. Hernandez-Ortiz; Karl A Ciuoderis; Katherine Laiton-Donato; Laura Silvana Perez; Lina M. Hurtado; Marcela Mercado-Reyes; Maria Angélica Maya; Maria Stella López; Rita Almanza Payares; Sandra Ines Cano; Simón Villegas Velásquez |
| EPI_ISL_2651119, EPI_ISL_2651120 | Temporary Specimen Collection Centre at the AsiaWorld-Expo | Hong Kong Department of Health | Alan K.L. Tsang; Dominic N.C. Tsang; Edman T.K. Lam; Ken H.L. Ng; Peter C.W. Yip; Rickjason C.W. Chan |
| EPI_ISL_2964673 | The Caribbean Public Health Agency | Carrington Lab, Department of Preclinical Sciences, Faculty of Medical Sciences, The University of the West Indies, St Augustine Campus | Anushka Ramjag; Arianne Brown-Jordan; Avery Hinds; Christine V. F. Carrington; Gabriel Escobar; Kenneth George; Nikita S. D. Sahadeo; Nuno Faria; Oliver Pybus; Risha Singh; Sarah Hill; SueMin Nathaniel; Vernie Ramkissoon |
| EPI_ISL_2993634, EPI_ISL_2993635 | The Centre for Clinical Infection & Diagnostics Research, KCL | The Centre for Clinical Infection & Diagnostics Research, KCL | Adela Alcolea-Medina; Gaia Nebbia; Jonathan D Edgeworth; Luke B Snell; Penny R Cliff; Rahul Batra; Sam T Douthwaite; Themoula Charalampous |
| EPI_ISL_2681041 | UM im. Karola Marcinkowskiego w Poznaniu Laboratorium UCA_Covid-19 Centrum Biologii Medycznej | 1. Tricity SARS-CoV-2 sequencing consortium: University of Gdansk, Medical University of Gdansk, Vaxican Ltd., Invicta Ltd. 2. National Institute of Public Health - National Institute of Hygiene, Warsaw, Poland | Celina Cybulska; Karolina Gackowska; Katarzyna Groth; Katarzyna Zacharczuk; Krystyna Bienkowska Szewczyk; Lukasz Rabalski; Maciej Grzybek; Maciej Kosinski; Magdalena Nowakowska; Marcin Lubocki; Małgorzata Sadkowska-Todys; Tomasz Wołkowicz |
| EPI_ISL_2136319, EPI_ISL_2321609 | UNIGEM | Universidad Nacional de Colombia - Laboratorio Genómico One Health | Andres F. Cardona-Rios; Carlos Franco-Muñoz; Carolina Muñoz-Arango; Celeny Ortiz; Daniel O. Maldonado-Perez; Diego A. Álvarez-Díaz; Hector Alejandro Ruiz-Moreno; Idabely Betancur Ortiz; Jorge E. Osorio; Juan P. Hernandez-Ortiz; Karl A Ciuoderis; Katherine Laiton-Donato; Laura Silvana Perez; Lina M. Hurtado; Marcela Mercado-Reyes; Maria Angélica Maya; Maria Stella López; Rita Almanza Payares; Sandra Ines Cano; Simón Villegas Velásquez |
| EPI_ISL_2628889, EPI_ISL_2628969, EPI_ISL_2645992, EPI_ISL_2810514, EPI_ISL_2810516, EPI_ISL_2810523, EPI_ISL_2895450 | | | |
| see above | UNILABS | Instituto Nacional de Saude (INSA) | Borges et al |
| EPI_ISL_2810398 | UNILABS | Instituto Nacional de Saude (INSA) and BioSystems & Integrative Sciences Institute (BioISI) Genomics Unit, FCUL | Borges et al |
| EPI_ISL_2796029, EPI_ISL_2796098, EPI_ISL_2796104 | UNILABS | Instituto Nacional de Saude (INSA) and Instituto Gulbenkian de Ciencia (IGC) | Borges et al |
| EPI_ISL_1820935 | UNIVERSIDAD DE CARTAGENA | Instituto Nacional de Salud- Dirección de Investigación en Salud Pública | Carlos Franco-Muñoz; Carmen Osorio; Christian Romero; Diana Malo; Diego A. Álvarez-Díaz; Diego Andrés Prada; Gerardo Santamaría; Hector Alejandro Ruiz-Moreno; Jhonnatan Reales-González; Jorge Rivera; Juan Camilo Martinez; Julian Naizaque; Katherine Laiton-Donato; Lisseth Pardo; Magdalena Wiesner; Marcela Mercado-Reyes; Maria T. Herrera-Sepúlveda; Marta Lopez Blanco; Martha Lucia Ospina Martinez; Paola Rojas; Patricia del Portillo; Sergio Gomez; Sheryll Corchuelo; Ángela Alarcon Cruz |
| EPI_ISL_1820926 | UNIVERSIDAD DE Magdalena | Instituto Nacional de Salud- Dirección de Investigación en Salud Pública | Carlos Franco-Muñoz; Carmen Osorio; Christian Romero; Diana Malo; Diego A. Álvarez-Díaz; Diego Andrés Prada; Gerardo Santamaría; Hector Alejandro Ruiz-Moreno; Jhonnatan Reales-González; Jorge Rivera; Juan Camilo Martinez; Julian Naizaque; Katherine Laiton-Donato; Lisseth Pardo; Magdalena Wiesner; Marcela Mercado-Reyes; Maria T. Herrera-Sepúlveda; Marta Lopez Blanco; Martha Lucia Ospina Martinez; Paola Rojas; Patricia del Portillo; Sergio Gomez; Sheryll Corchuelo; Ángela Alarcon Cruz |
| EPI_ISL_431116 | Unidad de Investigacion Biomedica de Zacatecas (UIBZ) | Unidad de Genomica Avanzada | ; Alejandra Garcia-Gasca; Alejandra Hernandez-Teran; Alejandro Sanchez-Flores; Alfredo Herrera-Estrella; Alicia Ocaña-Mondragon; Andreu Comas-Garcia; Angel Gustavo Salas-Lais; Antonio Loza Roman; Bernardo Martinez-Miguel; Blanca Taboada; Brenda Irasema Maldonado-Meza; Bruno Gomez-Gil; Carla Ivon Herrera-Najera; Carlos F. Arias; Celia Boukadida; Celida Duque Molina; Celida Martinez- Rodriguez; Clara Esperanza Santacruz-Tinoco; Concepcion Grajales-Muñiz; Consorcio Mexicano de Vigilancia Genomica (CoViGen-Mex). Authors (in alphabetical order): Julio Elias Alvarado-Yaah; Cristobal Chaidez-Quiroz; Daniel Fregoso-Rueda; Daniel Lira Morales; Eduardo Becerril-Vargas; Fernando Fontove-Herrera; Fidencio Mejia-Nepomuceno; Francisco Pulido; Gloria Elena Espinosa-Ayala; Gloria Maria Molina-Salinas; Gloria Vazquez; Hector Esteban Paz-Juarez; Hector Montoya-Fuentes; Helen Haydee Fernanda Ramirez-Plascencia; Irvin Gonzalez-Lopez; Jean Pierre Gonzalez; Jesus Hernandez; Joel Armando Vazquez-Perez.; Jorge Salas-Hernandez; Jose Antonio Enciso-Moreno; Jose Arturo Martinez-Orozco; Jose Esteban Muñoz-Medina; Jose de Jesus Nuñez-Contreras; Juan Bautista Chale-Dzul; Julissa Enciso-Ibarra; Luis Alberto Ochoa-Carrera; Margarita Matias-Florentino; Maria Guadalupe Santiago-Mauricio; Maria Guadalupe de Jesus Mireles-Rivera; Mario Mujica-Sanchez; Marissa Perez-Garcia; Nelly Selem-Mojica; Pavel Isa; Ricardo Ciria Merce; Ricardo Grande; Rosa Maria Gutierrez Rios; Santiago avila-Rios; Selene Zarate; Susana Lopez; Veronica Mata-Haro; Victor Eduardo Garcia-Arias; Victor Hugo Borja-Aburto |
| EPI_ISL_2438098 | Universidad Nacional de Colombia - Laboratorio Genómico One Health | Universidad Nacional de Colombia - Laboratorio Genómico One Health | Andres F. Cardona-Rios; Carlos Franco-Muñoz; Carolina Muñoz-Arango; Celeny Ortiz; Daniel O. Maldonado-Perez; Diego A. Álvarez-Díaz; Hector Alejandro Ruiz-Moreno; Idabely Betancur Ortiz; Jorge E. Osorio; Juan P. Hernandez-Ortiz; Karl A Ciuoderis; Katherine Laiton-Donato; Laura Silvana Perez; Lina M. Hurtado; Marcela Mercado-Reyes; Maria Angélica Maya; Maria Stella López; Rita Almanza Payares; Sandra Ines Cano; Simón Villegas Velásquez |
| EPI_ISL_1220045 | Universidad de Magdalena | Instituto Nacional de Salud- Dirección de Investigación en Salud Pública | Carlos Franco-Muñoz; Diego A. Álvarez-Díaz; Diego Andrés Prada; Gerardo Santamaría; Hector Alejandro Ruiz-Moreno; Jhonnatan Reales-González; Julian Naizaque; Katherine Laiton-Donato; Magdalena Wiesner; Marcela Mercado-Reyes.; Maria T. Herrera-Sepúlveda; Martha Lucia Ospina Martínez; Sheryll Corchuelo |
| EPI_ISL_1582993 | Universidad del Atlántico Laboratorio de Investigación en Biología Molecular | Instituto Nacional de Salud- Dirección de Investigación en Salud Pública | Carlos Franco-Muñoz; Carmen Osorio; Diana Malo; Diego A. Álvarez-Díaz; Diego Andrés Prada; Gerardo Santamaría; Hector Alejandro Ruiz-Moreno; Jhonnatan Reales-González; Juan Camilo Martinez; Julian Naizaque; Katherine Laiton-Donato; Lisseth Pardo; Magdalena Wiesner; Marcela Mercado-Reyes; Maria T. Herrera-Sepúlveda; Marta Lopez Blanco; Martha Lucia Ospina Martinez; Paola Rojas; Sergio Gomez; Sheryll Corchuelo; Ángela Alarcon Cruz |
| EPI_ISL_1424054, EPI_ISL_1424055 | Universidad del Magdalena | Instituto Nacional de Salud- Dirección de Investigación en Salud Pública | Carlos Franco-Muñoz; Diego A. Álvarez-Díaz; Diego Andrés Prada; Gerardo Santamaría; Hector Alejandro Ruiz-Moreno; Jhonnatan Reales-González; Julian Naizaque; Katherine Laiton-Donato; Magdalena Wiesner; Marcela Mercado-Reyes.; Maria T. Herrera-Sepúlveda; Martha Lucia Ospina Martínez; Sheryll Corchuelo |
| EPI_ISL_2355434, EPI_ISL_2517670 | University College London, Great Ormond Street Hospital for Children NHS Foundation Trust, Imperial College Healthcare NHS Trust | COVID-19 Genomics UK (COG-UK) Consortium | Alison Holmes; Charlotte Williams; Helena Tutill; Jacqueline Findlay; James Price; Judith Breuer; Julianne Brown; Kathryn Harris; Leysa Forrest; Mark Kristiansen; Paola Niola; Paola Resende Silva; Patricia Dyal; Paul Randell; Rachel Williams; Samuel Weeks; Sergi Castellano; Sunando Roy; Tony Brooks; Yasmin Panchbhaya |
| EPI_ISL_2405310, EPI_ISL_2405311, EPI_ISL_2601042, EPI_ISL_2601071, EPI_ISL_2601076, EPI_ISL_2601085, EPI_ISL_2885486, EPI_ISL_2885495, EPI_ISL_2885496, EPI_ISL_2981946 | | | |

| | | | |
|---|---|---|---|
| see above | University Hospitals of Geneva, Laboratory of Virology | HUG, Laboratory of Virology and the Health2030 Genome Center | Ana Rita Goncalves; Deborah Penet; Emmanouil Dermitzakis; Henri Pegeot; Ioannis Xenarios; Keith Harshman; Laurent Kaiser; Lorenzo Cerutti; Melyssa Elies; Samuel Cordey |
| EPI_ISL_2769134, EPI_ISL_2769148, EPI_ISL_2918872, EPI_ISL_2978325 | UniversitätsSpital Zürich | Institute of Medical Virology | Alexandra Trkola; Annette Audigé; Cyril Shah; Gabriela Ziltener; Guido Bloemberg; Jon Huder; Jürg Böni; Kevin Steiner; Maria Grünberg; Maryam Zaheri; Michael Huber; Riccarda Capaul; Stefan Schmutz; Verena Kufner |
| EPI_ISL_2390184 | Universitätsklinikum Bonn - Institut für Virologie | Robert Koch Institute | |
| EPI_ISL_1820955 | VIROLOGIA INS DRSP | Instituto Nacional de Salud- Dirección de Investigación en Salud Pública | Carlos Franco-Muñoz; Carmen Osorio; Christian Romero; Diana Malo; Diego A. Álvarez-Díaz; Diego Andrés Prada; Gerardo Santamaría; Hector Alejandro Ruiz-Moreno; Jhonnatan Reales-González; Jorge Rivera; Juan Camilo Martinez; Julian Naizaque; Katherine Laiton-Donato; Lisseth Pardo; Magdalena Wiesner; Marcela Mercado-Reyes; Maria T. Herrera-Sepúlveda; Marta Lopez Blanco; Martha Lucia Ospina Martinez; Paola Rojas; Patricia del Portillo; Sergio Gomez; Sheryll Corchuelo; Ángela Alarcon Cruz |
| EPI_ISL_2379728, EPI_ISL_2420116, EPI_ISL_2521342, EPI_ISL_2521344 | Valais Hospital, Central Institute | Valais Hospital, Central Institute | Alexis Dumoulin; Deborah Penet; Emmanouil Dermitzakis; Henri Pegeot; Ioannis Xenarios; Keith Harshman; Lorenzo Cerutti; Melyssa Elies |
| EPI_ISL_2152100, EPI_ISL_2544315, EPI_ISL_2662426, EPI_ISL_2724442, EPI_ISL_2820613 | Viollier AG | Department of Biosystems Science and Engineering, ETH Zürich | Andrea Patrignani; Andrea Patrizia Salzmann; Andreia Cabral de Gouvea; Catharine Aquino; Chaoran Chen; Christian Beisel; Christiane Beckmann; Christoph Noppen; Daniel Ehrsam; David Dreifuss; Doris Popovic; Elodie Burcklen; Griffin White; Henriette Kurth; Ina Nissen; Isabel Stürmer; Ivan Topolsky; Jay Tracy; Katharina Jahn; Kim Philipp Jablonski; Lara Fuhrmann; Laura Neff; Lennart Opitz; Maria Domenica Moccia; Maurice Redondo; Mirjam Feldkamp; Natascha Santacroce; Niko Beerenwinkel; Noemie Santamaria de Souza; Olivier Kobel; Philipp Jablonski; Ralph Schlapbach; Rebecca Denes; Sarah Nadeau; Simon Grüter; Sophie Seidel; Tanja Stadler; Timothy Sykes |
| EPI_ISL_2833685 | Zentrallabor Zürich | Institute of Medical Virology | Alexandra Trkola; Annette Audigé; Cyril Shah; Gabriela Ziltener; Guido Bloemberg; Jon Huder; Jürg Böni; Kevin Steiner; Maria Grünberg; Maryam Zaheri; Michael Huber; Riccarda Capaul; Stefan Schmutz; Verena Kufner |
| EPI_ISL_2600375 | cerballiance-IDF | Cerba lab | Aude Lessenne; Bénédicte Roquebert; Emmanuel Lecorche; Kader Merah; Laura Verdurme; Patrice Herisson; Sabine Trombert-Paolantoni; Stéphanie Haïm-Boukobza; Thierry Collin |
